# Safety and pathovariant-independent susceptibility in a *Salmonella* Typhimurium controlled human infection model: a phase 1, randomised, double-blind, dose-escalation study

**DOI:** 10.64898/2026.07.23.26358774

**Authors:** Christopher Smith, Anna Rydlova, Robert Varro, Emma Smith, Xinxue Liu, Yujie Ni, Emily Conibear, Zhengze Zhang, Chen Zhu, Shixin Wang, Sewoong Jun, Kyra Jankovich, Ryo Kusakari, Luke John, Mariam Alireza, Zainab Kiliddar, Anna Morkowska, Blanca Perez-Sepulveda, Xiaojun Zhu, Jen Mae Low, Germain Lam, Oshani Dissanayake, Vishal Pratap, Rocio Canals, Daniele De Simone, Francesca Mancini, Omar Rossi, Esmelda Chirwa, Peter W.S. Hill, Christopher Chiu, Robert K.M. Choy, Andrew J. Pollard, Melita A. Gordon, Graham S. Cooke, Jay C.D. Hinton, Malick M. Gibani

## Abstract

**Background:** Invasive non-typhoidal *Salmonella* (iNTS) disease causes an estimated 605,000 cases and 76,000 deaths each year, concentrated in sub-Saharan Africa, where the African *Salmonella* Typhimurium sequence type 313 (ST313) lineage predominates. Vaccine development is hampered by an absence of efficacy data and undefined correlates of protection.

**Methods:** We conducted a phase 1, randomised, double-blind, dose-escalation controlled human infection model (CHIM) in healthy UK-resident adults, who were randomly assigned 1:1 to oral challenge with S. Typhimurium 4/74 (ST19, associated with gastrointestinal disease) or D23580 (ST313, associated with invasive disease). Dose-escalation was guided by a Bayesian continual reassessment method (CRM). The primary endpoint was *Salmonella* diagnosis, defined as sustained fever ≥38°C on ≥2 occasions ≥12 hours apart and/or bacteraemia. Trial registration ClinicalTrials.gov (NCT05870150).

**Findings:** Between August 2023 and December 2024, 50 participants were enrolled (25 per strain). 10^5^ CFU was the maximum feasible dose, with CRM-estimated attack rates of 57·9% (95% credible interval 37·3–73·8) for D23580 and 47·4% (26·7–65·7) for 4/74. There were no serious adverse events. We found no clinical, microbiological, or immunological difference between the two pathovariants. Higher baseline serum anti-O-antigen IgG was associated with reduced disease (adjusted OR 0·42, 95% CI 0·17–0·90) and higher baseline faecal anti-lipopolysaccharide IgA with reduced colonisation (OR 0·12, 95% CI 0·01–0·59).

**Interpretation:** This *S*. Typhimurium CHIM is safe, reproducible, and provides a platform to generate early efficacy signals and candidate correlates of susceptibility, thereby de-risking future iNTS vaccine trials. The absence of a phenotypic difference between the invasive and gastrointestinal pathovariants in immunocompetent adults suggests that host factors, rather than pathogen adaptation alone, shape the invasive phenotype seen in endemic settings.

**Funding:** Wellcome Trust.

## Introduction

Non-typhoidal *Salmonella* (NTS) serovars are globally distributed enteric pathogens. The most common presentation is foodborne gastroenteritis, with an estimated 95·1 million cases annually.^1^ *Salmonella* enterica also ranks among the leading causes of bloodstream infection across Africa and Asia.^2^ Invasive NTS disease (iNTS), predominantly due to *Salmonella* Typhimurium and *Salmonella* Enteritidis, causes an estimated 605,000 cases and 76,000 deaths annually.^3,4^ Multidrug resistance has spread rapidly amongst iNTS isolates, which are now listed by the World Health Organization as priority antimicrobial-resistant pathogens.^5^ iNTS disease typically presents as a febrile illness with clinical signs of sepsis, often without gastrointestinal symptoms.^6^ Susceptibility is strongly linked with host immunosuppression, with risk factors including malaria, chronic anaemia (notably sickle cell disease) and malnutrition in children, alongside HIV infection and malignancy in adults.^6^ The case-fatality rate is high, estimated at 15–20%.^7^

In Africa, *S*. Typhimurium sequence type 313 (ST313) dominates invasive disease, whilst ST19 predominantly causes gastrointestinal illness in high-income settings.^8–10^ ST313 has undergone stepwise genomic degradation, including loss of genes linked to gut colonization,^11,12^ alongside distinctive prophage remodelling and other host-adaptive changes, suggesting evolution toward a systemic niche and enhanced immune evasion.^13,14^ However, the relative contributions of host immunosuppression and pathogen adaptation to invasive disease remain unknown.

Several iNTS vaccine candidates are in development, including O-antigen conjugates and outer-membrane vesicle-like platforms, with early clinical trials demonstrating encouraging immunogenicity.^15,16^ However, correlates of protection remain undefined, and phase 3 efficacy trials are operationally demanding, requiring large multi-centre designs in settings with focal epidemiology and limited surveillance.^17,18^ Controlled human infection models (CHIMs) offer a means to address both challenges, providing experimental evidence on correlates of protection and early proof-of-concept efficacy signals to de-risk phase 3 trial design, as demonstrated by the *Salmonella* Typhi CHIM that supported WHO prequalification of a Vi-tetanus toxoid conjugate vaccine^19^ and a paratyphoid CHIM demonstrating efficacy of a live-attenuated vaccine.^20^

Here, we report the development of a new *S*. Typhimurium controlled human infection model designed to address two interlinked questions. Firstly, whether the genomic, transcriptomic, and immune-evasive adaptations that distinguish African ST313 from globally distributed ST19 translate into divergent disease in humans, as more than a decade of preclinical work has predicted. Secondly, whether such a model can establish the clinical and immunological benchmarks needed to identify correlates of protection and accelerate evaluation of the iNTS vaccine candidates now entering clinical development. We conducted a Bayesian dose-escalation challenge study of healthy adults with two representative pathovariants (D23580 of the ST313 lineage and 4/74 of the ST19 lineage), characterising the clinical, microbiological, and immunological responses to define a platform that simultaneously interrogates *Salmonella* host–pathogen biology and provides the infrastructure for iNTS vaccine evaluation.

## Methods

### Study design and participants

The Challenge Non-typhoidal *Salmonella* (CHANTS) study was a phase 1, randomised, double-blind, dose-escalation controlled human infection model. Healthy UK-resident adults aged 18–50 years with no documented history of *Salmonella* infection were eligible. Full inclusion and exclusion criteria have been reported previously^21^ and are provided in the study protocol. Volunteers provided written informed consent and underwent structured screening to confirm absence of clinically significant systemic disease or key risk factors for invasive disease, including immunocompromised status, recent malaria, or sickle cell disease. All participants had abdominal ultrasonography to exclude gallstones or biliary pathology that might predispose to chronic carriage. Baseline anti-*Salmonella* serology was measured for exploratory purposes; results were blinded to enrolling physicians and did not inform eligibility. The study was conducted in inpatient and outpatient settings within Imperial College Healthcare NHS Trust. Approvals were obtained from the NHS Health Research Authority London-Fulham Research Ethics Committee (21/PR/0051; IRAS 301659). The trial is registered with ClinicalTrials.gov (NCT05870150) and is complete.

### Challenge strains

Participants received oral challenge with one of two strains of *S*. Typhimurium. Strain 4/74 (ST19; NCTC 14672), first isolated in 1974, is a globally distributed ST19 lineage associated with enterocolitis. Strain D23580 (ST313; NCTC 14677) was isolated in 2004 from a blood culture from a 24-month-old HIV-negative child in Blantyre, Malawi, and is a representative ST313 Lineage 2 isolate associated with bloodstream infection in sub-Saharan Africa. Strain selection, GMP manufacture, formulation, and characterisation have been reported previously.^22^

### Randomisation and masking

Randomisation was performed pre-challenge by computer-generated sealed envelope with permuted block sizes of two or four and 1:1 allocation. The first 10 participants comprised a sentinel safety cohort. The study was conducted double-blind from randomisation until unblinding after the last participant completed the day 28 visit. A locked allocation list was held by the study statistician and the unblinded laboratory team responsible for challenge agent preparation. The unblinded investigator who prepared challenge agents performed no further clinical procedures.

### Study Procedures

Participants were admitted to single-occupancy rooms in a hospital-based quarantine facility on day 0, remaining inpatient until day 7, with daily outpatient follow-up to day 14, then visits at days 28, 90, 180, and 365. Participants fasted for at least 120 minutes before challenge on day 0. Two minutes before challenge, a sodium bicarbonate solution (2·6g in 120ml water) was administered to neutralise gastric acid, followed by the challenge inoculum as a 30ml suspension in 0·9% saline. The administered dose was confirmed by triplicate plating. Dose escalation proceeded sequentially: five participants per strain at 1–5×10³ CFU, five per strain at 1–5×10^4^ CFU, and 15 per strain at 1–5×10^5^ CFU.

Safety was assessed with structured daily monitoring from challenge through at least day 14, including daily physician review, minimum four-times-daily vital signs, 24-hour nursing care during inpatient days, and continuous access to an on-call study physician. Safety bloods were obtained on alternate days, and daily for five days after meeting primary outcome criteria.

Solicited symptoms were recorded in daily diaries on a 5-point ordinal scale (grade 0 to grade 4). Daily symptom severity scores were calculated by summing the total across individually recorded symptom components. Bowel habits were recorded using the Bristol Stool Chart including frequency, consistency, and presence of visible blood or mucus. Daily diarrhoea severity scores were calculated by summing the total number of loose/liquid bowel movements in 24 hours.

Blood samples (10ml) were collected daily into aerobic culture bottles (day 0–14) and processed by automated blood culture. Stool was collected daily (day 0–14) and at days 28, 90, 180, and 365, processed in parallel by direct faecal PCR and selective enrichment culture. Sample processing was performed in the Department of Microbiology, North-West London Pathology, in accordance with national guidelines.^23–25^ Blood PCR for *Salmonella* detection was performed on 10ml of heparinised peripheral venous blood collected daily for seven days post-challenge, using quantitative PCR targeting the *Salmonella*-specific *ttrA* gene. Full protocols are provided in the supplementary appendix pp. 5–7.

Additional blood, saliva, and stool samples were collected at baseline and follow-up visits to measure plasma cytokines (IFN-γ, IL-1β, IL-2, IL-4, IL-6, IL-8, IL-10, IL-12p70, IL-13, TNF-α) by multiplex immunoassay (MSD V-PLEX), and faecal calprotectin and lactoferrin by ELISA. Antigen-specific serum IgG, IgA, and IgM, and salivary and faecal IgA, were measured against *S*. Typhimurium O-antigen (O-Ag), outer-membrane protein D (OmpD), and lipopolysaccharide (LPS) by ELISA; seroconversion was defined as a ≥4-fold rise from baseline to day 28. Serum bactericidal activity (SBA) was measured by a luminescence-based assay against each participant’s challenge strain. Full assay protocols are provided in the supplementary appendix pp. 5–7.

Antibiotics, active against both strains (first-line oral azithromycin 500 mg once daily or ciprofloxacin 500 mg twice daily for bloodstream infection), were started on meeting the primary endpoint or other pre-specified treatment criteria. An independent data safety monitoring committee oversaw the trial and were provided with interim unblinded reports following each cohort of 5 participants (per strain) to inform dose escalation or de-escalation decisions.

### Outcomes

The primary outcome was *Salmonella* diagnosis (SD), defined as fever ≥38·0°C on ≥2 occasions ≥12 hours apart and/or bacteraemia. Participants who received antibiotics before day 14 without meeting SD criteria were classified as antibiotic-treated without *Salmonella* diagnosis (AT-nSD) and excluded from the primary attack rate denominator. Pre-specified secondary outcomes included gastrointestinal colonisation (*Salmonella*-positive stool culture on ≥2 occasions ≥48 hours after challenge), gastroenteritis (≥3 loose or liquid stools in 24 hours), symptom and diarrhoea severity, inflammatory biomarkers, and antigen-specific antibody responses. For time-to-event analyses, event time was the time of blood culture collection for culture-confirmed cases, or the first oral temperature ≥38·0°C that subsequently persisted ≥12 hours for fever-defined cases; participants without an event were censored at day 14. Baseline immunological correlates of clinical outcome were a pre-specified exploratory analysis.

### Statistical analysis

No formal sample size calculation was performed and up to 80 participants (40 per strain) was a feasible target. Dose-finding used a model-based continual reassessment method (CRM) to model the dose– attack-rate relationship for each strain and guide escalation alongside the safety profile, with the dose yielding a predicted attack rate closest to 67·5% selected in the absence of safety concerns. To our knowledge this is the first CHIM to use CRM rather than rule-based escalation. The study opened at 1– 5×10³ CFU with permitted escalation to 1–5×10^6^ CFU, with an early-stopping rule once the model-recommended dose had accrued n=15. The primary attack rate was the proportion meeting SD, with model-estimated rates and 95% credible intervals derived by CRM for each strain. Time-to-event outcomes used Kaplan–Meier methods; categorical comparisons used Fisher’s exact test and continuous comparisons the Mann–Whitney U or Wilcoxon signed-rank test, with Benjamini–Hochberg adjustment across marker families. Associations between log_10_-transformed baseline biomarkers and binary outcomes (SD; colonisation) were assessed by logistic regression adjusted for age, sex, dose, and strain, with odds ratios per tenfold increase. Analyses were done in R (v4.5.0). The study was not powered for formal hypothesis testing, and between-strain comparisons were exploratory.

## Results

### Participants

Between Aug 31, 2023, and Dec 13, 2024, we enrolled 50 healthy volunteers (25 per strain) (figure 1). The median age was 27 years (range 18–43), 27 (54%) were women and 23 (46%) were men. Participants were randomly assigned 1:1 to oral challenge with 4/74 (ST19) or D23580 (ST313), with five per strain at 1–5×10³ CFU, five per strain at 1–5×10^4^ CFU, and 15 per strain at 1–5×10^5^ CFU. Baseline characteristics were well matched across strain and dose cohorts (table 1). All doses fell within target ranges with no between-strain differences (appendix pp. 20).

**Figure 1:**
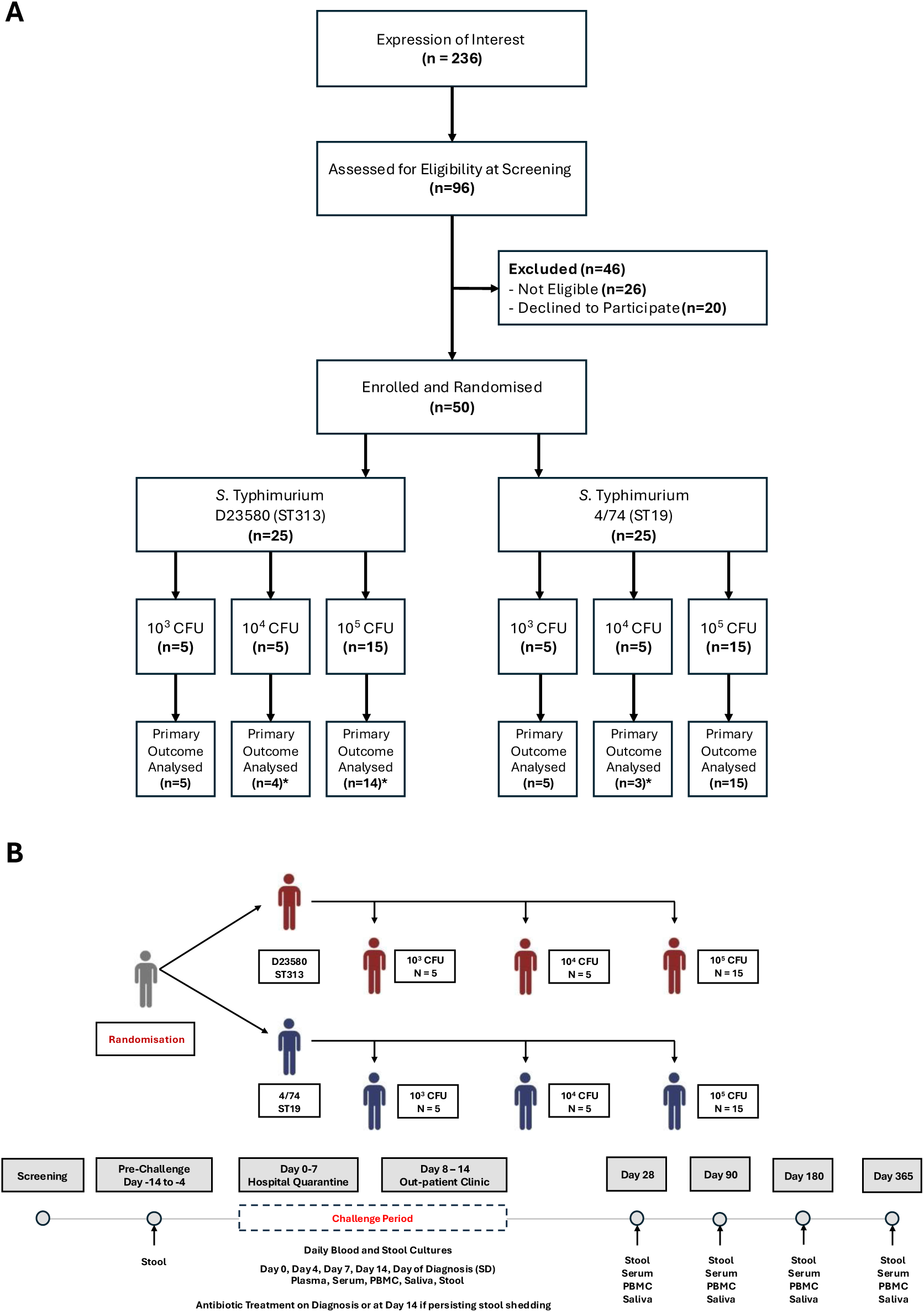
Study design and challenge dose administration. **(A)** CONSORT Diagram. The study was a dose escalation design conducted with continual reassessment method (CRM) modelling. 50 healthy adults were enrolled from n=236 expressions of interest and n=96 who attended screening. Screening exclusion and withdrawals are detailed in the supplementary appendix pp. 14. Participants were randomised 1:1 to challenge with either S. Typhimurium 4/74 or S. Typhimurium D23580 (n=25 per strain). Dose escalation was conducted sequentially with participants receiving 10^3^ CFU (n=10), 10^4^ CFU (n=10), or 10^5^ CFU (n=30). No participants were lost to follow-up, with all being monitored until at least day 28. *Participants excluded from primary outcome analysis (AT-nSD). **(B)** Study flow chart indicating in-patient (day 0-7) and out-patient (day 8-365) study visits with sampling schedules.

**Table 1:** Participant baseline demographics by challenge strain and dose allocation. 1 - Excludes Oral Ty21a – participants reporting prior oral live attenuated vaccination were not eligible for participation.

| Challenge Group |  |  |  |  |  |  |  |
| --- | --- | --- | --- | --- | --- | --- | --- |
|  | All | STm 4/74 (ST19) |  |  | STm D23580 (ST313) |  |  |
|  |  | 10<sup>3</sup> CFU | 10<sup>4</sup> CFU | 10<sup>5</sup> CFU | 10<sup>3</sup> CFU | 10<sup>4</sup> CFU | 10<sup>5</sup> CFU |
| Participant Baseline Characteristics |  |  |  |  |  |  |  |
| Participants | 50 | 5 | 5 | 15 | 5 | 5 | 15 |
| Male sex, n (%) | 23 (46%) | 4 (80%) | 2 (40%) | 8 (53·3%) | 2 (40%) | 2 (40%) | 5 (33·3%) |
| Female sex n(%) | 27 (54%) | 1 (20%) | 3 (60%) | 7 (46·7%) | 3 (60%) | 3 (60%) | 10 (66·7%) |
| Age, Years, Median (range) | 27 (18, 43) | 25 (21, 41) | 25 (19, 28) | 27 (19, 39) | 27 (25, 35) | 27 (22, 43) | 24 (18, 35) |
| Ethnicity, n (%) |  |  |  |  |  |  |  |
| White (British) | 19 (38%) | 0 (0%) | 4 (80%) | 6 (40%) | 3 (60%) | 1 (20%) | 5 (33·3%) |
| White (Other) | 12 (24%) | 3 (60%) | 1 (20%) | 3 (20%) | 2 (40%) | 1 (20%) | 2 (13·3%) |
| Asian | 7 (14%) | 2 (40%) | 0 (0%) | 2 (13·3%) | 0 (0%) | 1 (20%) | 2 (13·3%) |
| Black | 6 (12%) | 0 (0%) | 0 (0%) | 1 (6·7%) | 0 (0%) | 2 (40%) | 3 (20%) |
| Hispanic | 1 (2%) | 0 (0%) | 0 (0%) | 1 (6·7%) | 0 (0%) | 0 (0%) | 0 (0%) |
| Mixed | 5 (10%) | 0 (0%) | 0 (0%) | 2 (13·3%) | 0 (0%) | 0 (0%) | 3 (20%) |
| BMI (kg/m<sup>2</sup>), Median (range) | 23·1 (18·6, 37·6) | 22·9 (20·4, 28·6) | 22 (21·8, 25·5) | 23·4 (18·6, 36·3) | 28·2 (23·2, 37·6) | 24·9 (18·6, 30·6) | 22·7 (18·9, 36·5) |
| Previous Typhoid Vaccination, n (%)<sup>1</sup> | 9 (18%) | 0 (0%) | 1 (20%) | 3 (20%) | 0 (0%) | 0 (0%) | 5 (33·3%) |

### Primary endpoint, dose-response, and safety

A clear dose-response was observed. No participants met the primary SD endpoint at 10³ CFU. This increased to 2/10 at 10^4^ CFU (both D23580) and 16/30 at 10^5^ CFU (8/15 per strain). Of 18 participants meeting the SD endpoint, 17 (94%) did so through sustained fever. CRM-estimated attack rates at 10^5^ CFU were 57·9% (95% credible interval 37·3–73·8) for D23580 and 47·4% (26·7–65·7) for 4/74 (figure 2). Four participants received antibiotics without meeting SD criteria (AT-nSD; three at 10^4^ CFU, one at 10^5^ CFU) and were excluded from the primary attack rate denominators. Sensitivity analyses reclassifying AT-nSD participants as events gave consistent results (D23580 61·8% [42·3– 76·4]; 4/74 54·3% [34·6–70·4]; appendix pp. 11).

**Figure 2:**
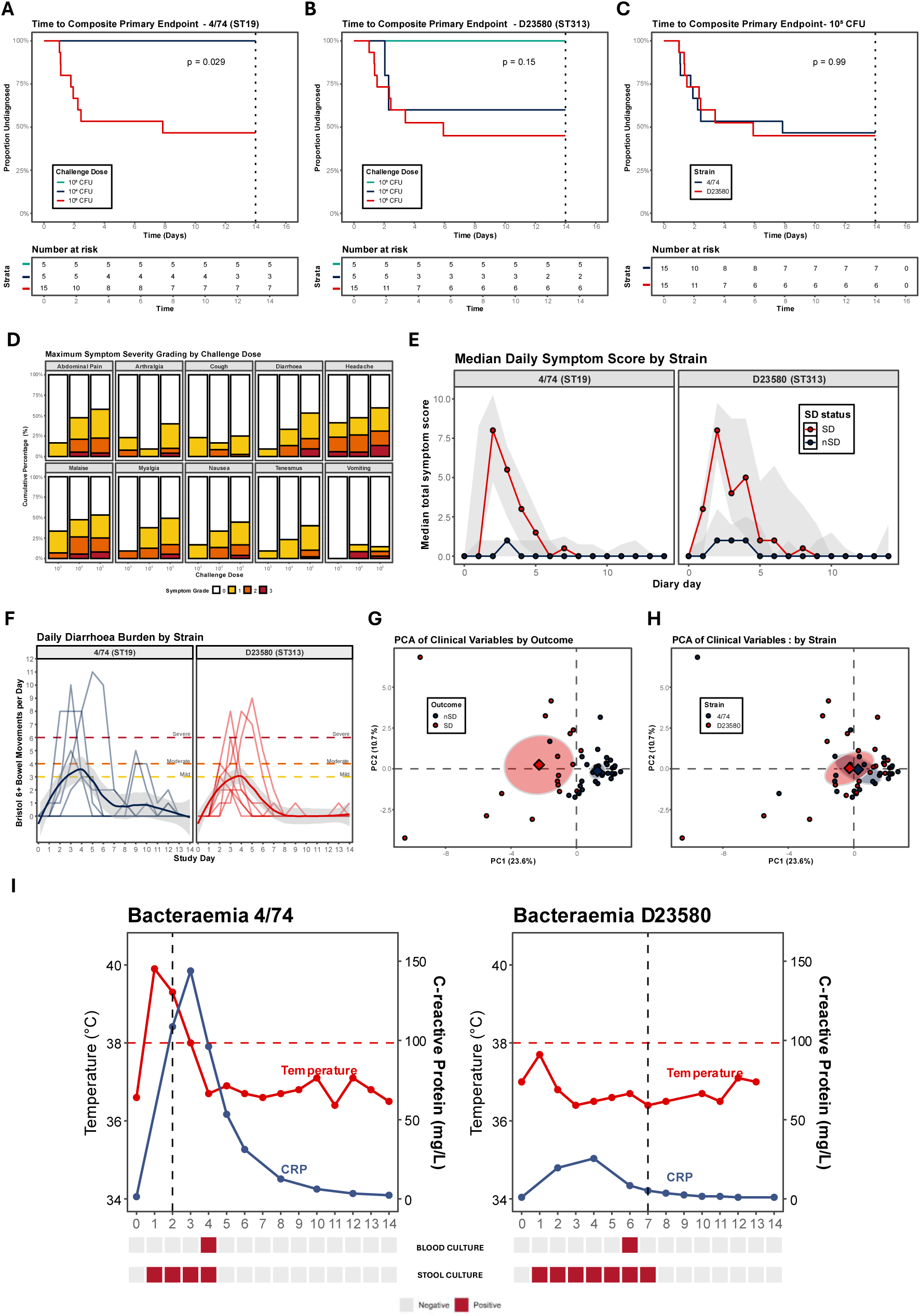
Primary outcome rates and clinical profiles following challenge. Kaplan–Meier curves show cumulative incidence of the primary outcome (fever ≥38.0°C for ≥12h and/or *S.* Typhimurium bacteraemia) across dose levels. Participants not meeting criteria were censored at antibiotic initiation (AT-nSD) or at day 14 (no event). (**A**) STm 4/74 (ST19); (**B**) STm D23580 (ST313); (**C**) strain comparison at 10^5^ CFU (log-rank test). (**D**) Maximum symptom severity scores across dose levels based on 10 solicited symptoms recorded daily (days 0–14). (**E**) Median daily symptom severity by strain and outcome (IQR shaded). (**F**) Daily diarrhoea burden (Bristol stool types 6/7) by strain, with individual trajectories and LOESS smoothing. Dashed lines indicate thresholds for mild, moderate, and severe gastroenteritis. (G–H) PCA of normalised clinical variables, coloured by outcome (**G**) or strain (**H**). Ellipses show 95% confidence intervals; diamonds mark centroids; axes indicate variance explained. (**I**) Clinical trajectories of two bacteraemia cases (#13, #48): D23580 was asymptomatic, whereas 4/74 was associated with fever and severe disease. Red dashed lines indicate fever threshold (≥38.0°C); black dashed lines indicate antibiotic initiation; red boxes = positive cultures, grey = negative.

No serious adverse events occurred. Per-protocol severe salmonellosis was observed in five participants (three D23580, two 4/74), arising from grade 4 laboratory abnormalities (n=2) or gastrointestinal bleeding associated with bloody diarrhoea (n=3); all events were self-limiting and resolved without complication (appendix pp. 15).

Dose escalation to 10^6^ CFU was not undertaken. Following review of unblinded safety and clinical data after the 10^5^ CFU cohort, the DSMC determined that the risk-benefit balance did not support further escalation. Observed attack rates at 10^5^ CFU were below the pre-specified 60–75% target. Despite this, 10^5^ CFU produced dose-dependent disease sufficient to address the primary study objectives and was therefore considered the maximum feasible dose within acceptable safety constraints. CRM modelling projected attack rates at 10^6^ CFU of 70·2% (95% credible interval 52·8–82·1) for D23580 and 61·7% (42·5–76·1) for 4/74, though these values are extrapolations rather than observed outcomes.

### Clinical phenotype and pathovariant comparison

Participants typically remained asymptomatic for the first 48 hours. In those who developed SD, symptoms peaked at days 2–3 across expected domains, including headache, diarrhoea, abdominal pain, and malaise, with a clear dose-response in frequency and severity (figure 2; appendix pp. 21). Systemic symptoms were frequently accompanied by acute watery diarrhoea (figure 2; appendix pp. 23). Overall, 15/50 participants (30%) met the case definition for *Salmonella* gastroenteritis, with no episodes at 10³ CFU, 2/10 (20%) at 10^4^ CFU, and 13/30 (43·3%) at 10^5^ CFU. The severity of gastroenteritis also increased with dose, with maximum stool frequency rising from five loose or liquid stools in 24 hours at 10^4^ CFU to 15 at 10^5^ CFU (table 2; appendix pp. 23). Routine haematology and biochemistry were comparable across strains (appendix pp. 25). Symptom type, severity, and duration, and gastroenteritis frequency and severity, were comparable between pathovariants across all dose levels. Disease phenotype, explored by principal component analysis (PCA) incorporating clinical features, solicited symptoms, and laboratory parameters, separated participants by disease outcome but not by challenge strain (figure 2; appendix pp. 26).

**Table 2:** Challenge dose administered, primary and secondary outcomes frequencies by challenge strain and dose allocation. 1 - Participants receiving antibiotic therapy prior to day 14 without meeting primary outcome criteria were excluded from primary outcome analyses (AT-nSD). 2 - Colonisation = 2 or more stool culture samples positive ≥48 hours from challenge. 3 - Gastroenteritis = ≥3 loose/liquid stool in 24h (Bristol Type ≥6); Mild ≥3 loose/liquid stool, Moderate 4-5 loose/liquid stool, Severe ≥6 loose/liquid stool. CRM: continual reassessment method.

|  | Challenge Group |  |  |  |  |  |  |
| --- | --- | --- | --- | --- | --- | --- | --- |
|  | All | STm 4/74 (ST19) |  |  | STm D23580 (ST313) |  |  |
|  |  | 10<sup>3</sup> CFU | 10<sup>4</sup> CFU | 10<sup>5</sup> CFU | 10<sup>3</sup> CFU | 10<sup>4</sup> CFU | 10<sup>5</sup> CFU |
| Participants | 50 | 5 | 5 | 15 | 5 | 5 | 15 |
| **Challenge Dose Administered** |  |  |  |  |  |  |  |
| Challenge Dose (CFU), Median (range) | - | 2500 (2300, 2700) | 38,000 (38,000, 42,000) | 330,000 (270,000 – 440,000) | 2200 (2000 – 2900) | 35,000 (35,000 – 48,000) | 340,000 (300,000 – 500,000) |
| **Primary Outcome** |  |  |  |  |  |  |  |
| Included in Primary Outcome Analysis<sup>1</sup> | 46 | 5 | 3 | 15 | 5 | 4 | 14 |
| *Salmonella* Diagnosis (SD), n (% attack rate), 95% CI | 18 (39.1%) (25.1 – 54.6) | 0 (0%) (0 – 52.2) | 0 (0%) (0 – 70.8) | 8 (53.3%) (26.6 – 78.7) | 0 (0%) (0 – 52.2) | 2 (50%) (6.8 – 93.2) | 8 (57.1%) (28.9 – 82.3) |
| SD = Fever | 17 | 0 | 0 | 8 | 0 | 2 | 7 |
| SD = Bacteraemia | 1 | 0 | 0 | 0 | 0 | 0 | 1 |
| CRM Model Predicted Attack Rate, 95% CI | - | 9.6% (1.6 – 26.6) | 26.7% (9.6 – 47.5%) | 47.4% (26.7 – 65.7) | 17.9% (4.5 – 38.5) | 38.0% (17.5 – 58.5) | 57.9% (37.3 – 73.8) |
| **Secondary Outcomes** |  |  |  |  |  |  |  |
| Included in Secondary Outcome Analyses | 50 | 5 | 5 | 15 | 5 | 5 | 15 |
| **Colonisation** |  |  |  |  |  |  |  |
| *Salmonella* Colonisation Rate, n (% total)<sup>2</sup> | 39 (78.0%) | 4 (80.0%) | 4 (80.0%) | 13 (86.7%) | 2 (40.0%) | 3 (60.0%) | 13 (86.7%) |
| Any Stool Culture Positive, n (%total) | 47 (94.0%) | 5 (100%) | 4 (80.0%) | 15 (100%) | 4 (80.0%) | 4 (80.0%) | 15 (100%) |
| **Gastroenteritis** |  |  |  |  |  |  |  |
| *Salmonella* Gastroenteritis Rate, n (% total)<sup>3</sup> | 15 (30.0%) | 0 (0%) | 1 (20.0%) | 6 (40.0%) | 0 (0%) | 1 (20.0%) | 7 (46.7%) |
| Mild Gastroenteritis | 15 (30.0%) | 0 (0%) | 1 (20.0%) | 6 (40.0%) | 0 (0%) | 1 (20.0%) | 7 (46.7%) |
| Moderate Gastroenteritis | 13 (26.0%) | 0 (0%) | 1 (20.0%) | 6 (40.0%) | 0 (0%) | 1 (20.0%) | 5 (33.3%) |
| Severe Gastroenteritis | 10 (20.0%) | 0 (0%) | 0 (0%) | 5 (33.3%) | 0 (0%) | 1 (20.0%) | 4 (26.7%) |

Bacteraemia was detected by daily blood culture in two participants, one from each challenge group, at 10^5^ CFU. The participant challenged with D23580 developed an afebrile bacteraemia on day 6 that cleared spontaneously by day 7 without antibiotics, whereas the 4/74 participant had already met SD criteria on day 2 through fever with raised inflammatory markers and severe gastroenteritis, with bacteraemia detected on day 4 (figure 2). No additional cases were identified by *ttrA*-targeted PCR on pre-enriched blood samples.

Plasma cytokines distinguished disease outcome but not strain: SD participants showed significant upregulation of IFN-γ, TNF-α, IL-6, IL-12p70, IL-8, and IL-10 compared with nSD, with no differential signature between pathovariants. Faecal calprotectin and lactoferrin rose transiently and returned toward baseline by day 7 with no inter-strain difference. PCA incorporating plasma cytokines and faecal markers again clustered by disease outcome but not by strain (figure 3).

**Figure 3:**
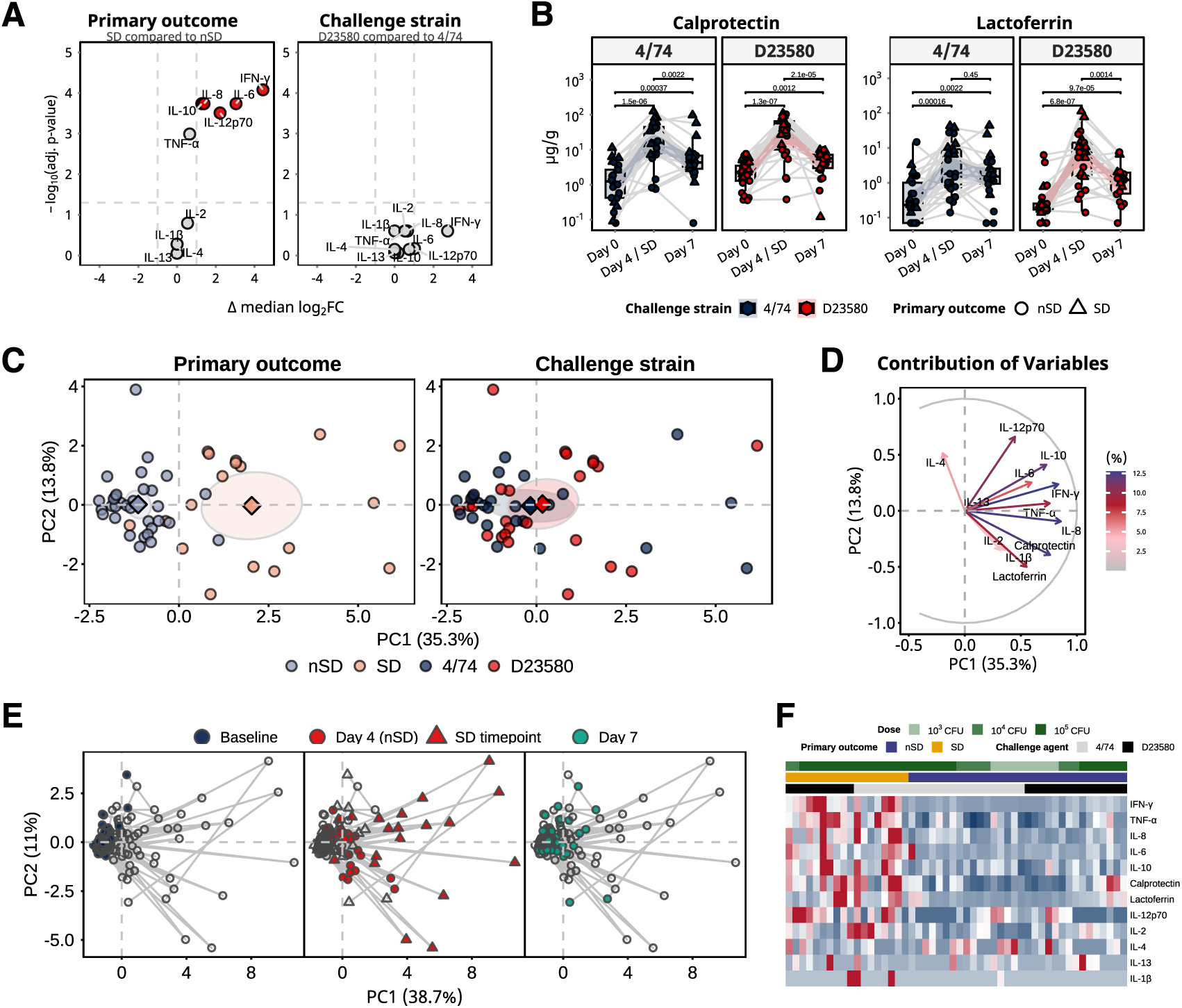
Inflammatory biomarkers in blood and stool following Salmonella Typhimurium challenge. (**A**) Volcano plots of log₂ fold-change in plasma cytokines at diagnosis (SD) or day 4 (nSD). Right panel compares strains 4/74 and D23580. Dashed lines indicate twofold change and adjusted p=0.05 (Mann– Whitney). (**B**) Faecal calprotectin and lactoferrin (µg g⁻^1^) measured by ELISA following challenge with 4/74 (blue) or D23580 (red). Circles = nSD; triangles = SD. Boxplots show median, IQR, and 1.5×IQR whiskers. Within-strain comparisons used Wilcoxon signed-rank tests. (**C**) PCA of normalised plasma cytokines and faecal markers, coloured by outcome (left) or strain (right). Ellipses show 95% confidence intervals; axes indicate variance explained; diamonds mark centroids. (**D**) Variable contributions to PC1 and PC2 (% variance). (**E**) PCA trajectories across baseline (day 0, blue), day 4/SD (red), and day 7 (green); grey lines connect individuals. (**F**) Heatmap of normalised inflammatory profiles (AUC, day 0–7). Rows = markers; columns = participants annotated by dose, outcome, and strain. nSD = no diagnosis; SD = *Salmonella* diagnosis; IQR = interquartile range.

### Gastrointestinal colonisation

Overall, 39/50 participants (78%) met colonisation criteria (*Salmonella*-positive stool culture on ≥2 occasions ≥48 hours after challenge), with a clear dose-response (6/10 [60%] at 10³ CFU; 7/10 [70%] at 10^4^ CFU; 26/30 [87%] at 10^5^ CFU; figure 4). Colonisation rates were comparable between strains at all dose levels (figure 4; table 2). Shedding patterns were heterogeneous across both strains (median 5 days, range 0–13), with positive cultures in most participants within 48 hours. Relapse, defined as recurrent shedding after clearance, occurred in 5/50 participants (10%) and was most often detected at day 28. All relapses were asymptomatic and required re-treatment per protocol. One participant remained colonised at day 180, achieving spontaneous clearance by day 365 (figure 4).

**Figure 4:**
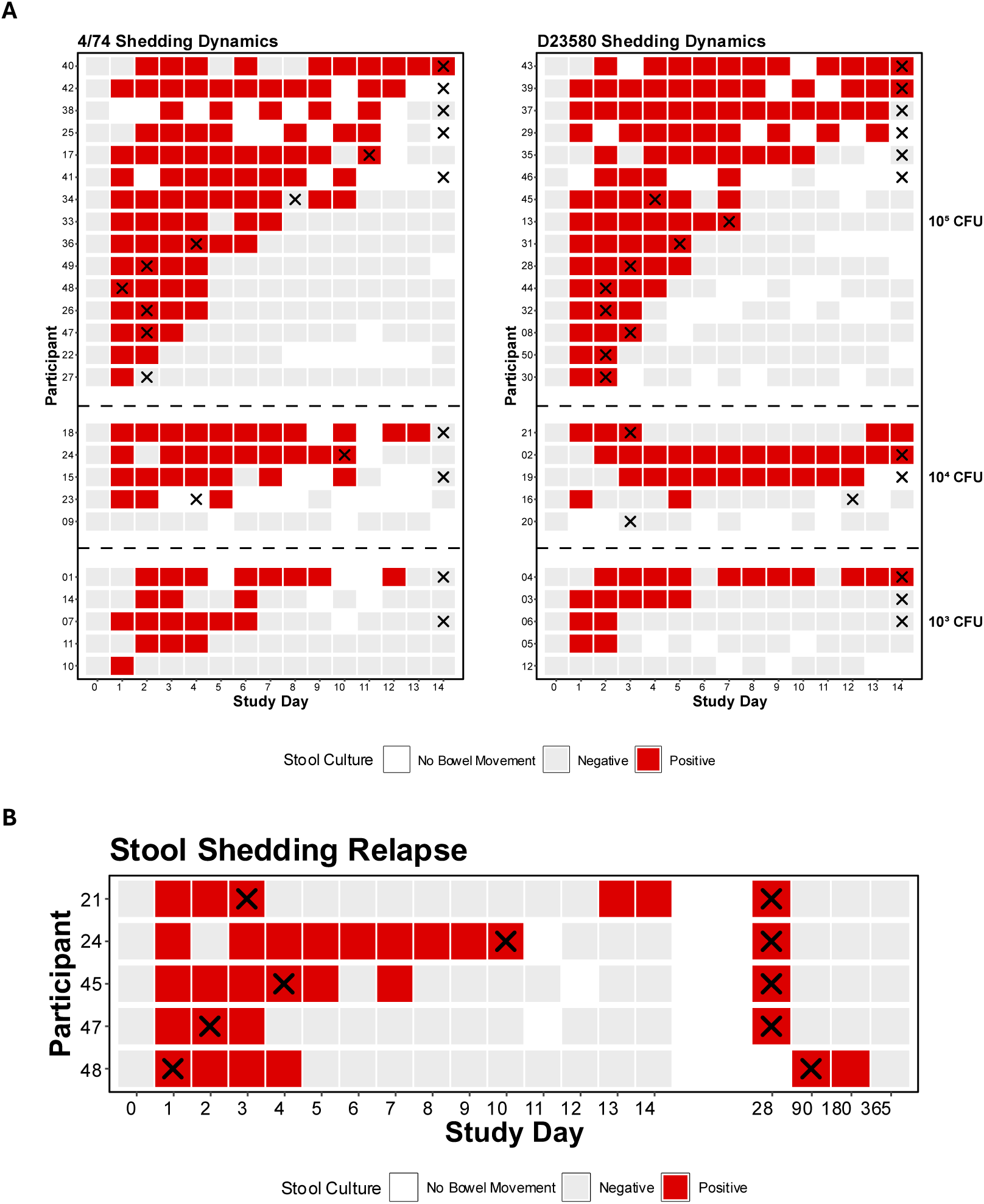
Gastrointestinal colonisation dynamics. (**A**) STm 4/74 (ST19) and (**B**) STm D23580 (ST313) stool shedding dynamics post-challenge by allocated challenge dose. Red boxes indicate stool culture positive, grey = negative, white = no bowel movement. Black crosses indicate day of antibiotic therapy for individuals receiving treatment. (**C**) Shedding patterns for 5 individuals who experienced asymptomatic relapse of stool shedding following an initial course of antibiotic therapy. Second black crosses indicate secondary courses of therapy. Participant identification numbers have been anonymised.

### Serological responses and humoral correlates of susceptibility

Antigen-specific antibodies increased significantly by day 28, with fold changes exceeding fourfold across most antigen-isotype combinations (figure 5; appendix pp. 30-31). At 10^5^ CFU, anti-O-antigen IgG seroconversion occurred in 80% of 4/74 and 93% of D23580 participants. Anti-O-antigen IgG, anti-OmpD IgG, and SBA IC_50_ all remained elevated from baseline through day 180. Significant increases in anti-LPS IgA were observed in serum, saliva, and stool across both challenge groups. Participants meeting SD showed markedly greater fold changes across IgG, IgM, and SBA compared with nSD participants (figure 5).

**Figure 5:**
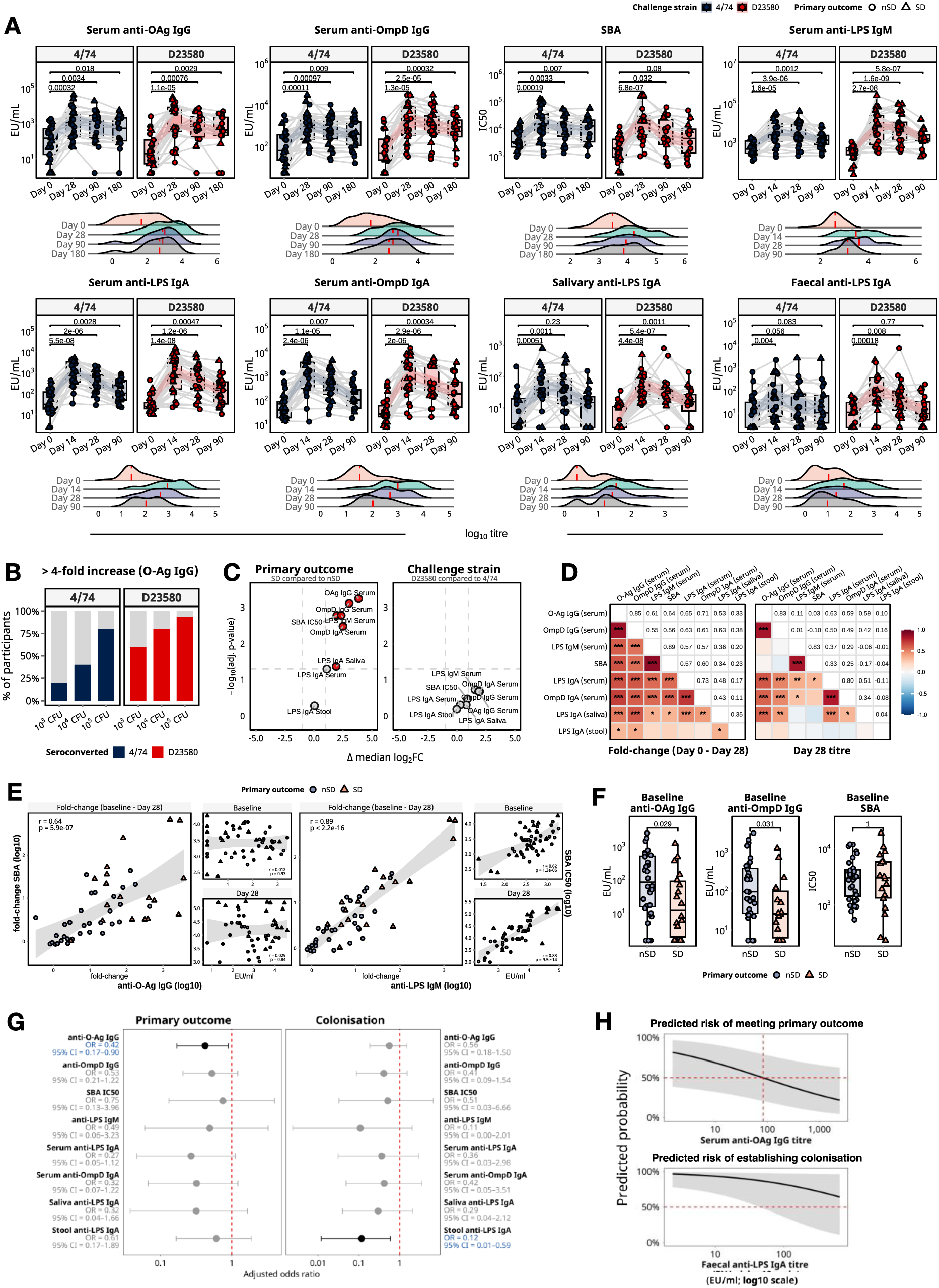
Antibody responses following controlled human infection with two strains of *Salmonella* Typhimurium. (A) Serum antigen-specific IgG, IgM, and IgA titres (EU mL⁻^1^) and serum bactericidal activity (SBA IC₅₀) were measured longitudinally (days 0, 14, 28, 90, 180) following challenge with strain 4/74 (blue) or D23580 (red). Participants with primary endpoint (SD) are triangles; others (nSD) circles. Boxplots show medians and IQRs; grey lines indicate individual trajectories. Within-strain comparisons used two-sided Wilcoxon tests; kernel density plots summarise temporal shifts. (B) Seroconversion (≥4-fold rise to day 28) for anti-O-antigen IgG by strain and dose. (C) Volcano plots of log₂ fold-change comparing SD vs nSD (left) and strains (right); dashed lines indicate twofold change and adjusted p=0.05. (D) Pairwise correlations of fold-change (left) and day 28 titres (right), coloured by Pearson r with significance indicated (*p<0·05,**p<0·01,***p<0·001). (E) Correlations between fold-change antibody responses and SBA; insets show baseline and day 28 titres (95% CI shaded). (F) Baseline titres compared by outcome (Mann–Whitney). (G) Adjusted univariate logistic regression of immune correlates of outcome and colonisation. (H) Model-predicted risk by antibody titre. IQR = interquartile range; SBA = serum bactericidal activity; FC = fold-change; nSD = no *Salmonella* diagnosis; SD = *Salmonella* diagnosis.

Participants who did not develop SD had significantly higher baseline serum anti-O-antigen IgG titres than those who met the primary endpoint (figure 5). After adjusting for age, sex, dose, and strain, each tenfold rise in baseline anti-O-antigen IgG corresponded to a 58% reduction in SD risk (odds ratio 0·42, 95% CI 0·17–0·90), whereas baseline SBA showed no association (0·75, 0·13–3·96). In parallel, each tenfold rise in baseline faecal anti-LPS IgA corresponded to an 88% reduction in colonisation risk (0·12, 0·01–0·59; figure 5). Serum anti-LPS IgM correlated strongly with SBA at baseline (r=0·62; p<0·0001) and day 28 (r=0·83; p<0·0001), independent of outcome or strain, whereas anti-O-antigen IgG showed no correlation with SBA (figure 5).

## Discussion

In this new *S*. Typhimurium controlled human infection model, we established a safe and reproducible model of colonisation, diarrhoeal disease, and fever in healthy adults, with a maximum feasible challenge dose of 10^5^ CFU producing systemic disease in approximately half of participants without serious adverse events. We observed no clinical, microbiological, or immunological difference between challenge with the African ST313 lineage strain D23580 and the globally distributed ST19 strain 4/74. Higher baseline serum anti-O-antigen IgG was associated with reduced systemic disease, and higher baseline faecal anti-LPS IgA with reduced gastrointestinal colonisation, providing experimental support in humans for correlates of susceptibility previously inferred from epidemiological observation.

Deliberate administration of *S*. Typhimurium to volunteers has been reported previously: dose-finding studies between 1936 and 1951 exposed participants to uncharacterised inocula without standardised endpoints or quantified attack rates^26–30^, and two later studies administered attenuated strains as candidate vaccine vectors rather than to reproduce disease^31,32^. This is therefore the first contemporary *S*. Typhimurium human infection model to use characterised GMP-manufactured inocula, pre-specified clinical endpoints, and a Bayesian dose-escalation design.

The primary objective of the dose-escalation phase was to define a challenge dose for *S.* Typhimurium. Using a Bayesian continual reassessment method, we mapped a clear dose–response. The pre-specified 60–75% attack rate was a pragmatic operational target, chosen for consistency with established enteric challenge models^19^ and to minimise future control-group sample sizes, rather than a criterion of model validity. At the maximum feasible dose of 10^5^ CFU, CRM-estimated attack rates approached rather than exceeded this target, at 57·9% (37·3–73·8) for D23580 and 47·4% (26·7–65·7) for 4/74. We regard this as a workable rather than an optimised attack rate, sufficient for early proof-of-concept vaccine evaluation at the cost of a modestly larger control group. We did not escalate to 10^6^ CFU because, although no serious adverse events occurred, symptom frequency and severity rose steeply with dose and the DSMC determined that the safety profile at 10^5^ CFU did not support further escalation; CRM projections suggested only modest additional gains would have been achieved.

The absence of a measurable phenotypic difference between ST313 and ST19 contrasts with more than a decade of *in vitro*, *in vivo*, and genomic data predicting an enhanced ability of D23580 to cause bloodstream infection.^33,34^ One interpretation is that the adaptive features of D23580 confer no virulence advantage in immunocompetent hosts, and that the ST313 invasive phenotype seen in sub-Saharan Africa reflects a combination of pathovariant-specific adaptation and host immunosuppression, with ST313-associated traits having more apparent consequences in individuals with recognised risk factors for invasive disease. Although the small number of bacteraemia cases (one per strain) precludes formal comparison, the contrasting clinical phenotypes – a silent, self-clearing bacteraemia with D23580 and a bacteraemia arising from fever and severe gastroenteritis with 4/74 – raise the possibility that ST313- associated immune-evasion strategies shape disease presentation without increasing the frequency of bloodstream infection in immunocompetent hosts.

Our serological findings indicate prior exposure to *Salmonella* in this population, and individuals with higher baseline serum anti-O-antigen IgG were less likely to develop clinical disease. Causality cannot be inferred from this association alone, given the small sample size and the observation that serum IgG responses across other surface antigens showed similar trends – for example, baseline anti-OmpD IgG showed a directionally consistent but non-significant association, compatible with co-recognition of structurally linked surface epitopes. Serum anti-O-antigen IgG likely promotes early opsonophagocytic clearance^35^, and the correlation of SBA with anti-LPS IgM but not IgG suggests IgM plays a greater role in cell-free killing. Although SBA has been epidemiologically linked with reduced iNTS risk in African populations^36^, no protective association was observed here, possibly reflecting greater importance of cellular clearance mechanisms. A tenfold increase in baseline faecal anti-LPS IgA corresponded to an 88% reduction in colonisation risk independent of dose, strain, age, and sex, suggesting that mucosal IgA contributes to *Salmonella* colonisation resistance in humans, plausibly by restricting luminal replication and epithelial attachment^37^.

We acknowledge the limitations of our study. It was conducted in healthy UK adults, a population distinct from those most affected by iNTS disease in sub-Saharan Africa. We examined only two pathovariants and cannot extrapolate to *S*. Enteritidis, the other major cause of iNTS globally. The trial was not powered to detect between-strain differences, although none were identified. Antibiotic therapy, applied per pre-trial expert consensus to safeguard participants^38^, may have affected carriage and immune responses, although robust seroconversion was observed. Participant diet was not controlled which may influence colonisation resistance^39^. Asymptomatic stool relapse occurred in 5/50 participants (10%), comparable with convalescent shedding after natural NTS infection – where pooled estimates show approximately 14% of asymptomatic individuals remain stool-culture positive at 8 weeks and 4% at 12 weeks^40^ – and was detectable only through protocolised surveillance not undertaken in routine clinical practice. These constraints are intrinsic to new challenge studies in immunocompetent volunteers and define the safety ceiling for further dose escalation.

In summary, this safe and reproducible controlled human infection model of *S*. Typhimurium establishes the experimental infrastructure needed to bridge early-phase immunogenicity studies and field efficacy trials for the iNTS vaccine candidates now in clinical development. By delivering controlled clinical and immunological endpoints in healthy volunteers, the model provides a route to evaluate correlates of protection and generate early proof-of-concept efficacy signals to inform vaccine licensure pathways. Beyond vaccine evaluation, it offers a tractable system for dissecting *Salmonella* host-pathogen biology in humans, including the contribution of host immunocompetence to the divergent outcomes seen between high-income and iNTS-endemic settings. Translation to populations and pathogens of greatest public-health relevance, including controlled studies in endemic settings and incorporation of host susceptibility factors such as malaria co-infection, is a realistic next step.

## Supporting information

Statistical Analysis Plan

Study Protocol

## Data Availability

Data sets generated and analysed in this study are available from the corresponding author upon reasonable request; no participant-identifiable information will be disclosed. The full study protocol and statistical analysis plan are provided as appendices. Genome sequences for the 4/74 and D23580 challenge strains are available as GenBank accessions CP002487.1 - CP002490.1 (4/74) and LS997973.1 - LS997977.1 (D23580).

## Contributors

Conceptualisation: MMG, MAG, RKMC, GSC, AJP, JCDH. Methodology (participant recruitment, clinical conduct, sample collection, and data collection): MMG, CS, ES, RV, AR. Investigation: CS, RV, MMG, AR, EC, ZK, LJ, AM, ZZ, CZ, RK, KJ, SW, SJ, BPS, XJ, RC, DDS, FM, OR, EC, PWH, MA, OD, JML, GL, VP. Formal analysis: XL, YN, CS. Funding acquisition: MMG, GSC. Supervision: GSC, CC. Project administration: CS, MMG, RV, ES. Writing – original draft: CS, AR. Writing – review and editing: all authors.

## Data Sharing

Data sets generated and analysed in this study are available from the corresponding author; no participant-identifiable information will be disclosed. The full study protocol and statistical analysis plan are provided as appendices. Genome sequences for the 4/74 and D23580 challenge strains are available as GenBank accessions CP002487.1–CP002490.1 (4/74) and LS997973.1–LS997977.1 (D23580).

## Acknowledgements

We thank all participants for their time and commitment to this study. We are grateful to the members of the Data Safety and Monitoring Committee: Thomas Darton (chair), Adam Dale, Daniela Ferreira, Melissa Kapula, and Moreno Ursino. We thank Andreea Badarau, Arturo Ledesma, and all ward staff at 15N Charing Cross Hospital; all staff at the NIHR Imperial Clinical Research Facility; Hugo Donaldson, Manfred Almeida, Sweenie Goonesekera, and the microbiology laboratory staff at North-West London Pathology; and Sonia Vidal, Carolyn Newell, Pete Gardner, and Shobana Balasingam at Wellcome Trust.

## References

1 GBD 2017 Non-Typhoidal Salmonella Invasive Disease Collaborators. The global burden of non-typhoidal salmonella invasive disease: a systematic analysis for the Global Burden of Disease Study 2017. Lancet Infect Dis 2019; 19: 1312–24.

2 Marchello CS, Fiorino F, Pettini E, Crump JA. Incidence of non-typhoidal Salmonella invasive disease: A systematic review and meta-analysis. J Infect 2021; 83: 523–32.

3 Lee J-S, Hwang Y, MacLennan CA, Jit M, Excler J-L, Kim JH. Estimating the economic burden of invasive non-typhoidal Salmonella infections in low- and middle-income countries. BMJ Glob Health 2025; 10. DOI:10.1136/bmjgh-2025-019370.

4 Vongpradith A, Dominguez R-MV, Tudor Car L, et al. Global burden of enteric infectious diseases, diarrhoeal diseases, and corresponding aetiologies, 1990–2023: a systematic analysis for the Global Burden of Disease Study 2023. The Lancet Infectious Diseases 2026; : S1473309926001945.

5 Sati H, Carrara E, Savoldi A, et al. The WHO Bacterial Priority Pathogens List 2024: a prioritisation study to guide research, development, and public health strategies against antimicrobial resistance. Lancet Infect Dis 2025; 25: 1033–43.

6 Feasey NA, Dougan G, Kingsley RA, Heyderman RS, Gordon MA. Invasive non-typhoidal salmonella disease: an emerging and neglected tropical disease in Africa. Lancet 2012; 379: 2489–99.

7 Marchello CS, Birkhold M, Crump JA, et al. Complications and mortality of non-typhoidal salmonella invasive disease: a global systematic review and meta-analysis. The Lancet Infectious Diseases 2022; 3099: 1–14.

8 Okoro CK, Kingsley RA, Connor TR, et al. Intracontinental spread of human invasive Salmonella Typhimurium pathovariants in sub-Saharan Africa. Nat Genet 2012; 44: 1215–21.

9 Langridge GC, Fookes M, Connor TR, et al. Patterns of genome evolution that have accompanied host adaptation in Salmonella. Proc Natl Acad Sci U S A 2015; 112: 863–8.

10 Van Puyvelde S, de Block T, Sridhar S, et al. A genomic appraisal of invasive Salmonella Typhimurium and associated antibiotic resistance in sub-Saharan Africa. Nat Commun 2023; 14: 6392.

11 Kingsley R a, Msefula CL, Thomson NR, et al. Epidemic multiple drug resistant Salmonella Typhimurium causing invasive disease in sub-Saharan Africa have a distinct genotype. Genome research 2009; 19: 2279–87.

12 Pulford CV, Perez-Sepulveda BM, Canals R, et al. Stepwise evolution of Salmonella Typhimurium ST313 causing bloodstream infection in Africa. Nature Microbiology 2020; : 1–12.

13 Aulicino A, Rue-Albrecht KC, Preciado-Llanes L, et al. Invasive Salmonella exploits divergent immune evasion strategies in infected and bystander dendritic cell subsets. Nature Communications 2018; 9. DOI:10.1038/s41467-018-07329-0.

14 Preciado-Llanes L, Aulicino A, Canals R, et al. Evasion of MAIT cell recognition by the African Salmonella Typhimurium ST313 pathovar that causes invasive disease. Proceedings of the National Academy of Sciences of the United States of America 2020; 117: 20717–28.

15 Hanumunthadu B, Demissie T, Greenland M, et al. Safety and immunogenicity of the invasive non-typhoidal Salmonella (iNTS)-GMMA vaccine: a first-in-human, randomised, dose escalation trial. EBioMedicine 2025; 119: 105903.

16 Chen WH, Barnes RS, Sikorski MJ, et al. A combination typhoid and non-typhoidal Salmonella polysaccharide conjugate vaccine in healthy adults: a randomized, placebo-controlled phase 1 trial. Nat Med 2025; 31: 4256–64.

17 Martin LB, Tack B, Marchello CS, et al. Vaccine value profile for invasive non-typhoidal *Salmonella* disease. Vaccine 2024; 42: S101–24.

18 Emary K, Bentsi-Enchill AD, Giersing BK, et al. Landscape analysis of invasive non-typhoidal salmonella (iNTS) disease and iNTS vaccine use case and demand: Report of a WHO expert consultation. Vaccine 2025; 55: None.

19 Jin C, Gibani MM, Moore M, et al. Efficacy and immunogenicity of a Vi-tetanus toxoid conjugate vaccine in the prevention of typhoid fever using a controlled human infection model of Salmonella Typhi: a randomised controlled, phase 2b trial. The Lancet 2017; 390: 2472–80.

20 McCann N, Vicentine MP, Ebrahimi N, et al. Safety, Efficacy, and Immunogenicity of a Salmonella Paratyphi A Vaccine. New England Journal of Medicine 2025; 393: 1704–14.

21 Smith C, Smith E, Rydlova A, et al. Protocol for the challenge non-typhoidal Salmonella (CHANTS) study: a first-in-human, in-patient, double-blind, randomised, safety and dose-escalation controlled human infection model in the UK. BMJ Open 2024; 14: e076477.

22 Smith C, Bzami A, Zhu X, et al. Comparative phenotypic, genomic and transcriptomic characterisation of two Salmonella Typhimurium strains for a first-in-human challenge model. 2025; : 2025.04.24.650425 [PREPRINT].

23 UK Standards for Microbiology Investigations: Gastroenteritis. 2024; published online Oct 8. https://www.rcpath.org/static/a05a43b2-1e67-401d-8ca4f1c3af59038b/S-7i22-Gastroenteritis-October-2024.pdf.

24 UK Standards for Microbiology Investigations: Identification of Salmonella Species. 2021; published online March 12. https://www.rcpath.org/static/ba491b57-5f61-4e69-b31e59dbb8883a8c/a1164110-57c1-45d3-9cfb711c1efe70ce/uk-smi-id-24i4-identification-of-salmonella-species-march-2021-pdf.pdf.

25 UK Standards for Microbiology Investigations: Sepsis and systemic or disseminated infections. 2025; published online April 24. https://www.rcpath.org/static/3f51b8e5-1ebe-469d-a79f3a3323bfaec9/uk-smi-s-12i1-1-sepsis-and-systemic-or-disseminated-infection-april-2025-pdf.pdf.

26 Hormaeche E, Peluffo CA, Aleppo PL. Zur Ätiologie der Sommerdiarrhöe bei Kindern mit besonderer Berücksichtigung der Salmonella-Infektionen. Zeitschr f Hygiene 1937; 119: 453–8.

27 Mccullough NB, Eisele CW. Experimental human salmonellosis. I. Pathogenicity of strains of Salmonella meleagridis and Salmonella anatum obtained from spray-dried whole egg. J Infect Dis 1951; 88: 278–89.

28 Mccullough NB, Eisele CW. Experimental human salmonellosis. II. Immunity studies following experimental illness with Salmonella meleagridis and Salmonella anatum. J Immunol 1951; 66: 595–608.

29 Mccullough NB, Eisele CW. Experimental human salmonellosis. III. Pathogenicity of strains of Salmonella newport, Salmonella derby, and Salmonella bareilly obtained from spray-dried whole egg. J Infect Dis 1951; 89: 209–13.

30 Mccullough N, Eisele CW. Experimental human salmonellosis. IV. Pathogenicity of strains of Salmonella pullorum obtained from spray-dried whole egg. J Infect Dis 1951; 89: 259–65.

31 Angelakopoulos H, Hohmann EL. Pilot study of phoP/phoQ-deleted Salmonella enterica serovar typhimurium expressing Helicobacter priori urease in adult volunteers. Infection and Immunity 2000; 68: 2135–41.

32 Hindle Z, Chatfield SN, Phillimore J, et al. Characterization of Salmonella enterica derivatives harboring defined aroC and Salmonella pathogenicity island 2 type III secretion system (ssaV) mutations by immunization of healthy volunteers. Infection and Immunity 2002; 70: 3457–67.

33 Hammarlöf DL, Kröger C, Owen SV, et al. Role of a single noncoding nucleotide in the evolution of an epidemic African clade of Salmonella. Proceedings of the National Academy of Sciences of the United States of America 2018; 115: E2614–23.

34 Canals R, Chaudhuri RR, Steiner RE, et al. The fitness landscape of the African Salmonella Typhimurium ST313 strain D23580 reveals unique properties of the pBT1 plasmid. PLOS Pathogens 2019; 15: e1007948.

35 Siggins MK, O’Shaughnessy CM, Pravin J, et al. Differential timing of antibody-mediated phagocytosis and cell-free killing of invasive African Salmonella allows immune evasion. Eur J Immunol 2014; 44: 1093–8.

36 Nyirenda TS, Gilchrist JJ, Feasey NA, et al. Sequential Acquisition of T Cells and Antibodies to Nontyphoidal Salmonella in Malawian Children. The Journal of Infectious Diseases 2014; 210: 56–64.

37 Iankov ID, Petrov DP, Mladenov IV, Haralambieva IH, Mitov IG. Lipopolysaccharide-specific but not anti-flagellar immunoglobulin A monoclonal antibodies prevent Salmonella enterica serotype enteritidis invasion and replication within HEp-2 cell monolayers. Infect Immun 2002; 70: 1615–8.

38 Smith C, Smith E, Chiu C, et al. The Challenge Non-Typhoidal Salmonella (CHANTS) Consortium: Development of a non-typhoidal Salmonella controlled human infection model: Report from a consultation group workshop, 05 July 2022, London, UK [version 2; peer review: 2 approved]. Wellcome Open Research 2023; 8. DOI:10.12688/wellcomeopenres.19012.2.

39 Kreuzer M, Hardt W-D. How Food Affects Colonization Resistance Against Enteropathogenic Bacteria. Annual Review of Microbiology 2020; 74: 787–813.

40 Buchwald DS, Blaser MJ. A review of human salmonellosis: II. Duration of excretion following infection with nontyphi Salmonella. Reviews of infectious diseases 1984; 6: 345–56.

