## Supplementary Appendix for "Safety and pathovariant-independent susceptibility in a *Salmonella* Typhimurium controlled human infection model: a phase 1, randomised, double-blind, dose-escalation study"

### Supplementary Methods

#### Study Design

**Eligibility Criteria:**

Full inclusion and exclusion criteria have been reported previously and are included in the attached study protocol.^1^ Briefly, volunteers provided written informed consent and underwent structured screening to confirm absence of clinically significant systemic disease or key risk factors, including immunocompromise, recent malaria, or sickle cell disease. Screening included assessment of medical history, clinical examination and baseline investigations to confirm eligibility. All participants had abdominal ultrasonography performed to exclude gallstones or biliary tract pathology that might predispose to chronic carriage. Baseline serology for anti-*Salmonella* antibody titres was measured for exploratory purposes; results were blinded to the enrolling physicians and did not inform eligibility.

**Challenge Strains**: Participants were challenged with one of two strains of *Salmonella* Typhimurium. S. Typhimurium 4/74 (ST19; NCTC 14672) was isolated in 1974 from the bowel of a calf with salmonellosis and is a globally distributed ST19 lineage associated with human enterocolitis. S. Typhimurium D23580 (ST313; NCTC 14677) was isolated in 2004 from a blood culture obtained from a 24-month-old HIV-negative child in Blantyre, Malawi and is considered a representative ST313 Lineage 2 isolate associated with bloodstream infection in sub-Saharan Africa. The rationale for choice of strain selection and details of GMP manufacture, formulation and characterisation have been reported previously.^2^

**Randomisation and Blinding**: Randomisation to challenge strain allocation was performed at the pre-challenge visit 4-7 days before challenge and conducted by computer generated sealed envelope with permuted block sizes of two or four and a 1:1 allocation to 4/74 (ST19) or D23580 (ST313). The first 10 participants comprised a sentinel safety cohort, for whom the pre-specified block sequence was 2, 4 and 4. The study was conducted double-blind from randomisation until unblinding after the last participant completed the Day 28 post-challenge visit. A locked, randomisation allocation list was maintained by the study statistician (XL, YN) and unblinded laboratory team responsible for challenge agent preparation. Challenge agents were prepared by an unblinded investigator who performed no further clinical procedures.

**Study Site**: The study was conducted in both in-patient and out-patient settings within Imperial College Healthcare NHS Trust. On enrolment (Day 0), participants were admitted to single-occupancy rooms in a hospital-based quarantine facility at Charing Cross Hospital and remained in-patient until Day 7. Thereafter (Days 8–365), all visits were conducted on an out-patient basis at the NIHR Imperial Clinical Research Facility, Hammersmith Hospital.

**Challenge Agent Preparation and Administration**: Stocks of *S*. Typhimurium D23580 and 4/74 were maintained at -80°C storage in cryovials containing glycerol (final concentration 16% (w/v)). Vials of D23580 (4·9 × 10⁸ CFU/ml) and 4/74 (5·2 × 10⁸ CFU/ml) were thawed and diluted immediately prior to administration to achieve target concentrations of 1-5 × 10³ CFU/ml to 1-5 × 10⁵ CFU/ml, according to the dose-escalation stage. The true administered dose was confirmed by triplicate plating on Tryptone Soya Agar (TSA) plates (Oxoid/PO0163) with colony counts performed after 24 hours of incubation at 37°C.

Participants fasted for at least 120 minutes prior to challenge. Two minutes before challenge, a sodium bicarbonate solution (2·6g NaHCO₃ in 120 ml water) was administered to neutralise gastric acid. The challenge inoculum was then administered as a 30 ml suspension in 0.9% saline. Prepared challenge agents were stored on ice in a sealed biojar and administered within 60 minutes of preparation. Participants were monitored closely for 90 minutes following challenge, including assessment of vital signs and adherence to a further 2-hours of fasting. Participants who vomited within 90 minutes of challenge would receive prompt antibiotic therapy and be withdrawn from the study.

**Clinical Assessments and Safety Monitoring**: Safety was assessed with structured daily monitoring from challenge through at least Day 14. During in-patient Days 0-7, participants underwent daily review by a study physician, minimum four-times-daily vital signs, had 24-hour nursing care and continuous access to an on-call study doctor. Participants were issued a thermometer and a temperature diary to record self-measured oral temperatures at least twice daily and at the onset of subjective fever, in addition to nursing observations. During out-patient Days 8-14, participants attended daily physician reviews with vital signs recorded; a 24-hour emergency phone line provided ongoing access to an on-call study physician. Safety bloods were obtained on alternate days, at additional timepoints at discretion of the study physician, and daily for 5 days after meeting primary outcome diagnostic criteria.

Solicited symptoms were captured using self-completed diaries once daily from Day 0 to Day 14. For each pre-defined symptom, both presence/absence and severity were recorded using a 5-point ordinal scale (Grade 0 = not present to Grade 4 = hospitalisation/extension of hospital stay). Daily symptom severity scores were calculated by summing the total grading across individually recorded symptom components.

Bowel habits were recorded on participant-completed stool charts documenting frequency, timing and consistency of each bowel movement. Consistency was graded on the 7-point Bristol Stool Chart, and the presence of visible blood or mucus was additionally recorded. Daily diarrhoea severity scores were calculated by summing the total number of loose/liquid bowel movements in 24 hours.

Antibiotics were started on meeting the composite primary outcome (SD) or other treatment criteria, using agents active against both challenge strains. First-line therapy was oral azithromycin 500 mg once daily or ciprofloxacin 500 mg twice daily, with protocol-listed alternatives permitted at the study physician’s discretion for intolerance, adverse effects, or inadequate response. Symptomatic treatments (for pain, nausea, cramping) were permitted; antipyretics were prohibited until antibiotics commenced.

**Outcomes:** The primary objective of the study was to determine, for each strain, the oral challenge dose (CFU) producing a 60-75% attack rate of *Salmonella* diagnosis (SD). The primary estimand was the proportion of participants developing SD by Day 14, where SD was defined as a composite of either detectable bloodstream infection or sustained fever (oral temperature ≥38°C on ≥2 occasions ≥12 hours apart), with participants receiving antibiotics without meeting SD criteria (AT-nSD) excluded from the primary analysis. Secondary endpoints compared strains and dose levels for: mode of diagnosis, time to first temperature ≥38.0°C, fever clearance time, solicited symptom profiles (recorded via diaries outlined above), stool colonisation dynamics, gastroenteritis profiles, haematological and biochemical parameters, plasma cytokine profiles, gastrointestinal inflammatory biomarker profiles, humoral responses, and frequency of adverse and severe adverse events. Stool and blood cultures and safety blood monitoring were processed by an accredited pathology laboratory at North-West London Pathology, Charing Cross Hospital.

**Diagnostic Criteria**: Participants who met the composite primary outcome criteria were classified as having ‘*Salmonella* Diagnosis (SD)’; those who did not were classified as ‘no *Salmonella* Diagnosis (nSD)’. To enable timely treatment in clinically compatible illness that did not meet SD, we applied an alternative diagnostic criterion - ‘Antibiotic-Treated – no *Salmonella* Diagnosis (AT-nSD)’ – triggered by prespecified features, including severity of solicited symptoms, moderate/severe gastroenteritis, fever, persistent stool shedding, or at study physician discretion. Full AT-nSD diagnostic criteria are provided in the study protocol.

Sensitivity analyses were conducted to evaluate the impact of alternative assumptions regarding AT-nSD participants: (i) reclassifying AT-nSD as SD, (ii) reclassifying AT-nSD as non-SD, and (iii) treating AT-nSD as right-censored at the time of antibiotic initiation within a time-to-event framework.

**Criteria for Severe Salmonellosis**: Severe Salmonellosis was defined as participants meeting any of the following criteria: oral temperature >40°C; systolic blood pressure <85 mmHg; significant lethargy or confusion; gastrointestinal bleeding; gastrointestinal perforation; or any grade 4 laboratory abnormality.

**Dose Modification**: We aimed to perform a dose escalation/de-escalation study between the range of 1-5 x 10^1^ to 1-5 x 10^6^ CFU. We elected to start at a dose of 1-5 x 10^3^ CFU based on data from outbreak studies and previous invasive *Salmonella* human challenge studies^6,7^. Dose escalation for both strains was performed in parallel. Participants were randomised to receive one of the two challenge agents at their specific target dose until the target dose was established. In the event that the target dose was achieved for one challenge agent, the remaining participants would be challenged with the other agent until target dose was achieved. The Data Safety and Monitoring Committee (DSMC) were provided with interim unblinded reports following each cohort of 5 participants (per strain) to inform dose escalation or de-escalation decisions. The dose modification algorithm is provided in the study protocol and statistical analysis plan. If the primary attack rate was lower than the target of 60-75%, further dose-escalation could only occur if deemed safe to do so following review of safety data and agreement between the study team, trial statistician and DSMC.

#### Laboratory Procedures

**Blood Culture** Blood samples (10ml) were collected daily from Day 0-14 into aerobic blood culture bottles (BACTEC Plus Aerobic Medium, BD). Sample processing was performed in the Department of Microbiology, North-West London Pathology, Charing Cross Hospital in accordance with national guidelines.^3^ Blood cultures were incubated for five days at 35-37°C. Samples were reported as negative if no growth was detected after five days of incubation. In the event of a positive culture, a Gram stain was performed, and further subculture and identification was undertaken in accordance with national guidelines. The study team were contacted upon completion of the Gram stain, and treatment was immediately commenced if Gram-negative rods were detected, prior to full identification and sensitivity profiling.

**Stool Processing:** Stool samples were collected daily when a bowel movement occurred from Day 0-14, including prior to challenge and at longer term follow-up appointments (Days 28, 90, 180, 365). Samples were collected at the closest possible time point to each study visit. These were collected for processing immediately during the in-patient admission or were transported from home to clinic during out-patient follow-up using insulated and temperature-controlled transport bags.

Stool samples were processed for isolation and detection of *Salmonella* or other enteric pathogens in accordance with national guidelines and the local standard operating procedures (SOP) of North-West London Pathology ^4^. In brief, 1g of faeces was inoculated into selenite F broth and mixed by vortex prior to overnight incubation. 10µl of selenite broth was then inoculated directly onto xylose lysine deoxycholate (XLD) agar (Oxoid/PO0931A) for incubation at 37°C for 18-24 hours. A further 10µl of selenite broth was inoculated onto chromogenic agar after 18-24 hours incubation onto *Salmonella* chromogenic agar (Oxoid/PO0958A). Presence of *Salmonella* colonies was determined by observation of red colonies with black centres on XLD plates (hydrogen sulphide (H2S) production) or pink colonies on chromogenic agar. Confirmation of *Salmonella* spp. identification was subsequently confirmed by MALDI-ToF, API 20E and serological slide agglutination tests per national guidelines.^5^

All stool samples also underwent multiplex real-time PCR testing using the EntericBio system (Serosep). This platform detects multiple enteric pathogen targets including *Salmonella* spp., *Shigella* spp., *Campylobacter* spp., *Cryptosporidium* spp., *Giardia lamblia* and Verotoxigenic *E. coli* (VTEC).

**Blood Culture PCR:** 10 mL of heparinised peripheral venous blood was collected daily for 7 days post-challenge. Blood samples were added to 20 ml (3% (w/v) ox bile) tryptone soya broth media containing 1.5 ml of micrococcal nuclease (New England Biolabs, M0247S) and incubated for 5h at 37°C, 225 rpm. Bacteria were then concentrated by centrifugation first at 4000 g for 30 minutes followed by 6000 g for 20 min and the supernatant removed. DNA was extracted from the bacterial pellet using Qiagen Blood and Tissue DNA extraction kit (Qiagen, 69504) following the manufacturer’s instructions. Quantitative PCR (qPCR) was performed using primers targeting the *Salmonella*-specific *ttrA* gene^8^ (Forward: 5′- GGCGGAAACCTTTACCAGTC-3′; Reverse: 5′-GCTAACGCCGTTGAATTTGC-3′) with SYBR Green chemistry. Amplification and fluorescence detection were carried out under standard cycling conditions, and data were analysed using QuantStudio Design and Analysis Software according to the manufacturer’s instructions.

**Plasma Cytokines**: MSD Proinflammatory Panel 1 Kit (V-PLEX, MSD®) was used to measure levels of human IFN-y, IL-1β, IL-2, IL-4, IL-6, IL-8, IL-10, IL-12p70, IL-13, TNF-α according to the manufacturer’s instructions. Cytokines were measured in plasma samples at baseline, day 4/diagnosis and at day 7. Data were acquired with MESO QuickPlex SQ 120MM and analysed using the MSD ® DISCOVERY WORKBENCH 4.0 Analysis Software. Results were expressed as pg/mL for each cytokine.

**Saliva and Stool Sample Processing for Protein Analysis**: 1g of stool was homogenised in 8mL of extraction buffer (PBS, 0·05% Tween-20, cOmplete Protease Inhibitor Cocktail Tablets, 4% 0.5M EDTA), centrifuged at 500 x g for 10 minutes and 2200 x g for 30 minutes; stabilisation buffer (0·1% wt/vol BSA, 0·02% azide) was added and aliquots were stored at -80C.

Saliva and oral fluid were collected using Oracol Plus device (Malmed), as described previously ^9^, centrifuged at 900 x g for 5 minutes and 5,500 x g for 5 minutes to pellet cells and debris. Equal volume of stabilisation buffer (PBS, 10% FBS, 0·2% Tween-20, 0·5% Geneticin, 0·2% Fungizone) was added and aliquots were stored at -80°C.

**Lactoferrin and Calprotectin Detection:** Faecal Calprotectin and Lactoferrin were measured using Human Calprotectin ELISA Kit (orb564703-96T) and Human LTF/LF (Lactoferrin) ELISA Kit (E-EL-H5200-96T), respectively, according to manufacturer’s instructions. All participant samples were run in technical duplicate or triplicate. Calprotectin and Lactoferrin were measured at baseline, day 4 (nSD)/day of diagnosis (SD), and at day 7.

**Anti-*S*. Typhimurium IgG, IgA, IgM ELISA, and Serum Bactericidal Activity (SBA) assays:** Anti-*S*. Typhimurium-O-Ag IgG titres were measured as described previously.^10^ Briefly, anti-*Salmonella* O-Ag serum IgG was quantified using a standardised ELISA with STm O-Ag (ST2192) as coating antigen (5 μg/mL) and a 1:5000 dilution of goat anti-human IgG-alkaline phosphatase secondary antibody (Sigma–Aldrich, A3187). Results are expressed as ELISA units (EU)/ml, with one EU unit defined as the reciprocal of the dilution of the standard serum that yields an absorbance value of 1 (OD490-OD405).

Serum IgG, IgA, and IgM, as well as salivary and faecal IgA, were measured against STm OmpD and STm LPS. In-house ELISAs were performed as follows: 96-well plates were coated overnight at 4°C with STm OmpD (provided by Professor Constantino Lopez-Macias, Instituto Mexicano del Segura Social, Mexico) or STm LPS (Merck, L6511) at 5µg/mL, diluted in coating buffer (Abcam; ab210899). Plates were then washed, and serially diluted participant sera in blocking buffer (ThermoScientific; 37528). Antibodies were detected using peroxidase-labelled goat anti-human IgG (Merck; A6029; 1: 50,000), IgA (Merck; A0295; 1:10,000), or IgM (Merck; A18835; 1:20,000) followed by TMB substrate (Abcam; ab171522).

For anti-O-Ag IgG assays, a calibrated standard serum was used. This standard was generated by pooling high-responder sera collected 28 days after second vaccination in the SALVO study (blinded).^11^ The pooled SALVO serum was calibrated against a primary standard, to ensure alignment with the established definition of 1 EU as the reciprocal dilution yielding an OD = 1, following published methods.^10^ All other antigen-specific assays used an internal positive control reference serum prepared by pooling post-challenge sera (day 14/28) from selected participants. The positive control reference serum was calibrated separately against each coating antigen (STm LPS, STm OmpD), and the coating concentration for each antigen was set at the point of signal saturation. Control serum was serially diluted and 10 dilutions, range 1:10 – 1:51,200 based on the isotype measured, were included on every plate in technical duplicate or triplicate. Participant sera from Days 0, 14, 28, 90, and 180 were run in duplicate or triplicate at different dilutions (range 1:10 – 1:12,800). An EU/mL measurement was accepted as valid when the coefficient of variation across dilutions with OD values in the linear range of the standard curve was <30%. In addition, the assay run was accepted only when the OD values of the standard curve and internal controls fell within predefined acceptance limits. Titres were calculated by interpolating absorbance values (OD450) onto a five-parameter logistic curve generated from the relevant standard or reference serum dilution series. Endpoint titres are reported as EU/ml. Seroconversion was defined as a ≥4-fold increase in the antibody titre from baseline to day 28.

Luminescence-Based High-Throughput Serum Bactericidal Assay (SBA) was performed as described previously.^12,13^ Briefly, individual serum samples were heat inactivated (56°C for 30 minutes) and tested against the respective *S*. Typhimurium strain of participant challenge. The assay results were expressed as the IC_50_, the reciprocal serum dilution that resulted in a 50% reduction of luminescence and thus corresponding to 50% growth inhibition of the bacteria present in the assay.

#### Statistical Analysis

All analyses were conducted in R (v4.5.0). Participant data and laboratory reports were entered into a secure web-based case report form (OpenClinica Enterprise, v4.3.0).

**Sample Size**: No formal sample size calculation was performed; up to 80 participants (40 per strain) was deemed a feasible target within timeline and budget. Dose finding used a model-based continual reassessment method (CRM) to model the dose-attack rate for each strain, to guide the dose escalation decision, together with the safety profile. The oral dose yielding a predicted attack rate of systemic disease closest to 67·5% was selected if there was no safety concern determined by the DSMC. To our knowledge, this is the first CHIM to use CRM rather than rule-based escalation for CHIM dose-finding. CRM borrows information across dose levels and, in simulations (parameters in statistical analysis plan), outperformed rule-based designs for correct dose selection, particularly at small sample sizes, with advantages in accuracy. The study opened at 1-5 x 10^3^ CFU with permitted escalation up to 1-5 x 10^6^ CFU. An early-stopping rule applied once the model-recommended dose had accrued n=15 challenged participants. Complete CRM implementation details, including prior distributions, likelihood formulation, dose coding, and the integration of model recommendations with DSMC safety oversight, are provided in the statistical analysis plan and supplementary tables below.

**Primary Outcome**: The primary attack rate was defined as the proportion of participants meeting *Salmonella* diagnosis (SD) criteria. The denominator excluded withdrawals and participants treated before Day 14 without meeting SD criteria (AT-nSD). For each strain, model-estimated attack rates were derived using the continual reassessment method (CRM) with 95% credible intervals.

**Time-to-event Analyses:** Time-to-event outcomes were assessed using Kaplan–Meier methods including all participants. Event time was defined as the time of blood culture collection for culture-confirmed cases, or the time of first oral temperature ≥38.0°C that subsequently persisted ≥12 h for fever-defined cases. Participants without an event were censored at Day 14 (end of monitoring) or at withdrawal.

**Statistical Testing**: Between-group comparisons of categorical outcomes, including attack rates, used Fisher’s exact test. Time-to-event curves were compared using the log-rank test. Continuous variables were compared using the Mann–Whitney U test (for independent groups) or the Wilcoxon signed-rank test (for paired data). For multiple comparisons, p-values were adjusted using the Benjamini–Hochberg false discovery rate (BH-FDR) procedure across markers within each comparison family. All tests were two-sided, and statistical significance was defined as p < 0.05. The study was not powered for formal hypothesis testing, and statistical comparisons were conducted for exploratory hypothesis-generating purposes.

**Volcano Plots:** Volcano plots were used to visualise differential inflammatory and humoral responses between study groups. For each marker, the fold-change (FC) from baseline to the specified timepoint was calculated per participant, and distributions of participant log_2_FC values were compared between groups using a two-sided Wilcoxon rank-sum test. Group-level comparisons were expressed as the difference in median log_2_FC between participants who met SD criteria and those who did not (nSD), and separately between participants challenged with *Salmonella* Typhimurium D23580 (ST313) and 4/74 (ST19). Significance thresholds were set at p < 0·05 adjusted using the BH-FDR. Marker values below the lower limit of quantification (LLOQ) were imputed as LLOQ/2.

**Correlation Analyses:** Correlations between continuous variables were evaluated using Pearson’s correlation coefficient (r). Corresponding r and p-values are reported. Where indicated, statistical significance is denoted as p < 0·05*, p < 0·01**, and p < 0·001***.

**Logistic Regression Analyses**: Associations between baseline immunological markers and clinical outcomes were assessed using logistic regression models (one model per biomarker). Outcomes (*Salmonella* diagnosis SD; and colonisation) were coded binary (1/0). Biomarkers were log₁₀-transformed prior to analysis (values ≤0 set to missing). Exponentiated coefficients are presented as odds ratios (ORs) per tenfold increase in biomarker concentration. All models adjusted for prespecified covariates: age (continuous), sex (binary), challenge dose (ordinal: 10³, 10⁴, 10⁵ CFU), and strain (ST313 D23580 vs ST19 4/74). For colonisation analyses, participants who were non-colonised but received early antibiotics (<Day 3) were excluded a priori to minimise misclassification. Models were fitted by maximum likelihood using a logit link, and ORs with Wald 95% confidence intervals and two-sided p-values are reported. Analyses were implemented in R (v4.5.0) using the glm function (stats package), with broom for model tidying and ggplot2/forcats for visualisation.

#### Approvals

Written informed consent was obtained from all volunteers prior to enrolment. Ethical approval was obtained from the NHS Health Research Authority (London-Fulham Research Ethics Committee 21/PR/0051; IRAS Project ID 301659). The study was registered with clinicaltrials.gov (11 May 2023; https://clinicaltrials.gov/study/NCT05870150) and was performed according to the provisions of the Declaration of Helsinki and Good Clinical Practice guidelines. An independent data safety monitoring committee oversaw the conduct of the trial.

#### Data Availability

Data sets generated and/or analysed in this study are attached. Additional data are available from the corresponding author. No participant identifiable information will be disclosed. The full study protocol and statistical analysis plan are included as supplementary files. Genome sequences for the STm 4/74 and D23580 challenge strains are available as Genbank Accessions: CP002487.1 - CP002490.1 (4/74) and LS997973.1 – LS997977.1 (D23580).

#### Patient and Public Involvement

Patient and public involvement (PPI) was incorporated through pre-trial workshops with members of the public, during which study design, participant information materials, and practical aspects of the quarantine stay were reviewed. Feedback from these sessions directly informed revisions to recruitment materials, consent documentation, and the organisation of in-patient procedures.

#### Methods References

1	Christopher Smith, Emma Smith, Anna Rydlova, *et al.* Protocol for the challenge non-typhoidal <em>Salmonella</em> (CHANTS) study: a first-in-human, in-patient, double-blind, randomised, safety and dose-escalation controlled human infection model in the UK. *BMJ Open* 2024; **14**: e076477.

2	Smith C, Bzami A, Zhu X, *et al.* Comparative phenotypic, genomic and transcriptomic characterisation of two *Salmonella* Typhimurium strains for a first-in-human challenge model. 2025; published online April 28. DOI:10.1101/2025.04.24.650425.

3	UK Standards for Microbiology Investigations: Sepsis and systemic or disseminated infections. 2025; published online April 24. https://www.rcpath.org/static/3f51b8e5-1ebe-469d-a79f3a3323bfaec9/uk-smi-s-12i1-1-sepsis-and-systemic-or-disseminated-infection-april-2025-pdf.pdf.

4	UK Standards for Microbiology Investigations: Gastroenteritis. 2024; published online Oct 8. https://www.rcpath.org/static/a05a43b2-1e67-401d-8ca4f1c3af59038b/S-7i22-Gastroenteritis-October-2024.pdf.

5	UK Standards for Microbiology Investigations: Identification of Salmonella Species. 2021; published online March 12. https://www.rcpath.org/static/ba491b57-5f61-4e69-b31e59dbb8883a8c/a1164110-57c1-45d3-9cfb711c1efe70ce/uk-smi-id-24i4-identification-of-salmonella-species-march-2021-pdf.pdf.

6	Gibani MM, Jin C, Shrestha S, *et al.* Homologous and heterologous re-challenge with Salmonella Typhi and Salmonella Paratyphi A in a randomised controlled human infection model. *PLoS Negl Trop Dis* 2020; **14**: e0008783.

7	Dobinson HC, Gibani MM, Jones C, *et al.* Evaluation of the Clinical and Microbiological Response to Salmonella Paratyphi A Infection in the First Paratyphoid Human Challenge Model. *Clinical Infectious Diseases* 2017; **64**: 1066–73.

8	Chirambo AC, Nyirenda TS, Jambo N, *et al.* Performance of molecular methods for the detection of Salmonella in human stool specimens. *Wellcome Open Res* 2020; **5**: 237.

9	Elias SC, Muthumbi E, Mwanzu A, *et al.* Complementary measurement of nontyphoidal Salmonella-specific IgG and IgA antibodies in oral fluid and serum. *Heliyon* 2023; **9**: e12071.

10	Aruta MG, Lari E, De Simone D, *et al.* Characterization of Enzyme-Linked Immunosorbent Assay (ELISA) for Quantification of Antibodies against Salmonella Typhimurium and Salmonella Enteritidis O-Antigens in Human Sera. *BioTech* 2023; **12**: 54.

11	Hanumunthadu B, Demissie T, Greenland M, *et al.* Safety and immunogenicity of the invasive non-typhoidal Salmonella (iNTS)-GMMA vaccine: a first-in-human, randomised, dose escalation trial. *EBioMedicine* 2025; **119**: 105903.

12	Aruta MG, De Simone D, Dale H, *et al.* Development and Characterization of a Luminescence-Based High-Throughput Serum Bactericidal Assay (L-SBA) to Assess Bactericidal Activity of Human Sera against Nontyphoidal Salmonella. *MPs* 2022; **5**: 100.

13	De Simone D, Pinto M, Aruta MG, *et al.* GMMA-based vaccine candidates against invasive nontyphoidal salmonellosis elicit bactericidal antibodies against a panel of epidemiologically relevant Salmonellae. *Front Immunol* 2025; **16**: 1610067.

### Supplementary Tables

| **Dose Allocation** | **4/74 (ST19): Attack Rate (95% CI)** | **D23580 (ST313): Attack Rate (95% CI)** |
| --- | --- | --- |
| **AT-nSD Excluded (Primary Report)** | | |
| 10^3^ CFU | 9·6 (1·6-26·6) | 17·9 (4·5-38·5) |
| 10^4^ CFU | 26·7 (9·6-47·5) | 38·0 (17·5-58·5) |
| 10^5^ CFU | 47·4 (26·7-65·7) | 57·9 (37·3-73·8) |
| 10^6^ CFU | 61·7 (42·5-76·1) | 70·2 (52·8-82·1) |
| **Sensitivity Analysis: AT-nSD as Events** | | |
| 10^3^ CFU | 14·6 (3·5-33·1) | 22·0 (6·7-42·9) |
| 10^4^ CFU | 33·9 (15·2-53·7) | 42·7 (21·8-62·1) |
| 10^5^ CFU | 54·3 (34·6-70·4) | 61·8 (42·3-76·4) |
| 10^6^ CFU | 67·3 (50·2-79·6) | 73·2 (57·3-84·0) |
| **Sensitivity Analysis: AT-nSD as Non-Events** | | |
| 10^3^ CFU | 8·0 (1·3-23·0) | 14·6 (3·5-33·1) |
| 10^4^ CFU | 24·1 (8·6-43·7) | 33·9 (15·2-53·7) |
| 10^5^ CFU | 44·7 (25·1-62·6) | 54·3 (34·6-70·4) |
| 10^6^ CFU | 59·4 (40·8-73·8) | 67·3 (50·2-79·6) |

Table S. 1: CRM-estimated attack rates by dose, strain and analysis approach.

Posterior mean attack rates with 95% Bayesian credible intervals (CI) are shown for each dose level. Three analyses assess the sensitivity of estimates to handling of participants treated with antibiotics before Day 14 without meeting Salmonella Diagnosis criteria (AT-nSD; n=4). In the primary analysis, AT-nSD participants were excluded from the denominator, reflecting the pre-specified estimand. Two bounding sensitivity analyses counted AT-nSD participants as events (upper bound, assuming systemic disease) or as non-events (lower bound, assuming antibiotics pre-empted an outcome that would not have occurred). A third approach, censoring AT-nSD participants at the time of antibiotic initiation, is presented in the Kaplan-Meier analysis in Figure 2A-C. Estimates at 10⁶ CFU are model-based extrapolations; this dose was not administered. CRM, continual reassessment method; CFU, colony-forming units; AT-nSD, antibiotic-treated without Salmonella diagnosis; CI, credible interval

| **Challenge agent: 4/74 (ST19)** | | | | |
| --- | --- | --- | --- | --- |
|  | **Dose 1**  **(1-5x10^3^ CFU)** | **Dose 2**  **(1-5x10^4^ CFU)** | **Dose 3**  **(1-5x10^5^ CFU)** | **Dose 4**  **(1-5x10^6^ CFU)** |
| Prior attack rate skeleton | 60% | 75% | 85% | 90% |
| **Initial dose** |  |  |  |  |
| Actual attack rate | 0/5 (0 %) | - | - | - |
| Posterior probability  (95% credible intervals) | 7·02%  (0·00%, 50·29%) | 22·41%  (0·31%, 67·90%) | 42·96%  (3·82%, 80·36%) | 57·82%  (12·05%, 86·78%) |
| **Model recommendation** |  | **Y** |  |  |
| **DSMC decision** |  | **Y** |  |  |
| **1^st^ Dose escalation** |  |  |  |  |
| Actual attack rate | - | 0/5 (0 %) | - | - |
| Posterior probability  (95% credible intervals) | 0·92%  (0·00%, 22·34%) | 7·12%  (0·03%, 43·00%) | 22·48%  (0·94%, 62·08%) | 38·00%  (4·84%, 73·41%) |
| **Model recommendation** |  |  | **Y** |  |
| **DSMC decision** |  |  | **Y** |  |
| **2^nd^ Dose escalation** |  |  |  |  |
| Actual attack rate | - | - | 2/5 (40 %) | - |
| Posterior probability  (95% credible intervals) | 2·94 %  (0·07%, 18·04%) | 13·73 %  (1·68%, 38·12%) | 32·57 %  (9·93%, 57·99%) | 48·33 %  (22·38%, 70·24%) |
| **Model recommendation** |  |  |  | **Y** |
| **DSMC decision** |  |  | **Y** |  |
| **3^rd^ Dose, stay** |  |  |  |  |
| Actual attack rate | - | - | 3/5 (60 %) | - |
| Posterior probability  (95% credible intervals) | 5·89 %  (0·56%, 21·26%) | 20·29 %  (5·41%, 41·81%) | 40·62 %  (19·24%, 61·10%) | 55·76 %  (34·36%, 72·66%) |
| **Model recommendation** |  |  |  | **Y** |
| **DSMC decision** |  |  | **Y** |  |
| **4^th^ Dose, stay** |  |  |  |  |
| Actual attack rate | - | - | 3/5 (60 %) | - |
| Posterior probability  (95% credible intervals) | 7·98 %  (1·29%, 23·00%) | 24·08 %  (8·63%, 43·70%) | 44·74 %  (25·06%, 62·65%) | 59·36 %  (40·77%, 73·85%) |
| **Model recommendation** |  |  |  | **Y** |
| **DSMC decision** |  |  | **Y** |  |

Table S. 2: CRM dose-escalation decision-making for S. Typhimurium 4/74 (ST19).

The table shows the evolution of posterior attack rate estimates and dose recommendations following each cohort enrolment. The CRM used a prior attack rate skeleton of 60%, 75%, 85%, and 90% for doses 1-4 respectively, with a target attack rate of 67.5%. After each cohort, the model provided posterior probability estimates with 95% credible intervals for all dose levels and recommended the dose closest to the target. The Data Safety and Monitoring Committee (DSMC) reviewed model recommendations alongside safety data before confirming dose allocation for the next cohort. Dose escalation continued until 15 participants were enrolled at the model-recommended dose (10⁵ CFU). CRM, continual reassessment method; CFU, colony-forming units; DSMC, Data Safety and Monitoring Committee; Y=Yes/recommend the selected dose for the next stage.

| **Challenge agent: D23580 (ST313)** | | | | |
| --- | --- | --- | --- | --- |
|  | **Dose 1**  **(1-5x10^3^ CFU)** | **Dose 2**  **(1-5x10^4^ CFU)** | **Dose 3**  **(1-5x10^5^ CFU)** | **Dose 4**  **(1-5x10^6^ CFU)** |
| Prior attack rate skeleton | 60% | 75% | 85% | 90% |
| Initial dose |  |  |  |  |
| Actual attack rate | 0/5 (0 %) | - | - | - |
| Posterior probability  (95% credible intervals) | 7·02%  (0·00%, 50·29%) | 22·41%  (0·31%, 67·90%) | 42·96%  (3·82%, 80·36%) | 57·82%  (12·05%, 86·78%) |
| Model recommendation |  | Y |  |  |
| DSMC decision |  | Y |  |  |
| 1^st^ Dose escalation |  |  |  |  |
| Actual attack rate | - | 2/5 (40·0 %) | - | - |
| Posterior probability  (95% credible intervals) | 14·91%  (1·38%, 42·90%) | 34·24%  (8·98%, 62·09%) | 54·58%  (25·63%, 76·40%) | 67·54%  (41·37%, 83·99%) |
| Model recommendation |  |  | Y |  |
| DSMC decision |  |  | Y |  |
| 2^nd^ Dose escalation |  |  |  |  |
| Actual attack rate | - | - | 4/5 (80 %) | - |
| Posterior probability  (95% credible intervals) | 21·77 %  (4·40%, 47·52%) | 42·38 %  (17·23%, 65·77%) | 61·57 %  (37·02%, 78·92%) | 73·02 %  (52·51%, 85·77%) |
| Model recommendation |  |  |  | Y |
| DSMC decision |  |  | Y |  |
| 3^rd^ Dose, stay |  |  |  |  |
| Actual attack rate | - | - | 1/5 (20 %) | - |
| Posterior probability  (95% credible intervals) | 13·55 %  (2·57%, 33·58%) | 32·44 %  (12·72%, 54·08%) | 52·94 %  (31·20%, 70·66%) | 66·21 %  (46·99%, 79·84%) |
| Model recommendation |  |  |  | Y |
| DSMC decision |  |  | Y |  |
| 4^th^ Dose, stay |  |  |  |  |
| Actual attack rate | - | - | 3/5 (60 %) | - |
| Posterior probability  (95% credible intervals) | 14·64 %  (3·54%, 33·12%) | 33·89 %  (15·25%, 53·67%) | 54·27 %  (34·56%, 70·36%) | 67·28 %  (50·22%, 79·62%) |
| Model recommendation |  |  |  | Y |
| DSMC decision |  |  | Y |  |

Table S. 3: CRM dose-escalation decision-making for *S*. Typhimurium D23580 (ST313).

The table shows the evolution of posterior attack rate estimates and dose recommendations following each cohort enrolment, using identical methodology to Table S2. Both strains used the same prior skeleton and target attack rate (67.5%), with dose escalation proceeding independently based on strain-specific CRM modelling. CRM, continual reassessment method; CFU, colony-forming units; DSMC, Data Safety and Monitoring Committee. Y=Yes/recommend the selected dose for the next stage.

| **Exclusion Rationale** | **Total** |
| --- | --- |
| **Total withdrawals/exclusions** | **46** |
| Declined to participate after completion of screening | 20 |
| HLA-B*27 antigen positive | 6 |
| Psychiatric co-morbidity | 5 |
| Cardiovascular co-morbidity | 2 |
| Not received SARS-CoV-2 vaccination | 2 |
| Gallstones detected on abdominal ultrasonography | 2 |
| Enteric (non-*Salmonella*) pathogen detected on stool testing | 2 |
| Metallic orthopaedic prosthesis | 2 |
| Weight <50kg | 1 |
| Prolonged QTc Interval on ECG | 1 |
| Household contact immunosuppression | 1 |
| Sickle cell trait | 1 |
| Recent Gram-negative bloodstream infection | 1 |

Table S. 4: Reasons for participant screening exclusions

**Antibiotic-Treated – no *Salmonella* Diagnosis (AT-nSD) Criteria**

Any participant with severe gastroenteritis

Any participant with moderate gastroenteritis plus:

- Oral temperature ≥38.0°C on one occasion **and/or**
- ≥1 Grade 2 solicited systemic symptoms

Any participant with 3 or more Grade 2 solicited systemic symptoms on the same day:

- Headache
- Fatigue/malaise
- Nausea
- Vomiting
- Diarrhoea
- Abdominal pain
- Tenesmus
- Myalgia
- Arthralgia
- Cough

Any participant from whom *Salmonella* has been detected from at least two stool culture/PCR tests 24hrs apart who has not received antibiotics by day 14 post-challenge, unless spontaneous clearance is confirmed on three consecutive serial stool cultures by day 14.

Any participant in whom antibiotic use is felt to be clinically necessary (as decided by a medically qualified study doctor)

Table S. 5: Antibiotic-Treated – no Salmonella Diagnosis (AT-nSD) Criteria

| Strain | Dose | Study Day | AT-nSD Criteria |
| --- | --- | --- | --- |
| D23580 | 1-5 x 10⁴ CFU | Day 12 | - 3 or more symptoms of grade 2 or higher on same day - Single oral temperature ≥38·0°C |
| 4/74 | 1-5 x 10⁴ CFU | Day 4 | - 3 or more symptoms of grade 2 or higher on same day |
| 4/74 | 1-5 x 10⁴ CFU | Day 10 | - Moderate gastroenteritis plus fever ≥38·0°C on one occasion - 3 or more symptoms of grade 2 or higher on same day |
| D23580 | 1-5 x 10^5^ CFU | Day 3 | - Severe gastroenteritis - Single oral temperature ≥38·0°C |

Table S. 6: Participants fulfilling AT-nSD Criteria

| **Challenge Group** | | | | | | | |
| --- | --- | --- | --- | --- | --- | --- | --- |
|  | **All** | **ST19 (4/74)** | | | **ST313 (D23580)** | | |
|  |  | **10^3^ CFU** | **10^4^ CFU** | **10^5^ CFU** | **10^3^ CFU** | **10^4^ CFU** | **10^5^ CFU** |
| Time to First Fever ≥38°C, Hours, Median (Range) – SD Participants | 46·2  (24·1, 188·6) | - | - | 44·4  (24·4, 188·6) | - | 52·0  (49·0, 55·0) | 36·7  (24·1, 81·7) |
| Time to First Fever ≥38°C, Hours, Median (Range) – All Participants, any fever | 51·4  (24·1, 284·5) | - | - | 50·0  (24·4, 188·6) | - | 55·0  (49·0, 284·5) | 36·7  (24·1, 82·0) |
| Fever Clearance Time, Hours, Median (Range) – SD Participants | 32·7  (14·3, 236·8) | - | - | 36·0  (14·3, 236·8) | - | 29·9  (27·1, 32·8) | 25·4  (21·1, 61·0) |
| Fever Clearance Time, Hours, Median (Range) – All Participants, any fever | 24·7  (0·1, 236·8) | - | - | 28·1  (0·6, 236·8) | - | 27·1  (0·9, 32·8) | 23·9  (0·1, 61·0) |

Table S. 7: Fever kinetics stratified by challenge dose and strain.

Median time to first fever (≥38.0 °C) and fever clearance times are shown for participants with systemic salmonellosis (SD) and for all participants who experienced any fever. Values are presented overall and within each challenge group (4/74 and D23580 at 10³, 10⁴, and 10⁵ CFU). Dashes indicate dose-strain groups in which no participants developed fever.

| Strain | Dose | Diagnostic Category | Severe Salmonellosis Criteria |
| --- | --- | --- | --- |
| D23580 | 10⁴ CFU | AT-nSD | Grade 4 laboratory abnormality (ALT 374.0 U/L) |
| D23580 | 10^5^ CFU | Primary Outcome  (Fever) | Grade 4 laboratory abnormality (CRP 218.9 mg/L) |
| D23580 | 10^5^ CFU | AT-nSD | Gastrointestinal Bleeding |
| 4/74 | 10^5^ CFU | Primary Outcome  (Fever) | Gastrointestinal Bleeding |
| 4/74 | 10^5^ CFU | Primary Outcome  (Fever) | Gastrointestinal Bleeding |

Table S. 8: Participants meeting criteria for severe salmonellosis.

Protocol-defined severe salmonellosis was triggered by Grade 4 laboratory abnormalities (ALT >5× upper limit of normal [>250 U/L]; CRP >200 mg/L) or gastrointestinal bleeding with bloody diarrhoea. All events were self-limiting and resolved without complications or sequelae. Primary outcome refers to participants meeting Salmonella Diagnosis (SD) criteria (sustained fever ≥38.0°C on ≥2 occasions ≥12 hours apart and/or bacteraemia). AT-nSD refers to participants treated with antibiotics before Day 14 without meeting SD criteria. ALT, alanine aminotransferase; AT-nSD, antibiotic-treated without Salmonella diagnosis; CFU, colony-forming units; CRP, C-reactive protein.

| **Challenge Group** | | | | | | | |
| --- | --- | --- | --- | --- | --- | --- | --- |
|  | **All** | **ST19 (4/74)** | | | **ST313 (D23580)** | | |
|  |  | **10^3^ CFU** | **10^4^ CFU** | **10^5^ CFU** | **10^3^ CFU** | **10^4^ CFU** | **10^5^ CFU** |
| *Salmonella* Gastroenteritis Rate,  n (% total) | 15/50  (30·0%) | 0 | 1/5  (20·0%) | 6/15  (40·0%) | 0 | 1/5  (20·0%) | 7/15  (46·7%) |
| Any loose/liquid T6-7 stool, n (%) | 26/50  (52·0%) | 0 | 2/5  (40·0%) | 10/15  (66·6%) | 1/5  (20·0%) | 2/5  (40·0%) | 11/15  (73·3%) |
| Moderate Gastroenteritis, n (%)  (4-5 T6/7 stool in 24h) | 13/50  (26·0%) | 0 | 1/5  (20·0%) | 6/15  (40·0%) | 0 | 1/5  (20·0%) | 5/15  (33·3%) |
| Severe Gastroenteritis, n (%)  (≥6 T6/7 stool in 24h) | 10/50  (20·0%) | 0 | 0 | 5/15  (33·3%) | 0 | 1/5  (20·0%) | 4/15  (26·7%) |
| Moderate Gastroenteritis plus 1 fever  T ≥ 38·0°C, n (%) | 11/50  (22·0%) | 0 | 0 | 6/15  (40·0%) | 0 | 1/5  (20·0%) | 4/15  (26·7%) |
| Moderate Gastroenteritis plus 1 or more grade 2 gastrointestinal symptoms, n(%) | 12/50  (24·0%) | 0 | 1/5  (20·0%) | 5/15  (33·3%) | 0 | 1/5  (20·0%) | 5/15  (33·3%) |
| Maximum diarrhoea frequency/24h,  Mean | 4·8 | 0 | 3·0 | 5·7 | 2 | 3·5 | 4·7 |
| Maximum diarrhoea frequency/24h, Individual Participant Level | 15 | 0 | 5 | 15 | 2 | 6 | 11 |
| Time to onset of diarrhoea, hours, (Range) | 52·5  (23·5, 213·9) | - | 156·4  (99·0, 213·9) | 49·7  (23·5, 201·6) | 32·4  (32·4, 32·4) | 81·9  (68·8, 95·0) | 49·9  (34·0, 152·3) |

Table S. 9: Gastroenteritis outcomes stratified by challenge dose and strain

| **Challenge Group** | | | | | | | |
| --- | --- | --- | --- | --- | --- | --- | --- |
|  | **All** | **STm 4/74 (ST19)** | | | **STm D23580 (ST313)** | | |
|  |  | **10^3^ CFU** | **10^4^ CFU** | **10^5^ CFU** | **10^3^ CFU** | **10^4^ CFU** | **10^5^ CFU** |
| **Participants** | **50** | **5** | **5** | **15** | **5** | **5** | **15** |
| **Safety Outcomes** | | | | | | | |
| **Severe Salmonellosis Rate, n (% total)** | 5 (10%) | 0 | 0 | 2 (13·3%) | 0 | 1 (20·0%) | 2 (13·3%) |
| GI Bleeding | 3 (6·0%) | 0 | 0 | 2 (13·3%) | 0 | 0 | 1 (6·7%) |
| Oral Temperature ≥40C | 0 | 0 | 0 | 0 | 0 | 0 | 0 |
| SBP ≤ 85mmHg | 0 | 0 | 0 | 0 | 0 | 0 | 0 |
| GCS ≤ 12 | 0 | 0 | 0 | 0 | 0 | 0 | 0 |
| GI Perforation | 0 | 0 | 0 | 0 | 0 | 0 | 0 |
| Extra-intestinal seeding of STm | 0 | 0 | 0 | 0 | 0 | 0 | 0 |
| Grade 4 laboratory abnormality | 2 (4·0%) | 0 | 0 | 0 | 0 | 1 (20·0%) | 1 (6·7%) |
| **Adverse Events** | | | | | | | |
| **Adverse Events (total count)** | 94 | 7 | 5 | 26 | 8 | 16 | 32 |
| **Serious Adverse Events** | 0 | 0 | 0 | 0 | 0 | 0 | 0 |
| **SUSAR** | 0 | 0 | 0 | 0 | 0 | 0 | 0 |

Table S. 10: Summary Safety Outcomes by challenge strain and dose.

| **Strain** | **Challenge Dose** | **Comment** |
| --- | --- | --- |
| D23580 | 1-5 x 10^4^ CFU | AESI: Microbiological relapse of stool shedding  **Relationship to challenge:** Definite  Initial shedding D1-3, terminated during treatment with PO ciprofloxacin 500mg bd. Relapse positive stool culture D13, 14, 28. Repeat treatment with PO azithromycin 500mg od. Stool clearance achieved Day 62. |
| 4/74 | 1-5 x 10^4^ CFU | AESI: Microbiological relapse of stool shedding  **Relationship to challenge:** Definite  Initial shedding D1-10, terminated during treatment with PO ciprofloxacin 500mg bd. Relapse positive stool culture Day 28. Repeat treatment with PO azithromycin 500mg od. Stool clearance achieved Day 63. |
| D23580 | 1-5 x 10^5^ CFU | AESI: Microbiological relapse of stool shedding  **Relationship to challenge:** Definite  Initial shedding D1-7, terminated during treatment with PO azithromycin 500mg od. Relapse positive stool culture Day 28. Repeat treatment with PO ciprofloxacin 500mg bd. Stool clearance achieved Day 57. |
| 4/74 | 1-5 x 10^5^ CFU | AESI: Microbiological relapse of stool shedding  **Relationship to challenge:** Definite  Initial shedding D1-3, terminated during treatment with PO azithromycin 500mg od. Relapse positive stool culture Day 28. Repeat treatment with PO ciprofloxacin 500mg bd. Stool clearance achieved Day 57. |
| 4/74 | 1-5 x 10^5^ CFU | AESI: Microbiological relapse of stool shedding  **Relationship to challenge:** Definite  Initial shedding D1-4, terminated during initial treatment with PO azithromycin 500mg od (indication primary outcome = fever), subsequently changed to PO ciprofloxacin 500mg bd (indication = bloodstream infection). Relapse positive stool culture Day 90. Repeat treatment with PO co-trimoxazole 960mg bd. Persistent shedding on interval testing at Day 115 and Day 155. Spontaneous clearance day 365. |

Table S. 11: Incidence of microbiological shedding relapse with narrative.

### Supplementary Figures

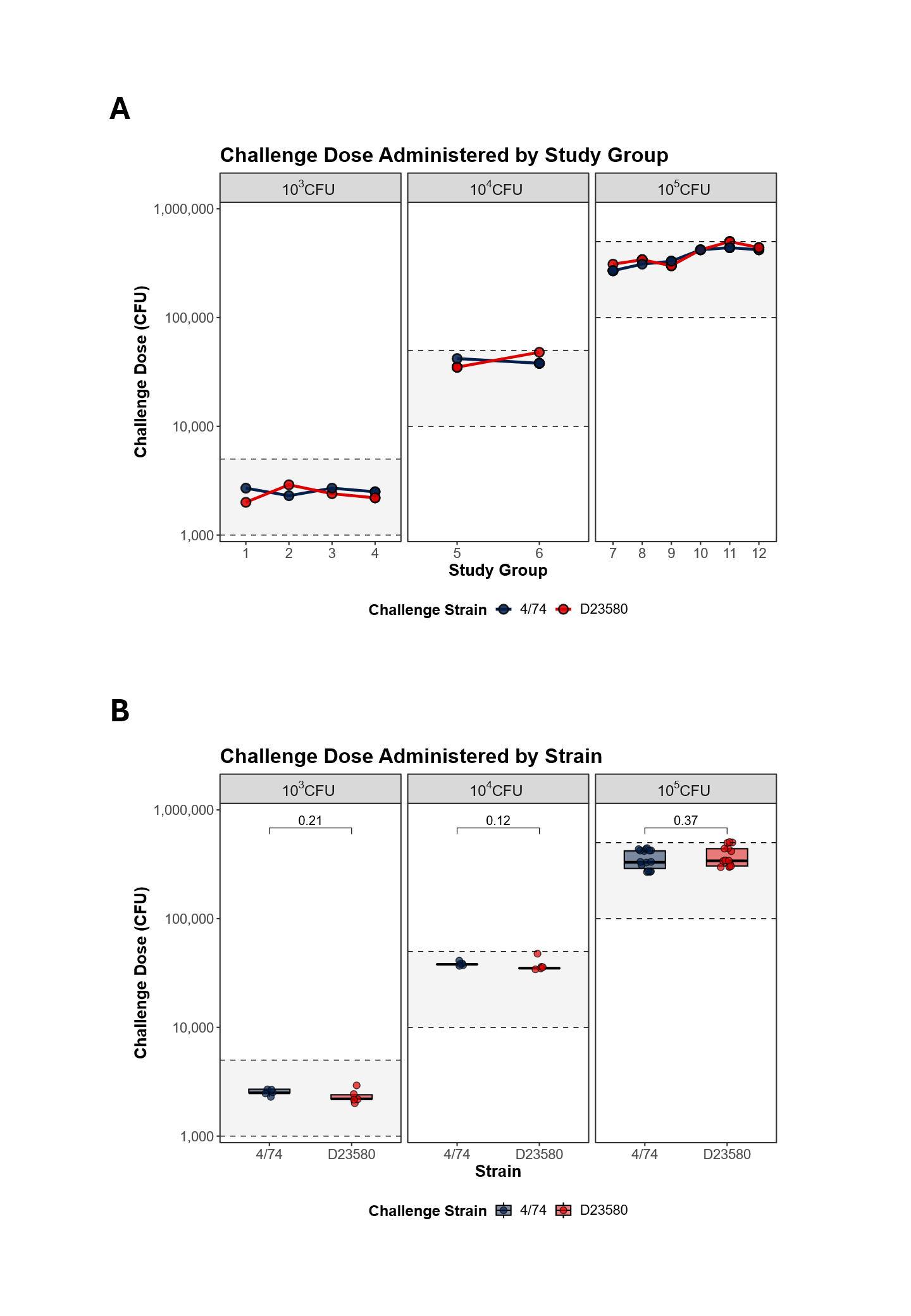

Figure S. 1: (A) Oral Challenge Dose Administration. Challenges were undertaken in 12 separate cohorts (group size 2-7). Administered challenges were all within target for allocated dose level, indicated by shaded grey boxes. (D) Challenge dose by strain. There were no significant differences between challenge doses administered between strains. Boxplots represent medians and interquartile ranges. P = Wilcoxon rank-sum test (two-sided).

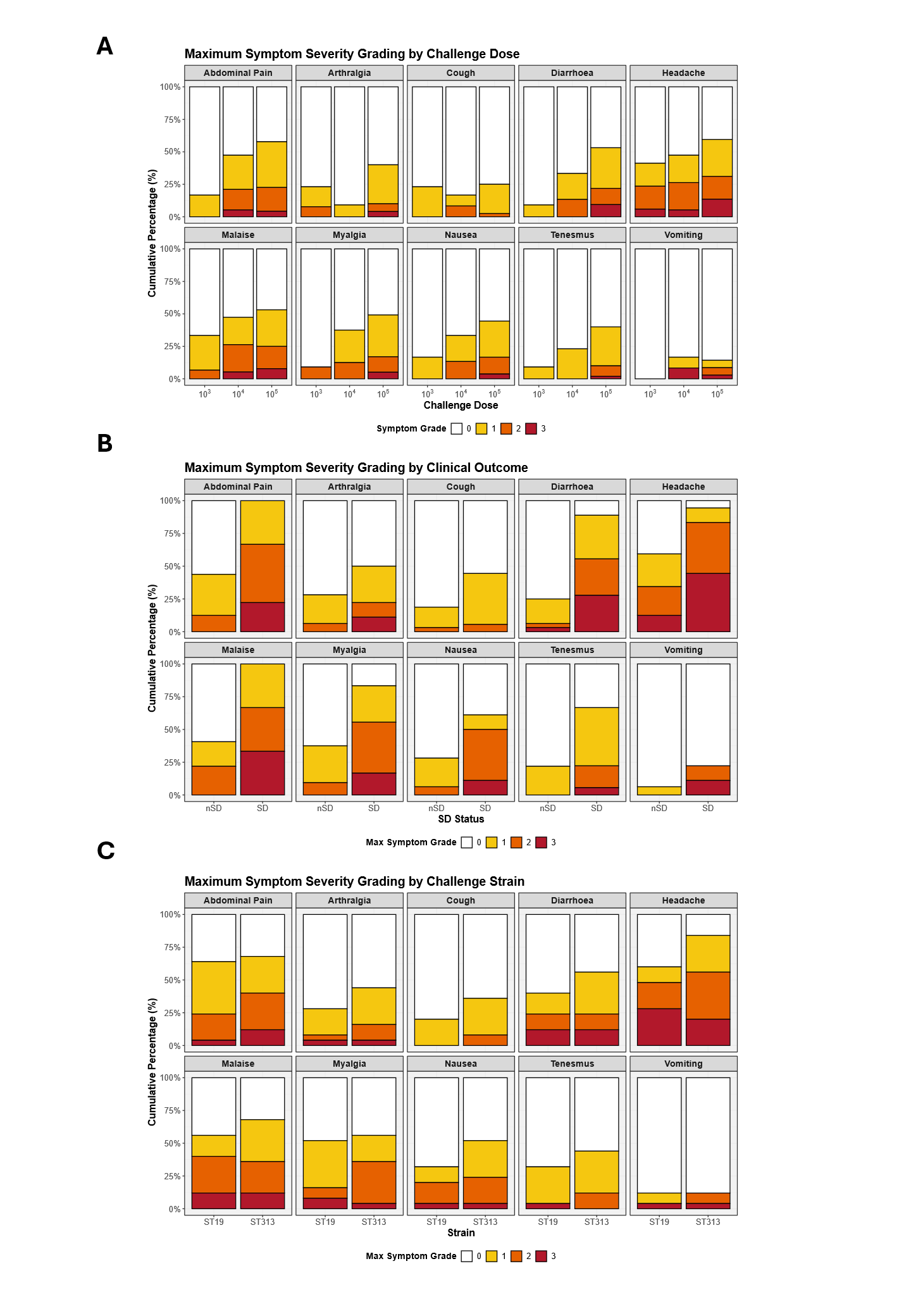

Figure S. 2: Maximum symptom severity scores across dose levels based on 10 solicited symptoms recorded daily (days 0–14) stratified by (A) challenge dose, (B) primary outcome; and (C) challenge strain. Stacked columns display percentage of participants reporting maximum symptom severity graded as mild (present but no interference with daily activity), moderate (some limitation of daily activity) or severe (unable to perform normal daily activity).

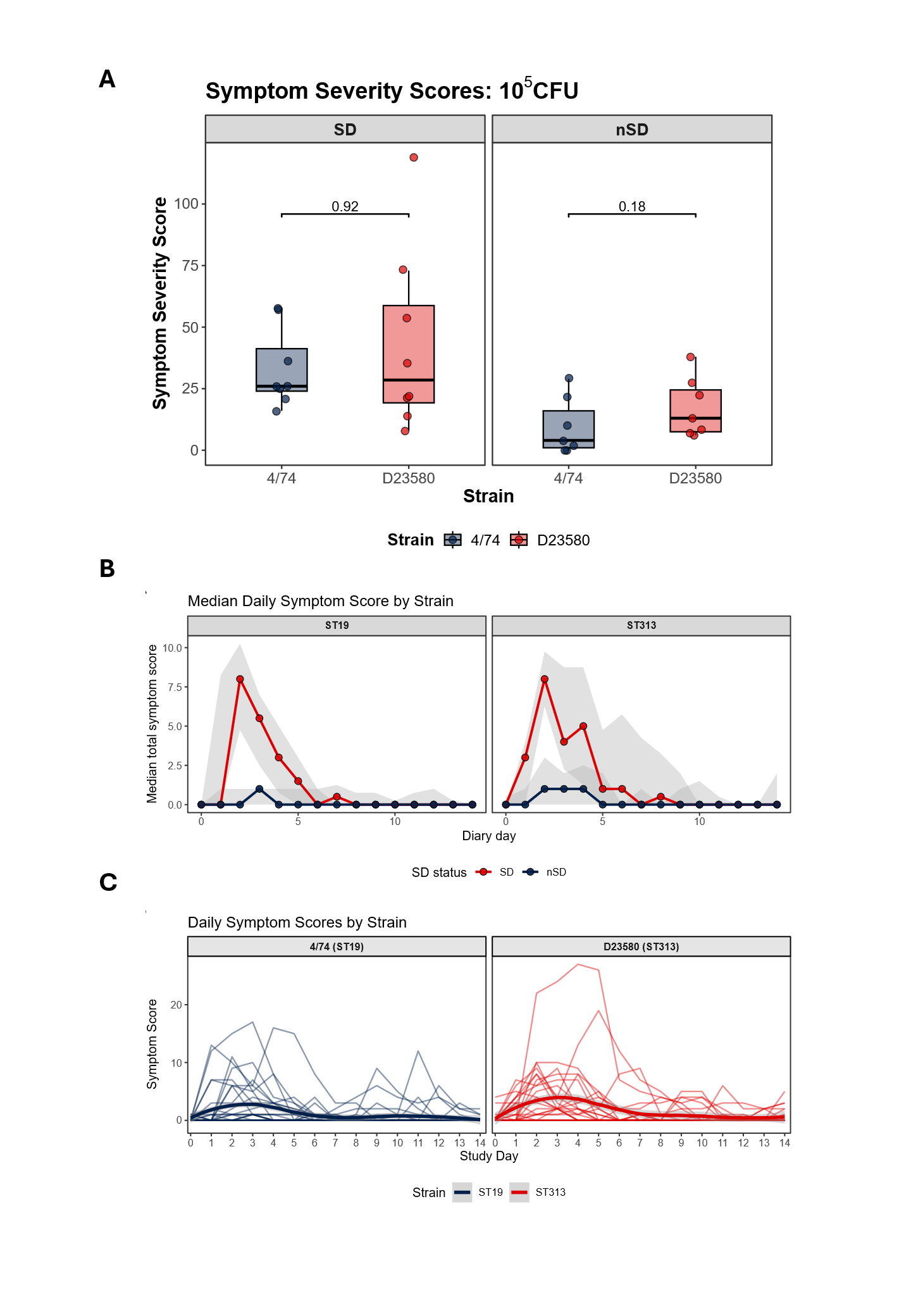

Figure S. 3: (A) Symptom severity scores stratified by strain and clinical outcome at 10^5^ CFU, nSD = No *Salmonella* Diagnosis, SD = *Salmonella* Diagnosis. Box plots represent median and interquartile range, P = Wilcoxon rank-sum test. (B) Median daily symptom severity by strain and outcome (IQR shaded). (C) Individual participant daily symptom score trajectories stratified by challenge strain.

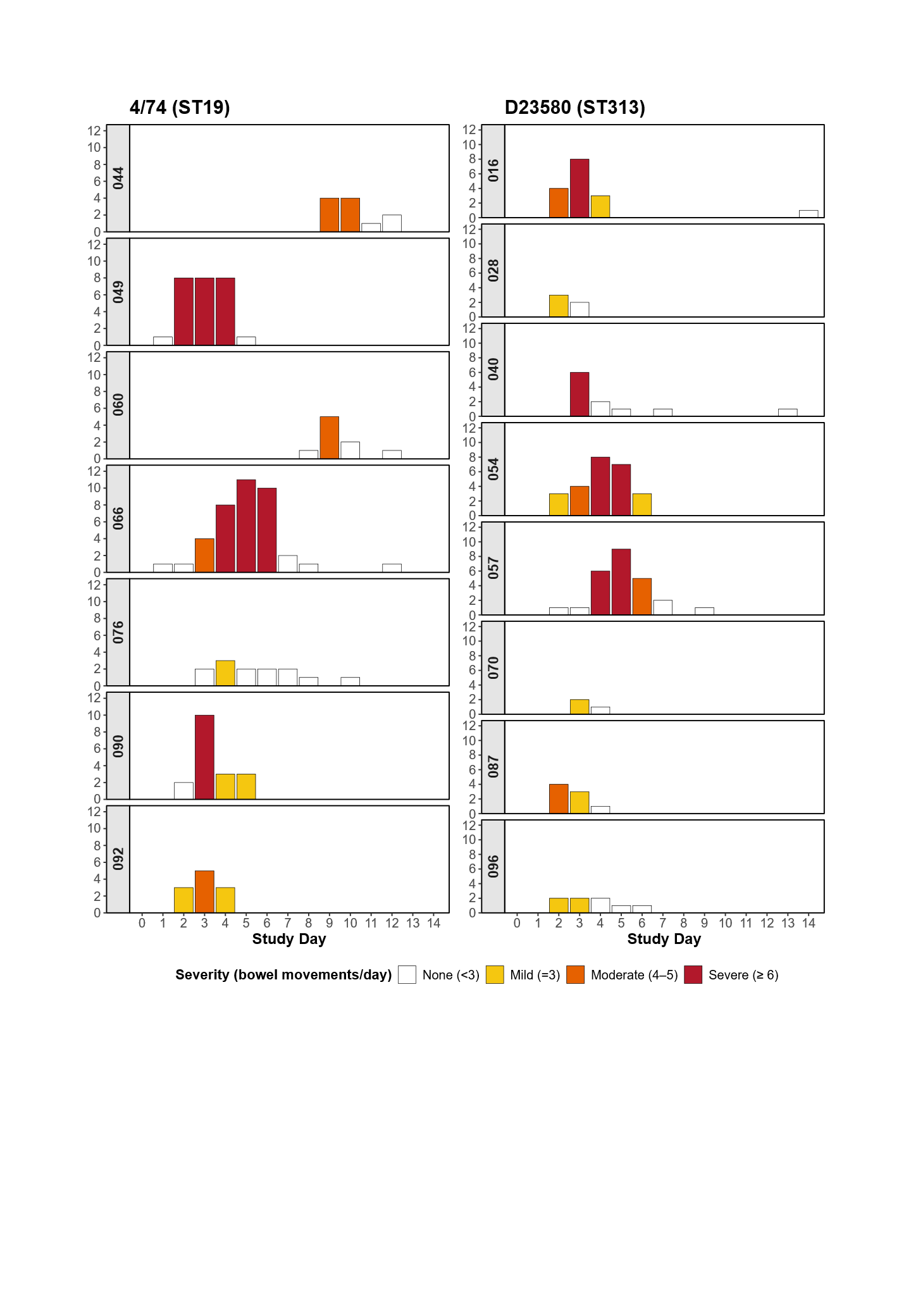

Figure S. 4: Daily participant level diarrhoea severity.

Individual diarrhoea trajectories from Day 0-14 for participants who developed gastroenteritis (7/25 receiving 4/74; 8/25 receiving D23580). Severity categories are defined as Mild = ≥3 loose/liquid stool in 24h; Moderate = 4-5 loose/liquid stool in 24h; Severe = ≥6 loose/liquid stool in 24h.

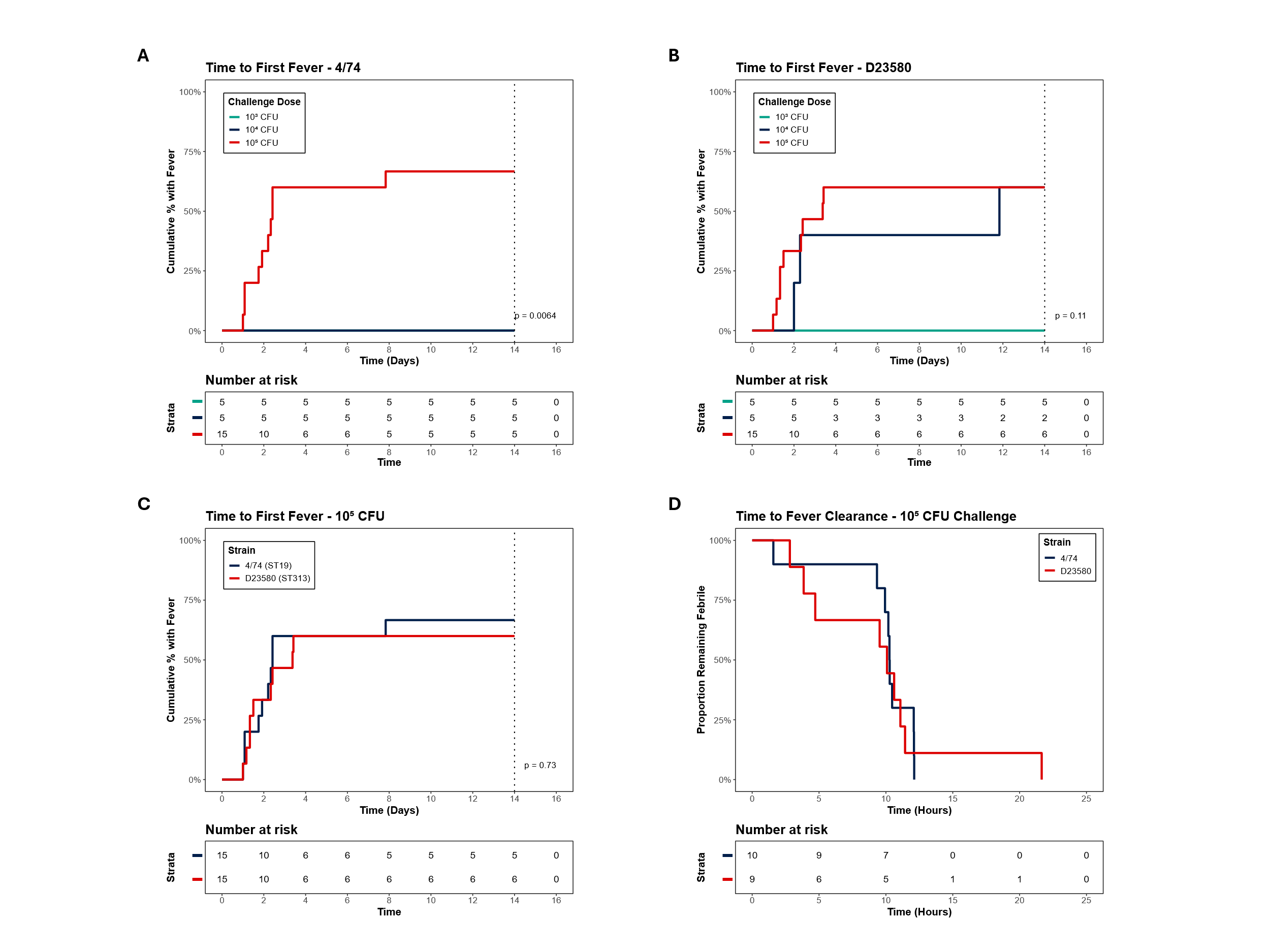

Figure S. 5: Time to fever onset and resolution by challenge dose and strain.

Kaplan-Meier plots illustrate the cumulative proportion of participants developing a first temperature recorded as fever (T ≥38.0°C). (A) STm 4/74 (ST19) by challenge dose, (B) STm D23580 (ST313) by challenge dose. (C) Strain comparison between groups challenged at 105 CFU, (D) Fever clearance times by strain for individuals who developed fever and were challenged at 105 CFU. P = log-rank test.

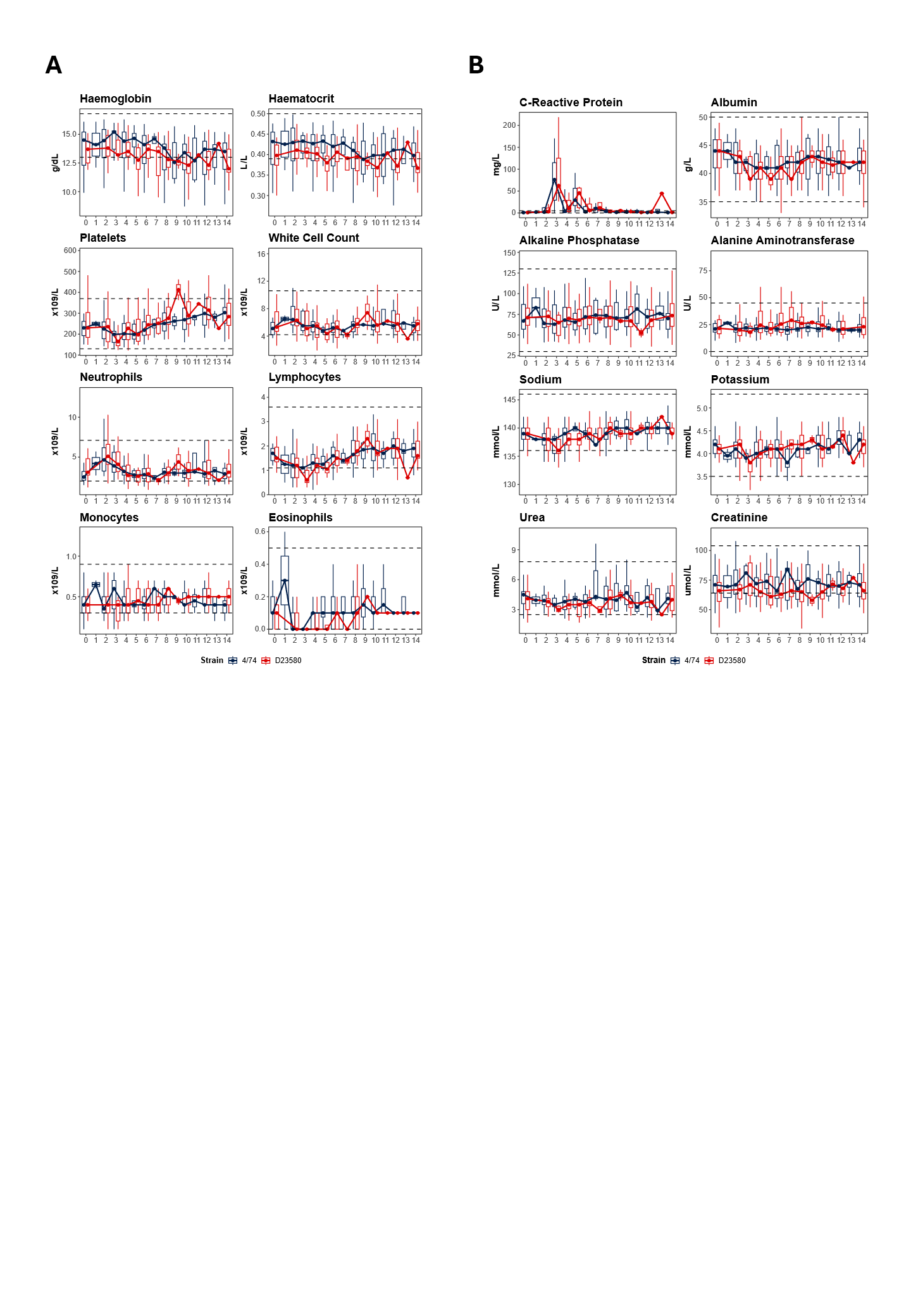

Figure S. 6: Routine safety laboratory parameter dynamics by challenge strain.

Collected throughout the challenge period from Day 0 – 14. (A) Haematology (Full Blood Count) and (B) Biochemistry (Renal Profile, Liver Function Tests, C-Reactive Protein). Boxplots represent median and interquartile range of all participants per day per strain. Dashed horizontal lines indicate upper and lower limits of normal in accordance with local reference ranges at North-West London Pathology.

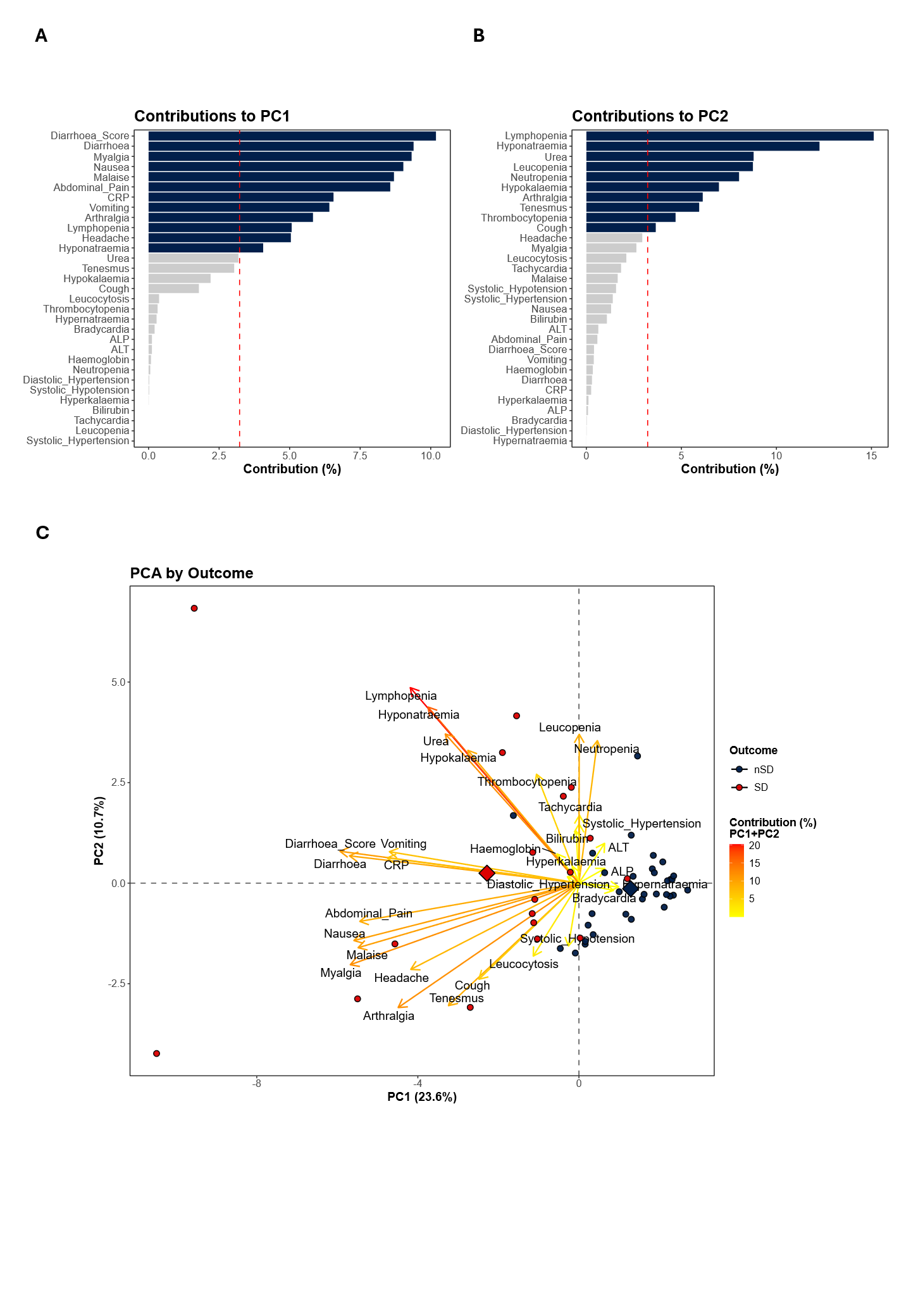

Figure S. 7: Principal component analysis (PCA) of all clinical features, solicited symptoms, and laboratory parameters.

Numeric variables were graded in accordance with Division of AIDS (DAIDS) Table for Grading the Severity of Adult and Pediatric Adverse Events. Principal component analysis was performed on z-scaled and centred numeric variables using the prcomp function in R. (A-B) Loadings plots showing variable contributions to PC1 and PC2, expressed as the percentage contribution of each variable to total component variance. Bars represent the relative influence of each variable on the component structure. The dashed vertical line indicates the expected average contribution (100% divided by the number of variables); variables exceeding this threshold contribute more strongly to that principal component. (C) PCA score plot stratified by diagnostic outcome (nSD = blue; SD = red). The percentage variance explained by each principal component is shown on the x- and y-axes. Biplot arrows represent variable loadings scaled according to their relative contribution.

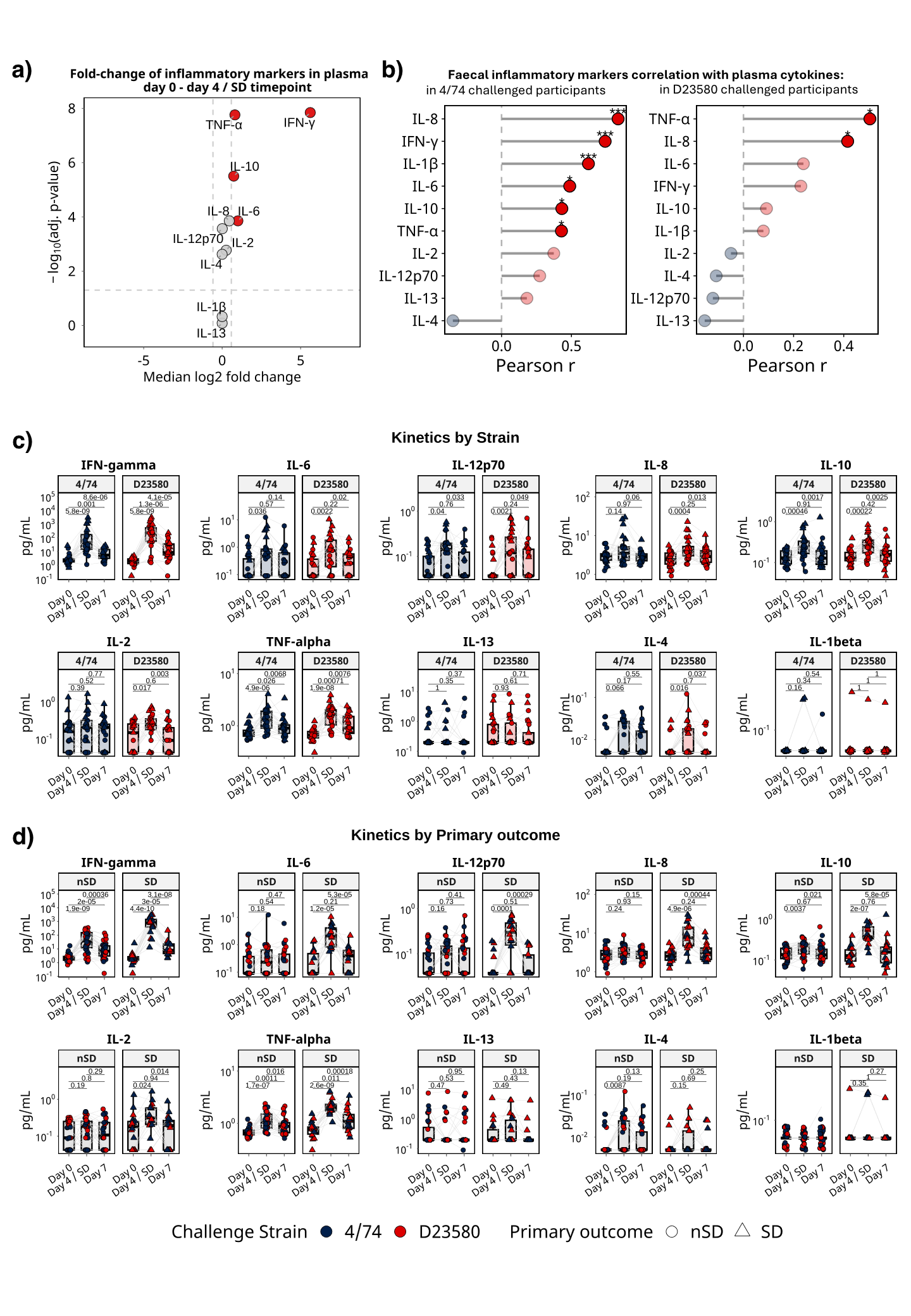

Figure S. 8: Plasma cytokine responses following *S*. Typhimurium challenge (A) Volcano plot showing the median log2 fold-change (FC) of plasma cytokines levels from baseline (Day 0) to Day 4/SD timepoint (measured in pg/mL). Horizontal dashed line shows the adjusted p = 0.05 cutoff (p-values were adjusted using the Benjamini–Hochberg procedure to control the false discovery rate (FDR)), the vertical dashed lines (-1, 1) show the cutoff for difference in FC between the specified cohorts (FC = 1.5). Red fill signifies the marker had FC > 1.5 and adjusted p-value < 0.05. (B) Strength and significance of Pearson correlation between the kinetics and magnitude of faecal markers (calprotectin and lactoferrin) and individual plasma cytokines, shown separately for participants challenged with 4/74 (left) and D23580 (right). Kinetics and magnitude were summarised by area-under-curve (AUC) calculated for the timepoints analysed (Day 0 – Day 4/ SD timepoint – Day 7). R value is plotted on x-axis, significance is denoted by as * p<0.05, ** p<0.01, *** p<0.001. (C, D) Individual cytokine response to NTS challenge from Day 0 to Day 7, stratified by challenge strain (C) with 4/74 (navy) or D23580 (red), and primary outcome (D): circle = (nSD); triangle = (SD). P values were calculated using two-sided Wilcoxon signed-rank tests (paired; unadjusted for multiple comparisons). nSD: participant did not meet primary outcome; SD: participant met primary outcome.

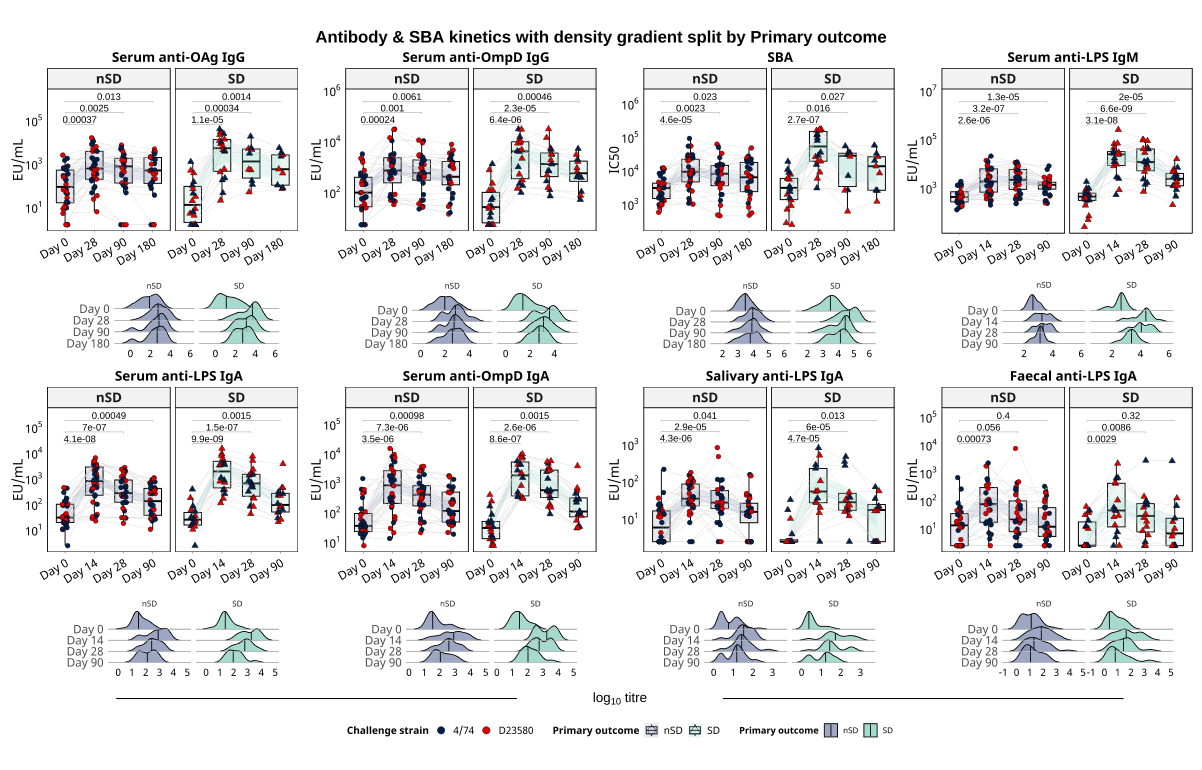

Figure S. 9: Antibody responses by challenge outcome.

Individual serum antigen-specific IgG, IgM and IgA titres were measured by ELISA and reported as ELISA units per millilitre (EU/mL). Serum IgG antibody and SBA IC50 response was measured at baseline (day 0), day 28, 90, and 180. Serum anti-LPS IgM titres were measured at baseline, day 14, 28 and 90. Serum, salivary and faecal anti-LPS and anti-OmpD IgA titres were measured at baseline, day 14, 28 and 90. Participants were challenged with one of two strains: 4/74 (navy) or D23580 (red). All plots are faceted by primary outcome (circle = participant did not meet primary outcome (nSD); triangle = Salmonella diagnosis (SD) / participant met primary outcome). Every individual measurement is plotted; thin grey lines connect repeated measurements from the same participant to show within-person trajectories. Boxplots summarise the group distribution at each timepoint (median line, box = IQR, whiskers = 1.5×IQR). Within strain, timepoints were compared using the Wilcoxon signed-rank test (paired, two-sided, p-value shown). Kernel density plots beneath each immune marker show the overall distributional shift (data pooled) within the primary outcome group. Curve height reflects the relative frequency of values, and the dashed vertical line marks the median at each timepoint.

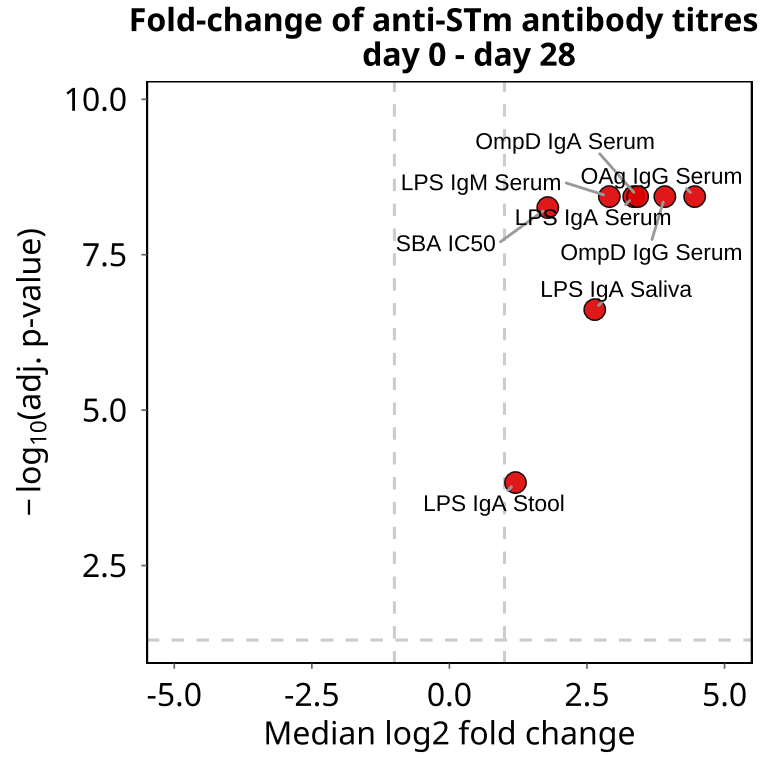

Figure S. 10: Volcano plot showing the median log2 fold-change (FC) of Salmonella-specific antibody levels from baseline (Day 0) to Day 28 (EU/mL).

Horizontal dashed line shows the adjusted p = 0.05 cutoff, the vertical dashed lines (-2, 2) show the cutoff for difference in FC between the specified cohorts (FC = 4). Red fill signifies FC > 4 and adjusted p-value < 0.05 (p-values were adjusted using the Benjamini–Hochberg procedure to control the false discovery rate (FDR)).

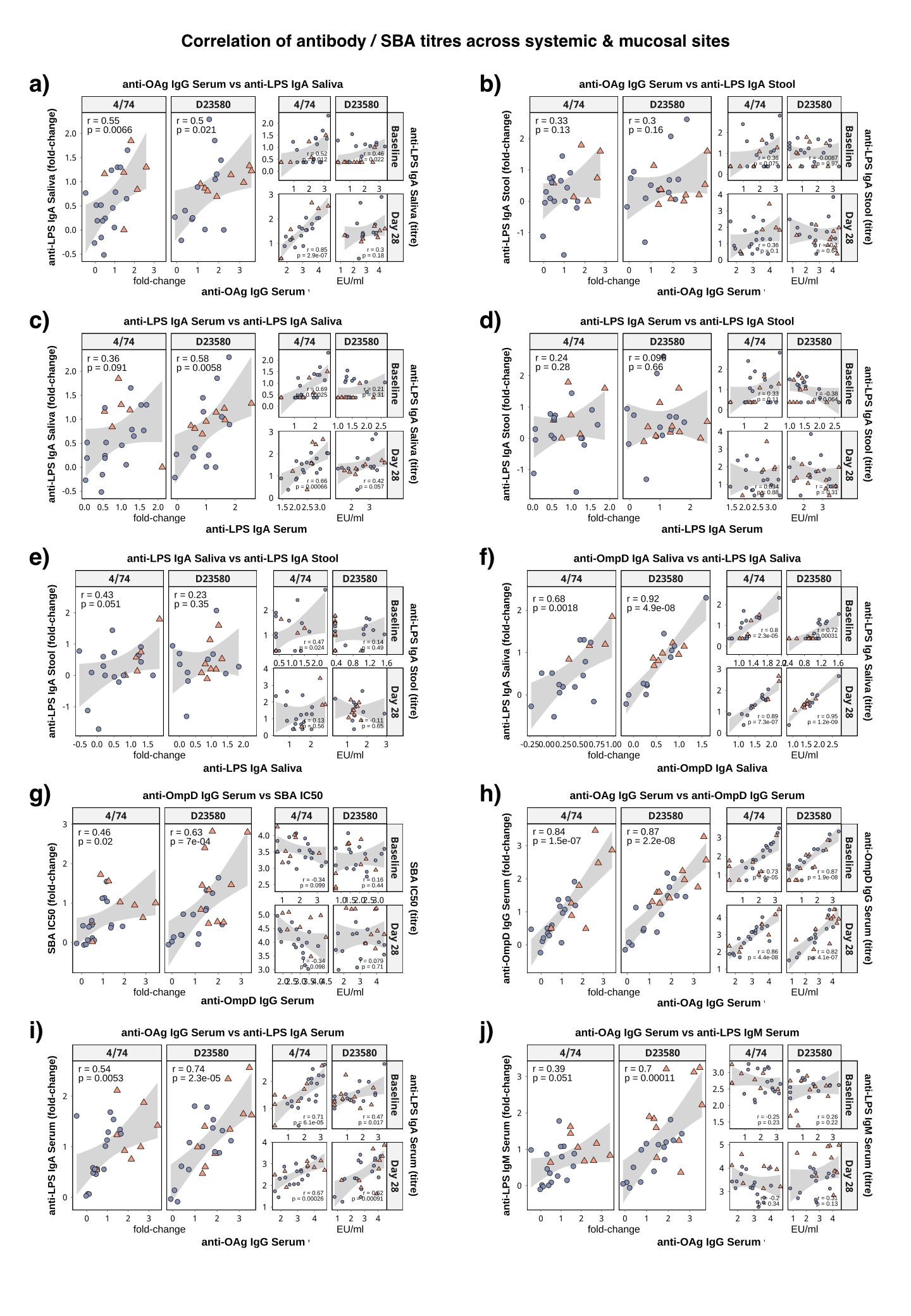

Figure S. 11: Correlation of antibody and SBA titres across systemic and mucosal sites. Each main panel shows the association between log₁₀ fold-change in antibody titre from baseline to Day 28 and log₁₀ fold-change in SBA IC50. Top right and bottom right inset panels show the correlation between absolute titres at baseline (top right) and day 28 (bottom right). Every point represents a participant (circles = nSD; triangles = SD). Pearson correlation coefficients (r) and p-values are shown; grey band shows 95% confidence interval. Statistics are calculated across all participants within each panel.

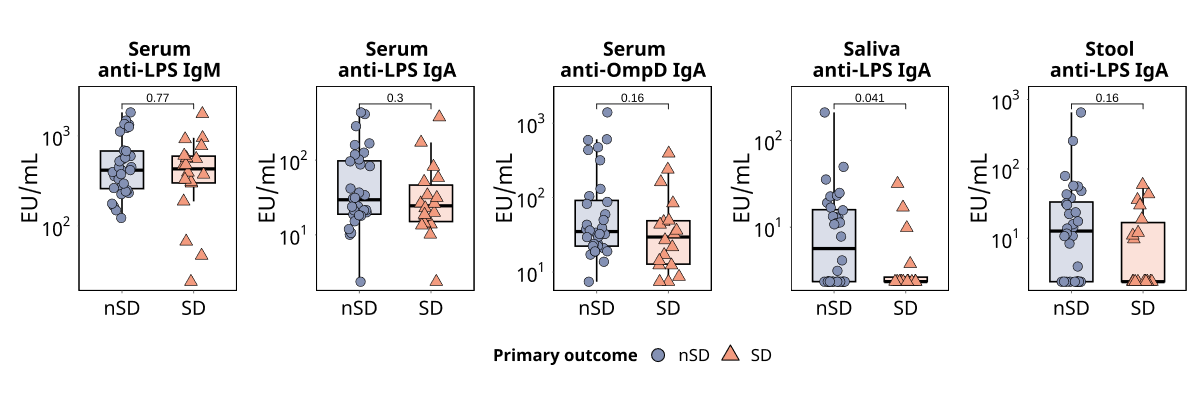

Figure S. 12: Baseline antibody titres by clinical outcome.

Participants who went on to develop systemic *Salmonella* diagnosis (SD, triangles) and those who did not (nSD, circles); two-sided Wilcoxon rank-sum test, p-value shown.
