## Supplementary material for "Safety and pathovariant-independent susceptibility in a *Salmonella* Typhimurium controlled human infection model: a phase 1, randomised, double-blind, dose-escalation study": Statistical Analysis Plan

Version 1.1 17 MAR 2025

**Study Title:** The Development of a Non-typhoidal *Salmonella* Human Challenge Model: A Safety and Dose Escalation Study
**Study Short Title:** Challenge Non-Typhoidal *Salmonella* (CHANTS) Study

**MAIN SPONSOR**: Imperial College London
**FUNDERS**: The Wellcome Trust (Award Reference 224029/Z/21/Z)
**STUDY COORDINATION CENTRE**: Imperial College London

**Sponsor study reference**: 21HH6989
**IRAS Project ID**: 301659
**REC reference**: 21/PR/0051

| | Name | Title | Signature | Date |
|---|---|---|---|---|
| Written by: | Yujie Ni | Statistician | | |
| | Dr Christopher Smith | Clinical Research Fellow | | |
| Reviewed by: | Dr Xinxue Liu | Lead Statistician | | |
| Approved by: | Dr Malick Gibani | Chief Investigator | | |

### 2 Introduction

#### 2.1 Study Summary

The Challenge Non-typhoidal *Salmonella* (CHANTS) study is a first-in-human phase 1, double-blind, randomised, dose-escalation controlled human infection model, conducted in healthy UK volunteers aged 18 to 50 years. The overall aim is to establish a model of NTS infection to better understand *Salmonella* infection biology and support candidate vaccine development.

A maximum of 80 eligible and consenting participants will receive oral challenge with one of two strains of *Salmonella enterica* subspecies *enterica* serovar Typhimurium (*S*. Typhimurium/STm) – 4/74 (ST19) and D23580 (ST313). Participants will be closely monitored for 14 days from challenge, with the first 7 days in an in-patient hospital setting, and subsequent 7 days by daily out-patient appointments. Antibiotic administration is commenced upon meeting the predefined primary outcome (sustained fever or bloodstream infection) or by meeting other predefined clinical safety criteria as outlined in CHANTS study protocol V3.1 25/Nov/2024.

### 3 Study Objectives

#### 3.1 Primary objective

To determine the dose in CFU of STm 4/74 and D23580 strains required for **60-75%** of volunteers to develop systemic Salmonellosis following oral challenge, defined as: *S*. Typhimurium bloodstream infection **AND/OR** sustained fever ≥38°C on ≥2 occasions ≥12 hours apart.

#### 3.2 Secondary objectives

- To describe the *Salmonella* colonisation rate following oral challenge with STm 4/74 and D23580 strains at different doses
- To describe the *Salmonella* gastroenteritis rate following oral challenge with STm 4/74 and D23580 strains at different doses.

- To determine the persistent fever rate following oral challenge with STm 4/74 and D23580 strains at different doses.
- To describe the *Salmonella* bacteraemia rate following oral challenge with STm 4/74 and D23580 strains at different doses.
- To describe the rate of systemic *Salmonellosis* according to an alternative composite diagnostic criterion following oral challenge with STm 4/74 and D23580 strains at different doses.
- To describe the safety of oral challenge with STm 4/74 and D23580 strains at different doses.
- To compare clinical features following oral challenge with STm 4/74 and D23580 strains at different doses.
- To compare microbiological features of gastrointestinal infection following oral challenge with STm 4/74 and D23580 strains at different doses.
- To compare microbiological features of bloodstream infection following oral challenge with STm 4/74 and D23580 strains at different doses.
- To compare biochemical and haematological laboratory parameters following oral challenge with STm 4/74 and D23580 strains at different doses
- To describe and compare the serum antibody response following oral challenge with STm 4/74 and D23580 strains at different doses.
- To describe and compare the mucosal antibody response following oral challenge with STm 4/74 and D23580 strains at different doses.
- To describe and compare cell-mediated immune response following oral challenge with STm 4/74 and D23580 strains at different doses.
- To describe participant experience following oral challenge with STm 4/74 and D23580 strains at different doses.

### 4 Study Design

#### 4.1 Study process

Eligible and consenting participants are randomised to receive oral challenge with one of two strains of STm (4/74 or D23580) at a starting dose of 1-5 x $10^3$ CFU (for both strains) administered following sodium bicarbonate pre-treatment. Dose escalation for both strains will happen in parallel. Participants will be randomised to receive one of the two challenge agents at their specific target dose until the target dose is establish for one strain. From that point, the remaining participants will be challenged with the other strain. Participants will be followed up for safety for a period of 12 months. A flowchart of the study design for dose escalation of each challenge strain utilising the continual reassessment method (CRM) model is presented in **Figure *1***.

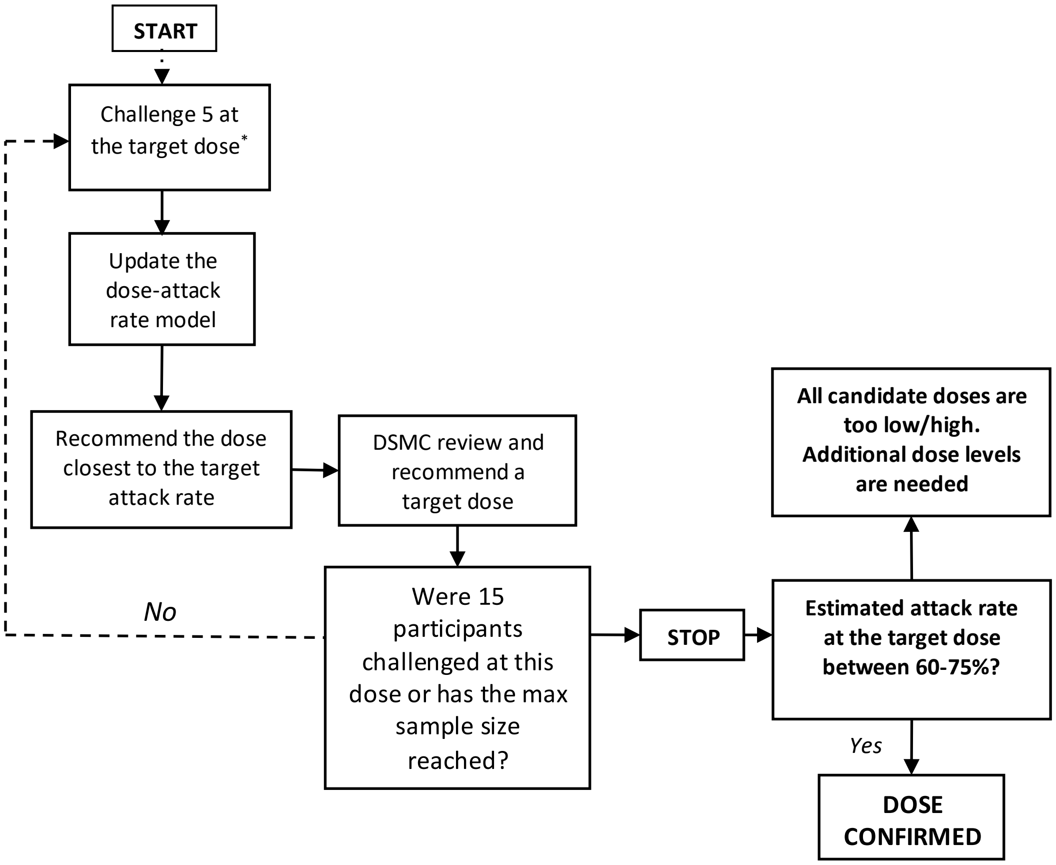

**Figure 1**: Decision making algorithm for dose escalation/de-escalation. Starting at 1-5 x $10^3$ CFU to reach the primary endpoint *The target dose for the first cohort will be 1-5 x $10^3$ CFU, and the first cohort of 5 participants will be divided into 3 groups (1, 2, and 2 participants) and challenge with an interval of at least 7 days.

##### 4.2 Study population

Study participants will be healthy volunteers aged 18 to 50. A full list of inclusion and exclusion criteria is outlined in CHANTS study protocol V3.1 25/Nov/2024.

##### 4.3 Randomisation and blinding

Randomisation of challenge agent will be carried out on or after Day -7. Randomisation will use random block sizes of two or four with an allocation ratio of 1:1 to receive 4/74 or D23580. The exception will be the three groups in the sentinel cohort, where the block sizes will be two, four and four, respectively. The study will be conducted double-blind from the time of randomisation until participant unblinding, which will occur once the last participant has completed the Day 28 post-challenge visit.

##### 4.4 Sample size

There is no formal sample size calculation for this dose-finding study. The maximum sample size for each challenge dose will be 40 participants. The sample finally recruited and used for dose identification are detailed within the continual reassessment method description.

##### 4.5 Continual Reassessment Method

Dose escalation or de-escalation will be guided by the continual reassessment method (CRM). The CRM uses a statistical model to estimate the relationship between dose and attack rate. After the actual attack rate from the challenged cohort is observed, the CRM will provide a posterior estimate of the attack rate for all dose levels using Bayesian inference. The dose recommended for the next cohort will be based on the allocation rule below. This process will continue until the total sample size is reached, or the early stopping rule (below) is triggered.

Below model and parameters will be used to run CRM:

| **CRM Model Parameters** | |
|---|---|
| Dose Levels | 1-5 x $10^3$ CFU, 1-5 x $10^4$ CFU, 1-5 x $10^5$ CFU, 1-5 x $10^6$ CFU |
| Dose coding | Ordinal labels 1-4 |
| Outcome | The binary likelihood for each participants's attack outcome |
| Target Attack Rate | 67.5% |
| Dose-Attack Rate Model | One-parameter power model<br>$p\ (d) = skeleton(d)^{exp(\alpha)}$ |
| Prior distribution of parameter α | Normal (mean = 0, variance = 1.34) |
| Dose-Attack Rate Skeleton | 60%, 75%, 85%, 90% |
| Inference | Bayesian |
| Decision Rule | Dose closest to target attack rate |
| Sample Size | 40 per each challenge agent (maximum) |
| Cohort Size | 5 |
| Safety Modifications | Starting at the lowest dose and only one dose escalation at a time |
| Stopping Rule | 15 participants challenged at the recommended dose |

The same rules and parameters are applied to both strains. The CRM will estimate the attack rate at each dose level, including 95% Bayesian posterior credible intervals, and dose recommendations will be provided independently for the two challenge strains. The Data and Safety Monitoring Committee (DSMC) will review model recommendations and retain the authority to override them based on safety or other considerations. The CRM will be implemented using the R package dfcrm (version 0.2-2.1).

#### 5 Analysis Populations

| Population | Description |
|---|---|
| **All participants** | All participants screened for the trial, to be used for reporting CONSORT diagram. |
| **Safety analysis population** | • Have consented to the study and met all the inclusion and exclusion criteria;<br>• Randomised participants who underwent study challenge will be included.<br>• Participants who withdraw consent will be included up to the date of their study withdrawal. |
| **Analysis population 1**<br>(Primary analysis) | • Have consented to the study and met all the inclusion and exclusion criteria;<br>• Have been successfully challenged with 4/74 or D23580;<br>• Participants withdrew or were treated prior to Day 14 without being diagnosed will be excluded. |
| **Analysis population 2**<br>(Secondary analysis) | • Have consented to the study and met all the inclusion and exclusion criteria;<br>• Have been successfully challenged with 4/74 or D23580;<br>• Have at least one evaluable post-challenge clinical specimen;<br>• Have at least one evaluable post-challenge safety data; |

#### 6 Variable of analysis

| Objective(s) | Endpoint(s) | Timepoints/visits |
|---|---|---|
| **Primary endpoint** | | |
| 1. To determine the dose in CFU of STm 4/74 and D23580 strains required for 60-75% of volunteers to develop systemic Salmonellosis following oral challenge. | 1. The proportion of participants who develop either:<br>1.1. Fever ≥38°C on ≥2 occasions ≥12 hours apart **AND/OR**<br>1.2. *Salmonella* Typhimurium bacteraemia | D0 – D28 |
| **Secondary endpoints** | | |
| 2.1 To describe the *Salmonella* colonisation rate following oral challenge with STm 4/74 and D23580 strains at different doses | 2.1 The proportion from whom *Salmonella* Typhimurium is isolated from stool on ≥2 occasions ≥48 hours from challenge at different doses of each strain. | D0 – D28 |
| 2.2 To describe the *Salmonella* gastroenteritis rate following oral challenge with STm 4/74 and D23580 strains at different doses. | 2.2 The proportion of participants at different doses at each strain developing<br>2.2.1 Severe diarrhoea **and/or**<br>2.2.2 Moderate diarrhoea plus<br>2.2.2.1 Fever ≥38°C on ≥1 occasion **and/or** | D0 – D28 |

| | | |
|---|---|---|
| | 2.2.2.2 ≥1 Grade 2 gastrointestinal symptoms (abdominal pain, nausea, vomiting, tenesmus) | |
| 2.3 To determine the persistent fever rate following oral challenge with STm 4/74 and D23580 strains at different doses. | 2.3 The proportion of participants who develop fever ≥38°C on ≥2 occasions ≥12 hours apart at different doses of each strain. | D0 – D28 |
| 2.4 To describe the *Salmonella* bacteraemia rate following oral challenge with STm 4/74 and D23580 strains at different doses. | 2.4 The proportion of participants developing *Salmonella* Typhimurium bacteraemia at any timepoint following challenge at different doses of each strain | D0 – D28 |
| 2.5 To describe the rate of systemic Salmonellosis according to **an alternative composite diagnostic criterion** following oral challenge with STm 4/74 and D23580 strains at different doses. | 2.5 The proportion of participants meeting the criteria for a composite diagnosis of *Salmonellosis* at different doses of each strain defined as any of 1) *Salmonella* Typhimurium is isolated from stool on ≥2 occasions ≥48 hours from challenge and/or 2) *Salmonella* gastroenteritis and/or 3) fever ≥38°C on ≥2 occasions ≥12 hours apart and/or 4) *Salmonella* Typhimurium bacteraemia | D0 – D28 |

| | | |
|---|---|---|
| 2.6 To describe the safety of oral challenge with STm 4/74 and D23580 strains at different doses. | 2.6 The proportion of participants at different doses of each strain reporting<br>• Adverse events,<br>• Adverse events of special interest,<br>• SAEs,<br>• SUSARs.<br>• Concomitant medication usage as outlined in the study protocol | Throughout study from challenge |
| 2.7 To compare **clinical** features following oral challenge with STm 4/74 and D23580 strains at different doses. | 2.7 A comparison of the clinical features of *Salmonella* infection after challenge with 4/74 or D23580 strains, with specific reference to<br>2.7.1 The proportion of participants in each group developing any diarrhoea.<br>2.7.2 The severity of diarrhoea in each group, as measured by<br>2.7.2.1 Stool volume in ml/24hrs<br>2.7.2.2 Frequency of bowel motions/24hrs<br>2.7.2.3 Total gastrointestinal symptom severity score calculated by summing numerical values assigned to the severity of all solicited gastrointestinal symptoms | D0 – D28 |

| | |
|---|---|
| | between Day 0 to Day 14 (0=not present; 1=mild; 2=moderate; 3=severe).<br>2.7.2.4 Total systemic symptom severity score calculated by summing numerical values assigned to the severity of all solicited gastrointestinal and systemic symptoms between Day 0 to Day 14 (0=not present; 1=mild; 2=moderate; 3=severe).<br>2.7.2.5 Area under the curve for gastrointestinal and systemic symptom severity score between Day 0 to Day 14<br>2.7.2.6 Time to onset of diarrhoea in each group, measured in hours since challenge to the first grade 6/7 stool.<br>2.7.3 The proportion of participants in each group developing any fever ≥38°C<br>2.7.4 Fever clearance time of febrile participants in each group measured by time in hours to first temperature <38°C lasting ≥12hrs |

| | |
|---|---|
| | 2.7.5 The proportion of participants in each group reporting the following solicited symptoms at any time:<br>2.7.5.1 headache,<br>2.7.5.2 malaise<br>2.7.5.3 anorexia<br>2.7.5.4 abdominal pain<br>2.7.5.5 nausea<br>2.7.5.6 vomiting<br>2.7.5.7 dysentery<br>2.7.5.8 myalgia<br>2.7.5.9 arthralgia<br>2.7.5.10 cough<br>2.7.5.11 rash<br>2.7.6 Duration of solicited symptoms measured in days from first symptom onset to complete resolution of symptoms<br>2.7.7 Severity of solicited symptoms measured by<br>2.7.7.1 The proportion of participants with maximum symptom severity score |

| | | |
|---|---|---|
| | graded as mild, moderate, or severe following challenge<br>2.7.7.2 The proportion of participants meeting the criteria for severe *Salmonellosis*<br>2.7.7.3 Symptom severity scores for individual solicited symptoms calculated by summing numerical values assigned to the severity of individual solicited symptoms between Day 0 to Day 14 (0=not present; 1=mild; 2=moderate; 3=severe) | |
| 2.8 To compare **microbiological** features of **gastrointestinal** infection following oral challenge with STm 4/74 and D23580 strains at different doses | 2.8 Gastrointestinal *Salmonella* infection after challenge with 4/74 or D23580 strains, with specific reference to<br>2.8.1 Time to onset in days in each group from challenge to first stool sample positive for *Salmonella* Typhimurium by culture and/or PCR<br>2.8.2 Duration of stool shedding in each group measured in days from first stool sample positive for *Salmonella* | D0 – D28 |

| | | |
|---|---|---|
| | 2.8.3 The duration of stool shedding in each group measured in days from first stool sample positive by culture and/or for *Salmonella* Typhimurium to first persistently negative stool sample (defined as three consecutive samples negative at least 48 hours apart)<br>2.8.4 The magnitude of stool shedding measured in CFU/g in quantitative stool culture analysis | |
| 2.9 To compare **microbiological** features of **bloodstream infection** following oral challenge with STm 4/74 and D23580 strains at different doses. | 2.9 *Salmonella* bloodstream infection after challenge with 4/74 or D23580 strains, with specific reference to<br>2.9.1 The proportion of participants in each group developing any bacteraemia<br>2.9.2 The severity of bacteraemia in each group as measured by<br>2.9.2.1 The duration of bacteraemia measured by time in hours from the first positive blood culture to first persistently negative blood culture | D0 – D28 |

| | | |
|---|---|---|
| | 2.9.2.2 Blood culture clearance time measured by time in hours from initiation of antibiotics treatment to first persistently negative blood culture<br>2.9.2.3 The time to onset of bacteraemia in each group, measured by hours from challenge | |
| 2.10 To compare **biochemical** and **haematological** laboratory parameters following oral challenge with STm 4/74 and D23580 strains at different doses | 2.10 Absolute values of laboratory parameters from time of challenge to Day 28 and/or Day 90, with specific reference to<br>2.10.1 Total haemoglobin (g/L)<br>2.10.2 Haemoglobin change from baseline (Hb g/l D0 – nadir Hb g/l)<br>2.10.3 Total white cell count ($x10^9$/l)<br>2.10.4 Neutrophil count ($x10^9$/l)<br>2.10.5 Lymphocyte count ($x10^9$/l)<br>2.10.6 Eosinophil count ($x10^9$/l)<br>2.10.7 Monocyte/lymphocyte ratio<br>2.10.8 Urea and electrolytes (Na+,K+, Urea, Creatinine – mmol/l)<br>2.10.9 C-reactive protein (mg/l) | D0 – D90 |

| | | |
|---|---|---|
| | 2.10.10 Liver function tests (Bilirubin [umol/l], alkaline phosphatase (ALP IU/l), alanine transaminase (ALT IU/l), Albumin (g/l) | |
| 2.11 To describe and compare the serum antibody response following oral challenge with STm 4/74 and D23580 strains at different doses. | 2.11 Absolute values of laboratory assays on serum samples measured at baseline and post challenge time points (e.g. Day 7, 14, 28, 90 and 365), including but not limited to<br>2.11.1 *S.* Typhimurium O-specific polysaccharide serum IgG and IgA concentration measured by ELISA<br>2.11.2 *S.* Typhimurium specific serum IgG and IgA concentration against other *S.* Typhimurium antigens measured by ELISA<br>2.11.3 Serum bactericidal antibody titres against *Salmonella* Typhimurium<br>2.11.4 *S.* Typhimurium specific antibody secreting cell and memory B-cell responses measured by ELISPOT | D0 – D365 |
| 2.12 To describe and compare the mucosal antibody response following oral challenge with *S.* Typhimurium 4/74 or *S.* Typhimurium D23580 strains. | 2.12 Absolute values of laboratory assays on saliva (and/or stool copro-antibodies) samples measured at baseline and post challenge time points (e.g. Day 7, 14, 28, 90 and 365) using laboratory assays including (but not limited to) | D0 – D180 |

| | | |
|---|---|---|
| | 2.12.1 S. Typhimurium specific IgG and IgA concentration measured by ELISA<br>2.12.2 Bactericidal antibody titres<br>2.12.3 Other functional antibody activity measurements including systems serology platforms | |
| 2.13 To describe and compare cell-mediated immune response following oral challenge with *S.* Typhimurium 4/74 or *S.* Typhimurium D23580 strains | 2.13 Absolute values and inter-group comparison of laboratory assays performed on peripheral blood mononuclear cells measured at baseline and post challenge time points including (but not limited to)<br>2.13.1 Description of lymphocyte populations at baseline and following challenge as measured by flow cytometry and/or CyTOF<br>2.13.2 Frequency and magnitude of S. Typhimurium specific cell-mediated immune responses as measured by ELISPOT and flow cytometry and/or CyTOF | |
| 2.14 To describe participant experience following oral challenge with STm 4/74 and D23580 strains at different doses. | 2.14 Descriptive statistics following administration of a structured questionnaire at D28 post-challenge | D28 |

### 7 Statistical Methodology

#### 7.1 General consideration

Continuous variables that follow an approximately normal distribution will be summarised using means and standard deviations. Skewed continuous variables will be summarised using medians, inter-quartile ranges. Categorical/binary variables will be summarised using frequencies and percentages.

Participant flow from screening, enrolment, through to randomisation, challenge, follow-up and analysis will be presented in a CONSORT flow diagram. This will contain the numbers of participants randomly assigned to each strain, receiving challenges, completing the study and analysed for the primary outcome.

The immune responses data are expected to be highly skewed, and the data will be log-transformed prior to analysis. The geometric mean concentration (GMC) and associated 95% confidence interval (CI) will be summarised by computing the anti-log of the mean of the log-transformed data. Data will be summarised by strains and dose levels at different time points. Values below the limit of detection for immune responses will be replaced by half the value of the lower limit.

The significance level for analysis of all endpoints will be 2-sided, and the significance level will be considered at 0.05, unless specified otherwise in the analysis section below.

#### 7.2 Baseline Characteristics

Descriptive statistics relating to participant characteristics at baseline (e.g., age, sex, ethnicity) will be calculated overall and by group. No formal statistical comparisons of baseline characteristics between randomised groups will be conducted.

##### 7.3 Primary objective analysis

The analysis of the primary endpoint will be descriptive only. The percentage of participants in each group, who meet the criteria for diagnosis of *Salmonella* Diagnosis (SD), will be calculated using a 95% Clopper-Pearson Exact confidence interval. The numerator will be the number of participants who meet the criteria for diagnosis and the denominator will include all participants excluding those who withdrew or were treated prior to Day 14 without being diagnosed.

A secondary analysis of the primary endpoint will be conducted using the Kaplan-Meier method which will include all participants. Participants who withdrew or were treated prior to Day 14 and had no *Salmonella* Diagnosis (nSD) will be censored in the analysis at the time of withdrawal or treatment. Participants who were not diagnosed and were treated at Day 14 will be counted as censoring at Day 14 in the analysis.

The CRM will provide final posterior attack rate estimates with 95% equal-tailed Bayesian credible intervals for all four dose levels and a final dose recommendation. The DSMC retains authority to confirm or override the final dose recommendation based on safety or other clinical considerations. The posterior probability that each dose achieves ≥60% attack rate (lower bound of the target window) will also be reported. The CRM will be run on the Safety Analysis Population during dose-escalation. A supplementary CRM analysis will be conducted on Analysis Population 1 to assess the sensitivity of posterior estimates to the exclusion of participants treated prior to Day 14 without a confirmed diagnosis (AT-nSD).

##### 7.4 Secondary objective analysis

The analysis for secondary endpoints will follow the primary endpoint analysis. For the time-to-event analyses of secondary endpoints (e.g., positive blood culture, oral temperature ≥ 38.0°C etc.) all participants will be included. Participants not meeting the criteria for an individual secondary endpoint will be censored in the analysis at, the time of withdrawal, the time of diagnosis or Day 14, whichever is earlier.

Area under the curve for gastrointestinal and systemic symptom severity score between Day 0 to Day 14 will be derived and summarise using the descriptive statistics (number, mean, standard deviation, median, minimum and maximum at each time point).

##### 7.5 Missing data

All available data will be used in the analyses and there will be no imputations for missing data. Participants will be analysed according to the type of challenge received for summaries by group.

##### 7.6 Procedures for Reporting deviations from the Original Statistical Plan

Any additional analysis or deviations from the analysis plan will be documented and updated according to the statistical standard operating procedure.

##### 7.7 Software

R will be used for all analysis.

#### 8 Amendment History

| Version Number | Date | Author | Reason |
|---|---|---|---|
| V1.1 | 17 March 2025 | Yujie Ni<br>Dr Christopher Smith | Initial version creation. |
