## Supplementary material for "Safety and pathovariant-independent susceptibility in a *Salmonella* Typhimurium controlled human infection model: a phase 1, randomised, double-blind, dose-escalation study": Study Protocol

Challenge Non-Typhoidal *Salmonella* (CHANTS) Study

THE CHANTS STUDY

The Development of a non-typhoidal *Salmonella* human challenge model: A safety and dose escalation study.

**Version 3.1 25NOV 2024**

MAIN SPONSOR: Imperial College London
FUNDERS: The Wellcome Trust (Award Reference 224029/Z/21/Z)
STUDY COORDINATION CENTRE: Imperial College London

Sponsor study reference: 21HH6989
IRAS Project ID: 301659
REC reference: 21/PR/0051

Protocol authorised by:

| Name & Role | Date | Signature |
|---|---|---|
| Dr Malick Gibani | 25 NOV 2024 | |

Study Management Group

Chief Investigator: Dr Malick Gibani

Co-investigators: Professor Graham Cooke; Professor Chris Chiu; Dr Chris Smith

Statistician: Dr Xinxue Liu

Study Management: Emma Smith

#### Study Coordination Centre

For general queries, supply of study documentation, and collection of data, please contact:

Study Coordinator: Dr Malick Gibani
Address: Variety Wing C, Medical School, St Mary's Hospital campus, Imperial College London, Praed St, London W2 1NY

Web address: https://www.imperial.ac.uk/people/m.gibani

**Clinical Queries**

Clinical queries should be directed to or Dr Malick Gibani or Dr Chris Smith who will direct the query to the appropriate person.

**Sponsor**

Imperial College London is the main research Sponsor for this study. For further information regarding the sponsorship conditions, please contact the Head of Regulatory Compliance at:

Research Governance and Integrity Team
Imperial College London and Imperial College Healthcare NHS Trust
Room 215, Level 2, Medical School Building
Norfolk Place
London, W2 1PG
Imperial College - Research Governance and Integrity Team (RGIT) Website

Funder
The Wellcome Trust (Reference UNS127883)

This protocol describes the CHANTS study and provides information about procedures for entering participants. Every care was taken in its drafting, but corrections or amendments may be necessary. These will be circulated to investigators in the study. Problems relating to this study should be referred, in the first instance, to the Chief Investigator.

This study will adhere to the principles outlined in the UK Policy Framework for Health and Social Care Research. It will be conducted in compliance with the protocol, the UK GDPR and other regulatory requirements as appropriate.

Table of Contents Page No

Glossary of abbreviations ... 11
Keywords ... 14
Study summary ... 15
Reference diagram ... 22
Lay summary ... 24
1. Introduction ... 25
1.1. Background ... 25
1.2. Vaccines for iNTS disease ... 26
1.3. Application of CHIM studies for vaccine development ... 27
1.4. *Salmonella* human challenge studies ... 28
1.4.1. Typhoidal *Salmonella* challenge studies ... 28
1.4.2. Non-typhoidal Salmonella challenge studies ... 29
1.5. Clinical Considerations in the development of an iNTS CHIM ... 30
1.5.1. Transmission ... 30
1.5.2. *Salmonella* Gastroenteritis ... 30
1.5.2.1. Post-infectious complications of gastrointestinal Salmonella infection ... 31
1.5.2.1.1. Reactive arthritis ... 31
1.5.2.1.2. Post-infectious irritable bowel syndrome ... 31
1.5.2.2. Management of *Salmonella* gastroenteritis ... 31
1.5.2.2.1. Supportive management ... 31
1.5.2.2.2. Antibiotics ... 32
1.5.3. Invasive *Salmonella* infection ... 33
1.5.3.1. Complications of invasive *Salmonella* infection ... 33
1.5.3.1.1. Endovascular infection ... 33
1.5.3.1.2. Meningitis ... 34
1.5.3.1.3. Osteomyelitis ... 34
1.5.3.2. Management of NTS bacteraemia. ... 34
1.5.4. Shedding ... 34
1.5.4.1. Acute and convalescent shedding ... 34
1.5.4.1.1. The impact of antibiotics on acute and convalescent shedding ... 34
1.5.4.2. Chronic carriage ... 35
1.5.5. Infectious Dose ... 35
1.5.6. Challenge strain selection ... 35
1.5.6.1. ST313 ... 35
1.5.6.2. ST19 ... 36
1.6. Rationale for the current study ... 36
2. Study objectives ... 37
3. Study design ... 44
3.1. Study design ... 44
3.2. Study outcome measures ... 46
3.2.1. Primary endpoint ... 46
3.2.2. Secondary disease endpoints ... 46
3.2.3. Severe Salmonellosis ... 47
3.3. Definitions ... 47

6.8.2. *Salmonella* diagnosis and treatment visits (SD, SD+24hrs, SD+48hrs, SD+72hrs, SD+96hrs) 84

#### Glossary of abbreviations

***Table 1 – Glossary of abbreviations***

| | |
|---|---|
| AE | Adverse event |
| AESI | Adverse event of special interest |
| ALP | Alkaline phosphatase |
| ALT | Alanine transaminase |
| AR | Adverse reaction or Attack Rate |
| ASC | Antibody secreting cell |
| AST | Aspartate transaminase |
| BD | bis in die (Latin: twice a day; prescription medicines) |
| BMI | Body Mass Index |
| CI | Chief investigator or confidence interval |
| CFU | Colony forming unit |
| CMI | Cell-mediated immunity |
| COPS | Core and O-polysaccharides conjugate vaccines |
| CRF | Case Report Form |
| CRO | Contract Research Organisation |
| CRP | C-reactive protein |
| CT | Clinical Trials |
| CTA | Clinical Trials Authorisation |
| CTL | Cytotoxic T-lymphocyte |
| DSMC | Data Safety and Monitoring Committee |
| DSUR | Development Safety Update Reports |
| EDTA | Ethylenediamine tetraacetic acid |
| ELISA | Enzyme linked immunosorbent assay |
| ELISPOT | Enzyme linked immunosorbent spot assay |
| EPI | Expanded Program on Immunisation |
| ESR | Erythrocyte sedimentation rate |
| GCP | Good Clinical Practice |
| GMT | Geometric Mean Titre |
| GP | General Practitioner |
| HADS | Hospital Anxiety and Depression Scale |
| HIV | Human Immunodeficiency Virus |
| HPU | Health Protection Unit |
| HRA | Health Research Authority |
| IB | Investigators Brochure |
| ICF | Informed Consent Form |
| ICH | International Conference of Harmonisation |
| ICHT | Imperial College Healthcare NHS trust |
| ICL | Imperial College London |
| ICRF | NIHR/Wellcome Trust Imperial Clinical Research Facility |
| IFN | Interferon |
| IgA | Immunoglobulin A |
| IgM | Immunoglobulin M |
| IgG | Immunoglobulin G |
| IL | Interleukin |
| IMP | Investigational medicinal product |

| | |
|---|---|
| iNTS | Invasive non-typhoidal *Salmonella* |
| IRB | Independent Review Board |
| ISF | Investigator site file |
| LFT | Liver function tests |
| LLN | Lower limit of normal |
| LPS | Lipopolysaccharide |
| MAX | Maximum (prescription medication) |
| MHRA | Medicines and Healthcare products Regulatory Agency |
| NIH | National Institute of Health |
| NRES | National Research Ethics Service |
| NTS | Non-typhoidal *Salmonella* |
| NWLP | North West London Pathology Laboratory (Charing Cross Hospital, Imperial Healthcare NHS Trust) |
| OSP | O-specific polysaccharide |
| OPK | Opsonophagocytic killing (OPK) assay |
| PBMC | Peripheral blood mononuclear cell |
| PCR | Polymerase chain reaction |
| PE | Protective Effect |
| PHE | Public Health England |
| PIL | Participant/ Patient information leaflet |
| PO | Per oral (by mouth) |
| PR | Per rectum (by rectum) |
| PRN | pro re nata (latin: as required; prescription medicines) |
| QDS | quater die sumendus (Latin: four times a day; prescription medicines) |
| R&D | NHS Trust Research & Development Department |
| REC | Research Ethics Committee |
| REC ref. | Research Ethics Committee reference |
| RPM | Revolutions per minute |
| RRT | Renal replacement therapy |
| SAE | Serious adverse event |
| SAR | Serious adverse reaction |
| SBA | Serum bactericidal assay |
| SCV | *Salmonella* Containing Vacuole |
| SMP(C) | Summary of Medicinal Product (Characteristics) |
| SOP | Standard Operating Procedure |
| SPI | *Salmonella* pathogenicity island |
| Spp. | Species (plural) |
| ST | Sequence type |
| SUSAR | Suspected unexpected serious adverse reactions |
| T3SS | Type 3 Secretion System |
| SD | *Salmonella* Diagnosis |
| TDS | ter die sumendum |
| TMF | Trial Master File |
| TOPS | The Over volunteering Prevention System (see: http://www.tops.org.uk) |
| TSB | Tryptone soya broth |
| TSC | Trial Steering Committee |
| ULN | Upper limit of normal |
| V[n] | Visit [number] |
| Vac | Vacutainer |

| | |
|---|---|
| Vi | Virulence antigen |
| WBC | White blood cell/count |
| XLD | Xylose lysine deoxycholate |
| 95%CI | 95% Confidence Interval |

#### Keywords

*Salmonella*
Non-typhoidal *Salmonella*
Human challenge model
Controlled human infection model
Dose escalation
D23580
4/74

#### Study summary

***Table 2 - Study summary***

| | | |
|---|---|---|
| **Study Title** | The Development of human challenge model for non-typhoidal *Salmonella*: A safety and dose escalation study. | |
| **Short title (internal ref.)** | CHANTS study | |
| **Clinical Phase** | Phase 1 | |
| **Study Design** | Double-blinded randomised human infection challenge study | |
| **Study Participants** | Healthy adults aged 18 to 50 years inclusive | |
| **Planned Sample Size** | 20 - 80 | |
| **Follow up duration** | 12 months | |
| **Planned Trial Period** | Clinical phase: April 2023 to April 2024 | |
| **Objectives and Endpoints** | | |
| | **Objective(s)** | **Endpoint(s)** |
| **Primary endpoints** | 1.1. To determine the dose in CFU of *Salmonella enterica* subspecies *enterica* serovar Typhimurium (*S.* Typhimurium) 4/74 and D23580 strains required for 60-75% of volunteers to develop systemic *Salmonellosis* following oral challenge | 1.1. The proportion of participants who develop either:<br>1.1.1. Fever ≥38°C on ≥2 occasions ≥12 hours apart **AND/OR**<br>1.1.2. *Salmonella* Typhimurium bacteraemia |
| **Secondary** | 2.1 To describe the *Salmonella* colonisation rate following oral challenge with *Salmonella* Typhimurium 4/74 and D23580 strains at different doses | 2.1 The proportion from whom *Salmonella* Typhimurium is isolated from stool on ≥2 occasions ≥48 hours from challenge at different doses of each strain. |
| | 2.2 To describe the *Salmonella* gastroenteritis rate following oral challenge of *Salmonella* Typhimurium 4/74 and D23580 strains at different doses. | 2.2 The proportion of participants at different doses at each strain developing<br>2.2.1 Severe diarrhoea **and/or**<br>2.2.2 Moderate diarrhoea plus<br>2.2.2.1 Fever ≥38°C on ≥1 occasion **and/or** |

| | | |
|---|---|---|
| | | 2.2.2.2 ≥1 Grade 2 gastrointestinal symptoms (abdominal pain, nausea, vomiting, tenesmus) |
| | 2.3 To determine the persistent fever rate following oral challenge with *Salmonella* Typhimurium 4/74 and D23580 at different doses. | 2.3 The proportion of participants who develop fever ≥38°C on ≥2 occasions ≥12 hours apart at different doses of each strain. |
| | 2.4 To describe the *Salmonella* bacteraemia rate following oral challenge with *Salmonella* Typhimurium 4/74 and D23580 at different doses. | 2.4 The proportion of participants developing *Salmonella* Typhimurium bacteraemia at any timepoint following challenge at different doses of each strain |
| | 2.5 To describe the rate of systemic Salmonellosis according to **an alternative composite diagnostic criterio**n following oral challenge of *Salmonella* Typhimurium 4/74 and D23580 at different doses. | 2.5 The proportion of participants meeting the criteria for a composite diagnosis of *Salmonellosis* at different doses of each strain defined as any of 1) *Salmonella* Typhimurium is isolated from stool on ≥2 occasions ≥48 hours from challenge and/or 2) *Salmonella* gastroenteritis and/or 3) fever ≥38°C on ≥2 occasions ≥12 hours apart and/or 4) *Salmonella* Typhimurium bacteraemia detected by blood culture and/or novel molecular methods |
| | 2.6 To describe the safety of oral challenge with *S*. Typhimurium 4/74 and D23580 strains | 2.6 The proportion of participants at different doses of each strain reporting<br>• Adverse events,<br>• Adverse events of special interest,<br>• SAEs,<br>• SUSARs.<br>• Concomitant medication usage as outlined in the study protocol |
| | 2.7 To compare **clinical** features following oral challenge with *S.* Typhimurium 4/74 or *S.* Typhimurium D23580 strains | 2.7 A comparison of the clinical features of *Salmonella* infection after challenge with 4/74 or D23580 strains, with specific reference to<br>2.7.1 The proportion of participants in each group developing any diarrhoea.<br>2.7.2 The severity of diarrhoea in each group, as measured by<br>2.7.2.1 Stool volume in ml/24hrs<br>2.7.2.2 Frequency of bowel motions/24hrs<br>2.7.2.3 Total gastrointestinal symptom severity score |

| | | |
|---|---|---|
| | | calculated by summing numerical values assigned to the severity of all solicited gastrointestinal symptoms between Day 0 to Day 14 (0=not present; 1=mild; 2=moderate; 3=severe).<br>2.7.2.4 Total systemic symptom severity score calculated by summing numerical values assigned to the severity of all solicited gastrointestinal and systemic symptoms between Day 0 to Day 14 (0=not present; 1=mild; 2=moderate; 3=severe).<br>2.7.2.5 Area under the curve for gastrointestinal and systemic symptom severity score between Day 0 to Day 14<br>2.7.2.6 Time to honest of diarrhoea in each group, measured in hours since challenge to the first grade 6/7 stool.<br>2.7.3 The proportion of participants in each group developing any fever ≥38°C<br>2.7.4 Fever clearance time of febrile participants in each group measured by time in hours to first temperature <38°C lasting ≥12hrs<br>2.7.5 The proportion of participants in each group reporting the following solicited symptoms at any time:<br>2.7.5.1 headache,<br>2.7.5.2 malaise<br>2.7.5.3 anorexia<br>2.7.5.4 abdominal pain<br>2.7.5.5 nausea<br>2.7.5.6 vomiting<br>2.7.5.7 dysentry<br>2.7.5.8 myalgia<br>2.7.5.9 arthralgia<br>2.7.5.10 cough<br>2.7.5.11 rash<br>2.7.6 Duration of solicited symptoms measured in days from first symptom onset to complete resolution of symptoms |

| | | |
|---|---|---|
| | | 2.7.7 Severity of solicited symptoms measured by<br>2.7.7.1 The proportion of participants with maximum symptom severity score graded as mild, moderate, or severe following challenge<br>2.7.7.2 The proportion of participants meeting the criteria for severe *Salmonellosis*<br>2.7.7.3 Symptom severity scores for individual solicited symptoms calculated by summing numerical values assigned to the severity of individual solicited symptoms between Day 0 to Day 14 (0=not present; 1=mild; 2=moderate; 3=severe |
| | 2.8 To compare **microbiological** features of **gastrointestinal infection** following oral challenge with *S.* Typhimurium 4/74 or *S.* Typhimurium D23580 strains | 2.8 Gastrointestinal *Salmonella* infection after challenge with 4/74 or D23580 strains, with specific reference to<br>2.8.1 Time to onset in days in each group from challenge to first stool sample positive for *Salmonella* Typhimurium by culture and/or PCR<br>2.8.2 Duration of stool shedding in each group measured in days from first stool sample positive for *Salmonella*<br>2.8.3 The duration of stool shedding in each group measured in days from first stool sample positive by culture and/or for *Salmonella* Typhimurium to first persistently negative stool sample (defined as three consecutive samples negative at least 48 hours apart)<br>2.8.4 The magnitude of stool shedding measured in CFU/ml in quantitative stool culture analysis |
| | 2.9 To compare **microbiological** features of **bloodstream infection** following oral challenge with *S.* Typhimurium 4/74 or *S.* Typhimurium D23580 strains | 2.9 *Salmonella* bloodstream infection after challenge with 4/74 or D23580 strains, with specific reference to<br>2.9.1 The proportion of participants in each group developing any bacteraemia |

| | | |
|---|---|---|
| | | 2.9.2 The severity of bacteraemia in each group as measured by<br>2.9.2.1 The duration of bacteraemia measured by time in hours from the first positive blood culture to first persistently negative blood culture<br>2.9.2.2 Blood culture clearance time measured by time in hours from initiation of antibiotics treatment to first persistently negative blood culture<br>2.9.2.3 Magnitude of bacteraemia measured in CFU/ml in quantitative blood culture samples collected immediately prior to the initiation of treatment<br>2.9.2.4 The time to onset of bacteraemia in each group, measured by hours since challenge |
| | 2.10 To compare biochemical and haematological laboratory parameters following oral challenge with *S.* Typhimurium 4/74 or *S.* Typhimurium D23580 strains | 2.10 Absolute values of laboratory parameters from time of challenge to Day 28 and/or Day 90, with specific reference to<br>2.10.1 Total haemoglobin (d/L)<br>2.10.2 Haemoglobin change from baseline (Hb g/l D0 – nadir Hb g/l)<br>2.10.3 Total white cell count (x$10^9$/l)<br>2.10.4 Neutrophil count (x$10^9$/l)<br>2.10.5 Lymphocyte count (x$10^9$/l)<br>2.10.6 Eosinophil count (x$10^9$/l)<br>2.10.7 Monocyte/lymphocyte ratio<br>2.10.8 Urea and electrolytes (Na+,K+, Urea, Creatinine – mmol/l)<br>2.10.9 C-reactive protein (mg/l)<br>2.10.10 Liver function tests (Bilirubin [umol/l], aspartate transaminase (AST IU/l), alkaline phosphatase (ALP IU/l), alanine transaminase (ALT IU/l), Albumin (g/l) |
| | 2.11 To describe and compare the serum antibody response following oral challenge with *S.* Typhimurium 4/74 or *S.* Typhimurium D23580 strains | 2.11 Absolute values of laboratory assays on serum samples measured at baseline and post challenge time points (e.g. Day 7, 14 , 28, 90 and 365), including but not limited to<br>2.11.1 *S.* Typhimurium O-specific polysaccharide serum IgG and |

| | | |
|---|---|---|
| | | IgA concentration measured by ELISA<br>2.11.2 *S*. Typhimurium specific serum IgG and IgA concentration against other S. Typhimurium antigens measured by ELISA<br>2.11.3 Serum bactericidal antibody titres against *Salmonella* Typhimurium<br>2.11.4 *S*. Typhimurium specific antibody secreting cell and memory B-cell responses measured by ELISPOT<br>2.11.5 Other functional antibody activity measurements including systems serology platforms. |
| | 2.12 To describe and compare the mucosal antibody response following oral challenge with *S*. Typhimurium 4/74 or *S*. Typhimurium D23580 strains. | 2.12 Absolute values of laboratory assays on saliva (and/or stool copro-antibodies) samples measured at baseline and post challenge time points (e.g. Day 7, 14 , 28, 90 and 365)using laboratory assays including (but not limited to)<br>2.12.1 S. Typhimurium specific IgG and IgA concentration measured by ELISA<br>2.12.2 Bactericidal antibody titres<br>2.12.3 Other functional antibody activity measurements including systems serology platforms |
| | 2.13 To describe and compare cell-mediated immune response following oral challenge with *S*. Typhimurium 4/74 or *S*. Typhimurium D23580 strains | 2.13 Absolute values and inter-group comparison of laboratory assays performed on peripheral blood mononuclear cells measured at baseline and post challenge time points including (but not limited to)<br>2.13.1 Description of lymphocyte populations at baseline and following challenge as measured by flow cytometry and/or CyTOF<br>2.13.2 Frequency and magnitude of S. Typhimurium specific cell-mediated immune responses as measured by ELISPOT and flow cytometry and/or CyTOF |

| | | |
|---|---|---|
| | 2.14 To describe participant experience following oral challenge with *S.* Typhimurium 4/74 or *S.* Typhimurium D23580 strains | 2.14 Descriptive statistics following administration of a structured questionnaire at baseline and post-challenge |
| **Tertiary** | 3.1. To investigate how the human microbiota interacts with oral challenge of *S.* Typhimurium 4/74 or *S.* Typhimurium D23580 strains | 3.1 Pathogen specific and metagenomic analysis of stool and/or saliva to measure constituent microbiological flora (and their metabolome, transcriptome and anti-microbial resistance profile) at baseline and post-challenge time points |
| | 3.2. To investigate novel diagnostic methods for detecting *S.* Typhimurium infection | 3.2 Exploratory analysis of blood and faecal samples, including novel molecular techniques |
| | 3.3. To describe and compare additional host immune responses following oral challenge of *S.* Typhimurium 4/74 or *S.* Typhimurium D23580 strains. | 3.3 Analysis of B-cell repetoire, plasma cytokine profile and kinetics, additional exploratory immunology |
| | 3.4. To evaluate environmental contamination with *Salmonella* enterica in the near patient environment | 3.4 The pattern and proportion of environmental samples testing positive for *Salmonella* Typhimurium by culture or molecular methods |
| | 3.5. To evaluate host-pathogen interactions following *Salmonella* infection including the presence and mechanisms of *Salmonella* persister infected macrophages | 3.5 Exploratory analysis of infected immune cells from blood including, but not limited to, single-cell RNA-seq. |
| Challenge agent – Dose and formulation | | |
| 1) | *Salmonella enterica* subspecies *enterica* serovar Typhimurium strain 4/74 | Dose range from $10^1$-$10^6$ CFU suspended in 0.9% saline and administered with sodium bicarbonate prior to oral ingestion |
| 2) | *Salmonella enterica* subspecies *enterica* serovar Typhimurium strain D23580 | Dose range from $10^1$-$10^6$ CFU suspended in 0.9% saline and administered with sodium bicarbonate prior to oral ingestion |

**Reference diagram**

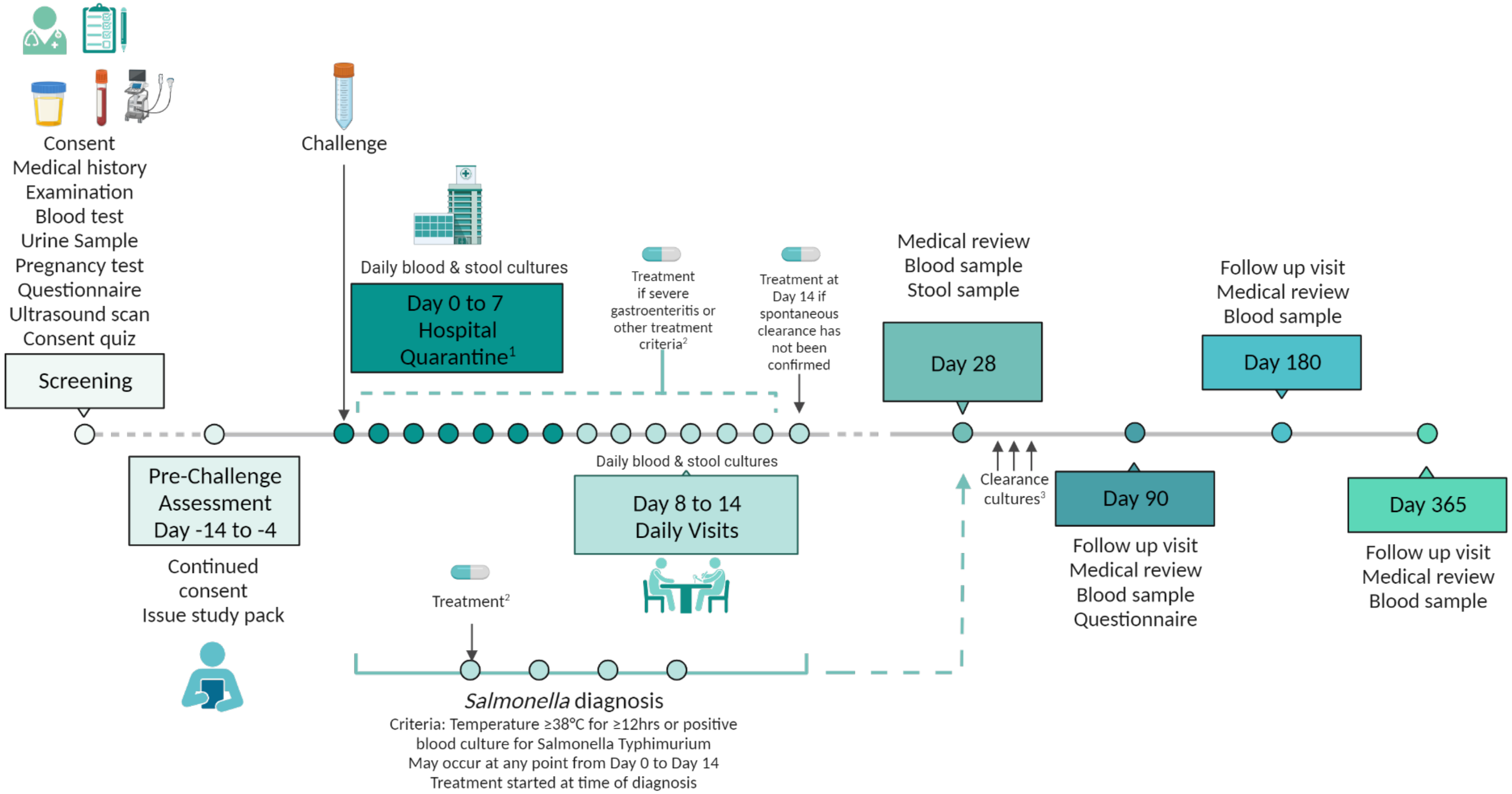

Figure 1 – Study reference diagram. 1) ***The quarantine period is expected to last 7 days. In some circumstances (i.e. if there are ongoing symptoms of diarrhoea after 7 days), participants may be asked to remain in quarantine until these symptoms have resolved. In rare circumstances, participants can be released from quarantine before day 7 – please see text below for details.*** 2) Treatment criteria detailed below. 3) Three consecutive stool samples testing negative for Salmonella (at least 48 hours apart) will be needed post-antibiotic treatment (unless spontaneous clearance has been confirmed).

#### Lay summary

This document details the study protocol for a human infection challenge study. Our research team are studying a large group of bacteria called *Salmonella*. There are over 2000 different types of *Salmonella,* each of which are subtly different. The specific species of bacteria we are studying is called ***Salmonella* Typhimurium**. This belongs to a group of *Salmonella* bacteria that we call **non-typhoidal *Salmonella*** – or **NTS** for short.

In most people, NTS will cause a gut infection, called gastroenteritis. The usual symptoms are diarrhoea, stomach-ache, and fever, which gets better after a few days. In some vulnerable people, the infection can be more severe. It can get inside the bloodstream or parts of the body outside the gut. We call this more severe form of the disease “invasive NTS” (or **iNTS** for short). This more severe form almost exclusively occurs in people who have a poorly functioning immune system (like HIV infection), or those who are very young, very old, or with specific risk factors like malaria or sickle cell disease.

Invasive *Salmonella* (INTS) is a very big problem globally, but especially in sub-Saharan Africa. We think it affects at least over half-a-million people every year. It mostly affects very young children (aged under 5), most of whom are malnourished or also have other illnesses, like malaria, sickle cell disease or HIV. We are interested in finding ways to prevent this infection, such as vaccines.

In this study, we will be undertaking a ‘challenge’ with one of two strains of NTS called *Salmonella* Typhimurium. Participants will be asked to swallow a drink containing live *Salmonella* under medical supervision. After this, they will be closely monitored for two weeks. The first 7 days will involve staying in hospital. The next seven days, they come to the clinic daily. They will be treated with antibiotics when they show symptoms of infection, like diarrhoea or fever. There is a very small risk of a more severe illness, but this is extremely rare in young, healthy people.

We will use **two strains** of the *Salmonella* Typhimurium bacteria. Half of the participants in this study will be challenged with a strain of *Salmonella* that is common in the UK and other half will be challenged with a strain that is more common in Africa. There is some data from the lab which suggests that these bacteria behave differently, but we don’t yet know the significance of this in healthy people. We want to decide which is the best strain of bacteria for future studies.

This study is called a “dose escalation” study. This means that we start the challenge with a low dose of bacteria and gradually increase (or decrease) until about 60% to 75% of people exposed to the bacteria develop particular symptoms of *Salmonella* infection. In addition, we will study how the immune system responds to the different strains of *Salmonella*, to understand how it prevents *Salmonella* disease. This will add to our general understanding of the bacteria and could aid vaccine development.

From start to finish, the study lasts for 1 year and includes 21 to 26 visits.

### 1. Introduction

This protocol describes the challenge non-typhoidal *Salmonella* (CHANTS) study. This is a first-in-human phase 1, double-blinded, randomised, dose-escalation human infection study, conducted in healthy volunteers aged 18 to 50 years.

The overall aim of this study is to establish a controlled human infection model of invasive non-typhoidal *Salmonella* (iNTS) infection. We will challenge healthy volunteers with two strains of *Salmonella* Typhimurium. Participants will be admitted to an inpatient quarantine facility after challenge and treated with antibiotics when they meet defined criteria. The primary objective of the study is to perform a dose escalation to determine the infectious dose required for 60-75% of volunteers to develop *Salmonellosis* using a composite diagnostic criterion. The secondary objectives of the study are to describe and compare the clinical and laboratory features following controlled human infection with *Salmonella* Typhimurium. It is hoped that the successful establishment of an NTS human challenge model can be used in the future to test candidate vaccines for NTS disease.

#### 1.1. Background

*Salmonella enterica* are amongst the most frequently isolated pathogens in patients with community onset bloodstream infections in Africa and Asia.[1] In particular, invasive disease caused by non-typhoidal *Salmonella* serovars (iNTS) is common across Africa, where the most recent data estimate that iNTS is responsible for 535 000 cases, 77 500 deaths and 4·26 million DALYs lost in 2017.[2] The precise burden of disease is, however, relatively poorly understood and estimates vary depending on case-definitions and methods of surveillance.[3,4] The WHO has recently listed *Salmonella enterica* as a priority pathogen, identified as one of 12 families of bacteria that pose the greatest risk to human health through rising antimicrobial resistance.[5]

Invasive NTS disease typically presents as a febrile illness with clinical signs of sepsis, often without gastrointestinal symptoms.[6] The case-fatality rate is estimated at between 15-20%.[3,7] The high mortality of iNTS is explained by its strong association with human immunodeficiency. Recognised risk factors include malnutrition, recent malaria and chronic anaemia (including sickle cell disease) in children, and HIV infection and malignancy in adults.[6] Primary NTS bacteraemia is also observed in patients with defects in neutrophil function (e.g. chronic granulomatous disease) and those with defects in IL-12/23 pathway.[4,8] Immune defects predisposing patients to iNTS have been studied most extensively in HIV disease, involving depletion of mucosal CD4+ T-cells, systemic cytokine dysregulation and dysregulated humoral immunity, coupled with hypergammaglobulinaemia and aberrant response to O-specific polysaccharides.[6,9] The incidence of iNTS initially rises in HIV-infected patients starting antiretroviral therapy and then falls, but remains much higher than in HIV-uninfected controls.[10]

The NTS serovars commonly isolated from bacteraemic patients vary by location, but usually include *Salmonella enterica* subspecies *enterica* serovars Typhimurium and Enteritidis.[11] *S.* Typhimurium accounts for approximately two-thirds of iNTS in Africa.[11] Sequencing of isolates from across Africa has identified *Salmonella* Typhimurium multilocus sequence type 313 (ST313) as the major cause of invasive disease, which is distinct from the sequence types 19 and 34 that are more frequently associated with gastrointestinal disease in high-income settings such as the UK.

ST313 appears to have undergone sequential evolutionary changes that converge towards a genotype associated with invasive disease, in the same way as reported for *Salmonella* Typhi and Paratyphi serovars. The key changes are characterized by stepwise genomic degradation typified by multiple gene deletions and pseudogene formation, often in genes associated with gastrointestinal survival.[12] The data suggests that ST313 has adapted to a human niche and can evade mucosal immunity.[13–15] Similar findings have been observed in invasive *S*. Enteritidis ST11 isolates from Africa.[16] Transmission of invasive isolates is complex and likely involves a combination of zoonotic reservoirs (particularly for *S*. Enteritidis) as well as human-to-human transmission.[17,18] Invasive NTS isolates are increasingly associated with multi-drug resistance, significantly limiting treatment options.[12,19–21]

##### 1.2. Vaccines for iNTS disease

There is increasing interest in the development of broadly protective vaccines for invasive *Salmonella* disease. Vaccines for NTS would represent a valuable public-health tool in Sub-Saharan Africa, in part because other effective methods for disease control are slow and cost prohibitive in many endemic countries.

Several candidate vaccines are in development, including conjugate vaccines, live-attenuated oral vaccines and those using the generalised modules for membrane antigens (GMMA) platform, typically in multivalent formulations.[22,23]

The University of Maryland and Bharat Biotech are developing a conjugate vaccine comprising core and O-polysaccharides conjugated to serovar specific phase 1 flagellin protein FliC (COPS conjugate vaccines).[24] *S.* Enteritidis and *S.* Typhimurium COPS vaccines are immunogenic and able to protect against homologous intra-peritoneal challenge with *S*. enteritidis and *S.* Typhimurium in a CD-1 mouse model.[25–27] This vaccine is being developed as a trivalent formulation comprising *S.* Typhi Vi-TT; *S.* typhimurium COPS FliC and *S.* enteritidis COPS FliC conjugates. Rabbits immunised with this formulation mount a strong anti-O-antigen IgG and bactericidal antibody response, which was able to protect mice from fatal challenge following passive transfer.[27] A phase 1 randomized, placebo-controlled dose-escalation study of this trivalent *Salmonella* vaccine is currently underway, with an estimated study completion date of September 2022 (NCT03981952).[28]

GSK vaccines for Global health are developing a bivalent *S.* enteritidis and *S.* typhimurium vaccine using the generalised modules for membrane antigens (GMMA) platform. GMMA vaccines are derived from wild-type parent strains engineered with *tolR* mutations to induce continuous over-blebbing. GMMA vaccines allow multiple antigens to be presented in their natural confirmational state, potentially increasing the breadth of vaccine induced immune responses. In pre-clinical studies, GMMA vaccines were at least as immunogenic as O-antigen $CRM_{197}$ conjugate comparators at inducing anti-O IgG. They also generated greater serum bactericidal antibody responses than glycoconjugates and were associated with reduced bacterial burden in animal models.[29] Phase 1 trials are scheduled to commence in the UK in 2022, followed by studies in Kenya, through the Vacc-iNTS consortium.

In addition, Boston Children's hospital are developing a bivalent vaccine against *S*. Typhimurium and *S*. Enteritidis using the Mulitple-Antigen Presenting System (MAPS). MAPS is a novel conjugation system that leverages biotin-rhizavidin pairing to link polysaccharides and proteins. The International Vaccine Institute (IVI) are also collaborating with SK Biosciences and Biofarma on the development of O-antigen conjugates, and new live-attenuated oral vaccines are also in pre-clinical development.[22,30]

Vaccine development for iNTS is hampered, in part, by an incomplete understanding of mechanisms and determinants of immunity during natural infection, as well as a lack of commercial incentives.[9,31] Non-typhoidal *Salmonella* are facultative intracellular pathogens adapted for survival with the intracellular niche of immune cells,  and also capable of cell-free survival. Much is known about the complex life-cycle and pathogenesis of *Salmonella,* consistent with a role for cellular immunity to clear disease and antibody to prevent fatal bacteraemia.[32]

Several lines of evidence indicate a role for antibodies in conferring a degree of protection to iNTS disease. In particular, acquisition of bactericidal antibodies inversely corresponds to the age at which African children are susceptible to iNTS disease.[33,34] Nevertheless, inhibition of killing of *S*. Typhimurium occurs in the presence of high-level of antibodies against O-antigen in HIV infected adults, thought to be due to aberrant response to O-specific polysaccharides.[9] Important unknowns include the antigenic targets of bactericidal antibodies beyond O-specific polysaccharides and the role of other antibody effector mechanisms, which remain to be fully elucidated.[35]

Data from animal models, coupled with the strong association of iNTS with advanced HIV infection, indicates that robust T-cell immunity is required for full elimination of *Salmonella*.[32]

The development of effective NTS vaccines can be improved by better understanding the basis of protective immunity against iNTS disease in humans.

##### 1.3. Application of CHIM studies for vaccine development

There is growing intertest and regulatory awareness of the utility of human infection challenge studies in vaccine development, particularly for diseases which occur sporadically, or which are difficult to conduct efficacy trials. A human infection challenge may significantly advance or accelerate vaccine development, but may conversely hamper product development if clinically meaningful endpoints are not used or the threshold for efficacy is too high.[36] A critical role for human challenge  studies is to better understand the nature and type of immune responses required for vaccine induced protection from disease, and to identify correlates of protection[37,38], such as the application of a *Shigella* CHIM to identify of serum anti-LPS IgG as a correlate of protection against *Shigellosis.*[39]

There are a number of instances where human challenge models have accelerated vaccine candidates on a pathway to licensure, in particular for enteric pathogens.[40–42] For example, the live attenuated cholera vaccine CVD 103-HgR was licensed as a travel vaccine on the basis of safety, immunogenicity and efficacy data arising from CHIM studies.[42]

Another example is a Vi-tetanus toxoid conjugate vaccine against typhoid fever (Typbar-TCV®, Bharat Biotech). This first demonstrated efficacy of 57% against culture confirmed typhoid fever in a human challenge model conducted in the UK.[40] Data from the human challenge model contributed in part to recommendations from WHO SAGE to support programmatic use of Vi-TT in high burden settings in children aged 6 months and over. The same vaccine progressed to phase 3 clinical trials in Nepal, where it demonstrated efficacy of 82% against culture confirmed typhoid fever in children aged 6 months to 16 years.[41] Notably, the efficacy in a clinical setting was comparable to a higher efficacy estimate of 89% was achieved in the challenge model using a secondary disease endpoint.[40]

The R21/Matrix-M malaria vaccine first demonstrated 81% efficacy in achieving sterile immunity in a malaria human challenge model in UK adults [43]– a finding which is supported by preliminary data from a phase 2 study in Burkina Faso where the vaccine demonstrated 77% efficacy against malaria at 1 year in children aged 5 to 17 months [44].

Despite these notable examples, human challenge studies are – in general – considered by regulators and policy makers as studies that can be suggestive of efficacy, but that typically require further evidence from field trials before directly contributing to licensure. In part, this stems from concerns that such studies fail to accurately reflect natural disease, with reference to the population under study, the inoculum used, and the endpoints measured. Nevertheless, the malaria [43,44] and typhoid [40,41] cases-studies indicate that efficacy signals from human challenge studies conducted in adults in high-income settings can – in some instances – translate to comparable efficacy readout in children in low-and-middle income countries.

##### 1.4. *Salmonella* human challenge studies

Human challenge studies using *Salmonella enterica* have existed for several decades. The majority of these have used the typhoidal *Salmonella* serovars Typhi and Paratyphi A. There is less experience in using non-typhoidal *Salmonella* serovars. The re-established typhoidal *Salmonella* challenge studies provide a framework to develop an NTS challenge model but will require disease specific modifications to the study design.

###### 1.4.1. Typhoidal *Salmonella* challenge studies

The rationale for the development of *Salmonella* Typhi and Paratyphi A human challenge studies stems – in part – from the fact that these serovars are human-host restricted pathogens. A human challenge model of typhoid infection was first developed at the University of Maryland in the 1950s to 1970s[45,46]. These studies involved almost 2,000 participants and described the development of humoral immunity after inoculation with *S*. Typhi in vaccinated and unvaccinated volunteers[47]. Studies in Maryland were terminated due to concern regarding the use of prisoners and other vulnerable groups in clinical research; the prison environment was considered overly coercive, despite the ethically progressive nature in which the Maryland studies were performed.

A human typhoid and paratyphoid challenge model was developed at the University of Oxford by the Oxford Vaccine Group between 2009 and 2012 (OVG 2009/10 Oxford A REC ref. 10/H0604/53, OVG 2011/02 Oxford A REC ref. 11/SC/0302). This new model used sodium bicarbonate as a gastric acid neutralising buffer, allowing a lower challenge dose to be used and resulting in a more consistent pattern of clinical infection. The model was also set up to be performed using outpatient community volunteers, and was demonstrated to be safe and acceptable to participants [48].

The first study in Oxford enrolled 41 healthy volunteers, who were challenged with escalating dose levels of *S.* Typhi delivered in a sodium bicarbonate buffer solution. Participants were then followed daily until diagnosed with typhoid infection or day 14, at which point they were treated with antibiotics. Participants developing infection tolerated the clinical course well, and all participant symptoms responded to antibiotic treatment. Four participants met predetermined criteria for severe infection (2 recorded temperatures exceeding 40°C and had >grade 3 laboratory abnormalities), however, no participants required hospital admission, intravenous antibiotics, or fluids. Participants were promptly treated on confirmation of typhoid diagnosis and no instances of stool shedding after treatment or transmission to secondary contacts were detected [48].

This model was then used in a subsequent clinical trial (OVG 2011/02 Oxford A REC ref. 11/SC/0302) to assess the protective efficacy of a novel, single dose oral typhoid vaccine (M01ZH09). The typhoid model was validated by demonstration of a consistent attack rate of 66% in the placebo vaccinated group (an attack rate of 65% had been demonstrated in the initial dose-finding study), and by demonstration of a protective effect by 3 doses of the licensed Ty21a vaccine (PE=35%). The study was performed safely, and challenge was well tolerated. The most significant study related SAE was related to use of ciprofloxacin antibiotics, rather than vaccination or illness related to challenge; the participant developed symptoms of acute depression on a background of low mood, which resolved over time without pharmacological treatment. One participant was withdrawn from the study following challenge, as they vomited (deliberately) within several hours of ingesting the challenge agent.

In 2015-2016, 103 participants were challenged with *S.* Typhi 28 days after randomisation to vaccination with one of three vaccines: a novel Vi-tetanus-toxoid conjugate vaccine; a licensed Vi-polysaccharide vaccine or a control Men-ACWY conjugate vaccine (OVG 2014/08 Oxford A REC ref. 14/SC/1427). This study demonstrated that a Vi-tetanus-toxoid conjugate vaccine conferred 57% protection against typhoid fever within the context of the model, supporting the further development of Vi-conjugate vaccines [40].

Building upon the experiences from these studies, the Oxford Vaccine Group has undertaken several additional challenge studies, including the first *Salmonella* Paratyphi A human challenge model (OVG 2013/07 Oxford A REC ref. 14/SC/0004), studies of the mechanisms and determinants of systemic and mucosal immunity to *S.* Typhi and Paratyphi A infection (OVG 2014/01 Oxford A REC ref 14/SC/1204).

Genetically modified strains of *S*. Typhi have also been used in the human challenge model. In 2017, a study was conducted to compare the response to oral challenge with wild-type *S.* Typhi with a comparator knock-out strain lacking a key virulence factor termed the typhoid toxin. This study demonstrated that strains lacking the typhoid toxin were associated with a more severe disease phenotype[49].

In total, over 400 participants have been successfully challenged to date, with an excellent safety profile. Importantly, the attack rate using the defined dose of $10^4$ CFUs has been consistent throughout all studies. Significant advances have been made in performing these human typhoid and paratyphoid challenge studies, including first confirmation of primary bacteraemia, identification of novel clinical and laboratory biomarkers and progression of our understanding of the host response to infection and microbiological dynamics[50].

##### 1.4.2. Non-typhoidal Salmonella challenge studies

Unlike human host-restricted *S*. Typhi, NTS serovars typically have a broad host range and several animal models have been developed to assess NTS candidate vaccines[51]. *Salmonella* spp. are frequently used as model organisms to study bacterial pathogenesis and host-microbe interactions. Many animal models have been described, including colonisation models; lethality models;

gastroenteritis models and invasive *Salmonella* models[52]. These include immunocompetent mouse models (such as those using NRAMP deficient strains e.g BALB/c or C57BL/6); immunodeficient mouse models; humanised mice; guinea-pig; chicken; rat and rabbit models; streptomycin-treated mouse models; calf ileal loop and rhesus macaque models. Unfortunately, many of these models do not fully recapitulate the manifestations of disease in a clinically relevant context[52,53].

Wild-type NTS challenge for gastroenteritis has been performed in a small number of volunteers. In the earliest published study from 1936, Hormache et al challenged a total of five volunteers with *Salmonella* Typhimurium administered in water, at a dose level of $2 \times 10^9$ (n=3) and $4 \times 10^9$ (n=2). Four out of five developed gastrointestinal disease and the fifth participant developed fever[54]. Other studies have described challenge with *Salmonella* Anatum, *Salmonella* Pullorum and *Salmonella* Meleagridis administered in eggnog[55]. Previous reports have described challenge of healthy volunteers with attenuated *S*. Typhimurium as candidate oral vaccines. Attenuated strains include **Δ*phoP*/Δ*phoQ*** [56] and **Δ*aroC*/Δ*ssaV*** (WT05) variants.[57] Challenge with attenuated *S*. Typhimurium was reportedly safe, immunogenic and – notably – not diarrhoeagenic. However in some instances oral challenge with WT05 was associated with prolonged stool shedding of vaccine lasting up to 21 days[57].

#### 1.5. Clinical Considerations in the development of an iNTS CHIM

##### 1.5.1. Transmission

Transmission of *Salmonella* is predominantly foodborne, from a wide range of different foodstuffs, including red and white meat, eggs, and dairy products. More than half of all *Salmonella* infections are likely to be asymptomatic.[58] Foodborne transmission occurs when cooked food is contaminated by raw food or by failure to reach adequate cooking temperatures. Person-to-person spread has been described, especially when patients are symptomatic with diarrhoea. Spread from asymptomatic food handlers is thought to contribute to only a small proportion of cases[59–65].

The reservoir of ST313 *Salmonella* pathovariants associated with iNTS disease in sub-Saharan Africa (Section 1.5.6.1) is less clearly understood. In studies to date, there is limited overlap between sequence types associated with invasive disease isolated from animal and human sources. [17,66,67] Whilst the data are inconclusive, some studies have suggested that person to person transmission may have an important role in transmission of ST313 *Salmonella* pathovariants associated with iNTS disease.[21,67]

##### 1.5.2. *Salmonella* Gastroenteritis

*Salmonella* gastroenteritis is most frequently caused by Typhimurium ST19 or ST34, and by the 'global epidemic clade' of *S*. Enteritidis.

The clinical features of *Salmonella* gastroenteritis are described in **Section 4.8.3**. Briefly, typical symptoms include watery diarrhoea, bloody diarrhoea, abdominal pain, nausea, vomiting, headache and/or fever. In most instances, stools are loose, of moderate volume and without blood. Other patients may report large volume watery stool or tenesmus. The symptom profile may be impacted by the infectious dose and symptoms may therefore change with dose escalation or de-escalation.[68]

The overall duration of symptoms typically ranges from 4 to 7 days. The total duration of illness is estimated to range between 3 to 19 days.[69]

###### 1.5.2.1. Post-infectious complications of gastrointestinal Salmonella infection

Post infectious complications can on occasion occur following gastrointestinal *Salmonella* infection. These can include erythema nodosum, reactive arthritis and post-infectious irritable bowel syndrome.[61–65,70]

###### 1.5.2.1.1. Reactive arthritis

Reactive arthritis is a subtype of spondyloarthropathy that is characterised by joint pain and swelling that develops soon after or during and infectious process, but in which no organism can be isolated from the affected joints. Gastrointestinal and genitourinary infections are commonly recognised triggers, including with *Salmonella* spp.

A systematic review from 2013 estimated that the incidence of reactive arthritis following *Salmonella* infection was 12 per 1000 patients [71]. This estimate was derived from 16 cohort studies (comprising a mix of hospital-based and population-based studies) totalling 39,148 patients with *Salmonella* infection. The frequency of reactive arthritis after asymptomatic *Salmonella* infection is difficult to accurately ascertain.

Symptom duration is highly variable, but most patients will have little-to-no symptoms at 6-12 months post onset. A small proportion of patients may develop symptoms lasting >12 months.[72] Long term antibiotics do not appear to have any benefit in the management of chronic reactive arthritis.[73–75] Symptom onset is typically 1 to 4 weeks following the preceding infection.[72,76] The prevalence of HLA-B27 appears to be increased in patients with reactive arthritis, although the estimates vary depending on illness definition and study design. [72,77,78] The presence of HLA-B27 is however not essential for the development of reactive arthritis.

Typical symptoms of reactive arthritis are described in **Section 4.8.4**

###### 1.5.2.1.2. Post-infectious irritable bowel syndrome

Post-infectious irritable bowel syndrome is estimated to occur in 3-10% of patients following bacterial diarrhoea. Symptoms generally resolve within 1 year. [70,79,80]

###### 1.5.2.2. Management of *Salmonella* gastroenteritis

###### 1.5.2.2.1. Supportive management

The mainstay of management for all symptomatic patients with *Salmonella* gastroenteritis is replacement of fluids and electrolytes. In general, oral rehydration is preferred using solutions containing water, sugar, and salt.[81] Individuals with severe hypovolaemia should receive intravenous fluid replacement and are typically transitioned to oral rehydration once they are fluid replete.

###### 1.5.2.2.2. Antibiotics

The use of antibiotics in the treatment of *Salmonella* gastroenteritis depends on the individual patient and the severity of disease.

Antibiotic treatment is generally not indicated in healthy, immunocompetent adults with mild-to-moderate disease. This is because gastroenteritis is typically self-limiting without treatment. In addition, antibiotic treatment of *Salmonella* gastroenteritis may also increase the risk of microbiological relapse and prolong the duration of shedding (see **Section 1.5.4.1.1**). [69 82,83] Data from Cochrane collaboration suggest that antibiotic treatment is not associated with a reduced risk of diarrhoea at days 2-4 or 5-7 post illness onset with non-quinolone antibiotics.[69]

Healthy immunocompetent adults are typically treated with antibiotics for *Salmonella* gastroenteritis when there is severe gastro-intestinal disease. Typically, severe disease is classified as gastroenteritis with:

- Severe diarrhoea (e.g >6 stools/days)
- Persistent fever
- A need for hospitalisation

Studies included the meta-analysis suggesting that there was no benefit of antibiotics for *Salmonella* gastroenteritis typically excluded patients with severe disease.[69] Other studies have shown that antibiotic treatment of severe community onset diarrhoea is associated with more rapid resolution of symptoms by 1-2 days, especially when quinolone therapy is used.[84–87] In healthy immunocompetent adults with severe *Salmonella* gastroenteritis, the benefit of shortening symptom duration needs to be balanced against the theoretical risk of prolonged shedding of *Salmonella* in the stool. Antibiotics treatment of severe *Salmonella* gastroenteritis is guided by the antibiotic susceptibilities. Typical treatment durations are between 3 to 7 days.

Antibiotic treatment of *Salmonella* gastroenteritis of any severity may be considered in specific patient groups, including:

- Adults aged >50 years
- Infants <12 months
- HIV positive individuals
- Other immunocompromised states
- Vascular disease, valvular heart disease, joint disease

These groups are at higher risk of complicated or invasive disease. Adults aged over 50 years, particularly those with atherosclerotic disease, are at higher risk of endovascular infection. Pre-emptive treatment of *Salmonella* gastroenteritis is typically recommended in those aged over 50 years, as endovascular infection is estimated to occur in up to 10% of over 50s with blood stream infection.[88,89]

Individuals are considered infectious whilst they are symptomatic. From a public health perspective, individuals are asked to exclude from high-risk activities until they are symptom free for at least 48 hours and have no diarrhoea. Clearance cultures are not typically indicated from a public health perspective, as prolonged shedding can occur.[68]

##### 1.5.3. Invasive *Salmonella* infection

Approximately ~5% of enteric infections with nontyphoidal *Salmonella* are thought to result in bacteraemia. This may increase to ~10% depending on underlying host risk factors and serotype.[90,91] Progression from enteric to systemic infection may occur more frequently in the context of altered gastric acid pH,[92] altered microbiota (including prior antibiotic treatment)[93] and concomitant rotavirus infection.[61–65,94]

Primary NTS bacteraemia typically occurs in the context of significant immunosuppression, and is not associated with gastroenteritis.[95] This likely reflects the majority of iNTS disease occurring in sub-Saharan Africa. *S*. Typhimurium has been shown to be present at only ~1 CFU/ml blood in HIV infected bacteraemic patients.[96] Bacteraemia may occur in the context of gastroenteritis, and may represent 'spill-over' from the gastrointestinal tract. This is typically seen in association with increased age, the presence of underlying co-morbidities and immunosuppression, and infection with specific serotypes.[91,97–100] In particular, infection with *Salmonella enterica* serovars Dublin, Cholerasuis, Newport and Infantis[101] appears to be associated with higher risk of invasive disease. The precise frequency of bacteraemia in gastrointestinal infection is unknown as many primary enteric infections are not microbiologically diagnosed and blood cultures are not routinely performed in all cases. It is plausible that occult bacteraemia may occur in a higher proportion of immunocompetent adults.

Diarrhoea is frequently absent in invasive non-typhoidal *Salmonella* infection. In heavily immunocompromised patients invasive disease can be associated with complicated infection, manifesting with septicaemia, anaemia, pneumonia, meningitis and/or extra-intestinal infections of bones, joints and vascular tissue.[7,61–65]

###### 1.5.3.1. Complications of invasive *Salmonella* infection

Complications of invasive *Salmonella* infection can include septicaemia, hypotension, tachycardia, pneumonia, anaemia, deep-seated infection, gastrointestinal perforation, or haemorrhage. Bloodstream infection can lead to extra-intestinal focal infections at any site, including aortitis, osteomyelitis, meningitis, or arthritis. These complications occur almost exclusively in clinically vulnerable and/or those who do not receive appropriate antibiotic treatment.[61–65]

###### 1.5.3.1.1. Endovascular infection

Endovascular infection is a rare complication of invasive *Salmonella* infection. It is associated with atherosclerotic disease or endovascular prosthesis. It typically affects the infra-renal abdominal aorta but can affect any site of damaged endothelium. Endovascular infection is estimated to occur in up to 10% of over 50s with blood stream infection.[61–65,88,89]

###### 1.5.3.1.2. Meningitis

Meningitis is a rare complication of invasive *Salmonella* infection. It typically occurs in neonates and infants <12years of and in the context of advanced HIV infection.[61–65]

###### 1.5.3.1.3. Osteomyelitis

*Salmonella* osteomyelitis is classically associated with sickle cell disease in children but can also occur in the context of other haemoglobinopathies and other immunosuppressed states. [61–65,102]

###### 1.5.3.2. Management of NTS bacteraemia.

All cases of bloodstream infection require prompt antibiotic therapy. Treatment typically consists of a fluroquinolone or third generation cephalosporin, depending on antibiotic susceptibilities. Treatment duration is typically 10-14 days. Extra-intestinal focal infection or endovascular infections may require longer durations of treatment and additional surgical intervention. [61–65]

##### 1.5.4. Shedding

Convalescent shedding of *Salmonella* is common after symptomatic or asymptomatic NTS infection.[60–65]

###### 1.5.4.1. Acute and convalescent shedding

The precise duration of shedding following symptomatic or asymptomatic NTS infection is poorly understood. Estimates vary depending on study experimental design, frequency of sampling, culture methods and choice of reported endpoints.[103]

One study has reported that 98.8% (254/257) adults infected with NTS had eradicated their infection within 12 days from the first positive stool sample.[104,105]

Shedding may persist up to 5 weeks, with prolonged shedding occurring more frequently in children under 5; in the context of symptomatic infection and in serotypes other that *S.* Typhimurium.[60]

Another retrospective study from Israel suggested that ~2% (1047/48345) of patients with microbiologically confirmed *Salmonella* infection had persistent shedding lasting at least 30 days.[83] Shedding persisted for a median of 55 days, and lasted months-to-years in some isolated cases. Over half of patients with persistent shedding reported a symptomatic disease with relapsing diarrhoea.

###### 1.5.4.1.1. The impact of antibiotics on acute and convalescent shedding

Antibiotic treatment of diarrhoeal disease secondary to NTS does not appear to shorten the duration of shedding and, in some instances, is associated with longer duration of shedding when using non-quinolone antibiotics.[69,83] In one study of children with *Salmonella* gastroenteritis (n=44), treatment with ampicillin (n=15) or amoxicillin (n=15) was associated with bacteriologic relapse in 53% of cases, as compared with 0% of placebo treated controls.[106] Some studies have suggested a similar pattern of prolonged shedding and higher rate of relapse was observed when using fluroquinolones for the treatment of *Salmonella* gastroenteritis in some studies.[107,108] Notably, this was not observed in the Cochrane review of antibiotic treatment for non-typhoidal *Salmonella* gastroenteritis, where fluroquinolone antibiotics were associated with reduced risk of microbiological failure compared with no treatment, (RR 0.33; 95%CI 0.20-0.56). In some instances, antibiotic treatment is associated with prolonged symptoms or higher rate of relapse. Overall antibiotic treatment has been associated with increased risk of Salmonella stool shedding at one month compared with no treatment (RR 1.96; 95% CI 1.29-2.98).[69]

###### 1.5.4.2. Chronic carriage

Chronic carriage is defined as the persistence of *Salmonella* in the stool for >12months from infection. The risk of chronic NTS carriage is lower than that for the typhoidal *Salmonella* serovars. The rate of carriage at 1 year is estimated to be <1%.[60] Chronic carriage is more common with host immunosuppression; in young children <5 and/or or in the context of biliary tract abnormalities.[61–65,109]

##### 1.5.5. Infectious Dose

The precise infectious dose required to induce *Salmonella* gastroenteritis (and/or invasive disease) is unknown.[110] The UK standards for microbiological investigation estimate a typical infectious dose of $10^3$ CFU. Early volunteer studies indicated that high infectious doses ($10^5$-$10^9$ CFU depending on serotype) were required to induce *Salmonella* infection in humans.[54] Outbreak investigations suggest that enteritis can be achieved with infectious doses ranging from $10^1$ to $10^{11}$ depending on the type of food-vehicle, serotype and method for dose calculation.[110] In healthy volunteer studies, administration of sodium bicarbonate prior to challenge with *Salmonella* Typhi, reduces the infectious dose required to cause symptomatic infection, compared with historical controls.[48,111] We anticipate that sodium bicarbonate pre-treatment would be associated with a lower inoculum and more reproducible attack rate following *S*. Typhimurium challenge, similar to *S*. Typhi.

##### 1.5.6. Challenge strain selection

The choice of challenge strain is a key variable in the design of a new challenge model. To maximise the utility of any challenge model, individual wild-type strains need to be i) phylogenetically representative of contemporary disease-causing isolates; ii) susceptible to commonly used antibiotics; iii) have a traceable history from isolation; iv) possess a representative repertoire of virulence factors and [with regards to ST313] v) associated with an invasive disease phenotype.

#### 1.5.6.1. ST313

*S.* typhimurium strains belonging to the lineage ST313 are thought have adapted to an extra-intestinal lifestyle and are responsible for the majority of iNTS cases across sub-Saharan Africa. ST313 consists of three discrete phylogenetic groups, denoted lineage 1, 2 and 3. Strains belonging to lineage 2 are the most frequently isolated from invasive infections in sub-Saharan Africa.

We have manufactured challenge stocks from the ST313 lineage 2 reference strain D23580, which is considered as an archetypal isolate representative of strains causing invasive disease in sub-Saharan Africa. D23580 was originally isolated from a blood culture taken from a 24-month-old child at Queen Elizabeth Central Hospital, Blantyre, Malawi. This strain has been extensively characterised at a genomic, transcriptomic, proteomic and phenotypic level. It has reduced catalase activity, cannot grow on melibiose, L-tartaric acid or dihydroxyacetone as sole carbon sources, and presents a negative red, dry and rough (RDAR) phenotype.[12,112] D23580 possesses five prophages (BTP1, Gifsy-2, ST64B, Gifsy-1, and BTP5), of which Gifsy-2, Gifsy-1, and ST64B are inactivated, and BTP1 is highly spontaneously induced.[113] The strain carries four plasmids, including pSLT-BT (a virulence plasmid pSLT with a Tn21-like element carrying antibiotic resistance genes); pBT1; pBT2 encoding CysS and pBT3.[114]

Several lines of evidence indicate that this strain is adapted to an extra-intestinal lifestyle, including disruption of genes associated with intestinal persistence (PipD, RatB, MacAB)[115–117] and increased dissemination (SseI)[118], alongside transcriptomic changes associated with escape from immune surveillance[15,119] and complement resistance.[114,120] D23580 possess a distinct prophage and plasmid repertoire as compared with ST19, which is – in turn - distinct from, ST313 lineage 1 and lineage 3 isolates.[12] D23580 is sensitive to ciprofloxacin, azithromycin and third generation cephalosporins, ensuring that a range of treatment options are available. The virulence plasmid pSLT encodes resistance to chloramphenicol, ampicillin, and sulphonamides – none of which would be used for treatment. Notably, D23580 is distinct from ST313 strains isolated from the UK and appears to associated with a higher risk of bloodstream infection.[12,121]

#### 1.5.6.2. ST19

Strains belonging to sequence type ST19 are considered as archetypal 'diarrhogenic' strains, although can cause invasive disease in susceptible hosts. We have selected a strain that has undergone detailed in vitro characterisation prior to cGMP manufacture, specifically the 4/74 strain, originally isolated from a calf in England in 1974. Genomic analysis indicates that it is an archetypal ST19 strain, representative of current clinical infections. It is frequently used as a model organism for laboratory infection biology experiments and has undergone detailed genotypic, transcriptomic, proteomic and phenotypic characterisation.[122]

##### 1.6. Rationale for the current study

The overall aim of this project is to establish a controlled human infection model of invasive non-typhoidal *Salmonella* (iNTS) infection. This model will be used to better understand the immunobiology and pathogenesis of iNTS disease in humans, to accelerate the development of vaccines for iNTS. We aim to determine the infectious dose required for 60-75% of volunteers to develop *Salmonellosis*. Our proposal is predicated on the hypothesis that the challenge of healthy volunteers with an invasive strain of *S*. Typhimurium ST313 will be associated with a distinct clinical phenotype as compared with participants challenged with a *S*. Typhimurium ST19 strain. There is a substantial body of phenotypic, genomic, and epidemiological evidence to indicate that specific lineages of *S*. Typhimurium ST313 are adapted to cause systemic illness and are associated with reduced gastrointestinal survival. This model will allow us: to characterise the clinical presentation and microbiological dynamics following NTS challenge with two different *S*. Typhimurium strains; to longitudinally describe the immune response to NTS challenge and to identify new diagnostic biomarkers. Our overarching goal is to use the human infection model to advance the development of the iNTS vaccines that are currently in early-stage testing.

#### 2. Study objectives

The study objectives and endpoints are detailed in **Table 3** .

***Table 3 - Study objectives and endpoints***

| | Objective(s) | Endpoint(s) |
| --- | --- | --- |
| **Primary endpoint** | 1.1. To determine the dose in CFU of *Salmonella enterica* subspecies *enterica* serovar Typhimurium (*S.* Typhimurium) 4/74 and D23580 strains required for 60-75% of volunteers to develop systemic *Salmonellosis* following oral challenge | 1.1. The proportion of participants who develop either:<br>1.1.1. Fever ≥38°C on ≥2 occasions ≥12 hours apart **AND/OR**<br>1.1.2. *Salmonella* Typhimurium bacteraemia |
| **Secondary endpoints** | 2.1 To describe the *Salmonella* colonisation rate following oral challenge with *Salmonella* Typhimurium 4/74 and D23580 strains at different doses | 2.1 The proportion from whom *Salmonella* Typhimurium is isolated from stool on ≥2 occasions ≥48 hours from challenge at different doses of each strain. |
| | 2.2 To describe the *Salmonella* gastroenteritis rate following oral challenge of *Salmonella* Typhimurium 4/74 and D23580 strains at different doses. | 2.2 The proportion of participants at different doses at each strain developing<br>2.2.1 Severe diarrhoea **and/or**<br>2.2.2 Moderate diarrhoea plus<br>2.2.2.1 Fever ≥38°C on ≥1 occasion **and/or**<br>2.2.2.2 ≥1 Grade 2 gastrointestinal symptoms (abdominal pain, nausea, vomiting, tenesmus) |
| | 2.3 To determine the persistent fever rate following oral challenge with *Salmonella* Typhimurium 4/74 and D23580 at different doses. | 2.3 The proportion of participants who develop fever ≥38°C on ≥2 occasions ≥12 hours apart at different doses of each strain. |
| | 2.4 To describe the *Salmonella* bacteraemia rate following oral challenge with *Salmonella* Typhimurium 4/74 and D23580 at different doses. | 2.4 The proportion of participants developing *Salmonella* Typhimurium bacteraemia at any timepoint following challenge at different doses of each strain |
| | 2.5 To describe the rate of systemic *Salmonellosis* according to **an alternative composite diagnostic criterio**n following oral challenge of *Salmonella* Typhimurium 4/74 and D23580 at different doses. | 2.5 The proportion of participants meeting the criteria for a composite diagnosis of *Salmonellosis* at different doses of each strain defined as any of 1) *Salmonella* Typhimurium is isolated from stool on ≥2 occasions ≥48 hours from challenge and/or 2) *Salmonella* gastroenteritis and/or 3) |

| | | |
|---|---|---|
| | | fever ≥38°C on ≥2 occasions ≥12 hours apart and/or 4) *Salmonella* Typhimurium bacteraemia |
| | 2.6 To describe the safety of oral challenge with *S*. Typhimurium 4/74 and D23580 strains | 2.6 The proportion of participants at different doses of each strain reporting<br>• Adverse events,<br>• Adverse events of special interest,<br>• SAEs,<br>• SUSARs.<br>• Concomitant medication usage as outlined in the study protocol |
| | 2.7 To compare **clinical** features following oral challenge with *S.* Typhimurium 4/74 or *S.* Typhimurium D23580 strains | 2.7 A comparison of the clinical features of *Salmonella* infection after challenge with 4/74 or D23580 strains, with specific reference to<br>2.7.1 The proportion of participants in each group developing any diarrhoea.<br>2.7.2 The severity of diarrhoea in each group, as measured by<br>2.7.2.1 Stool volume in ml/24hrs<br>2.7.2.2 Frequency of bowel motions/24hrs<br>2.7.2.3 Total gastrointestinal symptom severity score calculated by summing numerical values assigned to the severity of all solicited gastrointestinal symptoms between Day 0 to Day 14 (0=not present; 1=mild; 2=moderate; 3=severe).<br>2.7.2.4 Total systemic symptom severity score calculated by summing numerical values assigned to the severity of all solicited gastrointestinal and systemic symptoms between Day 0 to Day 14 (0=not present; 1=mild; 2=moderate; 3=severe).<br>2.7.2.5 Area under the curve for gastrointestinal and systemic symptom severity score between Day 0 to Day 14<br>2.7.2.6 Time to honest of diarrhoea in each group, measured in hours since challenge to the first grade 6/7 stool.<br>2.7.3 The proportion of participants in each group developing any fever ≥38°C<br>2.7.4 Fever clearance time of febrile participants in each group measured by time in hours to first temperature <38°C lasting ≥12hrs |

| | | |
|---|---|---|
| | | 2.7.5 The proportion of participants in each group reporting the following solicited symptoms at any time:<br>2.7.5.1 headache,<br>2.7.5.2 malaise<br>2.7.5.3 anorexia<br>2.7.5.4 abdominal pain<br>2.7.5.5 nausea<br>2.7.5.6 vomiting<br>2.7.5.7 dysentery<br>2.7.5.8 myalgia<br>2.7.5.9 arthralgia<br>2.7.5.10 cough<br>2.7.5.11 rash<br>2.7.6 Duration of solicited symptoms measured in days from first symptom onset to complete resolution of symptoms<br>2.7.7 Severity of solicited symptoms measured by<br>2.7.7.1 The proportion of participants with maximum symptom severity score graded as mild, moderate, or severe following challenge<br>2.7.7.2 The proportion of participants meeting the criteria for severe *Salmonellosis*<br>2.7.7.3 Symptom severity scores for individual solicited symptoms calculated by summing numerical values assigned to the severity of individual solicited symptoms between Day 0 to Day 14 (0=not present; 1=mild; 2=moderate; 3=severe |
| | 2.8 To compare **microbiological** features of **gastrointestinal infection** following oral challenge with *S.* Typhimurium 4/74 or *S.* Typhimurium D23580 strains | 2.8 Gastrointestinal *Salmonella* infection after challenge with 4/74 or D23580 strains, with specific reference to<br>2.8.1 Time to onset in days in each group from challenge to first stool sample positive for *Salmonella* Typhimurium by culture and/or PCR<br>2.8.2 Duration of stool shedding in each group measured in days from first stool sample positive for *Salmonella*<br>2.8.3 The duration of stool shedding in each group measured in days from first stool sample positive by culture and/or for *Salmonella* |

| | | |
|---|---|---|
| | | Typhimurium to first persistently negative stool sample (defined as three consecutive samples negative at least 48 hours apart)<br>2.8.4 The magnitude of stool shedding measured in CFU/ml in quantitative stool culture analysis |
| | 2.9 To compare **microbiological** features of **bloodstream infection** following oral challenge with *S.* Typhimurium 4/74 or *S.* Typhimurium D23580 strains | 2.9 *Salmonella* bloodstream infection after challenge with 4/74 or D23580 strains, with specific reference to<br>2.9.1 The proportion of participants in each group developing any bacteraemia<br>2.9.2 The severity of bacteraemia in each group as measured by<br>2.9.2.1 The duration of bacteraemia measured by time in hours from the first positive blood culture to first persistently negative blood culture<br>2.9.2.2 Blood culture clearance time measured by time in hours from initiation of antibiotics treatment to first persistently negative blood culture<br>2.9.2.3 Magnitude of bacteraemia measured in CFU/ml in quantitative blood culture samples collected immediately prior to the initiation of treatment<br>2.9.2.4 The time to onset of bacteraemia in each group, measured by hours since challenge |
| | 2.10To compare biochemical and haematological laboratory parameters following oral challenge with *S.* Typhimurium 4/74 or *S.* Typhimurium D23580 strains | 2.10Absolute values of laboratory parameters from time of challenge to Day 28 and/or Day 90, with specific reference to<br>2.10.1 Total haemoglobin (d/L)<br>2.10.2 Haemoglobin change from baseline (Hb g/l D0 – nadir Hb g/l)<br>2.10.3 Total white cell count (x$10^9$/l)<br>2.10.4 Neutrophil count (x$10^9$/l)<br>2.10.5 Lymphocyte count (x$10^9$/l)<br>2.10.6 Eosinophil count (x$10^9$/l)<br>2.10.7 Monocyte/lymphocyte ratio<br>2.10.8 Urea and electrolytes (Na+,K+, Urea, Creatinine – mmol/l)<br>2.10.9 C-reactive protein (mg/l) |

| | | |
|---|---|---|
| | | 2.10.10 Liver function tests (Bilirubin [umol/l], aspartate transaminase (AST IU/l), alkaline phosphatase (ALP IU/l), alanine transaminase (ALT IU/l), Albumin (g/l) |
| | 2.11 To describe and compare the serum antibody response following oral challenge with *S.* Typhimurium 4/74 or *S.* Typhimurium D23580 strains | 2.11 Absolute values of laboratory assays on serum samples measured at baseline and post challenge time points (e.g. Day 7, 14 , 28, 90 and 365), including but not limited to<br>2.11.1 *S*. Typhimurium O-specific polysaccharide serum IgG and IgA concentration measured by ELISA<br>2.11.2 *S*. Typhimurium specific serum IgG and IgA concentration against other S. Typhimurium antigens measured by ELISA<br>2.11.3 Serum bactericidal antibody titres against *Salmonella* Typhimurium<br>2.11.4 *S*. Typhimurium specific antibody secreting cell and memory B-cell responses measured by ELISPOT<br>2.11.5 Other functional antibody activity measurements including systems serology platforms. |
| | 2.12 To describe and compare the mucosal antibody response following oral challenge with *S.* Typhimurium 4/74 or *S.* Typhimurium D23580 strains. | 2.12 Absolute values of laboratory assays on saliva (and/or stool copro-antibodies) samples measured at baseline and post challenge time points (e.g. Day 7, 14, 28, 90 and 365) using laboratory assays including (but not limited to)<br>2.12.1 S. Typhimurium specific IgG and IgA concentration measured by ELISA<br>2.12.2 Bactericidal antibody titres<br>2.12.3 Other functional antibody activity measurements including systems serology platforms |
| | 2.13 To describe and compare cell-mediated immune response following oral challenge with *S.* Typhimurium 4/74 or *S.* Typhimurium D23580 strains | 2.13 Absolute values and inter-group comparison of laboratory assays performed on peripheral blood mononuclear cells measured at baseline and post challenge time points including (but not limited to)<br>2.13.1 Description of lymphocyte populations at baseline and following challenge as measured by flow cytometry and/or CyTOF<br>2.13.2 Frequency and magnitude of S. Typhimurium specific cell-mediated immune responses as measured by ELISPOT and flow cytometry and/or CyTOF |

| | | |
|---|---|---|
| | 2.14 To describe participant experience following oral challenge with *S.* Typhimurium 4/74 or *S.* Typhimurium D23580 strains | 2.14 Descriptive statistics following administration of a structured questionnaire at baseline and post-challenge |
| **Tertiary** | 3.1. To investigate how the human microbiota interacts with oral challenge of *S.* Typhimurium 4/74 or *S.* Typhimurium D23580 strains | 3.1 Metagenomic analysis of stol and/or saliva to measure constituent microbiological flora (and their metabolome/transcriptome) at baseline and post-challenge time points |
| | 3.2. To investigate novel diagnostic methods for detecting *S.* Typhimurium infection | 3.2 Exploratory analysis of blood and faecal samples, including novel molecular techniques |
| | 3.3. To describe and compare additional host immune responses following oral challenge of *S.* Typhimurium 4/74 or *S.* Typhimurium D23580 strains. | 3.3 Analysis of B-cell repetoir, plasma cytokine profile and kinetics, additional exploratory immunology |
| | 3.4. To evaluate environmental contamination with *Salmonella* enterica in the near patient environment | 3.4 The pattern and proportion of environmental samples testing positive for *Salmonella* Typhimurium by culture or molecular methods |
| | 3.5. To evaluate host-pathogen interactions following *Salmonella* infection including the presence and mechanisms of *Salmonella* persister infected macrophages | 3.5 Exploratory analysis of infected immune cells from blood including, but not limited to, single-cell RNA-seq. |

### 3. Study design

#### 3.1. Study design

This study is a phase 1, first-in-human, dose-escalation, randomised controlled human infection model of *Salmonella* Typhimurium infection using the continual reassessment method (CRM). Participants will be randomised (when possible) and challenged with one of two strains of S. Typhimurium (4/74 or D23580). Participants will be followed up for a period of 12 months. Study participants will be healthy volunteers aged 18 to 50. A full list of inclusion and exclusion criteria is provided in **Section 4 Participant entry.**

A flowchart of the study design for dose escalation of each challenge strain is presented in Figure 2 .

The parameters for the CRM are listed below:

- Four doses for each challenge agent: 1-5x$10^3$ CFU, 1-5x$10^4$ CFU, 1-5x$10^5$ CFU, 1-5x$10^6$ CFU;
- Target attack rate: 67.5%;
- Dose-attack rate model: one-parameter logistic model;
- Dose-attack rate skeleton: 60%, 75%, 85%, 90%;
- Inference: Bayesian;
- Decision rule: the dose closest to target attach rate (absolute difference);
- Sample size: 40 per each challenge agent;
- Cohort size=5;
- Safety modifications: starting at the lowest dose and only one dose escalation at a time;
- Stopping rule: 20 participants challenged at the recommended dose;

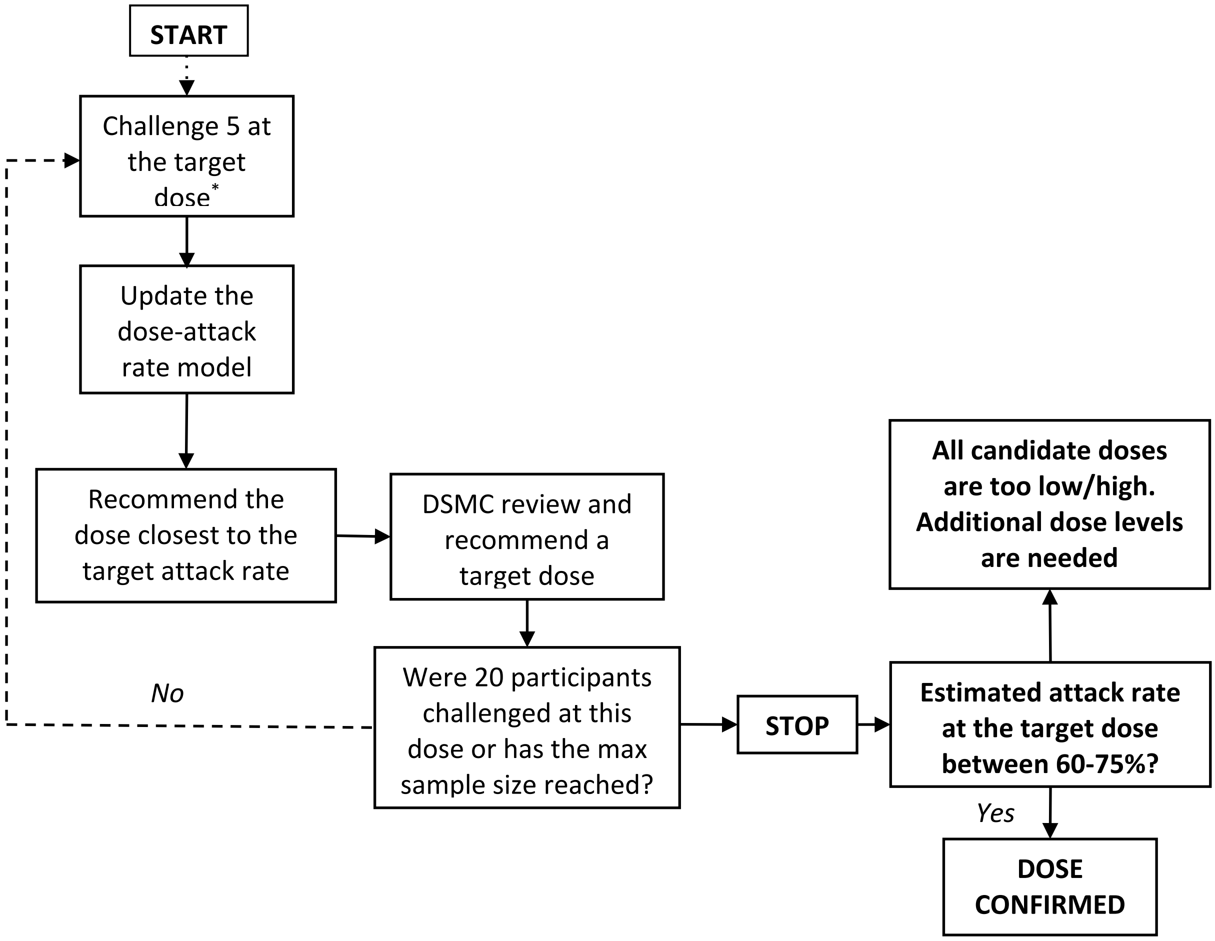

***Figure 2 - Decision making algorithm for dose escalation/de-escalation.***
***Starting at 1-5 x$10^3$ to reach the primary endpoint *The target dose for the first cohort will be 1-5 x $10^3$ CFU, and the first cohort of 5 participants will be divided into 3 groups (1, 2, and 2 participants) and challenge with an interval of*** **at least 7 days.** ***For the rest of the cohorts, they will be challenged on the same day, if possible.***

A sentinel cohort of ten participants will randomised and challenged (2,4,4) at a starting dose of $10^3$ CFU (for both strains) administered with sodium bicarbonate and monitored for 14 days. We will collect daily blood and stool cultures and regular safety bloods (See **Section 3.4** and **Table 8**). Participants will be treated if they meet pre-specified criteria outlined in **Section 6.8.3**. The primary disease endpoint will be *Salmonellosis*, as defined in **Section 6.8.1**. Safety data will be submitted to the DSMB to provide a go-no-go decision for further challenge.

Dose escalation for both strains will happen in parallel. Participants will be randomised to receive one of the two challenge agents at their specific target dose until the target dose is establish for one agent. From that point, the remaining participants will be challenged with the other agent. We aim to recruit between 40-80 participants in total for the two challenge agents (20-40 for each challenge agent). Details of the simulation results based on difference scenarios are provided in **section 9.1,** when comparing with the rule-based study design. The simulation results have clearly show that the model based have a higher chance to find the correct dose in different scenarios.

The challenge dose will be defined as one that achieves a 60-75% attack rate. The DSMC will be provided with an updated report after each cohort of 5 (per strain) completes challenged.

#### 3.2. Study outcome measures

The primary, secondary and exploratory study endpoints and outcome measures are detailed in **Table 3, Table 4** and **Table 5.**

##### 3.2.1. Primary endpoint

The primary endpoint is clinical or microbiological features of systemic Salmonellosis, defined in (**Table 4**)**.** Dose escalation and de-escalation decisions will be made based on the primary endpoint in consultation with the data safety monitoring committee.

***Table 4 – Primary endpoints for the diagnosis of Salmonella infection.***

| **Systemic Salmonellosis will be diagnosed if either of the following criteria are met** |
|---|
| • Fever ≥38°C on ≥2 occasions ≥12 hours apart |
| • *Salmonella* Typhimurium bacteraemia at any time point |

##### 3.2.2. Secondary disease endpoints

Exposure to *Salmonella* Typhimurium can result in a spectrum of outcomes. These range from asymptomatic colonisation through to bacteraemia. The secondary disease endpoints of most relevance to future model development are defined in **Table 5**. They are arranged in descending order of probability and ascending order of severity. Additional secondary endpoints are detailed in Table 3.

***Table 5 – Secondary endpoints.***

***1 -Severe diarrhoea is defined as ≥6 or ≥800g liquid stool samples (Grade 6-7 on Bristol stool scale) or ≥2 episodes of gross blood in stool (dysentery) in a rolling 24 hour period; 2 – Moderate diarrhoea is defined as 4-5 or 400-800g liquid stool samples (Grade 6-7 on Bristol stool scale) in a rolling 24 hour period; 3 – Grade 2 symptoms are self-defined as those causing discomfort enough to cause some limitation of usual activity and/or some medical intervention/therapy is required.***

| |
|---|
| 2.1 - *Salmonella* Colonisation - Isolation of *Salmonella* Typhimurium from stool on ≥2 occasions ≥48 hours from challenge **and/or** |

| |
|---|
| 2.2 – *Salmonella* gastroenteritis<br>• Severe diarrhoea[1] **and/or**<br>• Moderate diarrhoea[2] and<br>○ fever ≥38°C on one occasion OR<br>○ ≥1 Grade 2[3] systemic symptoms |
| 2.3 Fever<br>• Fever ≥38°C on ≥2 occasions ≥12 hours apart |
| 2.4 *Salmonella* bacteraemia alone<br>• *Salmonella* Typhimurium bacteraemia with or without fever |

###### 3.2.3. Severe Salmonellosis

Severe *Salmonellosis* will be defined as illness that includes any of the criteria satisfied in **Table 6**.

***Table 6 - Criteria for severe Salmonella diagnosis***

| **Severe *Salmonellosis* is diagnosed if ANY of the following apply** |
|---|
| Oral temperature ≥ 40.0°C |
| Systolic blood pressure ≤ 85 mmHg |
| Significant lethargy or confusion (GCS <12) |
| Gastrointestinal bleeding |
| Gastrointestinal perforation |
| Extra-intestinal seeding of *Salmonella* Typhimurium[1] |
| Any grade 4 or above laboratory abnormality |

##### 3.3. Definitions

- Diarrhoea – The passage of three or more loose or liquid stools per day. [2]
- Loose/Liquid stool – Stool score of 6 or 7 on the Bristol stool scale[3] [126]

  a. Type 6 stool – Fluffy pieces with ragged edges, a mushy stool
  b. Type 7 stool – Watery, no solid pieces, entirely liquid

[1] Including Salmonella osteomyelitis, arthritis, arteritis or other deep seated infection

[2] Frequent passage of formed stools is not considered as diarrhoea.

[3] The Bristol stool chart is frequently used graded visual scale of stool density. It is validated as a proxy for gastrointestinal transit time. Type 6-7 stool is used to define *Clostridioides difficile* diarrhoea in the most recent ESCMID guidance.[123] Type 6-7 stool are analogous to types 3-5 stool grading used in earlier diarrhoea challenge studies [124,125], where stools were graded on a five-point scale (grade 1 [fully formed], grade 2 [soft], grade 3 [thick liquid stool], grade 4 [opaque-watery], grade 5 [rice-water stool]).

- Diarrhoea severity[4]
    a. Mild
        i. 3 loose/liquid stools in a 24-hour period[5] or
        ii. 200-399g in any 24-hour period,
    b. Moderate
        i. 4–5 diarrheal stools in 24 hours or
        ii. 400–800 g within 24 hours or
    c. Severe
        i. ≥6 loose/liquid stools in 24 hours or
        ii. >800 g of loose/liquid stools in 24 hours.
        iii. ≥2 stools with gross blood in 24 hours

- Dysentery - ≥2 episodes of gross blood in a loose stool.

- Time to onset of symptoms – Time (Hours & Days) to first recorded solicited symptoms in the diary card OR first temperature ≥38°C

- Duration of illness – Time (Hours & Days) from first recorded individual solicited symptoms to complete resolution of individual recorded symptoms in the diary card.

- Fever clearance time – Time (Hours & Days) from first dose of treatment until temperature ≤37.5°C for a 48 hour period.

- Symptom severity
    a. The proportion of participants with maximum symptom severity score graded as mild, moderate or severe following challenge. See Appendix.
    b. The proportion of participants meeting the criteria for severe *Salmonellosis*
    c. The proportion of participants recording one or more severe solicited symptoms following challenge.
    d. Total symptom scores calculated by summing numerical values assigned to the severity of all solicited symptoms between Day 0 to Day 14 (0=not present; 1=mild; 2=moderate; 3=severe).
    e. Individual symptom severity scores calculated by summing numerical values assigned to the severity of individual solicited symptoms between Day 0 to Day 14 (0=not present; 1=mild; 2=moderate; 3=severe).

- Time to diagnosis – Time (Hours/Days) from challenge until fulfilment of diagnostic criteria (taken as date/time Gram negative rods are detected in blood culture AND/OR recorded temperature ≥38°C for 12hours)

---

[4] Diarrhoea severity grading is adapted from that used in a *Shigella* human challenge study endpoints [127] [124]

[5] This refers to a rolling 24-hour period that starts with the passing of each loose stool.

- Time to onset of bacteraemia – Time (Hours/Days) from challenge until the date/time first positive blood culture collected.

- Duration of bacteraemia – Time (Hours/Days) from collection of first positive blood culture until date/time of the first negative blood culture and blood cultures are persistently negative.

  Quantification of bacteraemia at time of diagnosis – Concentration of bacteria (CFU/ml) in 10ml blood taken at the time of diagnosis.

- Time to onset of stool shedding - Time from challenge (Hours/Days) to the first positive stool culture.

- Time to onset of diarrhoea - Time from challenge (Hours/Days) to the first grade 6/7 stool.

- Duration of stool shedding – Cumulative number of days where positive stool samples test positive for *Salmonella* Typhimurium by culture and/or PCR

- Pattern of stool shedding – Descriptive

- Haematological and biochemical parameters
  - Total Haemoglobin (g/L)
  - Haemoglobin change from baseline (Hb g/l D0 – Hb g/l D14)
  - Total White Cell Count ($x10^9/l$)
  - Platelet counts ($x10^9/l$)
  - Neutrophil count ($x10^9/l$)
  - Lymphocyte count ($x10^9/l$)
  - Monocyte count ($x10^9/l$)
  - Eosinophil count ($x10^9/l$)
  - Monocyte/Lymphocyte ratio
  - Urea & Electrolytes (Na, K+, Urea, Creatinine –mmol/l)
  - C-reactive protein (mg/l)
  - Liver function tests (Bilirubin [umol/l], aspartate transaminase (AST IU/l), alkaline phosphatase (ALP IU/l), alanine transaminase (ALT IU/l), Albumin (g/L)

#### 3.4. Visit Structure

##### 3.4.1. Primary challenge

Participants will be required to attend the study site for a maximum of 26 visits including the screening visit. The timing of visits and procedures carried out on each visit are described in **Table** 7. The protocol for visits will depend on whether the participant requires early treatment (prior to day 14) and/or meets the criteria for primary endpoint for *Salmonella* diagnosis, ('SD') defined in **Table 4**. This means that an individual participant may have fewer visits in total, depending on the day of diagnosis. In addition, further reviews may be required depending on the participant's condition, clinical indication or at the discretion of the study staff.

###### 3.4.2. Study visit window periods

Every effort will be made to perform the visits on time as indicated in the study protocol. Based on previous experience with challenge studies and similar trial protocols, we are aware that in some instances this may be difficult. A window of time to complete the visits will be as follows:

- Day -7 (-7 days/+3days (i.e. between Day -14 and Day -4)
- Day 28 +/- 4 days
- Day 90 +/- 14 days
- Day 180 +/- 28 days
- Day 365 +/- 56 days

*Table 7 – Summary of study procedures for patients undergoing primary challenge.*

| | Pre-challenge | | Challenge period | | | | | | | | | | | | | | | | | | | | Follow up | | | |
|---|---|---|---|---|---|---|---|---|---|---|---|---|---|---|---|---|---|---|---|---|---|---|---|---|---|---|
| | | | Inpatient quarantine | | | | | | | | Outpatient assessments | | | | | | | Post diagnosis/treatment | | | | | | | | |
| | Screening | Day -7 | Day 0 | Day 1 | Day 2 | Day 3 | Day 4 | Day 5 | Day 6 | Day 7 | Day 8 | Day 9 | Day 10 | Day 11 | Day 12 | Day 13 | Day 14 | SD | SD +24 h | SD +48 h | SD +72 h | SD +96 h | Day 28 | Day 90 | Day 180 | Day 365 |
| **Informed consent** | X | | | | | | | | | | | | | | | | | | | | | | | | | |
| **Consent quiz** | X | | | | | | | | | | | | | | | | | | | | | | | | | |
| **Medical history**[6] | X | X | | | | | | | | | | | | | | | | | | | | | | | | |
| **Vital signs**[7] | x | X | x | x | x | x | x | x | x | x | x | x | x | x | x | x | x | x | x | x | x | x | | | | |
| **Physical examination**[8] | x | X | | | | | | | | | | | | | | | | X | x | x | x | x | | | | |
| **Urine pregnancy test** | x | | x | | | | | | | | | | | | | | | x | | | | | | | | |
| **Urine dipstick** | X | | | | | | | | | | | | | | | | | | | | | | | | | |
| **SARS-CoV-2 Lateral flow** | | | x | | | | | | | | | | | | | | | | | | | | | | | |
| **12 lead ECG** | X | | | | | | | | | | | | | | | | | | | | | | | | | |
| **Ultrasound scan** | x | | | | | | | | | | | | | | | | | | | | | | | | | |
| **Mood assessment questionnaire**[9] | x | x | | | | | | | | x | | | | | | | | | | | | | | | | |
| **Irritable bowel syndrome questionnaire** | x | | | | | | | | | | | | | | | | | | | | | | x | x | x | |
| **Revalidation of consent** | | | x | | | | | | | | | | | | | | | | | | | | | | | |
| **Obtain 24 hr contact details** | | x | x | | | | | | | | | | | | | | | | | | | | | | | |
| **Issue study pack** | | x | | | | | | | | | | | | | | | | | | | | | | | | |
| **Admission to quarantine** | | | x | | | | | | | | | | | | | | | | | | | | | | | |
| **Discharge from quarantine**[10] | | | | | | | | | | x | | | | | | | | | | | | | | | | |
| **Adverse event recording** | | x | x | x | x | x | x | x | x | x | x | x | x | x | x | x | x | x | x | x | x | x | x | x | x | x |
| **Blood sample** | x | x | x | x | x | x | x | x | x | x | x | x | x | x | x | x | x | x | x | x | x | X | x | x | x | x |
| **Stool sample**[11] | | x | x | x | x | x | x | x | x | x | x | x | x | x | x | x | x | x | x | x | X | x | x | x | x | x |
| **Environmental sample**[12] | | | x | x | x | x | x | x | x | x | | | | | | | | | | | | | | | | |
| **Saliva sample** | | | x | | | | | | | | | | | | | | x | x | | | | | x | x | | |
| **Challenge** | | | x | | | | | | | | | | | | | | | | | | | | | | | |
| **Notification of UKHSA, HPU and GP**[13] | | | X | | | | | | | | | | | | | | x | | | | | | x | | | |
| **Letter and PIS for close contacts provided in discharge pack**[8] | | | | | | | | | | x | | | | | | | | | | | | | | | | |
| **Participant questionnaire** | | | | | | | | | | | | | | | | | | | | | | | x | | | |
| **Antibiotics** | | | | | | | | | | | | | | | | | x[14] | x | | | | | | | | |

6 Any interim medical history after day-7 will be recorded as an adverse event.

7 Vital signs may be taken and recorded at other times in the study if clinically indicated and at the discretion of the study team.

8 Performed at Screening, D-7, D0 and SD.Physical examination may be performed at other times in the study if clinically indicated and at the discretion of the study team.

9 Day 7 OR upon discharge from quarantine, if different to Day 7, and additional times if clinically indicated.

10 If participant meets discharge criteria.

11 Stool samples will be collected at least one week after completion of a 14 day course of antibiotics, until 3 successive stool samples are negative for Salmonella spp. If persistent stool shedding occurs after completion of antibiotics, participants will be referred to the Infectious Diseases Consultant at Imperial Healthcare NHS Trust.

[12] Surface swabs and contact plates in participant rooms for a randomly selected subset of participants (not all participants) and selected days during the 7 day quarantine at the discretion of the study team.
[13] UKHSA, the local health protection units and participant GPs will also be notified at the time of stool shedding clearance
[14] Antibiotic treatment may be started at any time post challenge as outlined in section 6.9.

| | Pre-challenge | | Challenge period | | | | | | | | | | | | | | | | | | | | Follow up | | | |
|---|---|---|---|---|---|---|---|---|---|---|---|---|---|---|---|---|---|---|---|---|---|---|---|---|---|---|
| | | | Inpatient quarantine | | | | | | | | Outpatient assessments | | | | | | | Post diagnosis/treatment | | | | | | | | |
| | Screening | Day -7 | Day 0 | Day 1 | Day 2 | Day 3 | Day 4 | Day 5 | Day 6 | Day 7 | Day 8 | Day 9 | Day 10 | Day 11 | Day 12 | Day 13 | Day 14 | SD | SD +24h | SD +48h | SD +72h | SD +96h | Day 28 | Day 90 | Day 180 | Day 365 |
| **Stool sample** | | x | x | x | x | x | x | x | x | x | x | x | x | x | x | x | x | x | x | x | x | x | x | x | x | x |
| **Blood culture BACTEC** | | | 10 | 10 | 10 | 10 | 10 | 10 | 10 | 10 | 10 | 10 | 10 | 10 | 10 | 10 | 10 | 10 | 10 | 10 | 10 | 10 | | | | |
| **FBC** | 1 | 1 | 1 | | 1 | | 1 | | 1 | | 1 | | 1 | | 1 | | 1 | 1 | 1 | 1 | 1 | 1 | 1 | 1 | 1 | |
| **U&E/CRP/LFT** | 2 | 2 | 2 | | 2 | | 2 | | 2 | | 2 | | 2 | | 2 | | 2 | 2 | 2 | 2 | 2 | 2 | 2 | 2 | 2 | |
| **Additional screening investigations**[15] | 10 | | | | | | | | | | | | | | | | | | | | | | | | | |
| **Bacterial quantification** | | | | | | | | | | | | | | | | | | 10 | | | | | | | | |
| **Serum sample** | | | 10 | | | | | | | 10 | | | | | | | 10 | 10 | | | | | 10 | 10 | 10 | 10 |
| **Plasma Sample** | | | 3 | | | | | | | 3 | | | | | | | 3 | 3 | | | | | 3 | | | |
| **PBMC** | | | 25 | | | | 25 | | | 25 | | | | | | | 25 | 25 | | | | 10 | 25 | 25 | 25 | 25 |
| **Functional genomics** | | | 3 | 3 | 3 | 3 | | | | 3 | | | | | | | | 3 | | | | | 3 | 3 | | |
| **Whole blood PCR** | | | 10 | 10 | 10 | 10 | 10 | 10 | 10 | 10 | | | | | | | | | | | | | | | | |
| **DNA sample (including epigenetics)** | | | x | | | | | | | | | | | | | | | | | | | | x | | x | |
| **Saliva sample** | | | x | | | | | | | x | | | | | | | x | | | | | | x | | x | |
| **Total blood volume** 708ml | | | | | | | | | | | | | | | | | | | | | | | | | | |

***Table 8 - Clinical sample collection plan for patients undergoing primary challenge. Values correspond to volume of blood collected in millilitres.***

[15] HLA-B27, Coeliac screen, HIV/HepB/HCV serology, haemoglobinopathy screen, HbA1c

### 4. Participant entry

#### 4.1. Pre-registration evaluations

All participants will attend a screening visit prior to enrolment in the study. Screening procedures are outlined in **Section 6.3.3**. Prior to enrolment participants will undergo the following investigations:

- Full blood count
- Urea and electrolytes
- Liver function tests
- C-reactive protein
- Serum IgA
- Coeliac serology
- HLA-B27 screening
- Coagulation screen
- Haemoglobinopathy screen
- HIV 1&2 antibody
- Hepatitis B surface antigen
- Hepatitis C IgG
- HbA1c
- Ultrasound of the biliary tract
- Ultrasound of the abdominal aorta.
- Stool culture and PCR for *Salmonella* spp.[1]
- Malaria screen (for participants who have travelled to malaria endemic areas within the preceding 6 months from the date of screening).
- Urine pregnancy test
- Urine dipstick test
- ECG
- Resting heart rate, respiratory rate, blood pressure, oxygen saturations and oral temperature.

#### 4.2. Inclusion criteria

Participants must satisfy all the following criteria to be considered eligible for the study:

- Agree to give informed consent for participation in the study.
- Aged between 18 and 50 years inclusive at time of challenge.
- In good health as determined by medical history, physical examination, and clinical judgment of the study team.
- Agree (in the study team's opinion) to comply with all study requirements, including capacity to adhere to good personal hygiene and infection control precautions.
- Agree to allow his or her General Practitioner (and/or Consultant if appropriate), to be notified of participation in the study.

[1] Testing will be performed at Day-7 visit and will comprise a multiplex PCR to assess for *Salmonella* spp., *Shigella* spp., *Campylobacter* spp., E. Coli O157, *Giardia* spp and *Cryptosporidum* spp.

- Agree to allow study staff to contact his or her GP to access the participant's medical history and vaccination records.
- Agree to allow UKHSA and the local health protection unit to be informed of their participation in the study.
- Agree to give his or her close contacts written information informing them of the participant's involvement in the study and offer them voluntary screening for *Salmonella* carriage.
- Agree to a period of inpatient quarantine whilst symptomatic (In exceptional circumstances, the duration of inpatient quarantine may be extended.)
- Agree to have 24-hour contact with study staff during the four weeks post challenge and can ensure that they are contactable by mobile phone for the duration of the study period until antibiotic completion.
- Agree to allow the study team to hold the name and 24-hour contact number of a close friend, relative or housemate who will be kept informed of the study participant's whereabouts for the duration of the challenge period after quarantine (from the time of challenge until completion of antibiotic course). This person will be contacted if study staff are unable to contact the participant after discharge from inpatient quarantine.
- Have internet access to be contactable by the study team by email for real-time safety monitoring.
- Agree to avoid antipyretic/anti-inflammatory treatment from the time of challenge (Day 0) until advised by a study doctor or until 14 days after challenge.
- Agree to refrain from donating blood for the duration of the study.
- Agree to provide their National Insurance/Passport number for the purposes of Trial over-volunteering prevention system (TOPS) registration and for payment of reimbursement expenses.
- Participants must have received at least two doses of a SARS-CoV-2 vaccines that has been approved for use by the MHRA (or other national regulatory authority) ≥ four weeks prior to enrolment.
- Proficient in English at a level sufficient to understand, retain, weigh up and communicate the study details as outlined in the participant information sheet, in the opinion of the study investigator.

##### 4.3. Exclusion criteria

The participant will not be enrolled if any of the following apply:

- History of microbiologically confirmed *Salmonella* infection.[1]
- History of **significant**[2] organ-specific and/or systemic disease that could interfere with trial conduct or completion. Including, for example, but not restricted to:

[1] Including enteric fever (*Salmonella* Typhi and/or *Salmonella* Paratyphi infection)

[2] Individuals with specific medical co-morbidities (e.g. childhood asthma) will be considered for enrolment after review with the principle investigator and clinical oversight group, following discussion with their GP and/or consultant as appropriate. The rationale for enrolment in these cases will be recorded in the CRF.

- Cardiovascular disease, including specifically
  - Atherosclerotic disease
  - Stable or unstable angina
  - Previous myocardial infarction
- Valvular heart disease
- Vascular disease, including specifically
  - Documented aneurysmal arterial disease
  - Peripheral arterial disease
  - Endovascular prosthesis
- Respiratory disease[1]
- Haematological disease including sickle cell disease and sickle cell trait.
- Endocrine disorders, including specifically
  - Diabetes mellitus
- Renal or bladder disease, including history of renal calculi
- Biliary tract disease, including specifically
  - biliary colic,
  - asymptomatic gallstones, or
  - previous cholecystectomy
- Gastro-intestinal disease including specifically,
  - a current requirement for antacids, $H_2$-receptor antagonists, proton pump inhibitors or laxatives
  - Inflammatory bowel disease
  - Confirmed diagnosis of irritable bowel syndrome as defined by the Rome IV criteria.[128]
- Neurological disease
- Metabolic disease
- Autoimmune disease
- Psychiatric illness requiring hospitalisation
- Known or suspected drug and/or alcohol misuse disorder
- Chronic/Active Infectious disease including active tuberculosis

[1] Participants with well controlled asthma may be considered for enrolment at the discretion of the study investigators and following consultation with their general practitioner

- Severe infection requiring hospitalisation for intravenous antibiotics within the last 10 years. Exceptions to this would include a short course of intravenous antibiotics for appendicitis, biliary sepsis, diverticulitis, and cellulitis.
- History of recent malaria infection within the past 12 months from screening.[1]
- History of joint replacement
- History of any orthopaedic/osseous implanted prosthesis
- Presence of other internal implanted device e.g. permanent pacemaker

- Have any known or suspected impairment of immune function (as defined in the green book[129]), alteration of immune function, or prior immune exposure that may alter immune function to *Salmonella* infection resulting from, for example:
  - Congenital or acquired immunodeficiency, including IgA deficiency
  - Human Immunodeficiency Virus infection or symptoms/signs suggestive of an HIV-associated condition
  - Evidence of severe primary immunodeficiency, for example, severe combined immunodeficiency, Wiskott-Aldrich syndrome, and other combined immunodeficiency syndromes.
  - Currently being treated for malignant disease with immunosuppressive chemotherapy or radiotherapy, or who have received such treatment within at least the last six months.
  - Individuals who have received a solid organ transplant and are currently on immunosuppressive treatment
  - Individuals who have received a bone marrow transplant, until within 12 months of finishing all immunosuppressive treatment,
  - Individuals receiving systemic high-dose steroids[2] until at least three months after treatment has stopped.
  - Individuals receiving other types of immunosuppressive[3] drugs (alone or in combination with lower doses of steroids) until at least six months after terminating such treatment.
  - Receipt of immunoglobulin or any blood product transfusion within 3 months of study start.
  - History of cancer (except squamous cell or basal cell carcinoma of the skin and cervical carcinoma in situ).
- Moderate or severe depression or anxiety as classified by the Hospital Anxiety and Depression Score at screening or challenge that is deemed clinically significant by the study doctors[4].

---

1 Volunteers with distant history of malaria infection may be considered for enrolment if they have been treated and can demonstrate a negative malaria test at screening.

2 Defined as those receiving at least 40mg of prednisolone per day for more than one week or equivalent

3 e.g. azathioprine, cyclosporin, methotrexate, cyclophosphamide, leflunomide and other cytokine inhibitors.

4 If elevated scores are due to temporary significant life events, the questionnaire may be repeated after resolution of the event with a view to inclusion if normal.

- Weight less than 50kg[1].
- Anyone taking long-term medication (e.g. analgesia, anti-inflammatories or antibiotics) that may affect symptom reporting or interpretation of the study results.
- Contraindication to cephalosporin, fluroquinolone, or macrolide antibiotics.
- Female participants who are pregnant, lactating or who are unwilling to ensure that they or their partner use effective contraception 30 days prior to challenge and continue to do so until two negative stool samples, a minimum of 3 weeks after completion of antibiotic treatment, have been obtained.
- Full-time, part-time, or voluntary occupations involving:
    - Clinical healthcare work[2]
    - Social work with direct contact with young children (defined as those attending pre-school groups or nursery or aged under 2 years), or other clinically vulnerable children, adolescents
    - Clinical or social work with direct contact with highly susceptible patients or persons in whom *Salmonella* infection would have particularly serious consequences (unless willing to avoid work until demonstrated not to be infected with *Salmonella* in accordance with guidance from Public Health England and willing to allow study staff to inform their employer).
- Full time, part time or voluntary occupations involving:
    - Commercial food handling (involving preparing or serving unwrapped foods not subjected to further heating)
- Close household contact with:
    - Young children (defined as those attending pre-school groups, nursery or those aged less than 2 years)
    - Any individual who is immunocompromised.
    - Pregnant women
    - Household contacts aged over 70 years
- Scheduled elective surgery or other procedures requiring general anaesthesia during the study period.
- Participants who have participated in another research study involving an investigational product that might affect risk of *Salmonella* infection or compromise the integrity of the

[1] Or a Body Mass Index (BMI) that, in the opinion of the study doctors, may adversely impair the interpretation of the study results or affect the safe performance of any study procedures.
[2] This includes medical, dental, nursing or midwifery students (or other allied-healthcare professionals) with direct patient contact. Such students who are not undertaking patient-facing rotations until demonstrated not to be infected with *Salmonella* species may be considered for enrolment.

study within the 30 days prior to enrolment (e.g. significant volumes of blood already taken in previous study)[1].

- Detection of any abnormal results from screening investigations, unless deemed not clinically significant.
- Any other social, psychological or health issues which, in the opinion of the study staff, may
    - Put the participant or their contacts at risk because of participation in the study,
    - Adversely affect the interpretation of the primary endpoint data,
    - Impair the participant's ability to participate in the study.
- Having previously received any experimental *Salmonella* vaccine as part of a clinical trial
- Have participated in previous *Salmonella* Typhi or Paratyphi challenge studies (with ingestion of challenge agent).
- Have a prolonged corrected QT interval (>450 milliseconds) on ECG screening
- Screening blood test positive for HLA-B27.
- Any employee of the sponsor or research site personnel directly affiliated with this study or their immediate family members.
- Inability to comply with any of the study requirements (at the discretion of the study staff).

##### 4.4. Temporary exclusion criteria at challenge

Participants will be temporarily excluded from challenge if presenting at the challenge visit with the following:

- Positive lateral flow test for SARS-CoV-2
- Significant acute or acute-on-chronic infection within the previous 7 days or have experienced fever (>37.5°C) or subjective febrile symptoms within the previous 3 days.
- History of any antibiotic therapy during the previous 5 days.
- Any systemic corticosteroid (or equivalent) treatment in the previous 14 days, or for more than seven consecutive days within the past 3 months.
- Therapy with antacids, proton pump inhibitors or $H_2$-receptor antagonists within 24 hours prior to challenge.
- Detection of gastrointestinal pathogens (other than *Salmonella*) in stool culture/PCR collected at the pre-challenge assessment (Day-7) including *Shigella* spp., *Campylobacter* spp., *E. Coli* O157, *Giardia* spp and *Cryptosporidum* spp, until two subsequent samples are negative. Detection of *Salmonella* spp. in stool prior to challenge (Day – 7) will result in exclusion from the study.

---

[1] As assessed by both participant questioning and registration with The Over Volunteering Prevention System (TOPS) database.

##### 4.5. Pregnancy and contraception

The possible adverse effects of *S.* Typhimurium infection or the effect of some antibiotics on the outcome of pregnancy are unknown. Therefore, pregnant women will be excluded from the study. If relevant, female participants will be required to use an effective form of contraception from 30 days prior to challenge until deemed to be clear of infection with three negative clearance stools.

*Salmonella* infection, with or without diarrhoea or vomiting, could reduce the efficacy of an oral hormonal contraceptive by altering absorption. For this reason, female participants who are taking oral contraception will be advised to use additional barrier contraception during the two-week challenge period and whilst taking antibiotics.

In the event of a female participant becoming pregnant during the study this outcome will be recorded and the Sponsor and the DSMC will be notified if appropriate. No further non-essential trial procedures will be performed (i.e. appropriate antibiotic treatment, screening for stool carriage and/or referral may be required but not further blood sampling etc.).

##### 4.6. Trial over-volunteering prevention system

The Over-volunteering Prevention System (TOPS) is a database to guard against the potential for harm that can result from excessive volunteering in clinical trials involving IMPs or blood donations. Participants will be registered with this system at screening using their national insurance number or passport number if they do not have a national insurance number. The system will be updated in the event of the participant not entering the trial, being withdrawn, or excluded. Alternatively, TOPS will be updated on the participant's last visit.

##### 4.7. Potential benefits to study participants

Participants will not directly benefit from participation in this study. However, it is hoped that the information gained from this study will contribute to the development of an understanding of *Salmonella* infection. The only benefits for participants will be information about their general health status.

##### 4.8. Potential risk to study participants

The general risks to participants in this study are associated with study-fatigue, phlebotomy, symptomatic infection, and the small risk of subsequent complications. In this section we outline potential risks to study participants and the relevant risk mitigation strategies that we will employ.

During the challenge phase (before treatment with antibiotics) participants will be admitted to an inpatient quarantine facility. They will be reviewed at least daily by a clinical study team member and be under observation 24/7. Participants will be made aware of all potential risks, including the potential symptoms of *Salmonella* infection. They will be monitored closely throughout the challenge for the development of these symptoms. Symptoms of fever, headache, malaise, anorexia, abdominal pain, nausea/vomiting, myalgias and arthralgias, cough, rash, diarrhoea, and constipation will be actively enquired about at each formal review. Participants will be instructed to record their oral temperature twice a day with a provided thermometer should they feel feverish.

The medical management of *Salmonella* infection is outlined below. If any participant is unexpectedly unwell, an additional formal review will be arranged by the clinical study team. If it is deemed necessary for safety reasons after the quarantine period has completed, a study nurse or doctor will be permitted to review the participant in their home.

##### 4.8.1. Study-fatigue

This may occur due to the intense nature of the study procedures, especially during the challenge period. Participants are compensated for their time and every effort will be made to make study investigations as swift and uncomplicated as possible.

###### 4.8.1.1. Risk mitigation – Study fatigue

Interventions and visits will be limited in number and will be arranged to fit with individual schedules and other obligations, as far as is practical.

##### 4.8.2. Phlebotomy

The volume of blood drawn over the study period should not compromise healthy adult participants. Risks from venepuncture or cannulation include mild tenderness, bruising, light-headedness and, rarely, syncope or arterial puncture.

###### 4.8.2.1. Risk mitigation - Phlebotomy

No more than 673ml of blood will be taken over the course of the study (**Table 8**). As a comparison, women can donate a maximum of 1410mls of blood per year, and men 1880mls, to the National Transfusion Service. At enrolment, any recent blood donations will be checked to ensure that the total volume of blood taken is safe to give. Venous cannulation can be considered to avoid repeated venepuncture, pain, and bruising, although samples blood cultures would need to be taken from a fresh puncture on each day to avoid contamination.

##### 4.8.3. Symptomatic gastroenteritis

We anticipate that some study participants will develop symptomatic *Salmonella* gastroenteritis infection following challenge. In real-world settings, more than half of all *Salmonella* infections are estimated to be asymptomatic.[58] One aim of this study is to determine the proportion of participants developing symptomatic infection at different challenge doses.

###### 4.8.3.1. Incubation period

The incubation period for Salmonella gastroenteritis is typically 12-48 hours but ranges from as short as 4 hours to a long as 120 hours. The incubation period may be impacted by the infectious dose and may therefore change with dose escalation or de-escalation.[68]

###### 4.8.3.2. Symptoms of Salmonella gastroenteritis

Typical symptoms include watery diarrhoea, bloody diarrhoea, abdominal pain, nausea, vomiting, headache and/or fever. In most instances, stools are loose, of moderate volume and without blood. Other patients may report large volume watery stool or tenesmus. The symptom profile may be impacted by the infectious dose and symptoms may therefore change with dose escalation or de-escalation.[68]

The overall duration of symptoms typically ranges from 4 to 7 days. Other studies from the Cochrane collaboration suggest that that the mean duration of diarrhoea is between 3 to 13 days. The mean duration of fever is estimated to be 1-2 days. The total duration of illness is estimated to range between 3 to 19 days.[69]

###### 4.8.3.3. Risk mitigation – *Salmonella* gastroenteritis

During the challenge phase participants will be admitted to an inpatient quarantine facility, such that any symptoms suggestive of severe diarrhoea or dehydration will be detected early. They will be reviewed at least daily by a clinical study team member and be under observation 24/7, to assess specifically for signs of dehydration. Daily stool output monitoring and daily stool cultures will be performed (See Table 8). Severe diarrhoea will be an indication for antibiotic treatment (See Table 10). Oral rehydration will be provided for patients with diarrhoea. Individuals with severe hypovolaemia will receive intravenous fluid replacement and are typically transitioned to oral rehydration once they are fluid replete.

##### 4.8.4. Invasive *Salmonella* infection

A proportion of participants may develop *Salmonella* bloodstream infection. The complications of invasive *Salmonella* infection can include septicaemia, hypotension, tachycardia, pneumonia, anaemia, deep-seated infection, gastrointestinal perforation, or haemorrhage.

###### 4.8.4.1. Risk mitigation – invasive *Salmonella* infection

Invasive Salmonella infection and associated complications occur almost exclusively in clinically vulnerable patients and/or those who do not receive appropriate antibiotic treatment. Participants at high risk of invasive disease and complications will be excluded from participation in the study (See **Section 4.3 and 6.3.3**). During the challenge phase participants will be admitted to an inpatient quarantine facility, such that any symptoms suggestive of invasive will be detected early. They will be reviewed at least daily by a clinical study team member and be under observation 24/7. Daily blood cultures will be collected to identify bacteraemia, were this to occur. Detection of bacteraemia would prompt early treatment with antibiotics (See Table 10).

##### 4.8.5. Reactive arthritis

Post-infectious reactive arthritis is described in **Section 1.5.2.1.1.**

In those patients who do develop reactive arthritis, typical symptoms are of mono-arthritis or oligo-arthritis, often affecting the lower limbs. Axial symptoms, enthesistis and/or dactylitis can also occur. Symptom duration is highly variable, but most patients will have little-to-no symptoms at 6-12 months post onset. A small proportion of patients may develop symptoms lasting >12 months.[72]

###### 4.8.5.1. Risk mitigation – Reactive arthritis

The overall risk of post-infectious reactive arthritis is low. HLA-B27 screening will be performed prior to enrolment (See **Section 4.1**). Any patient who develops symptoms suggestive of reactive arthritis will be referred to a rheumatology consultant at the Imperial Healthcare Hospitals NHS Trust for further investigation and management.

##### 4.8.6. Post-infectious irritable bowel syndrome

There is a theoretical risk of inducing post-infectious irritable bowel syndrome (PI-IBS) following *Salmonella* gastroenteritis. Post-infectious irritable bowel syndrome is estimated to occur in 3-10% of patients following bacterial diarrhoea.

###### 4.8.6.1. Risk mitigation – Post infectious irritable bowel syndrome

To mitigate this, and to estimate the frequency of this occurrence, we will administer an IBS questionnaire at baseline and post challenge as outlined in **Table 7**. In participants who do develop PI-IBS, symptoms generally self-resolve within 1 year. [70,79,80]

Participants will complete the Rome IV questionnaire[130] and the irritable bowel severity scoring system, described by Francis and colleagues [131] at baseline. This questionnaire has recently been used to describe the characteristics and frequency of post-infectious irritable bowel syndrome following *Campylobacter* enteritis [132]. Participants with baseline symptoms of irritable bowel syndrome will be excluded from participation.

##### 4.8.7. Relapse of Salmonella infection

Most immunocompetent patients will make a full recovery from *Salmonella* gastroenteritis. A small proportion develop microbiological relapse and prolonged shedding, which may be exacerbated by antibiotic treatment.

Relapses of *Salmonellosis* may develop post treatment in immunocompromised patients. This is particularly well described in patients with advanced HIV infection[133], chronic granulomatous disease[134], defects in specific cytokine pathways[4,8] and/or haematological malignancies[135].

###### 4.8.7.1. Risk mitigation – Relapse of *Salmonella* infection

In this instance, the risk of relapse will be mitigated primarily by exclusion of participants at risk (**Section 4.3**). Participants will be screened for shedding of *S*. Typhimurium in the stool commencing two weeks after completion of antibiotic therapy and reviewed if symptoms or laboratory results suggest evidence of relapse.

##### 4.8.8. Chronic carrier state

Long-term carriage of non-typhoidal *Salmonella* is rare. The median duration of nontyphoidal *Salmonella* shedding is estimated to be 5 weeks, with persistent excretion >1 years occurring in <1% patients.[60] One retrospective study from Israel suggested that ~2% (1047/48345) of patients with microbiologically confirmed *Salmonella* infection had persistent shedding lasting at least 30 days.[83] Shedding persisted for a median of 55 days, and lasted months-to-years in some isolated cases. Over half of patients with persistent shedding reported a symptomatic disease with relapsing diarrhoea.

###### 4.8.8.1. Risk mitigation -Chronic carrier state

To ensure clearance of infection and to exclude chronic carriage, stool samples for culture will be obtained upon completion of the initial antibiotic course. Should chronic carriage occur (defined as stool cultures being positive for *S.* Typhimurium four weeks after completion of antibiotics) then participants will be referred to an Infectious Diseases Consultant at the Imperial Healthcare NHS Trust for further management.

##### 4.8.9. Antibiotics

Treatment may be initiated with a fluroquinolone (e.g. ciprofloxacin), azithromycin or a third generation cephalosporins (see **Section 6.8.3**). Potential participants with may develop an intolerance or allergy to antibiotics used in the study. Antibiotic intolerance may manifest as gastro-intestinal upset, nausea, vomiting or other unspecified symptoms. An allergic reaction may present with rash, angio-oedema, difficulty in breathing or, rarely, anaphylaxis.

Azithromycin and quinolone antibiotics may be associated with a prolonged QT interval.

All antibiotics may be associated with abnormal liver function tests or other biochemical abnormalities.

Antibiotic treatment may – in theory – increase the risk of carriage of drug resistant bacteria.

###### 4.8.9.1. Risk mitigation – antibiotics

Antibiotic treatment will be avoided in specific instances where the risks of adverse events from treatment outweigh the benefits (see section 6.9).

The antibiotics to be used in this study are generally well tolerated and are only occasionally associated with side effects. Antibiotic treatment duration will be as short as possible (See **Section 6.9.1**). Participants with known antibiotic hypersensitivity or allergy to either of the first-line antibiotics (ciprofloxacin, azithromycin, or other macrolide antibiotics and cephalosporins) will be excluded (See **Section 4.3**).

Should an antibiotic cause allergy or intolerance this will be managed by a study doctor and a different antibiotic will be used for subsequent management. Participants will be admitted to an inpatient quarantine facility for the duration of the study and any adverse reactions will be noted promptly and managed by appropriately trained staff.

All patients will have a baseline ECG with QTc measurements taken at baseline. Participants with prolonged QTc interval will be excluded (See **Section 4.3**).

Regular measurements of liver function tests and electrolyte monitoring will take place during the study (See **Section 6).**

##### 4.9. Potential risk to close contacts of study participants

Person-to-person spread has been described, especially when patients are symptomatic with diarrhoea. Spread from asymptomatic food handlers is thought to contribute to only a small proportion of cases.[59,60 61–65]

Individuals are considered infectious whilst they are symptomatic with diarrhoea. From a public health perspective, individuals are asked to exclude from high-risk activities until they are symptom free for at least 48 hours and have no diarrhoea. Clearance cultures are not typically indicated from a public health perspective, as prolonged shedding can occur.[68]

It is acknowledged, however, that transmission within households can occur if the individual excreting *S.* Typhimurium fails to practice effective hand washing after defecation and is subsequently involved in uncooked food preparation. If food is kept at ambient temperatures, bacterial proliferation occurs such that an infective dose level is reached, and the food then may act as a vehicle for *Salmonella* transmission.

###### 4.9.1. Risk mitigation – risk to close contacts of study participants

In view of the low infectivity of *S.* Typhimurium without bicarbonate buffer and the high standard of hygiene and sanitation in the UK, secondary transmission of the challenge strain to household or other close contacts after discharge is considered highly unlikely.

Throughout the period of possible excretion of the challenge strain, participants must practice stringent hand washing techniques after defecation. Participants will be given soap and paper towels for use at home and detailed advice on how to prevent transmission. Participants will be taught and observed practising good hygiene technique at their initial challenge visit. The importance of adhering to sanitation advice will be emphasised to participants at each visit. It is important to note that participants in this trial will be fully informed about the risks of transmission and how to prevent this prior to challenge. As such, participants will be in the position to implement this from the point of infection which will reduce the chance of secondary transmission.

When occasional transmission of *Salmonella* infection occurs, it is usually related to unknowingly infected food handlers. For this reason, food handlers will be excluded from this study. Potential participants employed in clinical or social work with direct contact with young children (those attending pre-school groups, nursery, or aged less than 2 years) or highly susceptible patients or persons in whom *Salmonella* infection would have particularly serious consequences (such as the elderly) also represent an increased risk and will be excluded unless willing to not work until it has been demonstrated that they are not infected with *S.* Typhimurium (See **Section 4.3**).

##### 4.10. Withdrawal criteria

Each participant can exercise their right to withdraw from the study at any time. If, however, the participant decides to withdraw after they are challenged, they may require additional follow up for public health reasons. In some instances, antibiotic treatment may be required and participants may be required to attend additional hospital/non-study visits to ensure compliance.

In addition, the investigator may terminate a participant's involvement in the study at any time if the investigator considers it necessary for any reason including, though not exclusive to, the following:

- Ineligibility (either arising during the study or in the form of new information not declared or detected at screening),
- Significant protocol deviation,
- Significant non-compliance with study requirements or risk to public health,
- Any adverse event which requires discontinuation of the study procedures or results in an inability to continue to comply with study procedures,
- Lost to follow up.

Withdrawal from the study will not result in exclusion of the data generated by that participant from analysis. The reason for withdrawal, if given, will be recorded in the CRF.

#### 5. Interventions

##### 5.1. *S.* Typhimurium challenge strains

We have selected two bacterial challenge stocks that will be used in the clinical study. The rationale for the selection of these specific challenge strains is detailed in **Section 1.5.6**. One strain (4/74) belongs to the ST19 lineage, whereas the second strain (D23580) belongs to the ST313 lineage. We hypothesise the challenge of healthy volunteers with *S.* Typhimurium ST313 (D23580) will be associated with a distinct clinical phenotype as compared with participants challenged with a *S.* Typhimurium ST19 (4/74) strain. There is a substantial body of phenotypic, genomic, and epidemiological evidence to support this, but has yet to be proven clinically. [12,115–117,121]

We have manufactured two challenge strains to a GMP standard, equivalent to that used for IMP production. Manufacture was undertaken at an accredited facility (Pilot Bioproduction Facility [PBF], Walter Reed Army Institute of Research, MD, USA [WRAIR]). Both strains have a detailed record of isolation and storage, and have been deposited in the UK National Collection of Type Cultures (NCTC).[136] The 4/74 (NCTC14672) strain was originally isolated from a calf in England in 1974 and was linked to an outbreak of gastroenteritis. It has been sourced from the University of Liverpool. The D23580 (NCTC14677) strain was originally isolated from a blood culture taken from a 24-month-old child at Queen Elizabeth Central Hospital, Blantyre, Malawi. It was supplied to the University of Liverpool from the Malawi-Liverpool-Wellcome unit and was deposited in the NCTC under the terms of the MTA. Vials of both strains were supplied by the University of Liverpool to the PBF at WRAIR, who undertook manufacture to GMP standard of both strains. Batch production records have been produced for both strains. Microbial limits testing was performed to confirm absence of other specified pathogenic bacteria, namely *Escherichia coli*, *Pseudomonas spp.*, *Bacillus cereus*, *Staphylococcus aureus*, *Salmonella spp*. (other than *S.* Typhimurium), yeasts and moulds and <100 non-pathogenic bacteria and <20 fungi/ ml of the strain material. The culture media and selective media used are outlined in the batch production record. Microbial limits testing was performed by WRAIR. Full antibiotic sensitivity and microbial purity of both strains in the master cell banks was demonstrated and further characterisation work including genome sequencing has been completed at the University of Liverpool. Samples will be taken at defined time points to evaluate the stability of the frozen GMP manufactured product. Stability testing will be performed by WRAIR, in accordance to established SOPs.

###### 5.1.1. Storage

The S. Typhimurium (4/74 and D23580 strains) for challenge of participants are stored as a frozen suspension in soya tryptone medium containing 10% sucrose. Suspensions have been labelled with the contents (S. Typhimurium 4/74 or S. Typhimurium D23580), date of manufacture, CFU information, storage conditions, batch and vial number.

Vials of the required concentration of S. Typhimurium will be thawed and diluted immediately prior to use. The actual challenge dose administered will be back calculated after each administration. Details of sample storage and accountability are outlined in the Laboratory Analysis plan and in local SOPs.

##### 5.2. Accountability for the challenge strain

The investigator will be responsible for adequate and accurate accounting of *S.* Typhimurium vials prepared for administration to participants. The investigator or designee will administer the study *S.* Typhimurium vials only to individuals included in this study following the procedures set out in this study protocol and the associated local SOPs and Study Plans. The date, dosage and time of administration will be recorded.

The study team will track all vials of *S.* Typhimurium that have been used, administered to participants, and wasted within an accountability log.

##### 5.3. Compliance with Trial Treatment

The challenge agents will be administered by trained doctors and nurses. Administration will be documented according to GCP guidelines and relevant local SOPs. Issues related to compliance are therefore the responsibility of the study staff that have received appropriate training.

##### 5.4. Concomitant Medication

Any medication taken by the participant at the time of enrolment into the study or during the first 90 days of the study period will be recorded on the eCRF. Study medications as outlined in **Table 11** will be provided by study staff according to clinical indication.

##### 5.5. Post-trial Treatment

Study medication will not be continued beyond the study period. Unused medications prescribed during the study will be returned to the study team for disposal 28 days after NTS challenge.

#### 6. Study procedures

##### 6.1. Recruitment

###### 6.1.1. Identification of study participants

Several strategies may be employed to recruit the required cohort of participants, including:

- Direct mail out NHS Database: Potential study participants will be identified via National Health Applications and Infrastructure Services (NHAIS) who hold the central NHS patient database (NHS Digital) or their equivalent. Use of the database will be approved by the Caldecott Guardian for that specific area. These databases will identify all persons within the local area who are in the appropriate age range. First contact to potential participants will not be made by the researchers at Imperial College London. The initial information about the study will be sent out from this agency to preserve the confidentiality of potential participants. Potentially eligible participants will be sent an invitation letter, which describes the study, with a reply slip. Anyone who is interested in taking part will be able to contact the study team by telephone, email, registering on the study website (https://www.imperial.ac.uk/infectious-disease/research/human-challenge/chants) or by returning the attached reply slips.

- Poster advertising: Display of posters advertising the study throughout local hospitals and doctor's surgeries, tertiary education institutions and other public places with the permission of the owner/ proprietor.
- Direct mail-out: This will involve direct mailing of the study information sheet to adults whose names and addresses have been obtained via the Electoral register. Those people who have indicated they do not wish to receive postal mailshots will have their names removed prior to the study team being given the names and addresses. The company providing this service is registered under UK GDPR.
- E-mail communication: We will contact representatives of local tertiary education establishments and local employers and ask them to circulate posters and information, and to circulate a link to study information on the study website by email.
- NIHR Imperial Clinical Research Facility healthy volunteer database - Direct email and link to members of the public who have registered their interest in potentially volunteering for clinical trials conducted by the ICRF. This secure database is maintained by the NIHR ICRF and members of the public registered here have given consent to have their details recorded and be contacted expressly for this purpose of being notified when a trial opens for recruitment. They understand this is not a commitment to volunteering for any trial they are contacted about.
- Media advertising: Local media, newspaper, website, and social media advertisements placed in locations relevant for the target age group with brief details of the study and contact details for further information.

- Website advertising: Description of the study and copy of information booklet on the study website.
- Exhibitions: Advertising material and/or persons providing information relating to the study will exhibit using stalls or stands at exhibitions and/or fairs, such as University Fresher's Fairs.
- Royal Mail Leaflet: Royal Mail door-to-door service with delivery of invitation letters in Imperial College envelopes to every household within certain postcode areas.

#### 6.2. Pre-screening

Volunteers who are interested in study participation will be able to contact the study team by telephone, email, by visiting the study website online registration or by returning a paper reply slip.

##### 6.2.1. Initial eligibility assessment of potential study participants

Once an expression of interest has been received by the study team, potential volunteers will be asked to complete a pre-screening questionnaire. Volunteers will be directed to the study website to complete an online pre-screening questionnaire and submit their contact details. Alternatively, study investigators can complete the pre-screening questionnaire and collected contact details by telephone.

Volunteers who successfully complete the pre-screening questionnaire will be sent an information booklet by email or post to read at their leisure. Participants can also be directed to the study website, where the information booklet will be available.

If participants are willing to proceed, they are invited for a screening and consent visit, where a member of the clinical research team will assess their eligibility.

#### 6.3. Screening visit

##### 6.3.1. Consultation and review of study procedures

This screening will begin with a consultation with a study investigator. This acts as an opportunity for the study investigator to explain the study procedures in detail and supplements the information provided in the participant information sheet. Volunteers will be expected to have read the PIS in detail before the screening visit and will have an opportunity to ask questions directly to study investigators.

If, after this consultation, volunteers are still interested in participating, they will be asked to sign a consent form.

##### 6.3.2. Informed Consent

The participant will personally sign and date the latest approved version of the informed consent form before any study specific procedures are performed. Consent will be sought as described in relevant local SOPs and the clinical study plan.

Written and verbal versions of the participant information booklet and informed consent form will be presented to the participant, detailing no less than:

- the exact nature of and the rationale for performing the study
- implications and constraints of the protocol
- The risks and benefits involved in taking part (as outlined in **Sections 4.7** and **4.8**)
- To contact their GP (or other clinician involved in their long-term care) and/or access a shared electronic NHS care record to confirm their medical and immunisation history and participation in the study.
- To inform close contacts of their involvement in the study by giving them letters provided by the study team. The letter details the study and offers screening for *Salmonella* carriage. The low risk of spread will be emphasised to contacts to avoid undue anxiety.
- For a study staff member to hold the name and 24-hour contact number of a close friend, relative or housemate who lives nearby and will be kept informed of the study participant's whereabouts after the study. This person is to be contacted if study staff are unable to contact the participant for clinical follow up. The 24-hour contact will receive written information, and complete and sign a reply slip that the participant will give the study doctor/nurse before challenge.
- For UKHSA and the local health protection unit to be informed of study participation, challenge outcome, commencement and completion of antibiotic course, and clearance results.
- If the participant is involved in the provision of health or social care to vulnerable groups, then consent will be taken to inform his/her employer of their participation in the study.

It will be clearly stated that the participant is free to withdraw from the study at any time, for any reason and that they are under no obligation to give the reason for withdrawal. The participant will be allowed adequate time to consider the information, and the opportunity to question the researcher, their GP, or other independent parties to decide whether they will participate in the study. Written informed consent will be obtained by means of a dated signature of the participant and a signature of the study staff member who presented informed consent. A copy of the signed informed consent will be given to the participant and the original signed form will be retained at the study site. A doctor or nurse, who has been trained in the consent process, will conduct the informed consent discussion.

Participants will be asked to complete a consent quiz after completing the informed consent form to ensure they have properly understood the study.

###### 6.3.3. Eligibility Assessment

Once informed written consent is obtained, the following baseline assessments and information will be collected as part of the assessment of inclusion/exclusion criteria:

- Participant demographics; age, sex, ethnicity, country of birth
- Medical history to reflect study inclusion and exclusion criteria (**Sections 4.2 and 4.3**), with reference to:

- Details of any significant medical or surgical history based on participant recall and supplemented by GP records and/or by accessing a shared electronic NHS care record. If medical clarification is required, medical notes may be obtained and/or discussion with other medical practitioners will be undertaken.
- Confirmation from the participant's GP that they know of, or do not know of, any medical condition that may affect their suitability as a participant
- History of past blood donation history and plans for future donations.
- Immunisation history (particularly receipt of previous oral typhoid vaccines)
- Contraception use (Female participants only)

- Use of concomitant medication including
    - over the counter medications,
    - vitamin supplements,
    - recreational drug use
    - herbal supplements.
- Physical examination including as a minimum:
    - Cardiovascular examination,
    - Respiratory examination,
    - Abdominal examination,
    - Gross neurological examination,
    - Calculation of Body Mass Index.
- Investigations, including:
    - Blood samples for:
        - Full blood count
        - Urea and electrolytes
        - Liver function tests
        - C-reactive protein
        - Serum IgA
        - Coeliac serology
        - HLA-B27 screening
        - Coagulation screen
        - Haemoglobinopathy screen
        - HIV 1&2 antibody
        - Hepatitis B surface antigen
        - Hepatitis C IgG
        - HbA1c
        - Malaria screen (for participants who have travelled to malaria endemic areas within the preceding 6 months from the date of screening).
    - Urine dipstick (and laboratory analysis if appropriate)

- Urine pregnancy test (for female participants only)
- 12-lead ECG
- Abdominal ultrasound (to screen for gallbladder disease and abdominal aortic aneurysm).
- Mood assessment by the Hospital Anxiety and Depression Score.

- Responses regarding any personal or domestic reason that may lead to concern regarding a participant's ability to maintain good personal hygiene.
- Provision of the following documents.
    - 24-hour contact letter (to be returned completed and signed),
    - Letter to participant's close contacts.

The medical history, vaccination history and prescribed medication lists are based primarily on participant recall. The participants' GP will be contacted to confirm the history by providing a copy of the participant's medical summary and stating whether they know of any medical reason why the participant should not be included. Alternatively, a summary care record can be accessed through a shared electronic NHS care record where available. Review of the medical summary with no concerns is required prior to study enrolment.

Consent will be taken to register the participant onto TOPS (see **Section 4.6**)

Participants will be informed that they would also be eligible for the Imperial College Healthcare Tissue bank.[137] Separate consent is sought for this.

##### 6.3.4. Incidental medical findings

All laboratory results will be reviewed and collated by the study team who will record these in the electronic source database. If a test result is deemed clinically significant, it may be repeated, to ensure it is not a single occurrence. If a test remains clinically significant, the participant will be informed, and appropriate medical care arranged with the permission of the participant in liaison with their GP. Decisions to exclude potential participants from enrolling in the trial or to withdraw a participant from the trial will be at the discretion of the Chief and Co-Investigators.

##### 6.3.5. Eligible participants

After all screening procedures are completed, the study investigators will review all results. An eligibility checklist will be documented in the CRF by two separate study investigators.

If eligibility checklist is satisfactory, a volunteer will be deemed eligible to participate. Volunteers will at that point be invited to attend a pre-challenge visit, where they will be enrolled in the study.

###### 6.3.5.1. Eligible but declined to participate

Some individuals may be deemed eligible to participate after screening, but subsequently decline to participate. For the purposes of CONSORT reporting, these participants will be designated as 'Declined to Participate'. This will be recorded in the CRF for the purposes of maintaining a screening log.

###### 6.3.6. Ineligible participants

After screening, some volunteers may be deemed ineligible to participate if they meet any of the study exclusion criteria or if screening investigations are abnormal. Decisions to exclude potential participants from enrolling in the trial or to withdraw a participant from the trial will be at the discretion of the Chief and Co-Investigators. Participants will be informed of the reason for screening failure and if any further action is required. They will be reimbursed for the screening visit and investigations as outlined below. For the purposes of CONSORT reporting, these participants will be designated as screening failures. The reason for screening failure will be documented in the CRF for the purposes of maintaining a screening log.

##### 6.4. Pre-challenge visit

Volunteers who are deemed eligible for challenge after screening will be asked to attend a study visit prior to challenge. After eligibility is confirmed, the study team will contact the patient by telephone or email to inform them of their eligibility. The study team will arrange a proposed date for challenge to take place. Volunteers will be invited to attend a study visit one to two weeks (Day -14 to Day -4) prior to challenge (designated as the "Day – 7 visit"), where they will be formally enrolled in the study.

The primary purpose of this visit is to prepare participants for the challenge period. Participants will be asked to sign a continued consent form to confirm that they are still willing to participate. We will repeat a medical history and examination to ensure that there have been no changes to participant health since the screening visit. Study procedures are outlined in **Table** 7 and sample collection is outlined in **Table 8**.

###### 6.4.1. Definition of Enrolment

A participant is considered enrolled at the time of the Day -7 visit. A maximum period of 120 days between initial screening and enrolment into the study will be accepted. If the period between screening and enrolment exceeds 120 days, rescreening will consist of repeat informed consent, interim medical history, clinical examination, urinalysis, and blood tests as a minimum. Additional screening investigations, including repeat ultrasound gallbladder scan, may be performed at the discretion of the study doctor following consultation with the senior study investigator.

###### 6.4.2. Stool collection kit

Participants will be asked to provide a stool sample prior to challenge. The purpose of this is to ensure that participants are not colonised with *Salmonella* spp. or other enteric pathogens prior to challenge.

To facilitate sample collections, participants will be issued with a stool collection kit at the Day-7 visit. This stool collection kit includes an opaque transport bag, cool packs, stool pots, transport tubes, toilet seat stool collection paper and stool collection instructions.

Participants will be asked to return the stool sample at the earliest opportunity and no later than 72 hours prior to challenge.

The stool sample will be processed for

- Stool culture and PCR for *Salmonella* spp. and other enteric pathogens.

The result of the stool sample will be recorded in the CRF.

###### 6.4.2.1. Temporary exclusion due to positive stool samples

The result of the stool sample collected at Day-7 will be reviewed prior to the challenge visit. Detection of gastrointestinal pathogens (other than *Salmonella*) in stool culture/PCR collected at the pre-challenge assessment (Day-7) will result in temporary exclusion from the study. This included detection of the following organisms:

- *Shigella* spp.,
- *Campylobacter* spp.,
- *E. Coli* O157,
- *Giardia* spp
- *Cryptosporidum* spp,
- 

Participants will be managed according to the local SOP for management of incidental medical findings. They may subsequently be eligible for challenge at the discretion of the chief investigator if they are able to provide two subsequent samples, 48 hours apart, that test negative. These samples would be collected at *ad hoc* study visits arranged directly with study participants.

Detection of *Salmonella* spp. in stool prior to challenge (Day – 7) will result in exclusion from the study.

###### 6.4.3. Diary

At the Day – 7 visits, participants will be provided with a diary card. They will be instructed in how to complete the diary card and when to report to the study team. Participants will be asked to record solicited symptoms from challenge through to 21 days. The diary card will be reviewed daily during the inpatient stay and during the daily visits from day 8 to day 14. Participants may be contacted outside of clinic visits by telephone or email to confirm that the diary card has been completed and to review solicited and unsolicited symptoms. Details from the diary card will be uploaded into the eCRF. Unsolicited symptoms will be recorded from challenge up to one year post challenge.

###### 6.4.4. Randomisation and blinding

Randomisation of challenge agent will be carried out on or after Day -7. Randomisation will use random block sizes of two or four with an allocation ratio of 1:1 to receive the ST19 or ST313 strains.

The exception will be the three groups in the sentinel cohort, where the block sizes will be two, four and four, respectively.

###### 6.4.4.1. Challenge agent blinding

The study will be conducted double-blind from the time of randomisation until participant unblinding, which will occur once the last participant has completed their Day 28 post-challenge visit. The randomisation schedules will be prepared by the trial statistician. Trained unblinded laboratory staff will prepare challenge agents on Day 0, according to the randomisation at Day-7 and labelled the agent with participant identification number, in accordance with local SOPs. The participants, clinical staff administering the challenge agents and those undertaking follow-up visits will be blinded to the challenge agent and dose (ST19 or ST313 strains). Both strains will be prepared suspended in 0.9% saline and will have an indistinguishable appearance (transparent, colourless liquid).

Aside from designated unblended laboratory staff preparing the challenge agents, other laboratory staff members will remain unaware of challenge agent assignment while conducting the immunogenicity assays. Participants will be informed by the study team of the challenge agent they have received at the time of unblinding. A letter will also be sent to the participant's GP notifying them of unblinding and challenge agent assignment.

Unblinding may occur at an earlier time point in the event of participant withdrawal, or the occurrence of SAEs, SARs or SUSARs. This will be conducted under the guidance of the Data Safety and Monitoring Committee.

#### 6.5. Day 0 – S. Typhimurium challenge

Challenge with either wild type *S.* Typhimurium ST19 (4/74) or *S.* Typhimurium ST313 (D23580) will take place on Day 0. These procedures are described below, and in further detail in the relevant SOPs, Clinical and Laboratory Study Plans.

##### 6.5.1. Pre-challenge assessment

- Check observations and perform physical examination if clinically indicated; complete continued consent form; check eligibility criteria (specifically for temporary exclusion criteria to challenge) and check for occurrence of any new adverse or medically significant events since previous visit.

- Check details of the 24-hour contact (who will be kept informed by the participant of their whereabouts for the subsequent 28 days) (see **Section 6.3**)

- Record oral temperature, pulse, and blood pressure.

- Perform urinary pregnancy test for all females.

- Perform a SARS-CoV-2 lateral flow test.

- Collect clinical specimens as per Table 8

- Confirm the participant has fasted for a minimum of 90 minutes prior to ingestion of the challenge agent.

##### 6.5.2. Preparation of challenge agent

The solution for ingestion containing either wild type *S.* Typhimurium 4/74 (ST19) strain or *S.* Typhimurium D23580 (ST313) will be prepared in a Class II biological safety cabinet within containment level 3 laboratory that is solely used for the purpose of preparing the challenge solution. Challenge solution preparation is conducted by laboratory staff and checked by two laboratory staff members. Two clinical study team members check the challenge solution immediately prior to ingestion by the participant. The water, 0.9% saline and bicarbonate used for the preparation will be commercially available food products. The strain will be prepared, checked, and given to participants as outlined in relevant SOPs and Laboratory and Clinical Study Plans. Following solution ingestion, the single-use containers will be returned to the laboratory for inspection, autoclaving, and disposal.

##### 6.5.3. Administration of S. Typhimurium 4/74 (ST19) and D23580 (ST313)

Both *S.* Typhimurium challenge agents used in this study will be administered by the oral route with sodium bicarbonate at a starting dose of 1-5x$10^3$ CFU. Participants will fast for 90 minutes before and after challenge. The procedure for administration is:

- Remove the *S.* Typhimurium suspension from the BIOJAR.
- Check label to ensure contents of solution match challenge agent allocation.
- Ask the participants to drink the 120 ml of bicarbonate solution.
- Ask the participants to ingest the 30ml *S.* Typhimurium 0.9% saline solution.
- Return containers to the containment level 3 laboratory for disposal in accordance with local SOPs.

##### 6.5.4. Assessment after challenge

- Admit the participant to the quarantine unit.
- Fast for a further 90 minutes.
- Participants who vomit for any reason within 90 minutes of the challenge will be withdrawn from the trial and treated with antibiotics as described in **Section 6.8.3**.
- Instruct participant to notify staff any serious adverse events/reactions that occur prior to next review.
- Instruct participant not to use antipyretics.
- Instruct participant to notify study team if they require the use of any medications.
- Instruct participant to notify study team if they have a temperature ≥38.0$^{o}$C.
- Provide participant with a diary card for recording systemic effects and oral temperatures.

- Check details of a mobile telephone number that the participant will be carrying with them for the 14 days post-challenge. Counsel the participant on the importance of maintaining contact with the study team and keeping the mobile always switched on and with them. Explain to the participant that if they are uncontactable, and their 24-hour contact is unsure of their whereabouts, the study team will make every effort to locate them.

- Issue participant with information on enteric precautions.

- Educate participant on correct hand washing technique, including demonstration and observation.

- Advise participant to inform the study team if any breaches of enteric precautions occur such that another individual meets excreta from the participant.

- Issue participants with liquid hand soap and paper towels to aid with adherence to enteric precautions.

- Instruct participant on obtaining stool specimens (as outlined in the relevant SOPs) and provide the participant with sampling equipment.

Notification of challenge will be provided to the local Health Protection Unit, UKHSA and to the participant's GP.

#### 6.6. Day 0 to Day 7 - Inpatient quarantine

Participants will be admitted to a quarantine facility immediately after challenge. The inpatient quarantine beds will be in the ICRF at the Hammersmith Hospital Campus, Ward 15S at Charring Cross Hospital Campus (Imperial College Healthcare NHS trust), or Ron Johnson Ward, Chelsea and Westminster Hospital (Chelsea and Westminster Hospitals NHS foundation trust).

##### 6.6.1. Post-challenge assessments during inpatient quarantine

During the inpatient quarantine period, the participants will be reviewed at least daily by a member of the study team. Designated assessments will take place during this time, including the following procedures:

- Check continued consent
- Review symptoms profile, diary entries, and laboratory blood results (if available)
- Check for occurrence of any new adverse or medically significant events since previous visit

- Record oral temperature, pulse, and blood pressure
- Perform mood assessment using HADS form (only on Day 7, and the day of antibiotic commencement either if diagnosis is made)
- Perform sample collection as per Table 8
- Prescribe and issue concomitant medication for symptom control if required (see Table 11)

- Re-iterate participant requirements such as completion of the diary, refraining from use of antipyretics, notification of any medication administration, and notification of any fever ≥38.0°C (where appropriate).
- Re-iterate hand hygiene precautions

The quarantine facility will be staffed 24/7 by trained nurses who will work alongside the research team. A clinician will be on-call 24/7 and will be alerted by the staff nurses in case of any abnormal observations. Out of hours visits will be arranged as necessary.

###### 6.6.1.1. Infection control - Enteric precautions

During the inpatient quarantine participants will be managed using infection control protocol appropriate for enteric diseases.

###### 6.6.1.1.1. Hand hygiene

Study investigators will perform hand hygiene using soap-and-water before and after contact with the patient, the patient's immediate environment, and any contaminated articles. Gloves should be removed between procedures, hands washed and dried thoroughly. Typically, alcohol hand rub should not be used in patients with loose stool and diarrhoea.

Participants will be instructed on hand hygiene during the inpatient quarantine stay. Hand washing should be performed regularly, particularly after defecation. Participants will be issued with disinfectant wipes to clean toilet handles, toilet seats and other touch points after toilet use.

###### 6.6.1.1.2. Personal protective equipment Side rooms

Study investigators will wear a disposable apron and gloves during study procedures. Gloves will be changed between procedures. Hand hygiene will be performed after contact with the patient, contaminated articles, or the patient's immediate environment. Participants will be instructed on hand washing procedures. Face masks or other facial protection are not strictly needed unless splashing of body fluids is anticipated. Local guidance may recommend routine face mask use for routine clinical care (including outpatient clinic visits) as per local infection prevention and control team/UKHSA guidance.

###### 6.6.1.1.3. Side rooms

Participants will be managed in single-occupancy side rooms with dedicated toilet facilities.

###### 6.6.1.1.4. Linen

Linen will be treated as contaminated, according to local policies. Linen changes should take place daily.

###### 6.6.1.1.5. Crockery and cutlery

Crockery and cutlery will be handles according to local SOPs and washed using a routine domestic hot wash.

###### 6.6.1.1.6. Clinical waste

Clinical waste will be disposed on in a yellow bag inside the isolation area. If the outside of the bag becomes contaminated, this should be placed inside a second yellow clinical waste bag at the door of the isolation room.

###### 6.6.1.1.7. Room cleaning

Room cleaning will take place daily in accordance with local procedures. Door handles, taps, toilet flushes and bed rails should be cleaned regularly throughout the quarantine period in accordance with local policies.

Terminal cleaning of the room will take place at the end of the quarantine period. All clinical waste must be removed prior to terminal cleaning. The specifics of terminal cleaning are outlined in local policies, but typically this will involve washing of all horizontal surfaces, equipment, taps, door handles and toilet flushes. The floor will be mopped thoroughly with a 1000ppm chlorine solution (or equivalent disinfectant according to local SOPs).

###### 6.6.1.1.8. Bedpans

Disposable cardboard bedpans will be used to measure stool volume and stool type. The bedpan will be covered after use. After collection of the stool samples, the bedpan will be transported to the local macerator for disposal. Ideally, a second member of staff will be present to open doors and lid of the macerator. The bedpan holder/toilet will be cleaned with detergent and wiped with disinfectant.

###### 6.6.1.1.9. Visitors

Access for visitors will be limited, to minimise the risk of transmission. A limited number of visitors would be allowed if participants are not considered actively infectious i.e. in the absence of ongoing type 6/7 stools for 48 hours. Any visitors you have would need to follow hospital policy for enteric precautions, including (but not limited to), wearing a disposable apron and gloves, plus handwashing with soap and water.

###### 6.6.1.2. Food

There will be no specific restrictions to the food that can be eaten during the inpatient quarantine period. Participants will be provided with breakfast, lunch, and dinner during their inpatient stay. Participants will be allowed to bring snacks and order external meals. Participants will be asked to record details of their diet in the diary card.

###### 6.6.1.3. *Salmonella* diagnosis during inpatient quarantine

At any point from Day 0 to Day 7, a participant may meet the criteria for *Salmonella* diagnosis (Table 4) and/or the criteria for commencing treatment initiation (Table 10). These criteria will be assessed at each daily assessment. Participants who meet the criteria for *Salmonella* diagnosis and treatment timepoints will follow an alternate parallel visit schedule as outlined below.

##### 6.6.2. Discharge criteria

The period of quarantine will be for 7 days after challenge (i.e. 168 hours). Participants can be discharged on Day 7 if they meet the criteria for discharge as outlined in **Table 9**. Some participants may be discharged prior to Day 7 if they have been started antibiotic treatment and completed 96 hours of follow up after starting treatment. If these criteria are not met on Day 7, then participants will be asked to remain in quarantine until the discharge criteria are met. An algorithm outlining criteria for discharge is shown in **Figure 3**.

***Table 9 – Criteria for discharge.***

***1 – Resolution of diarrhoea will be taken as 48hrs measured from the time of first type 1-5 stool, with no type-6-7 stools in the interim.***

| *Participants will be discharged from the inpatient quarantine unit if the following criteria are met:* |
|---|
| • Medically fit for discharge in the opinion of the study physician **AND** |
| • Complete resolution of diarrhoea (Bristol stool type 6-7) for 48 hours[1] **AND** |
| • Seven days (168 hours) have elapsed since challenge |
| For patients diagnosed with Salmonellosis from day 0 to day 7, the following criteria apply: |
| • Antibiotic treatment has been initiated and patient has completed SD+96hrs follow up **OR** |
| • Resolution of *Salmonella* Typhimurium bacteraemia (if applicable) |

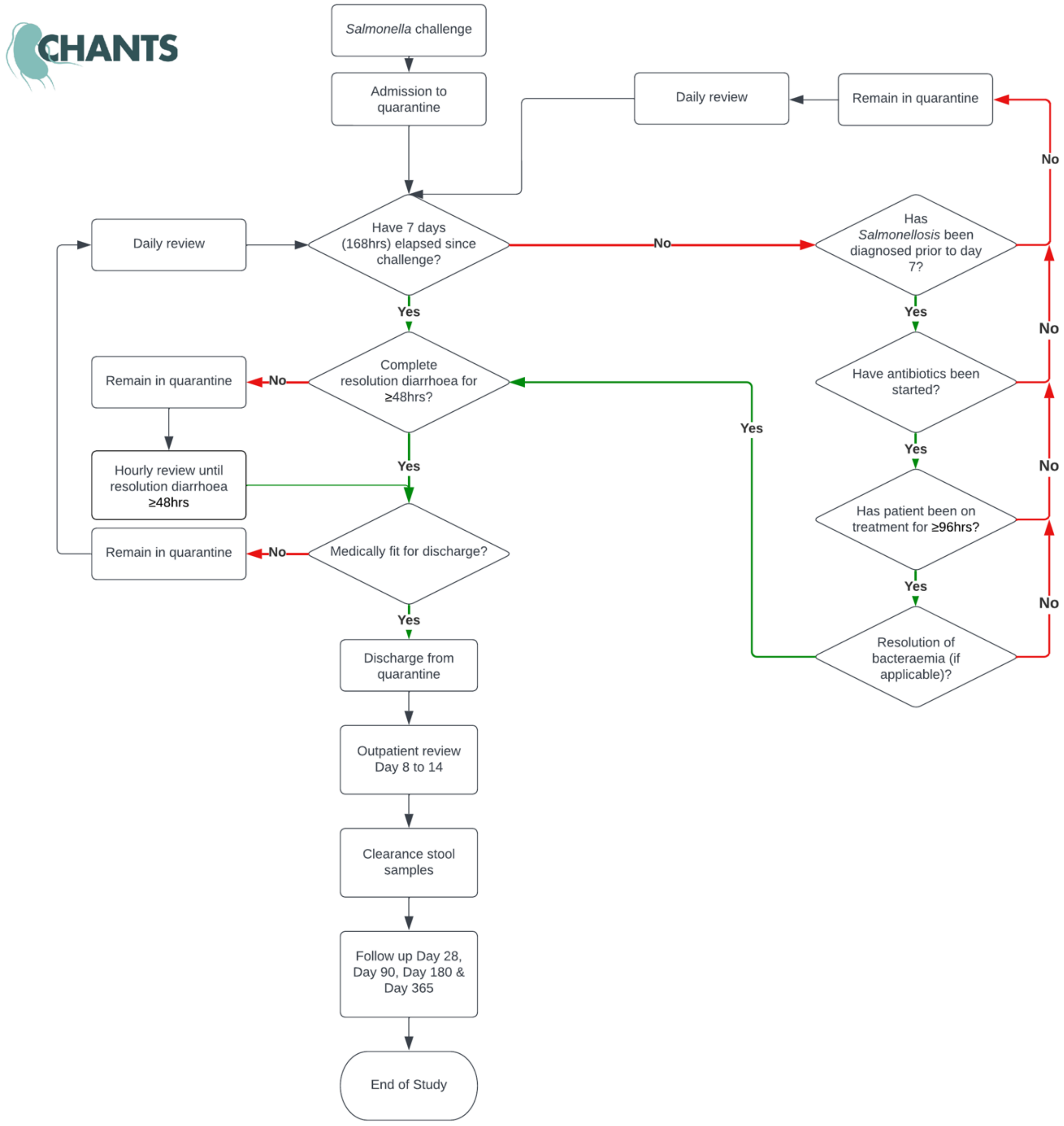

***Figure 3 - Discharge algorithm***

#### 6.7. Day 8 to Day 14 - Post-quarantine visits

After the quarantine period has ended, participants will be asked to attend the clinical facility daily for a further 7 days. If, for any reason they cannot attend, a study doctor will contact them by telephone. Daily blood and stool cultures will be collected at the post-quarantine visits (Day 8 to Day 14). Participants with evidence of *Salmonella* blood stream infection after Day 7 may be required to be re-admitted for observation.

Designated assessments will take place during this time, including the following procedures:

- Check continued consent
- Check for occurrence of any new adverse or medically significant events since previous visit
- Review symptoms profile, diary entries, and laboratory blood results (if available),
- Record oral temperature, pulse, and blood pressure
- Perform mood assessment using HADS form (only on Day 7, and the day of antibiotic commencement if diagnosis is made)
- Perform sample collection as per **Table 8**
- Prescribe and issue concomitant medication for symptom control if required (see Table 11)
- Re-iterate participant requirements such as completion of the diary, refraining from use of antipyretics, notification of any medication administration, and notification of any fever ≥38.0°C (where appropriate).
- Re-iterate hand hygiene precautions
- Check details of a mobile telephone number that the participant will be carrying with them for the 14 days post-challenge. Counsel the participant on the importance of keeping the mobile always switched on and with them. Explain to the participant that if they are uncontactable, and their 24-hour contact is unsure of their whereabouts, the study team will make every effort to locate them. This may include notifying the police.
- Provide details of next clinic appointment

##### 6.7.1. *Salmonella* diagnosis from Day 8 to Day 14

During the outpatient assessment period from Day 8 to Day 14, a participant may meet the criteria for *Salmonella* diagnosis (Table 4) and/or the criteria for commencing treatment initiation (Table 10). These criteria will be assessed at each daily assessment. Participants who meet the criteria for *Salmonella* diagnosis and treatment timepoints will follow an alternate parallel visit schedule as outlined below.

#### 6.8. *Salmonella* diagnosis and treatment timepoints

Participants who meet the primary endpoint for systemic *Salmonellosis* (**Table 4**) and/or those who are started on antibiotic treatment (**Table 10**) prior to day 14 will follow an alternate parallel visit schedule.

##### 6.8.1. Criteria for diagnosis of Salmonellosis

For the purposes of data analysis and reporting of cases to UKHSA and the local health protection team, *Salmonella* infection will be defined as specified in **Table 4**.

###### 6.8.2. *Salmonella* diagnosis and treatment visits (SD, SD+24hrs, SD+48hrs, SD+72hrs, SD+96hrs)

The following procedures will be performed at the diagnosis and treatment visit timepoints.

- Obtain and document the criteria for diagnosis and/or treatment.
- Perform a physical examination at SD and at SD+24 to SD+96 if applicable
- Assessment by a study doctor at the time of SD to assess severity and potential need to remain in the quarantine facility and/or be re-admitted to the quarantine facility.
- Review diary entries and laboratory results (if available),
- Record oral temperature, pulse, and blood pressure,
- Perform sample collection as per **Table 8**.
- Prescribe and issue antibiotic treatment.
- Prescribe and issue concomitant medication for symptom control (if required)
- Schedule next visit and re-iterate participant requirements such as continuing diary entries, adherence to antibiotic therapy and maintaining contact with the study team.

###### 6.8.3. Location of SD to SD96 visits

If a participant meets the criteria for primary diagnosis and/or treatment during the inpatient quarantine period between day 0 to 7, the post diagnosis/treatment visits will take place in the inpatient facility until the participant meets the discharge criteria. If a participant meets the criteria for primary diagnosis and/or treatment during the outpatient follow up period between day 8 to 14, the post diagnosis/treatment visits will take place as an outpatient unless there is a need to re-admit the patient to the quarantine facility in the opinion of the study investigator. The precise location of visits SD to SD96 will depend on the time at which a participant meets the criteria for primary diagnosis. It is anticipated that some post diagnosis visits will take place in an inpatient setting and others in an outpatient setting.

##### 6.9. Antibiotic treatment

Antibiotic therapy will be offered as soon as possible when one or more of the criteria in **Table 10** are satisfied or at Day 14 unless spontaneous clearance has been confirmed (defined in Section 6.9.1.2).

Antibiotics treatment of *Salmonella* bacteraemia will be initiated on the identification of Gram-negative rods on microscopy from a positive blood culture taken from a challenged participant. Treatment will not await speciation.

Severe gastroenteritis is defined in **Section 3.2.1** and is defined as ≥6 loose/liquid stools[1] and/or >800 g of loose/liquid stools in a rolling 24-hour period and/or ≥2 stools with gross blood in 24 hours.

[1] Type 6 -7 on Bristol stool chart

*Salmonella* gastroenteritis is typically self-limiting in healthy immunocompetent adults and treatment may not be indicated in all patients.  Antibiotic treatment can be associated with side effects and in mild cases maybe associated with a risk of prolonged shedding. Participants with *Salmonella* shedding not meeting the criteria for immediate treatment in Table 10 or those with mild diarrhoea, may not benefit from treatment. These participants will be offered the option of 1) no-treatment or 2) antibiotic treatment treated at Day 14 post challenge after being counselled about the risks and benefits of either approach. Treatment decisions will be recorded in the CRF.

An algorithm for treatment decisions is provided in **Figure 3**.

| Antibiotics are commenced if ANY of the following apply |
|---|
| Any participant with *Salmonella* Typhimurium bacteraemia |
| Fever ≥38°C for ≥12hrs |
| Any participant with severe gastroenteritis |
| Moderate gastroenteritis plus:<br>• fever ≥38°C on one occasion and/or<br>• ≥1 Grade 2 systemic symptoms |
| Any participant with 3 or more of the following symptoms on the same day at Grade 2 or higher*;*<br>• Headache<br>• Fatigue/Malaise<br>• Anorexia<br>• Abdominal pain<br>• Nausea<br>• Vomiting<br>• Myalgia<br>• Arthralgia<br>• Cough |
| Any participant from whom *Salmonella* has been detected from at least two stool culture/PCR and 24hrs apart who has not received antibiotics by day 14 post-challenge, unless spontaneous clearance is confirmed on three consecutive serial stool cultures by day 14[a]. |
| Any participant in whom antibiotic use is felt to be clinically necessary (as decided by a medically qualified study doctor) |

***Table 10 – Criteria for commencing antibiotic treatment. a) – Clearance during the challenge period is defined as three consecutive stool cultures testing negative for Salmonella (with no subsequent positive samples) by Day 14 post challenge. If no other treatment criteria are met, spontaneous clearance of Salmonella is assumed. In this instance, the adverse effects of antibiotic treatment may outweigh the benefits. Antibiotic treatment will not routinely be recommended but may be offered to the patient if i) antibiotics are requested by the patient and/or ii) there are compelling alternative reasons for offering antibiotic treatment at the discretion of a study physician in consultation with the study PI.***

##### 6.9.1. Treatment protocols

###### 6.9.1.1. Transient asymptomatic shedding

Treatment will **not** be routinely recommended to patients if all the following criteria are met:

- Isolation of *Salmonella* Typhimurium from stool samples on <2 occasions after challenge
- All blood cultures for *Salmonella* Typhimurium are negative
- Absence of fever ≥38°C for ≥12hrs
- Absence of systemic symptoms as outlined in **Table 10**
- Absence of mild, moderate or severe gastro-enteritis

In these instances, the risks of antibiotic treatment are considered to outweigh the potential benefits. In these cases, treatment may still be offered if felt to be clinically indicated by the senior study investigator.

###### 6.9.1.2. Spontaneous clearance

Treatment will **not** be routinely recommended to patients if all the following criteria are met:

- Isolation of *Salmonella* Typhimurium by culture from stool samples on ≥1 occasion after challenge between days 0 and 11 (regardless of symptoms) AND
- All blood cultures for *Salmonella* Typhimurium are negative.
- Absence of fever ≥38°C for ≥12hrs
- Absence of systemic symptoms as outlined in **Table 10**
- Three consecutive stool cultures have tested negative for *Salmonella* by Day 14.

In these instances, participants will be considered to have spontaneously cleared *Salmonella,* despite evidence of colonisation early in the course of infection. In this instance, the risks of antibiotic treatment are considered to outweigh the potential benefits. In these cases, treatment may still be offered if felt to be clinically indicated by the senior study investigator. Treatment may be offered if i) antibiotics are requested by the patient or ii) there are compelling alternative reasons for offering antibiotic treatment at the discretion of a study physician in consultation with the study PI.

If treatment is offered of it comprise azithromycin 500mg once daily for 5 days. Second line treatment will comprise ciprofloxacin 500mg twice daily for 5 days.

###### 6.9.1.3. Asymptomatic/Pauci-symptomatic Stool shedding

Participants with *Salmonella* stool shedding on ≥2 occasions who do not develop gastroenteritis and who do meet other treatment criteria outlined in **Table 10** will not routinely be offered treatment at day 14, if the following criteria are met.

- Isolation of *Salmonella* Typhimurium by culture from stool samples on ≥2 occasion after challenge between days 0 and 11 (regardless of symptoms) AND
- Three consecutive stool cultures have tested negative for *Salmonella* by Day 14.
- All blood cultures for *Salmonella* Typhimurium are negative
- Absence of fever ≥38°C for ≥12hrs
- Absence of systemic symptoms as outlined in **Table 10**

In these instances, participants will be considered to have spontaneously cleared *Salmonella,* despite evidence of colonisation early in the course of infection. In this instance, the risks of antibiotic treatment are considered to outweigh the potential benefits. In these cases, treatment may still be offered if felt to be clinically indicated by the senior study investigator.

If treatment is offered of it comprise azithromycin 500mg once daily for 5 days. Second line treatment will comprise ciprofloxacin 500mg twice daily for 5 days.

###### 6.9.1.4. Salmonella gastroenteritis

###### 6.9.1.4.1. Treatment for severe gastroenteritis

Participants with severe *Salmonella* gastroenteritis will receive treatment upon meeting the criteria for severe gastroenteritis as defined in **Section 3.3** and **Table 10**. Treatment will comprise azithromycin 500mg once daily for 5 days. Second line treatment will comprise ciprofloxacin 500mg twice daily for 5 days.

###### 6.9.1.4.2. Treatment for moderate gastroenteritis

Participants with moderate *Salmonella* gastroenteritis as defined in **Section 3.3** will be offered treatment upon meeting the criteria for moderate gastro enteritis if accompanied by:

- fever ≥38°C on one occasion and/or
- ≥1 Grade 2 systemic symptoms

Treatment will comprise azithromycin 500mg once daily for 5 days. Second line treatment will comprise ciprofloxacin 500mg twice daily for 5 days.

###### 6.9.1.4.3. Treatment for mild gastroenteritis

Participants with mild *Salmonella* gastroenteritis as defined in **Section 3.3** will initially be symptomatically managed with analgesia and rehydration. Antibiotic treatment will not be initiated at the onset of mild diarrhoea, unless they transition to moderate gastroenteritis with fever and systemic symptoms or severe gastro-enteritis prior to Day 14.

###### 6.9.1.5. *Salmonella* bloodstream infection

Participants with *Salmonella* bloodstream infection will receive treatment if/when blood cultures test positive and Gram negative rods are identified on microscopy. Treatment will comprise ciprofloxacin 500mg twice daily for 10-14 days. Second line treatment will comprise ceftriaxone IV OD for up to 14 days.

###### 6.9.1.6. Systemic illness

Participants may be treated for systemic illness following *S.* Typhimurium challenge as outlined in **Table 10** (i.e. fever ≥38°C for ≥12hrs or 3 or more systemic symptoms severe enough to interfere with all normal activity). Treatment will comprise azithromycin 500mg once daily for 5 days. Second line treatment will comprise ciprofloxacin 500mg twice daily for 5 days.

###### 6.9.2. Treatment of clinical or microbiological relapse

A second course of antibiotic treatment may be offered to participants in instances where *Salmonella* Typhimurium is isolated from faecal specimens after a course of primary treatment (with or without diarrhoea and/or systemic symptoms compatible with *Salmonella* disease). In such instances, the antibiotic offered will belong to a different antibiotic class to that offered for primary treatment. For example, if ciprofloxacin is offered for primary treatment as outlined in Table 10, then azithromycin will be offered for the second course of treatment (and vice versa). In instances of microbiological or clinical relapse, enhanced hand washing precautions will be emphasised. We will confirm clearance by collected three faecal specimens taken at least 48hrs apart, the first of which will be collected at least 7 days after completing the re-treatment course. If, *Salmonella* is persistently isolated from faecal specimens after re-treatment, then a third course of treatment will not be routinely offered within the remit of the study, as asymptomatic shedding with spontaneous clearance is well recognised. [60] Further faecal cultures will be collected at follow up timepoints including Day 90, 180 and 365 as per Table 8. The participant will be offered a referral to an NHS infectious disease specialist as an outpatient if they meet the criteria for chronic carriage i.e. persistent stool culture positivity at 12 months post challenge.

##### 6.10. Concomitant medication for treatment of Salmonella infection

Concomitant medication can be provided for symptomatic control of *Salmonella* infection, before and after diagnosis. Concomitant medication after *Salmonella* diagnosis includes antibiotics and antipyretics if required.

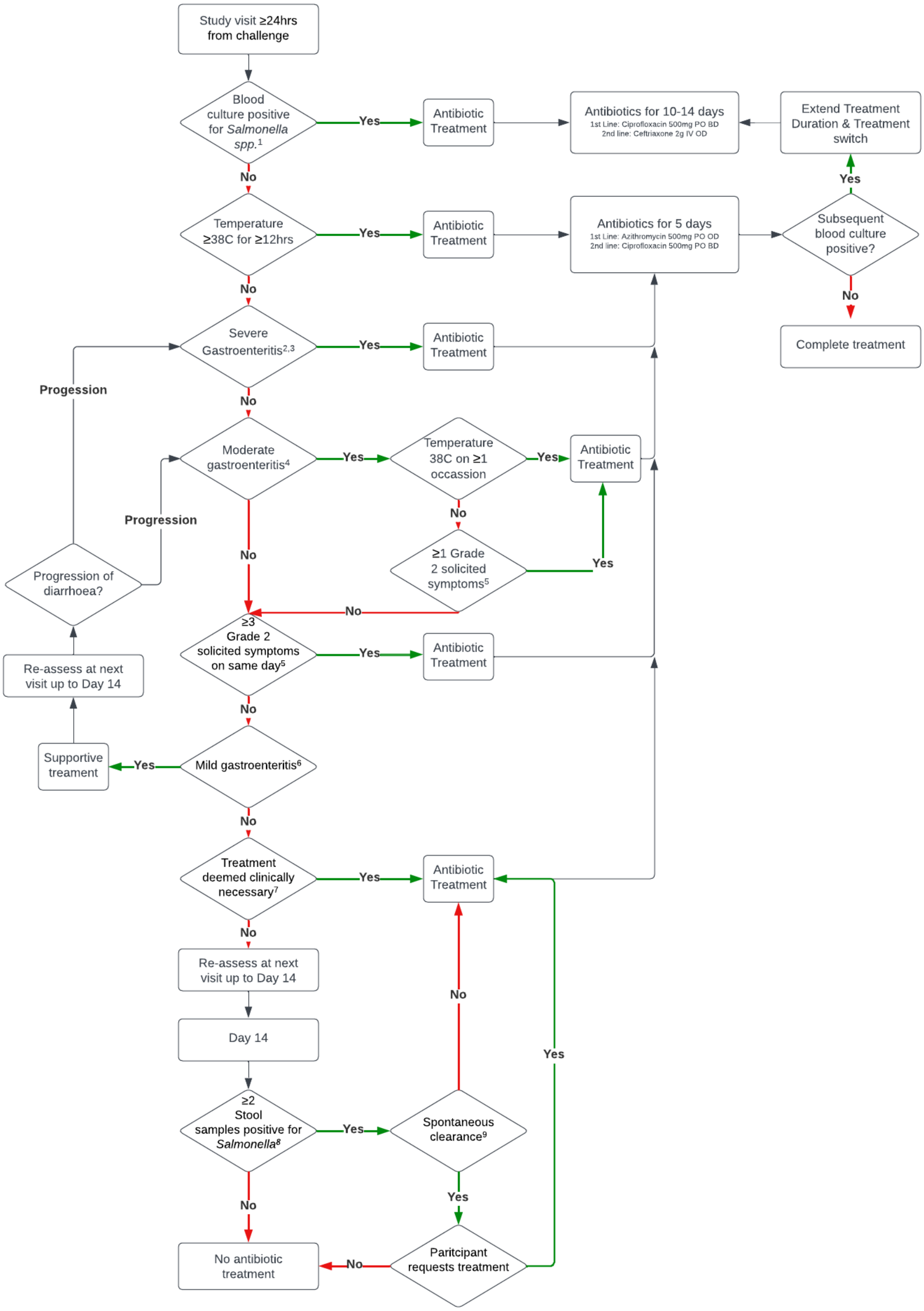
Study visit ≥24hrs from challenge
Blood culture positive for *Salmonella spp.*[1]
Yes
Antibiotic Treatment
Antibiotics for 10-14 days
1st Line: Ciprofloxacin 500mg PO BD
2nd line: Ceftriaxone 2g IV OD
Extend Treatment Duration & Treatment switch
Yes
No
Temperature ≥38C for ≥12hrs
Yes
Antibiotic Treatment
Antibiotics for 5 days
1st Line: Azithromycin 500mg PO OD
2nd line: Ciprofloxacin 500mg PO BD
Subsequent blood culture positive?
No
Complete treatment
No
Severe Gastroenteritis[2,3]
Yes
Antibiotic Treatment
Progession
No
Moderate gastroenteritis[4]
Yes
Temperature 38C on ≥1 occassion
Yes
Antibiotic Treatment
No
≥1 Grade 2 solicited symptoms[5]
Yes
Progression
Progression of diarrhoea?
No
No
≥3 Grade 2 solicited symptoms on same day[5]
Yes
Antibiotic Treatment
Re-assess at next visit up to Day 14
No
Supportive treament
Yes
Mild gastroenteritis[6]
No
Treatment deemed clinically necessary[7]
Yes
Antibiotic Treatment
No
Re-assess at next visit up to Day 14
No
Day 14
Yes
≥2 Stool samples positive for *Salmonella*[8]
Yes
Spontaneous clearance[9]
No
Yes
No antibiotic treatment
No
Paritcipant requests treatment

***Figure 4 - Antibiotic treatment algorithm. 1 - Treatment initiated upon identification of Gram negative rods in blood culture, prior to formal speciation; 2- Loose/liquid stools defined as Type 6 or 7 stools on the Bristol stool chart; 3 - Severe gastroenteritis defined over a rolling 24-hour period as: ≥6 loose/liquid stools and/or >800 g of loose/liquid stools and/or ≥2 stools with gross blood; 4 - Moderate Gastroenteritis defined over a rolling 24-hour period as:4-5 loose/liquid stools and/or 400-800g loose/liquid stools; 5 - Solicited symptoms defined as: Headache, Fatigue/Malaise, Anorexia, Abdominal pain, Nausea, Vomiting, Myalgia, Arthralgia, Cough; 6 - Mild Gastroenteritis defined over a rolling 24-hour period as: 3 loose/liquid stools and/or 200-399g loose/liquid stools; 7 - At the discretion of the principal investigator in consultation with the trial steering committee; 8 - As detected by culture and/or PCR; 9 - Three consecutive stool cultures testing negative for Salmonella by Day 14 post challenge.***

| Drug | Indication | Dose | Route | Frequency |
| --- | --- | --- | --- | --- |
| Ciprofloxacin | *Salmonella* bloodstream infection (1st line) and *Salmonella* gastroenteritis (2nd line) | 500mg | Oral | BD |
| Azithromycin | *Salmonella* gastroenteritis (1st line) | 500mg | Oral | OD |
| Ceftriaxone | *Salmonella* bloodstream infection (2nd line) | 2g | Intravenous | OD |
| Paracetamol | Fever and discomfort | 500mg - 1 gram | Oral | PRN, max QDS |
| Codeine | Pain including headache | 30-60mg | Oral | PRN (max. 240mg/24 hours) |
| Senna | Constipation | 2-4 tablets | Oral | PRN |
| Cyclizine | Nausea and/or vomiting | 50mg | Oral | PRN (max. 150mg/24 hours) |
| Chlorpheniramine | Allergy | 4mg | Oral | PRN TDS-QDS (max. 24mg/24 hours) |
| Oral rehydration solution | Prevention of Dehydration | 200-400ml after each loose stool – approx 2000ml daily | Oral | PRN |
| IV fluids – Sodium chloride 0.9% +/- 20-40mmol KCl | Dehydration – unable to tolerate oral fluids | 250-1000ml | IV | PRN |

***Table 11 - Concomitant medication for treatment and symptom control.***

#### 6.11. Stool culture for assessment of shedding

Clearance cultures following NTS infection are not typically indicated according to UKHSA guidance.[68] This is in contrast to UK public health guidance following infection with the typhoidal *Salmonella* serovars (*S*. Typhi and Paratyphi) Prolonged shedding is known to occur and is thought to contribute to only a small proportion of onward transmission (see **Section 1.5.4**).

In this study we will collect clearance stool cultures to detect differences in patterns of stool shedding between the two challenge strains (See **Objective 2.8**).

To detect convalescent or chronic carriage, stool samples will be collected daily from day 0 to day 14 and at all follow up visits (**Table 7**). In patients treated with antibiotics, three stool samples obtained at least 48 hours apart will be tested one week after completion of the antibiotic course.

Participants with three successive negative stool samples will be designated as cleared of *S.* Typhimurium infection.

If ≥1 clearance samples test positive for *Salmonella*, further clearance stool samples will be collected until at least 3 consecutive samples test negative. Participants with persistent shedding will be referred for Infectious Diseases follow up for further assessment and management.

Patients with chronic carriage of *S.* Typhimurium – designated as persistent shedding 12 months after infection – will be referred for Infectious Diseases follow up for further assessment and management.

UKHSA and the local health protection unit will be informed of all participants with chronic carriage. The employer of any participant involved in the provision of health or social care to vulnerable groups will be notified in writing once clearance is confirmed.

#### 6.12. Reporting to local health protection team

The Northwest London Health Protection Team[28] (UK Health security agency) will be informed of the name, address, and date of birth of all participants who:

- Have been challenged with *S.* Typhimurium
- Have isolated *S.* Typhimurium from at least one stool specimen post challenge
- Have commenced and completed antibiotics, and
- Have completed clearance stool sampling (with additional information and continued contact if persistence stool shedding occurs)

In addition, any breaches in enteric precautions that result in another individual who comes into contact with the excreta of a participant will be reported to the local health protection team.

#### 6.13. Screening of close contacts

The participant will be offered information from the study team to give to close contacts, including household contacts, if they wish. Upon request, contacts will be offered the opportunity to be screened for *Salmonella* infection, which will involve obtaining three stool samples 48-hours apart a

[28] North West London HPT UK Health Security Agency, 61 Colindale Avenue, London, NW9 5EQ;, Telephone 020 3326 1658

minimum of seven days after the participant has begun antibiotic treatment. If either stool culture of a household contact is positive, he/she will be referred to a Consultant in Infectious Diseases for appropriate antibiotic management and the local health protection unit will be informed.

##### 6.14. Transport of samples

All samples from participants must be a labelled with a 'Danger of Infection' sticker when transported outside of the study site and to the designated microbiology laboratory. If a specimen sample bag is to be used, this should also be labelled 'Danger of Infection'. Samples should be transported in accordance with local SOPs.

##### 6.15. Discontinuation/Withdrawal of participants

Each participant can exercise his or her right to withdraw from the study at any time. If, however, the participant decides to withdraw after they are challenged, they may – for personal and public health reasons - be asked to complete a course of antibiotics. This may require attendance at additional hospital/non-study visits to ensure compliance.

In addition, the investigator may terminate a participant's involvement in the study at any time if the investigator considers it necessary for any reason including, though not exclusive to, the following:

- Ineligibility (either arising during the study or in the form of new information not declared or detected at screening),
- Significant protocol deviation,
- Significant non-compliance with study requirements or risk to public health,
- Any adverse event which requires discontinuation of the study procedures or results in an inability to continue to comply with study procedures,
- Consent withdrawn,
- Lost to follow up.

Withdrawal from the study will not result in exclusion of the data generated by that participant from analysis. The reason for withdrawal, if given, will be recorded in the CRF.

##### 6.16. Irritable bowel syndrome questionnaire

There is a theoretical risk of inducing post-infectious irritable bowel syndrome (PI-IBS) following *Salmonella* gastroenteritis. To mitigate this, and to estimate the frequency of this occurrence, we will administer an IBS questionnaire at baseline and post challenge as outlined in **Table 7**.

Participants will complete the Rome IV questionnaire[130] and the irritable bowel severity scoring system, described by Francis and colleagues [131]. This questionnaire has recently been used to describe the characteristics and frequency of post-infectious irritable bowel syndrome following *Campylobacter* enteritis [132].

##### 6.17. Participant Experience Questionnaire

Following completion of their last diary entry participants will be directed to an optional questionnaire regarding their experience of the study. The results of this questionnaire will be anonymised. The questionnaire will be closely based on one used in the previous challenge studies, which aimed to examine participant motivations, attitudes and factors influencing participation in human challenge research.[138]

##### 6.18. Out of schedule visits

Unscheduled visits will be arranged, if required, to ensure participant safety as further history, examination and investigation may be needed. These visits will be at the discretion of the clinical study team. If participants are unwell and unable to attend the study site for a visit, they may be visited at home by one of the study team members.

##### 6.19. Definition of End of Trial

The clinical phase of the study has ended when the last participant completes their last visit. The definition of the end of the study is when the last laboratory assay has been performed.

#### 7. Laboratory

##### 7.1. Blinding of laboratory samples

Details regarding the process of blinding laboratory samples and the laboratory methods to be used are given in relevant SOPs and study plans. Samples processed by departments at the Northwest London Pathology laboratory will be processed using the participant study number and participant initials.

##### 7.2. Bacteriology

###### 7.2.1. Blood culture

After inoculation of aerobic broth with 10 mL of the participant's blood (*BACTEC* PLUS Aerobic/F culture vial; BD, Oxford, UK), culture will be performed using the BACTEC 9240 continuous monitoring system in the microbiology laboratory of the North West London Pathology Laboratory (Charing Cross Hospital, Imperial Healthcare NHS Trust; NWLP) according to current SOPs and UK Standards for Microbiology Investigations ID 24: identification of *Salmonella* species.[139]

Identification of organisms cultured will be by biochemical (API, Analytical Profile Index; bioMérieux, Basingstoke, UK) and serological methods, latterly by agglutination with *S.* Typhimurium anti-sera. Confirmed isolates will be tested for antibiotic susceptibility using standard methods. All *Salmonella* isolates will be sent to the gastrointestinal reference lab for whole genome sequencing.

Quantitative culture of whole blood will be performed to determine the number of organisms in the blood, using the Abbott Isostat® Isolator system (Oxoid Ltd, Basingstoke). Enumeration of *S.*

Typhimurium organisms in the blood will be performed by lysis centrifugation followed by direct plating onto non-selective media.

##### 7.2.2. Stool culture & PCR

Stool samples supplied by participants should be delivered to NWLP within 24 hours of being taken. If possible, the samples should be kept cool until delivered to NWLP and then stored at 2$^{o}$C to 8$^{o}$C. The time of sampling will be noted on the sample form.

*Salmonella* will be detected in stool by both PCR and culture.

All faecal samples will be routinely run on Serosep (real time PCR) for the detection of *Salmonella* sp., *Shigella* sp., Enteroinvasive *E coli*, *Campylobacter jejuni/ lari/ coli*, *Cryptosporidium parvum/ hominis*, *Giardia lamblia* and Verotoxin- producing E.coli (VTEC), according to local SOPs (MIC-LP-169-X Serosep Bacterial Enteric PCR).

Routine stool cultures and screening for enteric pathogens will be performed in parallel by the microbiology laboratory, according to relevant local SOPs (MIC-LP-009-X Enteric C&S SOP) and UK Standards for Microbiology Investigations ID 24: identification of *Salmonella* species.[139]. Stool will be inoculated directly onto XLD agar for semi-quantitative culture and into Selenite F enrichment broth for qualitative culture. After overnight incubation (at 37.0°C), each sample will be sub-cultured onto *Salmonella*-selective chromogenic agar. Suspicious colonies will be identified as per the blood culture method described above.

Stool cultures will be taken at Day 0 (challenge), throughout the 14 day post-challenge period and at visits after *Salmonella* diagnosis. Participants will be required to supply further stool samples until proven not to be shedding *S.* Typhimurium in three consecutive samples.

Isolates of *S*. Typhimurium (from blood and stool culture) will be retained for whole genome sequencing or further investigation by the reference laboratory if challenge strain confirmation is required by PHE/UKHSA.

Additionally, quantitative stool cultures or PCR may be performed at ICL, to assess the burden of stool shedding. Isolates from stool samples will be stored frozen for future analysis, which may include WGS and/or metagenomic sequencing.

Analysis of the relative composition of bacterial populations within the bowel may be performed on collected stool samples. The faecal microbiome at baseline and its effects on challenge outcome, bacterial dynamics and response to antibiotic treatment may be assayed using techniques including pyrosequencing. After collection, stool will be stabilised in RNA-later and stored at -80°C, prior to further assessment.

##### 7.2.3. Microbiological sampling of healthcare environment surfaces

Microbiological sampling of the patient environment will be performed using swabs and contact plates.[140,141] Details of sampling locations are provided in the clinical study plan.

##### 7.2.4. Blood PCR detection

We aim to develop a novel PCR/RT-PCR method for detection of *S.* Typhimurium in the blood of participants after challenge. This will be based on a previously used TSB-bile blood culture-PCR assay which has been used to detect low levels of *S.* Typhi in the blood of participants after challenge in previous studies (OVG 2009/10, OVG 2011/02). Whole blood (5-10ml) is inoculated into 20ml tryptone soya broth containing 3.0% ox bile and micrococcal nuclease. The blood/broth culture is shaken at 220rpm in a 37°C incubator for 5 hours, and then centrifuged at 5,000rpm for 20 minutes. The supernatant is then discarded and the pellet used for DNA isolation.[142] Other novel molecular methods to detect *Salmonella* DNA/RNA may also be used.

#### 7.3. Plasma cytokines

The kinetics of the inflammatory response will be measured in stored plasma samples. A custom commercial multiplex bead-array kit is in use in the laboratory (including, but not limited to, IL-1β, IL-2, IL-6, IL-8, IL-10, IL-12, IL-17A, TNF-α and IFN-γ.). Plasma samples will be isolated from heparinised blood and stored at -80°C until assays are performed.

#### 7.4. Markers of gastrointestinal inflammation

Faecal samples will be collected and assayed for markers of gastrointestinal inflammation, for markers such as (but not limited to) myeloperoxidase, calprotectin and lactoferrin.[1]

#### 7.5. Antibody responses

##### 7.5.1. Serum

Serum will be isolated from blood and stored at -80°C prior to assays being performed. *S.* Typhimurium-specific antibody responses to multiple antigens including O-specific polysaccharides and other antigens baseline and various time points post-challenge. Antibody responses will be assessed using a combination of commercial kits and in-house developed ELISAs.

Functional antibody responses may also be determined using serum bactericidal or opsonophagocytic assays.

We will apply systems serology to further characterise antibody functionality in relation to NTS challenge. These platforms comprise multiplexed high-throughput functional assays alongside high-throughput biophysical profiling of antibody isotype/subclass and Fc-region glycosylation. These are linked to systems biology/machine learning algorithms, which collectively aim to define the specific characteristics of a protective humoral immune to iNTS from within a polyclonal pool of antibodies. These assays are currently being run to characterise the serological response to iNTS disease in field trials. Consequently, we will adopt a hypothesis driven approach, focussing on antigenic targets and functional antibody responses that are associated with protective humoral immune responses against iNTS in the field. System serology studies have been successfully applied by our group to identify new putative correlates of protection in a *S*. Typhi CHIM and can be readily adapted to an iNTS CHIM.[37] Additional studies may include the use of proteome arrays, to define novel target antigens following controlled *Salmonella* Typhimurium infection.[143]

##### 7.6. Mucosal antibody responses

Total and antigen specific salivary IgA will be measured using ELISA. Saliva will be collected using a sponge device inside the mouth, from the mucosa inside the cheek (buccal). The saliva will be transferred from the sponge device into a tube by centrifugation.

IgG and IgA and specific IgG and IgA antibodies against *S*. Typhimurium will be measured in stool samples provided by participants and stored. If specific IgA is detected, IgA1 and IgA2 subclass determination will be performed. The stool specimen will be suspended in a 10% solution of supplemented PBS, centrifuged, and the supernatant assayed for antibody by ELISA. *S.* Typhimurium-specific antibodies will be measured by qualified members of the study team in the local laboratory.

##### 7.7. B-cell responses

Antibody-secreting cells (ASCs) that secrete antibodies against *S.* Typhimurium specific antigens including O-specific polysaccharides and other antigens by using ELISPOT.[144] Briefly, peripheral blood mononuclear cells (PBMCs) will be separated by density gradient centrifugation and added to antigen-coated ELISPOT plates. In the ELISPOT, antigen specific antibody secreted by individual ASCs will be quantified. Memory B-cells responses will be detected after *in vitro* polyclonal stimulation.

##### 7.8. Cellular immune responses

Cellular immune responses will be analysed using PBMCs isolated using density gradient centrifugation.  Assays performed may include activation, proliferation, and cytokine and surface marker measurements using techniques such as intracellular staining and multi-chromatic flow cytometry of cells. Additional analysis may be performed using heavy metal ion tags (CyTOF) to increase the number of parameters being investigated.

##### 7.9. DNA analysis

Analysis of DNA sequencing or the association with single-nucleotide polymorphisms (SNPs) and epigenetics will be performed using DNA extracted from clotted blood, derived from collected serum samples. With consent, these samples will be used to identify host-susceptibility factors for susceptibility and response to challenge using a candidate gene approach. Samples will be taken at baseline and at post-challenge time points to assess for epigenetic changes in DNA transcription.

##### 7.10. Metabolomics

Serum, plasma and/or urine will be collected from participants at baseline and various time points after challenge for metabolomics and proteomic analysis, to compare difference in peripheral signatures following challenge with either D23580 or 4/74 strains of *S*. Typhimurium. Comparable samples from other controlled human infection studies – including viral challenge (influenza, SARS-Cov-2, influenza) and/or previous *S*. Typhi/Paratyphi human challenge studies conducted in Oxford – may be retrieved for comparative analysis.

##### 7.11. Functional genomics

In brief, blood will be collected in Tempus™ or PaxGene Blood RNA Tubes and stored at -80°C until processing. RNA will be extracted and used for study of gene expression profiles. Gene expression profiles will be determined using gene expression microarrays and/or mRNA-seq technologies at baseline and post-challenge time points.

#### 8. Safety Reporting & Adverse events

##### 8.1. Definitions

**Adverse Event (AE):** any untoward medical occurrence in a patient or clinical study subject.

**Serious Adverse Event (SAE):** any untoward medical occurrence or effect that:

- Results in death
- **Is life-threatening** – refers to an event in which the subject was at risk of death at the time of the event; it does not refer to an event which hypothetically might have caused death if it were more severe

- Requires hospitalization, or prolongation of existing inpatients' hospitalization.[29]
- Results in persistent or significant disability or incapacity
- Is a congenital anomaly or birth defect

Medical judgement should be exercised in deciding whether an AE is serious in other situations. Important AEs that are not immediately life-threatening or do not result in death or hospitalisation but may jeopardise the subject or may require intervention to prevent one of the other outcomes listed in the definition above, should also be considered serious.

##### 8.2. Reporting procedures

All adverse events should be recorded at each scheduled and unscheduled study visit as they occur. Depending on the nature of the event the reporting procedures below should be followed. Any questions concerning adverse event reporting should be directed to the Chief Investigator in the first instance.

###### 8.2.1. Non serious AEs

All such events, whether expected or not, should be recorded- it should be specified if only some non-serious AEs will be recorded, any reporting should be consistent with the purpose of the trial end points.

###### 8.2.2. Serious AEs

[29] Not including expected inpatient quarantine as outlined in the study protocol. Re-admission to the quarantine facility following discharge will be considered as an SAE.

An SAE form should be completed and emailed to the Chief Investigator within 24 hours. However, hospitalizations for elective treatment of a pre-existing condition do not need reporting as SAEs.

All SAEs should be reported to the London Fulham REC where in the opinion of the Chief Investigator, the event was:

- 'related', ie resulted from the administration of any of the research procedures; and
- 'unexpected', ie an event that is not listed in the protocol as an expected occurrence

Reports of related and unexpected SAEs should be submitted within 15 days of the Chief Investigator becoming aware of the event, using the NRES SAE form for non-IMP studies. The Chief Investigator must also notify the Sponsor of all related and unexpected SAEs.

Local investigators should report any SAEs as required by their Local Research Ethics Committee, Sponsor and/or Research & Development Office.

Contact details for reporting SAEs


Please send SAE forms to: (sponsor) and (REC).

#### 8.3. Expected adverse events

##### 8.3.1. Definition of an Expected Adverse Event

Any expected adverse event related to symptomatic *Salmonella* Typhimurium infection (as specified in **Table 12**) will be deemed an 'Expected Adverse Event' and will be documented in the eCRF, however, not in the 'Adverse Event' section of the eCRF. These symptoms will be considered 'Expected Adverse Events' if they occur from Day 1 and Day 14 post-challenge (inclusive) or between Day 3 and SD +96 hours. Expected events can be recorded as adverse events, at the discretion of study staff, if they are believed to be unusual (for example, in severity, frequency, duration or character).

Additionally, symptoms occurring outside of the specified time frame will also be considered adverse events.

| **Expected Events – *Salmonella* Typhimurium Infection** |
|---|
| Diarrhoea |
| Abdominal pain |
| Loss of appetite |
| Nausea |
| Vomiting |
| Tenesmus |
| Headache |
| Fever >37.5°C but <40.0°C (fever ≥40.0°C will be considered an AE) |
| Malaise |
| Myalgia |
| Arthralgia |
| Cough |

***Table 12 - Events which will be attributed to development of symptomatic Salmonella infection***

##### 8.3.2. Causality

Causality assessment will follow the guidelines from existing SOPs, except for causality attribution and classification that will be based on the following criteria.

- **No relationship -** No temporal relationship to *S*. Typhimurium ingestion, *and* alternative aetiology (clinical, environmental or other intervention), *and* does not follow pattern of recognised response to NTS infection.
- **Possible -** Reasonable temporal relationship to *S*. Typhimurium ingestion, *or* event not readily explained by alternative aetiology (clinical, environmental, or other interventions), *or* similar pattern of response to that seen to NTS infection.
- **Probable -** Reasonable temporal relationship to *S*. Typhimurium ingestion, *and* event not readily produced by alternative aetiology (clinical, environment, or other interventions), *or* known pattern of response with NTS infection.

- **Definite -** Reasonable temporal relationship to *S.* Typhimurium ingestion, *and* event not readily produced by alternative aetiology (clinical, environment, or other interventions), *and* known pattern of response to NTS infection.

##### 8.3.3. Adverse Events of Special Interest (AESI)

The following events will be considered AESIs and reported to the DSMC in the same manner as for reporting SAEs:

***Table 13 – Adverse events of special interest***

| **Adverse Events of Special interest – *Salmonella* Typhimurium Infection** |
|---|
| Severe *Salmonella* Typhimurium infection (**Table 6**). |
| Clinical or microbiological relapse of *Salmonella* gastroenteritis following completion of therapy |
| Clinical or microbiological relapse of *Salmonella* bacteraemia following completion of therapy |
| Progression to a chronic carrier state[30] |
| Reactive arthritis |
| Pregnancy |
| Transmission of *S.* Typhimurium to a contact of a participant |
| AEs requiring a physician visit or Emergency Department visit which, in the opinion of study staff, are related to the challenge with *S*. Typhimurium. |

Participants with severe *Salmonella* Typhimurium infection – as outlined in **Table 6** – will be closely monitored during the initial study phase, until a course of antibiotic is completed, minimising the risk of complications.

The possible adverse effects of *S.* Typhimurium infection or the effect of some antibiotics on the outcome of pregnancy are unknown. Therefore, pregnant women will be excluded by history and laboratory tests, and female participants will be specifically instructed to prevent conception during the challenge period of the study until completion of antibiotic therapy and clearance of *Salmonella* infection is confirmed. Should pregnancy occur, information about outcome of the pregnancy will be sought.

##### 8.3.4. Safety Profile Review

A sentinel group of two participants will be challenged with either one of the 4/74 or D3580 strains. The sentinel group will be randomised using a block size of two. If there are no safety concerns (as assessed by a study doctor and discussed with the senior investigator and/or clinician as appropriate) by Day 14 post-challenge, the remaining participants will be challenged. Randomisation using varying block sizes will occur thereafter. The DSMC chair and/or other members of the DSMC committee will be updated at regular time points following the initial challenge safety profile will be reviewed after the first 10 participants have been challenged and upon completion of all participants reaching Day 28. Any concerns will be referred to the DSMC Chair.

[30] Defined as isolation of *Salmonella* Typhimurium from the stool 1 year post challenge

##### 8.4. Data safety monitoring committee

A Data and Safety Monitoring Committee (DSMC) will be appointed to provide real-time oversight of safety and trial conduct.

The DSMC is independent and will review unblinded safety data throughout the study according to the DSMC Charter. The DSMC will have access to data and, if required, will monitor these data and make recommendations to the study investigators on whether there are any ethical or safety reasons why the trial should not continue. The DSMC will particularly review the attack rate and advise of dose-escalation/de-escalation decisions. They will review unblinded data to confirm there are no significant safety differences between the two challenge strains. A summary of all AESIs and SAEs to date will be provided to the DSMC on request. The DSMC will also be notified if the study team have any concerns regarding the safety of a participant or the general public (e.g. if a participant is not contactable after *S*. Typhimurium challenge and potentially infectious to others).

The outcome of each DSMC review will be communicated directly to the study investigators and documentation of all reviews will be kept in the TMF. The Chair of the DSMC will also be contacted for advice when the Chief Investigator feels independent advice or review is required.

###### 8.4.1. Stopping and pausing rules

Further challenge of participants will be temporarily paused if a participant develops an adverse event of special interest (**Table 13**) or if a participant develops bacteraemia. All AESIs and cases of bacteraemia will be reviewed by the DSMC in an unblinded fashion. The DSMC will provide recommendations to the trial steering committee as to whether if it is safe and appropriate to challenge further participants and/or if protocol adjustments are required.

###### 8.4.2. Review of bacteraemia

All cases of *Salmonella* Typhimurium bacteraemia will be reviewed by the DSMC. If a participant develops *Salmonella* Typhimurium bacteraemia, no further participants will be challenged with either strain until that participant has completed treatment and further recommendations are given by the DSMC. The DSMC will make recommendations to the trial steering committee if it is safe and appropriate to challenge further participants and/or if protocol adjustments are required.

##### 8.5. Trial Management Committee

The trial investigators will form the trial management committee and will provide on-going management of the trial.

##### 8.6. Procedure to be followed in the event of an abnormal finding

Abnormal clinical findings from medical history, examination, or blood tests, will be assessed as to their clinical significance using the severity grading criteria for Adverse Events. If a test result is deemed clinically significant, it may be repeated, to ensure it is not a single occurrence. If a test remains clinically significant, the participant will be informed, and appropriate medical care will be arranged with the permission of the participant. Decisions to exclude potential participants from enrolling in the trial or to withdraw a participant from the trial will be at the discretion of the study team.

| Symptom | Grade 0 | Grade 1 | Grade 2 | Grade 3 | Grade 4 |
|---|---|---|---|---|---|
| **Vomiting** | Not present | Present but no interference with activity or 1-2 episodes/24 hours | Some interference with activity or >2 episodes/24 hours | Prevents daily activity OR requires outpatient IV hydration | ED visit or hospitalization for hypotensive shock |
| **Nausea** | Not present | No interference with activity | Some interference with activity not requiring medical intervention | Significant; prevents daily activity | ED visit or hospitalization |
| **Diarrhoea** | Not present | 2-3 loose stools or <400g /24 hrs | 4-5 loose stools or 400-800g/24 hrs | 6 or more watery stools or >800g/24 hrs or requires outpatient IV hydration | ED visit or hospitalization for hypotensive shock |
| **Abdominal pain** | Not present | No interference with activity | Repeated use of non-narcotic pain reliever > 24 hours or some interference with activity | Significant; any use of narcotic pain reliever or prevents daily activity | ED visit or hospitalization |
| **Tenesmus** | Not present | No interference with activity | Some interference with activity not requiring medical intervention | Significant; prevents daily activity | ED visit or hospitalization |
| **Headache** | Not present | No interference with activity | Repeated use of non-narcotic pain reliever > 24 hours or some interference with activity | Significant; any use of narcotic pain reliever or prevents daily activity | ED visit or hospitalization |

| | | | | | |
|---|---|---|---|---|---|
| **Fatigue/Malaise** | Not present | No interference with activity | Some interference with activity not requiring medical intervention | Significant; prevents daily activity | ED visit or hospitalization |
| **Myalgia** | Not present | No interference with activity | Some interference with activity not requiring medical intervention | Significant; prevents daily activity | ED visit or hospitalization |
| **Arthralgia** | Not present | No interference with activity | Some interference with activity not requiring medical intervention | Significant; prevents daily activity | ED visit or hospitalization |
| **Cough** | Not present | No interference with activity | Some interference with activity not requiring medical intervention | Significant; prevents daily activity | ED visit or hospitalization |

***Table 14 - Grading the severity of solicited and unsolicited systemic Adverse Events***

| Observation | Grade 0 | Grade 1 | Grade 2 | Grade 3 | Grade 4 |
|---|---|---|---|---|---|
| **Oral temperature (°C)** | 35.5-37.5 | 37.6 – 38.0 | 38.1 – 38.9 | 39-40.0 | >40.0 |
| **Tachycardia (beats/min)** | 55-100 | 101-115 | 116-130 | >130 | ED visit or hospitalization for arrhythmia |
| **Bradycardia (beats/min)** | 100-55 | 50-54 | 45-49 | <45 | ED visit or hospitalization for arrhythmia |
| **Systolic hyper-tension (mmHg)** | 90-140 | 141-150 | 151-155 | >155 | ED visit or hospitalization for malignant hypertension |
| **Diastolic hyper-tension (mmHg)** | <90 | 91-95 | 96-100 | >100 | ED visit or hospitalization for malignant hypertension |
| **Systolic hypo-tension (mmHg)** | >90 | 85-89 | 80-84 | <80 | ED visit or hospitalization for hypotensive shock |

***Table 15 - Grading the severity of visit observed Adverse Events***

| Parameter | Grade 1 | Grade 2 | Grade 3 | Grade 4* |
|---|---|---|---|---|
| **Haemoglobin: decrease from baseline value (g/l)** | < 15 | 15-20 | 21-50 | >5 |
| **White cell count: elevated (cell/mm$^3$)** | 10,800–15,000 | 15,001–20,000 | 20,001–25,000 | >25,000 |
| **White cell count: depressed (cells/mm3)** | 2500-3500 | 1500-2499 | 1000-1499 | <1000 |
| **Neutrophil count (cells/mm$^3$)** | 1500-2000 | 1000-1499 | 500-999 | <500 |
| **Platelets (cells/mm$^3$)** | 125,000-140,000 | 100,000-124,000 | 25,000-99,000 | <25,000 |
| **Sodium: hyponatraemia (mmol/L)** | 132–134 | 130–131 | 125–129 | <125 |
| **Sodium: hypernatraemia (mmol/L)** | 144–145 | 146–147 | 148–150 | >150 |
| **Potassium: hyperkalaemia (mmol/L)** | 5.1–5.2 | 5.3–5.4 | 5.5–5.6 | >5.6 |
| **Potassium: hypokalaemia (mmol/L)** | 3.5–3.6 | 3.3–3.4 | 3.1–3.2 | <3.1 |
| **Urea (mmol/L)** | 8.2–8.9 | 9.0–11 | >11 | RRT |
| **Creatinine (µmol/L)** | 132-150 | 151-176 | 177-221 | >221 or RRT |
| **ALT and/or AST (IU/L)** | 1.1–2.5 x ULN | >2.6–5.0 x ULN | 5.1-10 x ULN | >10 x ULN |
| **Bilirubin, with increase in LFTs (µmol/L)** | 1.1–1.25 x ULN | 1.26–1.5 x ULN | 1.51–1.75 x ULN | >1.75 x ULN |
| **Bilirubin, with normal LFTs (µmol/L)** | 1.1–1.5 x ULN | 1.6–2.0 x ULN | 2.1–3.0 x ULN | >3.0 x ULN |
| **Alkaline phosphatase (IU/L)** | 1.1–2.0 x ULN | 2.1–3.0 x ULN | 3.1–10 x ULN | >10 x ULN |
| **Amylase (IU/L)** | 1.1–1.5 x ULN | 1.6–2.0 x ULN | 2.1–5.0 x ULN | >5.0 x ULN |
| **Albumin: hypoalbuminaemia (g/L)** | 28–31 | 25–27 | <25 | Not applicable |
| **C-reactive protein** | >10-30 | 31-100 | 101-200 | >200 |

***Table 16 - Grading the severity of laboratory Adverse Events***

##### 8.7. Staff and Investigator Safety

All staff working on the project will be required to follow strict infection control techniques as outlined in local SOPs. All staff members working at the research sites will be informed of the commencement of the challenge study.

##### 8.8. Annual progress report

An Annual Progress Report (APR) of the trial will be submitted once a year by the Chief Investigator to the Research Ethics Committee and the Sponsor. The APR will be submitted within 30 days the anniversary date of the favourable opinion of that REC.

#### 9. Statistics and Data Analysis

##### 9.1. Description of statistical methods

This dose-finding study will use the continual reassessment method. The CRM is a model-based design, which was first proposed in 1990. It is widely applied in cancer drug dose-finding early phase studies to replace the traditional 3+3 design (rule-based design).[145,146] The most attractive characteristics of CRM compared with the traditional rule-based dose finding studies is that the CRM method borrows information across all dose levels, which means it will find the true dose with higher probability than rule-based method in most situations, especially when the maximum sample size is small. To our best knowledge, this will be the first dose-finding study using CRM for CHIM, while rule-based method (**Figure 4** ) was used in previous CHIM dose-finding studies. [48]

The parameters for the CRM are presented in **section 3.1**.

In this section, we will present the simulation results by comparing the CRM and rule-based method as to the probability of choosing the correct dose and average sample size needed under the below scenarios:

- Scenario 1 target dose at dose 1 with attack rate: 0.675, 0.8, 0.9, 0.95;
- Scenario 2 target dose at dose 2 with attack rate: 0.5, 0.675, 0.8, 0.9;
- Scenario 3 target dose at dose 4 with attack rate: 0.3, 0.4, 0.5, 0.675;
- Scenario 4 target dose beyond dose 4 with attack rate: 0.2, 0.3, 0.4, 0.5;
- Scenario 5 target dose under dose 1 with attack rate: 0.8, 0.9, 0.95, 0.975.

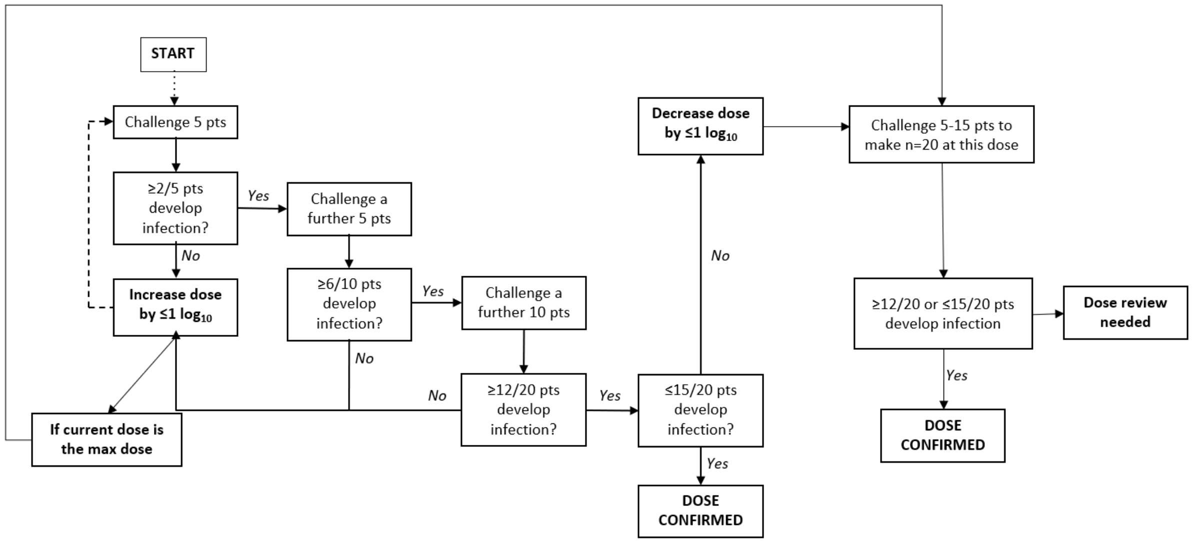

***Figure 5 - Rule-based dose finding study design***

#### 9.2. Simulation results

***Table 17 - Simulation results comparing CRM with rule-based model***

| | **Modified model-based** | Model-based | Rule-based |
|---|---|---|---|
| ***Scenario 1*** | | | |
| Median sample size (IQR) | 15 (15-20) | 20 (20-25) | 20 (20-25) |
| Probability of choosing the dose | | | |
| First dose too high | 24.4% | 17.2% | 16.4% |
| **1 (67.5%)** | **59.3%** | **69.1%** | **58.4%** |
| 2 (80%) | 16% | 13.7% | 8.6% |
| 3 (90%) | 3% | - | - |
| 4 (95%) | - | - | - |
| Fail to find a dose | - | - | - |
| ***Scenario 2*** | | | |
| Median sample size (IQR) | 25 (20-30) | 30 (25-35) | 30 (25-35) |
| Probability of choosing the dose | | | |
| First dose too high | 1.7% | 0.7% | 0.8% |
| 1 (50%) | 27% | 20.6% | 17.3% |
| **2 (67.5%)** | **58.7%** | **64.9%** | **49.4%** |
| 3 (80%) | 12.3% | 13.7% | 3.9% |
| 4 (90%) | 0.3% | 0.1% | - |
| Fail to find a dose | - | NA | 28.6% |

| | | | |
|---|---|---|---|
| ***Scenario 3*** | | | |
| Median sample size (IQR) | 30 (30-35) | 35 (35-40) | 30 (20-40) |
| Probability of choosing the dose | | | |
| First dose too high | | | |
| 1 (30%) | 1% | 0.02% | 0.6% |
| 2 (40%) | 4.5% | 2.2% | 4.4% |
| 3 (50%) | 40.3% | 34.1% | 14.1% |
| **4 (67.5%)** | **50%** | **59.1%** | **11%** |
| Last dose too low | 4.2% | 4.4% | - |
| Fail to find a dose | NA | NA | 69.9% |
| ***Scenario 4*** | | | |
| Median sample size (IQR) | 30 (30-35) | 35 (35-40) | 30 (25-35) |
| Probability of choosing the dose | | | |
| First dose too high | | | |
| 1 (20%) | - | - | - |
| 2 (30%) | 0.3% | 0.1% | 0.8% |
| 3 (40%) | 8.7% | 5.3% | 3.1% |
| 4 (50%) | **51.1%** | 46.0% | 10.7% |
| **Last dose too low** | **39.9%** | **48.6%** | **-** |
| Fail to find a dose | NA | NA | 85.4% |
| ***Scenario 5*** | | | |
| Median sample size (IQR) | 15 (15-15) | 20 (20-20) | 20 (20-20) |
| Probability of choosing the dose | | | |
| **First dose too high** | **66.1%** | **64.7%** | **63.2%** |
| 1 (80%) | 33.4% | 35% | 32.3% |
| 2 (90%) | 0.5% | 0.3% | 0.1% |
| 3 (95%) | - | - | - |
| 4 (97.5%) | - | - | - |
| Fail to find a dose | NA | NA | 4.4% |

The dose selection process was be evaluated using three methods: a modified model-based approach, a standard model-based approach, and a rule-based approach, across five predefined scenarios. The modified model-based method employs an early stopping rule with a sample size of n=15 if challenged at the recommended dose, as opposed to n=20, allowing for earlier trial termination. Overall, the modified model-based method demonstrates superior performance compared to the rule-based approach in terms of dose recommendation accuracy. For instance, in scenarios where the first dose is too high (e.g., Scenario 1), the modified model-based approach reduces the probability of selecting an excessively high dose (24.4%) compared to rule-based methods (16.4%). Additionally, the modified model-based approach achieves a median sample size of 15 (IQR 15–20), significantly smaller than the 20–25 participants required by the rule-based method. However, a potential limitation arises in scenarios where the last dose is too low (Scenario 4). In this case, the modified model is more likely to recommend Dose 4 than the correct "too-low" recommendation, which could lead to suboptimal conclusions. Despite this limitation, the modified model consistently balances safety, efficiency, and accuracy, making it the preferred approach for dose selection in this study.

To meet the primary objective of determining the challenge inoculum required for a clinically reproducible attack rate of 60% to 75%, we will use dose escalation strategy with a starting dose of 1-5x$10^3$ CFU. The dose escalation strategy is described in **Figure 2**.

###### 9.2.1. Sample size

There is no formal sample size calculation for this dose-finding study. The maximum sample size for each challenge dose will be 40 participants. The study and the parameters for the CRM are specified in **Section 3.1** and **Figure 2.**

###### 9.2.2. Replacements

As the study is double blinded, participants will be NOT replaced if they withdrew within 14 days post challenge and before the diagnosis of *Salmonellosis has been reached*.

##### 9.3. Analysis of demographic and baseline characteristics

Descriptive statistics relating to participant characteristics at baseline will be calculated overall and by group. No formal statistical comparisons of baseline characteristics between randomised groups will be conducted.

##### 9.4. Analysis of the primary endpoint

The analysis of the primary endpoint will be descriptive only. The percentage of participants in each group, who meet the criteria for diagnosis of *Salmonellosis*, will be calculated using a 95% Clopper-Pearson Exact confidence interval. The numerator will be the number of participants who meet the criteria for diagnosis and the denominator will include all participants excluding those who withdrew or were treated prior to Day 14 without being diagnosed.

A secondary analysis of the primary endpoint will be conducted using the Kaplan-Meier method which will include all participants. Participants who withdrew or were treated prior to Day 14 and had no diagnosis of *Salmonellosis* will be censored in the analysis at the time of withdrawal or treatment. Participants who were not diagnosed with *Salmonellosis* and were treated at Day 14 will be counted as censoring at Day 14 in the analysis.

The difference in proportions of diagnosed participants between the two challenge strains will be analysed using Pearson's chi-squared test (or Fisher's Exact test if expected counts in any group are less than 5).

##### 9.5. Analysis of secondary endpoints

The analysis for secondary endpoints will follow the primary endpoint analysis.
For the time-to-event analyses of secondary endpoints (e.g. positive blood culture, oral temperature ≥ 38.0°C etc.) all participants will be included. Participants not meeting the criteria for an individual secondary endpoint will be censored in the analysis at, the time of withdrawal, the time of diagnosis or Day 14, whichever is earlier.

A detailed SAP will be signed off before the final analysis.

##### 9.6. Endpoint definitions

Study endpoint definitions are detailed in **section 3.2.1.**

##### 9.7. Level of statistical significance

The main study outcome is a proportion, which will be quoted with 95% confidence intervals. 95% confidence intervals will be used throughout.

##### 9.8. Criteria for termination of the study

The trial steering committee with the Data and Safety Monitoring Committee will have the right to terminate the study at any time on grounds of participant safety. If the study is prematurely terminated the clinical study team will promptly inform the participants and will ensure appropriate therapy and follow-up. If the study is halted, the Sponsor, Imperial College Healthcare NHS Trust and the relevant Ethics Committee will be notified within 15 days of this occurring.

##### 9.9. Procedure for accounting for missing, unused and spurious data

All available data will be used in the analyses and there will be no imputations for missing data. Participants will be analysed according to the type of challenge received for summaries by group.
Participants will be replaced if the diagnosis of *Salmonellosis* has not been reached by the time of withdrawal. Similarly, additional participants will replace participants who require treatment before the formal diagnosis of *Salmonellosis* or Day 14. Participants withdrawn after this time period (i.e. after being diagnosed with *Salmonellosis*, or after 14 days post-challenge) may be replaced at the discretion of the Chief Investigator.

##### 9.10. Procedures for Reporting deviations from the Original Statistical Plan

Any additional analysis or deviations from the analysis plan will be documented and updated according to the statistical standard operating procedure.

#### 10. Regulatory issues

##### 10.1. Declaration of Helsinki

The Investigator will ensure that this study is conducted in accordance with the principles of the current version of the Declaration of Helsinki.

##### 10.2. Guidelines for Good Clinical Practice

The Investigator will ensure that this study is conducted in accordance with relevant regulations and with Good Clinical Practice.

##### 10.3. Ethics approval

The Study Coordination Centre has obtained approval from the London - Fulham Research Ethics Committee (REC) and Health Research Authority (HRA). The study must also receive confirmation of capacity and capability from each participating NHS Trust before accepting participants into the study or any research activity is carried out. The study will be conducted in accordance with the recommendations for physicians involved in research on human subjects adopted by the 18th World Medical Assembly, Helsinki 1964 and later revisions.

##### 10.4. Consent

Consent to enter the study must be sought from each participant only after a full explanation has been given, an information leaflet offered, and time allowed for consideration. Signed participant consent should be obtained. The right of the participant to refuse to participate without giving reasons must be respected. After the participant has entered the study, the clinician remains free to give alternative treatment to that specified in the protocol at any stage if he/she feels it is in the participant's best interest, but the reasons for doing so should be recorded. In these cases, the participants remain within the study for the purposes of follow-up and data analysis. All participants are free to withdraw at any time from the protocol treatment without giving reasons and without prejudicing further treatment.

##### 10.5. Confidentiality

The study staff will ensure that participants' anonymity is maintained. Study participants will be identified by initials and a participant ID number on the CRF. Any electronic databases and documents with participant identifying details will be stored securely and will only be accessible by study staff and authorised personnel. The study will comply with the Data Protection Act, which requires data to be anonymised as soon as it is practical to do so.

The Chief Investigator will preserve the confidentiality of participants taking part in the study and is registered under the Data Protection Act.

Data will be pseudonymised.

Pseudonymised data may be transferred to the University of Oxford for analysis of endpoints.

##### 10.6. Indemnity

Imperial College London holds negligent harm and non-negligent harm insurance policies which apply to this study.

##### 10.7. Sponsor

Imperial College London will act as the main Sponsor for this study. Delegated responsibilities will be assigned to the NHS trusts taking part in this study.

##### 10.8. Funding

Funding for the study has been provided by the Wellcome trust.

##### 10.9. Audits

The study may be subject to audit by Imperial College London under their remit as sponsor and other regulatory bodies to ensure adherence to GCP and the UK Policy Framework for Health and Social Care Research.

An assigned quality assurance manager will operate an internal audit program to ensure that the systems used to conduct clinical research are present, functional, and enable research to be conducted in accordance with study protocols and regulatory requirements. Audits include laboratory activities covering sample receipt, processing and storage and assay validation. The internal audits will supplement the external monitoring process and will review processes not covered by the external monitor.

The Sponsor may carry out audits to ensure compliance with the protocol, GCP and appropriate regulations.

##### 10.10. Participant reimbursement

Participants who attend all routine study visits will receive up to £3002.05 if they remain in the study for the entire period and attend all routinely scheduled study visits. All participants will be reimbursed for their time, travel and for inconvenience based on the following figures:

- Travel expenses: £15 per visit (total for up to 21 visits including screening and delivery of clearance samples = £270)
- Inconvenience of blood tests: £10 per blood donation (total for up to 21 blood tests, including screening = £210)
- Inconvenience of screening ultrasound scan: £50 (total for 1 scan = £50)
- Inconvenience of providing clearance stool samples: £10 per visit (total for three visits £30)
- Time off work compensation – calculated using the London living wage rate of £11.05 an hour[31] - Total : £2442.05

Some participants may be asked to attend additional visits for the purposes of safety follow up. For example, if a participant is diagnosed with *Salmonellosis* at the end of the 14-day monitoring period, they will be asked to attend additional clinic visits and have additional blood tests taken. Additional reimbursement is provided for up to 5 extra visits, which will be reimbursed on a pro-rata basis. If a participant attends all extra 5 visits, the maximum additional reimbursement will be £235.50.

Time off work reimbursement is limited to the 10 days after challenge, to account for the 7 day quarantine period plus additional days off that may be required post quarantine. Payments will be

[31] An hourly rate will be paid depending on the duration of the visit and will include travel time to and from the clinic. The screening visit will be compensated for 4 hours. The pre-screening visit on Day -7 will be compensated for 3 hours. Visits on day 11, 12, 13, 14, 28, 90, 180, 365 and additional study visits will be compensated for 2 hours. The days in quarantine (Day 0 to 7) will be compensated for 18 hours. Days 8, 9 and 10 will also be compensated for 18 hours in case of the need for a prolonged admission.

made via bank transfer. Participants will be required to provide banking details including account name, sort code and account number. All personal banking details will be stored confidentially and retained by the study team while the participant is actively involved in the study. Consent will be obtained prior to requesting and storing personal bank account details.

Participant payments will be requested at the following visits: Screening, Day 28, Day 90, Day 180, Day 365. Due to the reimbursement for scheduled visits, participants will not be given extra reimbursement for unscheduled visits.

#### 11. Study management

The day-to-day management of the study will be co-ordinated through Imperial college London.

##### 11.1. Quality assurance procedures

The study will be conducted in accordance with the current approved protocol, GCP, relevant regulations and standard operating procedures. Approved and relevant Standard Operating Procedures (SOPs) and Laboratory and Clinical Study Plans will be used at all clinical and laboratory sites.

##### 11.2. Monitoring

Monitoring will be performed according to Good Clinical Practice (GCP) by parties appointed by the Chief Investigator. Following written SOPs, the monitors will verify that the clinical trial is conducted and data are generated, documented and reported in compliance with the protocol, GCP and the applicable regulatory requirements. The investigator sites will provide direct access to all trial related source data/documents and reports for the purpose of monitoring and auditing by the Sponsor and inspection by local and regulatory authorities.

##### 11.3. Direct access to source data/documents

Direct access will be granted to authorised representatives from the Sponsor, host institution and the regulatory authorities to permit trial-related monitoring, audits and inspections.

##### 11.4. Modifications to the protocol

No amendments to this protocol will be made without consultation with – and agreement from -- the Sponsor. Any amendments to the study that appear necessary during the study must be discussed with the Investigator and Sponsor concurrently. If agreement is reached concerning the need for an amendment, it will be produced in writing by the Chief Investigator and will be made a formal part of the protocol following ethical and regulatory approval.

The Investigator is responsible for ensuring that changes to an approved study, during the period for which NHS REC approval has already been given, are not initiated without NHS REC review and approval except to eliminate apparent immediate hazards to the participant.

##### 11.5. Protocol deviation

Any deviations will be documented in a protocol deviation form and filed in the TMF.

#### 12. Publication policy

The Chief Investigator will co-ordinate dissemination of data from this study.  All publications (e.g., manuscripts, abstracts, oral/slide presentations, book chapters) based on this study will be reviewed by each sub-investigator and by the sponsor prior to submission. All communication or publications concerning the project, including at a conference or seminar, shall acknowledge the Parties and the Wellcome trust financial contribution

#### 13. References

1. Marchello, C. S., Dale, A. P., Pisharody, S., Rubach, M. P. & Crump, J. A. A Systematic Review and Meta-analysis of the Prevalence of Community-Onset Bloodstream Infections among Hospitalized Patients in Africa and Asia. *Antimicrob. Agents Chemother.* **64**, (2020).
2. Stanaway, J. D. *et al.* The global burden of non-typhoidal salmonella invasive disease: a systematic analysis for the Global Burden of Disease Study 2017. *Lancet Infect. Dis.* **19**, 1312–1324 (2019).
3. Ao TT, Feasey NA, Gordon MA, Keddy KH, A. F. Global burden of invasive nontyphoidal Salmonella disease, 2010. *Emerg. Infect. Dis.* (2015).
4. Crump, J. A., Sjölund-Karlsson, M., Gordon, M. A. & Parry, C. M. Epidemiology, clinical presentation, laboratory diagnosis, antimicrobial resistance, and antimicrobial management of invasive Salmonella infections. *Clin. Microbiol. Rev.* **28**, 901–937 (2015).
5. Tacconelli, E. *et al.* Discovery, research, and development of new antibiotics: The WHO priority list of antibiotic-resistant bacteria and tuberculosis. *Lancet Infect. Dis.* **18**, 318–327 (2017).
6. Feasey, N. A., Dougan, G., Kingsley, R. A., Heyderman, R. S. & Gordon, M. A. Invasive non-typhoidal salmonella disease: an emerging and neglected tropical disease in Africa. *Lancet* **379**, 2489–2499 (2012).
7. Marchello, C. S. *et al.* Complications and mortality of non-typhoidal salmonella invasive disease: a global systematic review and meta-analysis. *Lancet Infect. Dis.* **3099**, 1–14 (2022).
8. Klemm, E. J. *et al.* Emergence of host-adapted Salmonella Enteritidis through rapid evolution in an immunocompromised host. *Nat. Microbiol.* **1**, (2016).
9. MacLennan, C. A. *et al.* Dysregulated humoral immunity to nontyphoidal Salmonella in HIV-infected African adults. *Science* **328**, 508–12 (2010).
10. Feasey, N. A. *et al.* Modelling the Contributions of Malaria, HIV, Malnutrition and Rainfall to the Decline in Paediatric Invasive Non-typhoidal Salmonella Disease in Malawi. *PLoS Negl. Trop. Dis.* **9**, e0003979 (2015).
11. Reddy, E. A., Shaw, A. V. & Crump, J. A. Community-acquired bloodstream infections in Africa: a systematic review and meta-analysis. *Lancet Infect. Dis.* **10**, 417–432 (2010).
12. Pulford, C. V. *et al.* Stepwise evolution of Salmonella Typhimurium ST313 causing bloodstream infection in Africa. *Nat. Microbiol.* 1–12 (2020) doi:10.1038/s41564-020-00836-1.
13. Kingsley, R. a *et al.* Epidemic multiple drug resistant Salmonella Typhimurium causing invasive disease in sub-Saharan Africa have a distinct genotype. *Genome Res.* **19**, 2279–87 (2009).
14. McClelland, M. *et al.* Comparison of genome degradation in Paratyphi A and Typhi, human-restricted serovars of Salmonella enterica that cause typhoid. *Nat. Genet.* **36**, 1268–74 (2004).
15. Preciado-Llanes, L. *et al.* Evasion of MAIT cell recognition by the African Salmonella Typhimurium ST313 pathovar that causes invasive disease. *Proc. Natl. Acad. Sci. U. S. A.* **117**, 20717–20728 (2020).
16. Feasey, N. A. *et al.* Erratum: Distinct Salmonella Enteritidis lineages associated with enterocolitis in high-income settings and invasive disease in low-income settings. *Nat. Genet.* **49**, 651–651 (2017).
17. Crump, J. A. *et al.* Investigating the Meat Pathway as a Source of Human Nontyphoidal Salmonella Bloodstream Infections and Diarrhea in East Africa. *Clin. Infect. Dis.* 4–6 (2020) doi:10.1093/cid/ciaa1153.
18. Post, A. S. *et al.* Supporting evidence for a human reservoir of invasive non-Typhoidal Salmonella from household samples in Burkina Faso. *PLoS Negl. Trop. Dis.* **13**, e0007782 (2019).
19. Musicha, P. *et al.* Trends in antimicrobial resistance in bloodstream infection isolates at a large urban hospital in Malawi (1998–2016): a surveillance study. *Lancet Infect. Dis.* **17**, 1042–1052 (2017).
20. Van Puyvelde, S. *et al.* An African Salmonella Typhimurium ST313 sublineage with extensive drug-resistance and signatures of host adaptation. *Nat. Commun.* **10**, 1–12 (2019).
21. Kariuki, S. *et al.* High relatedness of invasive multi-drug resistant non-typhoidal Salmonella genotypes among patients and asymptomatic carriers in endemic informal settlements in Kenya. *PLoS Negl. Trop. Dis.* **14**, e0008440 (2020).
22. MacLennan, C. A., Martin, L. B. & Micoli, F. Vaccines against invasive Salmonella disease: Current status and future directions. *Hum. Vaccines Immunother.* **10**, 1478–1493 (2014).
23. Baliban, S. M., Lu, Y. J. & Malley, R. Overview of the nontyphoidal and paratyphoidal Salmonella vaccine pipeline: Current status and future prospects. *Clin. Infect. Dis.* **71**, S151–S154 (2020).
24. Simon, R. *et al.* Salmonella enterica Serovar Enteritidis core O Polysaccharide conjugated to H:g,m Flagellin as a candidate vaccine for protection against invasive infection with S. Enteritidis. *Infect. Immun.* **79**, 4240–4249 (2011).

25. Baliban, S. M. *et al.* Development of a glycoconjugate vaccine to prevent invasive Salmonella Typhimurium infections in sub-Saharan Africa. *PLoS Negl. Trop. Dis.* **11**, e0005493 (2017).
26. Baliban, S. M. *et al.* Immunogenicity and efficacy following sequential parenterally-administered doses of Salmonella Enteritidis COPS:FliC glycoconjugates in infant and adult mice. *PLoS Negl. Trop. Dis.* **12**, (2018).
27. Baliban, S. M. *et al.* Immunogenicity and induction of functional antibodies in rabbits immunized with a trivalent typhoid-invasive nontyphoidal Salmonella glycoconjugate formulation. *Molecules* **23**, (2018).
28. ClinicalTrials.gov. Salmonella Conjugates CVD 1000: Study of Responses to Vaccination With Trivalent Invasive Salmonella Disease Vaccine - Full Text View - ClinicalTrials.gov. https://clinicaltrials.gov/ct2/show/NCT03981952.
29. Micoli, F. *et al.* Comparative immunogenicity and efficacy of equivalent outer membrane vesicle and glycoconjugate vaccines against nontyphoidal Salmonella. *Proc. Natl. Acad. Sci. U. S. A.* (2018) doi:10.1073/pnas.1807655115.
30. Tennant, S. M. *et al.* Refined live attenuated Salmonella enterica serovar typhimurium and enteritidis vaccines mediate homologous and heterologous serogroup protection in mice. *Infect. Immun.* **83**, 4504–4512 (2015).
31. Tennant, S. M., MacLennan, C. A., Simon, R., Martin, L. B. & Khan, M. I. Nontyphoidal salmonella disease: Current status of vaccine research and development. *Vaccine* **34**, 2907–2910 (2016).
32. Dougan, G., John, V., Palmer, S. & Mastroeni, P. Immunity to salmonellosis. *Immunol. Rev.* **240**, 196–210 (2011).
33. MacLennan, C. A. *et al.* The neglected role of antibody in protection against bacteremia caused by nontyphoidal strains of Salmonella in African children. *J. Clin. Invest.* **118**, 1553–1562 (2008).
34. Nyirenda, T. S., Mandala, W. L., Gordon, M. A. & Mastroeni, P. Immunological bases of increased susceptibility to invasive nontyphoidal Salmonella infection in children with malaria and anaemia. *Microbes Infect.* **20**, 589–598 (2018).
35. Mastroeni, P. & Rossi, O. Antibodies and protection in systemic salmonella infections: Do we still have more questions than answers? *Infect. Immun.* **88**, (2020).
36. Darton, T. C. *et al.* Using a Human Challenge Model of Infection to Measure Vaccine Efficacy: A Randomised, Controlled Trial Comparing the Typhoid Vaccines M01ZH09 with Placebo and Ty21a. *PLoS Negl. Trop. Dis.* **10**, e0004926 (2016).
37. Jin, C. *et al.* Vi-specific serological correlates of protection for typhoid fever. *J. Exp. Med.* **218**, (2020).
38. WHO EXPERT COMMITTEE ON BIOLOGICAL STANDARDIZATION. *Human Challenge Trials for Vaccine Development: regulatory considerations*. (2016).
39. Cohen, D. *et al.* Serum IgG antibodies to Shigella lipopolysaccharide antigens–a correlate of protection against shigellosis. *Hum. Vaccines Immunother.* **15**, 1401–1408 (2019).
40. Jin, C. *et al.* Efficacy and immunogenicity of a Vi-tetanus toxoid conjugate vaccine in the prevention of typhoid fever using a controlled human infection model of Salmonella Typhi: a randomised controlled, phase 2b trial. *The Lancet* **390**, 2472–2480 (2017).
41. Shakya, M. *et al.* Phase 3 Efficacy Analysis of a Typhoid Conjugate Vaccine Trial in Nepal. *N. Engl. J. Med.* **381**, 2209–2218 (2019).
42. Chen, W. H. *et al.* Single-dose Live Oral Cholera Vaccine CVD 103-HgR Protects Against Human Experimental Infection With *Vibrio cholerae* O1 El Tor. *Clin. Infect. Dis.* **62**, 1329–1335 (2016).
43. High level efficacy in humans of a next-generation plasmodium falciparum anti-sporozoite vaccine: r21 in matrix-mTM adjuvant | Cochrane Library. https://www.cochranelibrary.com/central/doi/10.1002/central/CN-01462017/full.
44. Datoo, M. S. *et al.* Efficacy of a low-dose candidate malaria vaccine, R21 in adjuvant Matrix-M, with seasonal administration to children in Burkina Faso: a randomised controlled trial. *Lancet Lond. Engl.* **397**, 1809–1818 (2021).
45. Hornick, R. & Greisman, S. Typhoid fever: pathogenesis and immunologic control. 2. *N. Engl. …* **283**, 739–46 (1970).
46. Waddington, C. S. *et al.* Advancing the management and control of typhoid fever: a review of the historical role of human challenge studies. *J. Infect.* **68**, 405–18 (2014).
47. Hornick, R. B. *et al.* Typhoid fever: pathogenesis and immunologic control. *N. Engl. J. Med.* **283**, 686–91 (1970).
48. Waddington, C. S. *et al.* An outpatient, ambulant-design, controlled human infection model using escalating doses of Salmonella Typhi challenge delivered in sodium bicarbonate solution. *Clin. Infect. Dis. Off. Publ. Infect. Dis. Soc. Am.* **58**, 1230–40 (2014).
49. Gibani, M. M. *et al.* Investigation of the role of typhoid toxin in acute typhoid fever in a human challenge model. *Nat. Med.* **25**, 1082–1088 (2019).
50. Gibani, M. M. *et al.* The impact of vaccination and prior exposure on stool shedding of salmonella typhi and salmonella paratyphi in 6 controlled human infection studies. *Clin. Infect. Dis.* **68**, (2019).
51. Higginson, E. E., Simon, R. & Tennant, S. M. Animal models for salmonellosis: Applications in vaccine research. *Clin. Vaccine Immunol.* **23**, 746–756 (2016).
52. Ault, A. *et al.* Safety and tolerability of a live oral Salmonella typhimurium vaccine candidate in SIV-infected nonhuman primates. *Vaccine* **31**, 5879–5888 (2013).

53. Tsolis, R. M., Xavier, M. N., San1. Tsolis RM, Xavier MN, Santos RL, Bäumler AJ. How to become a top model: Impact of animal experimentation on human Salmonella disease research [Internet]. Vol. 79, Infection and Immunity. Infect Immun; 2011 [cited 2021 Jun 1]. p. 1806–14. Available f, R. L. & Bäumler, A. J. How to become a top model: Impact of animal experimentation on human Salmonella disease research. *Infect. Immun.* **79**, 1806–1814 (2011).
54. Hormaeche, E., Peluffo, C. & Aleppo, P. Nuevo contrabucion al estudio etiologico de las 'Diarreas infantiles de Verano'. *Arch. Urug. Med.* **9**, 113–163 (1939).
55. McCullough, N. B. & Eisele, C. W. Experimental human salmonellosis. *J. Infect. Dis.* **88**, 278–289 (1951).
56. Angelakopoulos, H. & Hohmann, E. L. Pilot study of phoP/phoQ-deleted Salmonella enterica serovar typhimurium expressing Helicobacter priori urease in adult volunteers. *Infect. Immun.* **68**, 2135–2141 (2000).
57. Hindle, Z. *et al.* Characterization of Salmonella enterica derivatives harboring defined aroC and Salmonella pathogenicity island 2 type III secretion system (ssaV) mutations by immunization of healthy volunteers. *Infect. Immun.* **70**, 3457–3467 (2002).
58. Jertborn, M., Haglind, P., Iwarson, S. & Svennerholm, A. M. Estimation of Symptomatic and Asymptomatic Salmonella Infections. *Scand. J. Infect. Dis.* **22**, 451–455 (2009).
59. Roberts, D. Factors contributing to outbreaks of food poisoning in England and Wales 1970-1979. *J. Hyg. (Lond.)* **89**, 491–498 (1982).
60. Buchwald, D. S. & Blaser, M. J. A review of human salmonellosis: II. Duration of excretion following infection with nontyphi Salmonella. *Rev. Infect. Dis.* **6**, 345–356 (1984).
61. Bennett, J. E., Dolin, R. & Blaser, M. J. *Mandell, Douglas, and Bennett's Principles and Practice of Infectious Diseases*. (Elsevier, 2019). doi:19-09-2019.
62. Hohmann, E. L. Nontyphoidal Salmonella: Gastrointestinal infection and carriage - UpToDate. https://www.uptodate.com/contents/nontyphoidal-salmonella-gastrointestinal-infection-and-carriage?search=salmonella gastroenteritis&source=search_result&selectedTitle=1~150&usage_type=default&display_rank=1#H6.
63. Hohmann, E. L. Nontyphoidal Salmonella: Microbiology and epidemiology - UpToDate. https://www.uptodate.com/contents/nontyphoidal-salmonella-microbiology-and-epidemiology?search=salmonella gastroenteritis&topicRef=2697&source=see_link.
64. Kotton, C. N. & Hohmann, E. L. Pathogenesis of Salmonella gastroenteritis - UpToDate. https://www.uptodate.com/contents/pathogenesis-of-salmonella-gastroenteritis?search=salmonella gastroenteritis&topicRef=2697&source=see_link.
65. Hohmann, E. L. Nontyphoidal Salmonella bacteremia - UpToDate. https://www.uptodate.com/contents/nontyphoidal-salmonella-bacteremia?search=salmonella gastroenteritis&topicRef=2697&source=see_link.
66. Dione, M. M. *et al.* Clonal Differences between Non-Typhoidal Salmonella (NTS) Recovered from Children and Animals Living in Close Contact in The Gambia. *PLoS Negl. Trop. Dis.* **5**, e1148 (2011).
67. Koolman, L. *et al.* Case-control investigation of invasive Salmonella disease in Africa reveals no evidence of environmental or animal reservoirs of invasive strains. *medRxiv* 2022.01.31.22270114 (2022) doi:10.1101/2022.01.31.22270114.
68. Public Health England. Management of Gastrointestinal Infections 2019. (2019).
69. Onwuezobe, I. A., Oshun, P. O. & Odigwe, C. C. Antimicrobials for treating symptomatic non-typhoidal Salmonella infection. *Cochrane Database Syst. Rev.* **11**, (2012).
70. Shane, A. L. *et al.* 2017 Infectious Diseases Society of America Clinical Practice Guidelines for the Diagnosis and Management of Infectious Diarrhea. *Clin. Infect. Dis.* **65**, e45–e80 (2017).
71. Ajene, A. N., Fischer Walker, C. L. & Black, R. E. Enteric Pathogens and Reactive Arthritis: A Systematic Review of Campylobacter, Salmonella and Shigella-associated Reactive Arthritis. *J. Health Popul. Nutr.* **31**, 299 (2013).
72. Hannu, T. Reactive arthritis. *Best Pract. Res. Clin. Rheumatol.* **25**, 347–57 (2011).
73. Laasila, K., Laasonen, L. & Leirisalo-Repo, M. Antibiotic treatment and long term prognosis of reactive arthritis. *Ann. Rheum. Dis.* **62**, 655 (2003).
74. Yli-Kerttula, T. *et al.* Effect of a three month course of ciprofloxacin on the outcome of reactive arthritis. *Ann. Rheum. Dis.* **59**, 565 (2000).
75. Kvien, T. K. *et al.* Three month treatment of reactive arthritis with azithromycin: a EULAR double blind, placebo controlled study. *Ann. Rheum. Dis.* **63**, 1113–1119 (2004).
76. Townes, J. M. Reactive arthritis after enteric infections in the United States: the problem of definition. *Clin. Infect. Dis. Off. Publ. Infect. Dis. Soc. Am.* **50**, 247–254 (2010).
77. Aho, K., Ahvonen, P., Lassus, A., Sievers, K. & Tiilikainen, A. HL-A antigen 27 and reactive arthritis. *Lancet Lond. Engl.* **2**, 157 (1973).

78. Leirisalo-Repo, M., Hannu, T. & Mattila, L. Microbial factors in spondyloarthropathies: insights from population studies. *Curr. Opin. Rheumatol.* **15**, 408–412 (2003).
79. Dupont, H. L. Gastrointestinal infections and the development of irritable bowel syndrome. *Curr. Opin. Infect. Dis.* **24**, 503–508 (2011).
80. Haagsma, J. A., Siersema, P. D., De Wit, N. J. & Havelaar, A. H. Disease burden of post-infectious irritable bowel syndrome in The Netherlands. *Epidemiol. Infect.* **138**, 1650–1656 (2010).
81. Avery, M. E. & Snyder, J. D. Oral Therapy for Acute Diarrhea - The Underused Simple Solution. *N. Engl. J. Med.* **323**, 891–894 (1990).
82. Leinert, J. L., Weichert, S., Jordan, A. J. & Adam, R. Non-Typhoidal Salmonella Infection in Children: Influence of Antibiotic Therapy on Postconvalescent Excretion and Clinical Course—A Systematic Review. *Antibiot. 2021 Vol 10 Page 1187* **10**, 1187 (2021).
83. Marzel, A. *et al.* Persistent Infections by Nontyphoidal Salmonella in Humans: Epidemiology and Genetics. *Clin. Infect. Dis.* **62**, 879–886 (2016).
84. Goodman, L. J. *et al.* Empiric Antimicrobial Therapy of Domestically Acquired Acute Diarrhea in Urban Adults. *Arch. Intern. Med.* **150**, 541–546 (1990).
85. Dryden, M. S., Gabb, R. J. E. & Wright, S. K. Empirical treatment of severe acute community-acquired gastroenteritis with ciprofloxacin. *Clin. Infect. Dis. Off. Publ. Infect. Dis. Soc. Am.* **22**, 1019–1025 (1996).
86. Wistrom, J. *et al.* Empiric treatment of acute diarrheal disease with norfloxacin. A randomized, placebo-controlled study. Swedish Study Group. *Ann. Intern. Med.* **117**, 202–208 (1992).
87. Mattila, L. *et al.* Short-term treatment of traveler's diarrhea with norfloxacin: a double-blind, placebo-controlled study during two seasons. *Clin. Infect. Dis. Off. Publ. Infect. Dis. Soc. Am.* **17**, 779–782 (1993).
88. Benenson, S. *et al.* The risk of vascular infection in adult patients with nontyphi Salmonella bacteremia. *Am. J. Med.* **110**, 60–63 (2001).
89. Cohen, P. S., O'Brien, T. F., Schoenbaum, S. C. & Medeiros, A. A. The risk of endothelial infection in adults with salmonella bacteremia. *Ann. Intern. Med.* **89**, 931–932 (1978).
90. Hohmann, E. L. Nontyphoidal salmonellosis. *Clin. Infect. Dis.* **32**, 263–269 (2001).
91. Parry, C. M. *et al.* A retrospective study of secondary bacteraemia in hospitalised adults with community acquired non-typhoidal Salmonella gastroenteritis. *BMC Infect. Dis.* **13**, 107 (2013).
92. Neal, R. K., Brij, O. S., Slack, B. C. R., Hawkey, J. C. & Logan, R. F. A. Recent treatment with H2 antagonists and antibiotics and gastric surgery as risk factors for salmonella infection. *BMJ* **308**, 176 (1994).
93. Pavia, A. T. *et al.* Epidemiologic evidence that prior antimicrobial exposure decreases resistance to infection by antimicrobial-sensitive Salmonella. *J. Infect. Dis.* **161**, 255–260 (1990).
94. Hung, T. Y., Liu, M. C., Hsu, C. F. & Lin, Y. C. Rotavirus infection increases the risk of bacteremia in children with nontyphoid Salmonella gastroenteritis. *Eur. J. Clin. Microbiol. Infect. Dis.* **28**, 425–428 (2009).
95. Gordon, M. A. Salmonella infections in immunocompromised adults. *J. Infect.* **56**, 413–422 (2008).
96. Gordon, M. A. *et al.* Invasive non-typhoid salmonellae establish systemic intracellular infection in HIV-infected adults: An emerging disease pathogenesis. *Clin. Infect. Dis.* **50**, 953–962 (2010).
97. Ramos, J. M., García-Corbeira, P., Aguado, J. M., Alés, J. M. & Soriano, F. Classifying extraintestinal non-typhoid Salmonella infections. *QJM - Mon. J. Assoc. Physicians* **89**, 123–126 (1996).
98. Thuluvath, P. J. & Mckendrick, M. W. *Salmonella and Complications Related to Age Sheffield Experience*. *Quarterly Journal of Medicine* vol. 67 497–503 https://academic.oup.com/qjmed/article/67/3/497/1571129 (1988).
99. McCarron, B. A 3-year retrospective review of 132 patients with Salmonella enterocolitis admitted to a regional infectious diseases unit. *J. Infect.* **37**, 136–139 (1998).
100. Mandal, B. K. & Brennand, J. Bacteraemia in salmonellosis: A 15 year retrospective study from a regional infectious diseases unit. *Br. Med. J.* **297**, 1242–1243 (1988).
101. Jones, T. F. *et al.* Salmonellosis outcomes differ substantially by serotype. *J. Infect. Dis.* **198**, 109–114 (2008).
102. Kaplan, J., Ikeda, S., McNeil, J. C., Kaplan, S. L. & Vallejo, J. G. Microbiology of Osteoarticular Infections in Patients with Sickle Hemoglobinopathies at Texas Children's Hospital, 2000-2018. *Pediatr. Infect. Dis. J.* **38**, 1251–1253 (2019).
103. Gal-Mor, O. Persistent infection and long-term carriage of typhoidal and nontyphoidal salmonellae. *Clin. Microbiol. Rev.* **32**, (2019).
104. Sirinavin, S. *et al.* Norfloxacin and azithromycin for treatment of nontyphoidal salmonella carriers. *Clin. Infect. Dis. Off. Publ. Infect. Dis. Soc. Am.* **37**, 685–691 (2003).
105. Sirinavin, S., Pokawattana, L. & Bangtrakulnondh, A. Duration of Nontyphoidal Salmonella Carriage in Asymptomatic Adults. *Clin. Infect. Dis.* **38**, 1644–1645 (2004).
106. Nelson, J. D., Kusmiesz, H., Jackson, L. H. & Woodman, E. Treatment of Salmonella Gastroenteritis with Ampicillin, Amoxicillin, or Placebo. *Pediatrics* **65**, (1980).

107. Neill, M. A. *et al.* Failure of ciprofloxacin to eradicate convalescent fecal excretion after acute salmonellosis: Experience during an outbreak in health care workers. *Ann. Intern. Med.* **114**, 195–199 (1991).
108. Carlstedt, G. *et al.* Norfloxacin treatment of salmonellosis does not shorten the carrier stage. *Scand. J. Infect. Dis.* **22**, 553–556 (1990).
109. Musher, D. M. & Rubenstein, A. D. Permanent Carriers of Nontyphosa Salmonellae. *Arch. Intern. Med.* **132**, 869–872 (1973).
110. Blaser, M. J. & Newman, L. S. *A Review of Human Salmonellosis: I. Infective Dose*. *REVIEWS OF INFECTIOUS DISEASES* • vol. 4 https://academic.oup.com/cid/article/4/6/1096/288158 (1982).
111. Hornick R.B, Greisman S.E, Woodward T.E, DuPont H.L, Dawkins A.T, S. M. J. Typhoid fever: pathogenesis and immunologic control (first of two parts). *N. Engl. J. Med.* **283**, 686–691 (1970).
112. Mackenzie, K. D. *et al.* Parallel evolution leading to impaired biofilm formation in invasive Salmonella strains. *PLoS Genet.* **15**, e1008233 (2019).
113. Owen, S. V. *et al.* Characterization of the prophage repertoire of African Salmonella Typhimurium ST313 reveals high levels of spontaneous induction of novel phage BTP1. *Front. Microbiol.* **8**, 235 (2017).
114. Canals, R. *et al.* Adding function to the genome of African Salmonella Typhimurium ST313 strain D23580. *PLoS Biol.* **17**, e3000059 (2019).
115. Wood, M. W. *et al.* Identification of a pathogenicity island required for Salmonella enteropathogenicity. *Mol. Microbiol.* **29**, 883–891 (1998).
116. Okoro, C. K. *et al.* Signatures of Adaptation in Human Invasive Salmonella Typhimurium ST313 Populations from Sub-Saharan Africa. *PLoS Negl. Trop. Dis.* **9**, e0003611 (2015).
117. Honeycutt, J. D. *et al.* Genetic variation in the MacAB-TolC efflux pump influences pathogenesis of invasive Salmonella isolates from Africa. *PLoS Pathog.* **16**, e1008763 (2020).
118. Carden, S. E. *et al.* Pseudogenization of the Secreted Effector Gene sseI Confers Rapid Systemic Dissemination of S. Typhimurium ST313 within Migratory Dendritic Cells. *Cell Host Microbe* **21**, 182–194 (2017).
119. Aulicino, A. *et al.* Invasive Salmonella exploits divergent immune evasion strategies in infected and bystander dendritic cell subsets. *Nat. Commun.* **9**, (2018).
120. Hammarlöf, D. L. *et al.* Role of a single noncoding nucleotide in the evolution of an epidemic African clade of Salmonella. *Proc. Natl. Acad. Sci. U. S. A.* **115**, E2614–E2623 (2018).
121. Ashton, P. M. *et al.* Public health surveillance in the UK revolutionises our understanding of the invasive Salmonella Typhimurium epidemic in Africa. *Genome Med.* **9**, 1–13 (2017).
122. Kröger, C. *et al.* An infection-relevant transcriptomic compendium for salmonella enterica serovar typhimurium. *Cell Host Microbe* **14**, 683–695 (2013).
123. van Prehn, J. *et al.* European Society of Clinical Microbiology and Infectious Diseases: 2021 update on the treatment guidance document for Clostridioides difficile infection in adults. *Clin. Microbiol. Infect.* **27**, S1–S21 (2021).
124. Talaat, K. R. *et al.* Consensus Report on Shigella Controlled Human Infection Model: Conduct of Studies. *Clin. Infect. Dis.* **69**, S580–S590 (2019).
125. Levine, M. M. *et al.* ESCHERICHIA COLI STRAINS THAT CAUSE DIARRHŒA BUT DO NOT PRODUCE HEAT-LABILE OR HEAT-STABLE ENTEROTOXINS AND ARE NON-INVASIVE. *The Lancet* **311**, 1119–1122 (1978).
126. Lewis, S. J. & Heaton, K. W. Stool form scale as a useful guide to intestinal transit time. *Scand. J. Gastroenterol.* **32**, 920–924 (1997).
127. Maclennan, C. A. *et al.* Consensus Report on Shigella Controlled Human Infection Model: Clinical Endpoints. *Clin. Infect. Dis.* **69**, S591–S595 (2019).
128. Rome Foundation. Rome IV Criteria - Rome Foundation. https://theromefoundation.org/rome-iv/rome-iv-criteria/.
129. Public Health England. Immunisation against infectious disease.
130. Palsson, O. S. *et al.* Development and Validation of the Rome IV Diagnostic Questionnaire for Adults. *Gastroenterology* **150**, 1481–1491 (2016).
131. Francis, C. Y., Morris, J. & Whorwell, P. J. The irritable bowel severity scoring system: a simple method of monitoring irritable bowel syndrome and its progress. *Aliment. Pharmacol. Ther.* **11**, 395–402 (1997).
132. Berumen, A. *et al.* Characteristics and Risk Factors of Post-Infection Irritable Bowel Syndrome After Campylobacter Enteritis. *Clin. Gastroenterol. Hepatol.* **19**, 1855-1863.e1 (2021).
133. Gordon, M. A. *et al.* Non-typhoidal salmonella bacteraemia among HIV-infected Malawian adults: high mortality and frequent recrudescence. *AIDS Lond. Engl.* **16**, 1633–41 (2002).
134. Safe, A. F., Maxwell, R. T., Howard, A. J. & Garcia, R. C. Relapsing Salmonella enteritidis infection in a young adult male with chronic granulomatous disease. *Postgrad. Med. J.* **67**, 198 (1991).
135. Mori, N., Szvalb, A. D., Adachi, J. A., Tarrand, J. J. & Mulanovich, V. E. Clinical presentation and outcomes of non-typhoidal Salmonella infections in patients with cancer. *BMC Infect. Dis.* **21**, (2021).
136. NCTC Culture Collections. https://www.culturecollections.org.uk/collections/nctc.aspx.

137. Imperial College Healthcare Tissue Bank | Faculty of Medicine | Imperial College London. https://www.imperial.ac.uk/imperial-college-healthcare-tissue-bank.
138. Oguti, B. *et al.* Factors influencing participation in controlled human infection models: a pooled analysis from six enteric fever studies. *Wellcome Open Res. 2019 4153* **4**, 153 (2019).
139. PHE. UK Standards for Microbiology Investigations Identification of Salmonella species. (2021).
140. Public Health England. Examining food, water and environmental samples from healthcare environments Microbiological guidelines. (2020).
141. Rawlinson, S., Ciric, L. & Cloutman-Green, E. How to carry out microbiological sampling of healthcare environment surfaces? A review of current evidence. *J. Hosp. Infect.* **103**, 363–374 (2019).
142. Darton, T. C. *et al.* Blood culture-PCR to optimise typhoid fever diagnosis after controlled human infection identifies frequent asymptomatic cases and evidence of primary bacteraemia. *J. Infect.* (2017) doi:10.1016/j.jinf.2017.01.006.
143. Darton, T. C. *et al.* Identification of novel serodiagnostic signatures of typhoid fever using a Salmonella proteome array. *Front. Microbiol.* **8**, 1–14 (2017).
144. Sztein, M. B. Cell-mediated immunity and antibody responses elicited by attenuated Salmonella enterica Serovar Typhi strains used as live oral vaccines in humans. *Clin. Infect. Dis. Off. Publ. Infect. Dis. Soc. Am.* **45 Suppl 1**, (2007).
145. Wheeler, G. M. *et al.* How to design a dose-finding study using the continual reassessment method. *BMC Med. Res. Methodol.* **19**, 1–15 (2019).
146. O'Quigley, J., Pepe, M. & Fisher, L. Continual Reassessment Method: A Practical Design for Phase 1 Clinical Trials in Cancer. *Biometrics* **46**, 33 (1990).

#### 14. Appendix - Amendment history

| Amendment No. | Protocol Version No. | Date issued | Author(s) of changes | Details of Changes made |
|---|---|---|---|---|
| 5 | 3.1 | | Malick Gibani, Xinxiue Liu | **Section 9.2 -** The study has moved from using a standard model-based approach to a modified model-based approach for determining the appropriate dose of *Salmonella* to use in the study. This change was made to reduce the number of participants exposed to *Salmonella* while still ensuring that the data collected remains reliable and meaningful. The modified approach allows the study to stop earlier in certain cases, meaning fewer participants are needed overall, which helps to minimize risks and burdens on volunteers. This amendment was recommended by the trial statisticians after reviewing interim data. |
| 5 | 3.0 | 03-APR-2024 | Malick Gibani | • Interim Medical History Recording:<br> ○ Removed the requirement to record interim medical history post-enrolment.<br> ○ All changes in health status, abnormal test results, or symptoms will now be recorded as adverse events instead.<br>• Management of Relapse:<br> ○ Added clarification on the treatment and management of clinical or microbiological relapse ("second-line" treatment) after the primary course of antibiotics (Section 6.9.2).<br> ○ No changes to the type, dose, or duration of antibiotics specified in the protocol.<br>• Screening of Close Contacts:<br> ○ Revised the process to provide information for close contacts only upon participant request, rather than automatically.<br>• Convalescent/Chronic Carriage Management:<br> ○ Participants experiencing relapse after the primary treatment may be offered a second short course of second-line antibiotics as per the protocol.<br> ○ If bacteria persist after the second course or are still detectable at 1 year, participants will not receive further treatment but will be referred to an infectious disease (ID) specialist in cases of clinical relapse. |
| 3 | 2.2 | 19-FEB-2024 | Malick Gibani | Change to the order of the first line and second line antibiotic treatments for prolonged shedding and/or gastroenteritis. The first line antibiotic treatment for prolonged shedding and gastroenteritis is changed from from ciprofloxin (500mg twice daily for 5 days) to azithromycin (500mg once daily for 5 days, formerly the second line treatment) in response to a recently issued advice form the MHRA regarding the safety of fluoroquinolene antibiotics. Both Salmonella |

| | | | | |
|---|---|---|---|---|
| | | | | strains are sensitive to azithromycin and ciprofloxacin. This change can be implemented within existing resource in place.<br><br>Sections 6.9 and Figure 4 have been amended. |
| **2** | **2.1** | **14-JAN-2024** | **Malick Gibani** | Antibiotic treatment criteria - The following sections have been amended to clarify the policy on antibiotic treatment. It is recognised that in some instances, the risk of antibiotic related adverse effects outweighs the potential benefit of antibiotic treatment. This is reflected below:<br><br>• Section 4.8.9.1<br>• Section 6.9.1.2 Transient asymptomatic shedding<br>• Section 6.9.1.2 Spontaneous clearance<br>• Section 6.9.1.3 Asymptomatic/Pauci-symptomatic shedding<br>• Table 10<br>• Figure 1<br>• Figure 4<br><br>DSMB Updates - The following sections have been added to clarify timings of DSMC review. Specifically:<br>• Section 3. 1 Study design<br>The DSMC will be provided with an updated report after each cohort of 10 completes challenged.<br><br>• Section 8.4.2. The following clause has been removed “has attended their day 28 follow up visit”. All bacteraemia cases will still be reviewed by the DSMC.<br><br>Visitors - The following section has been added to clarify the policy on visitors to the inpatient quarantine facility.<br>• Section 6.6.1.1.9 Visitors<br><br>Modification of temporary exclusion criteria to remove screening for resistant organisms.<br><br>Screening tests changed to test HbA1c in place of capillary blood glucose and inclusion of urine dipstick test in the pre-registration evaluation (section 4.3).<br><br>Table 7 – Footnote updated to clarify that physical examination only performed if clinically indicated at the discretion of the attending study physician. Footnote updated to record that any intermin medical history to be recorded as an adverse event.<br><br>Study window period for Day -7 visit changed to 7 days beforehand and 3 days after.<br><br>Mood questionnaire removed at Day 14. |

| | | | | |
|---|---|---|---|---|
| **1** | **2.0** | **18-SEP-2023** | **Malick Gibani** | The study exclusion criteria (section 4.4) have been clarified. Specifically:<br><br>• History of recent malaria infection within the past 12 months from screening. Volunteers with distant history of malaria infection may be considered for enrolment if they have been treated and can demonstrate a negative malaria test at screening.<br><br>• Screening blood test positive for HLA-B27<br><br>Table 2, Section 6.4.4.1, Section 6.5.2 - We have amended details of the challenge agent preparation, to reflect results of laboratory validation experiments. In short, stability of the challenge agent was superior when suspended in 0.9% saline as compared with sodium bicarbonate. The total sodium bicarbonate buffer administered will remain the same.<br><br>Protocol section 6.6.1.2 Food - There will be no specific restrictions to the food that can be eaten during the inpatient quarantine period. Participants will be provided with breakfast, lunch and dinner during their inpatient stay. Participants will be allowed to bring snacks and order external meals. Participants will be asked to record details of their diet in the diary card.<br><br>Protocol – Section 6.4.3 – Update to change from electronic diary to paper diary.<br><br>Table 8 - An extra 5ml of blood will be collected at day 7 and day 14, an extra 10ml of blood will be collected at day 365 and an extra 15ml blood will be collected at day 4. This will bring the total volume of blood collected from 673ml to 708ml. An extra saliva sample will be collected on Day 7. These changes have been made following finalisation of laboratory analysis plan and to ensure that there are sufficient samples to meet secondary objectives. The PIS has been updated accordingly.<br><br>References to online diary removed and changed to paper diary. |
